# Exposome changes in primary school children following the wide population non-pharmacological interventions implemented due to COVID-19 in Cyprus: a national survey

**DOI:** 10.1101/2020.10.21.20216978

**Authors:** C. Konstantinou, X.D. Andrianou, A. Constantinou, A. Perikkou, E. Markidou, C.A. Christophi, K.C. Makris

**Affiliations:** Cyprus International Institute for Environmental and Public Health, Cyprus University of Technology, Limassol, Cyprus; Department of Nutrition, Cyprus Ministry of Health, Nicosia, Cyprus

**Keywords:** exposome, SARS-CoV-2, pandemic, COVID-19, confinement, mitigation, public health response, lockdown, primary school children, compliance, EWAS

## Abstract

**Background:** School closures were part of a series of non-pharmacological intervention (NPI) measures addressing the COVID-19 pandemic in Cyprus. We aimed to study changes in the environment, diet, behavior, personal hygiene, contacts, lifestyle choices and the degree of compliance to NPI measures by primary school children in Cyprus at school and at home for two periods, i.e., before lockdown and during the school re-opening using the methodological context of the human exposome.

**Methods:** During June 2020, an online survey questionnaire was forwarded to parents of primary school children through schools’ administrations, with questions about the children’s lifestyle/behaviours for two periods; school re-opening, following the population-wide lockdown (May 21-June 26, 2020), and the school period before lockdown (before March 2020). Descriptive statistics and exposome wide association analysis were implemented to agnostically assess associations of demographic, lifestyle and behavioral parameters with the degree of compliance to NPI measures.

**Findings:** A total of 1509 children from more than 180 primary schools (out of 330 schools) in Cyprus were included. Median number of contacts per day at home, school and other places during weekdays was lower (p<0.001) in the post-lockdown period compared to the pre-lockdown period (5 vs 12, 10 vs 29 and 6 vs 14, respectively). Vulnerable contacts with children also decreased from 2[1, 3] in the pre-lockdown to 1[0, 2] in the post-lockdown period (p<0.001). Differences in sugary and ready-made food consumption, physical activity, screen time, digital communication and hand hygiene were noted between the two periods. More than 72% of children complied with the NPI measures, with the exception of the decrease in number of vulnerable contact(s) indicator (48%). Eating meat more frequently post-lockdown and doing less physical activity during school break post-lockdown were positively associated with increased time spent at home post-lockdown. Furthermore, the odds of compliance, as indicated by the time spent at home post-lockdown were lower with days elapsing from school re-opening, living in smaller town and using antiseptic more frequently pre-lockdown.

**Interpretation:** In this national survey, children showed a high degree of compliance to most NPI measures for the community and primary school settings in Cyprus. The initial NPI measures may have affected the children’s exposome profile in the following months, by altering their diet, physical activity, sedentary lifestyle and hand hygiene habits.

**Funding:** The study was partially funded by the EXPOSOGAS project, H2020 under grant agreement #810995

**Panel: Research in context:** *Evidence before this study:* We searched PubMed for studies published until September 30, 2020 using the search terms: COVID-19, children and lifestyle. Only six peer-reviewed, English-language studies were retrieved on the effect of COVID-19 measures on children’s lifestyle. The impact of non-pharmacological intervention (NPI) measures on children’s health during the pandemic period has been sporadically studied by focusing on a few risk factors at a time without using the exposome’s methodological framework, which is defined as the comprehensive characterization of all environmental exposures during one’s lifetime.

*Added value of this study:* A survey targeted all primary schools of Cyprus to comprehensively study the impact of the initial population-wide NPI measures (lockdown) (March 13-May 4) on the children’s exposome during the school re-opening period (May 21 – June 26). To the best of our knowledge, this is the first study looking at the post-confinement (lockdown) exposome profile changes of children during schools’ re-opening, after the initial population-wide NPI measures of COVID-19 response. The comprehensive and agnostic description of the children’s exposome profile may help to comprehensively account for both known and possibly unknown effects of NPI measures on children’s health and for delineating the children’s degree of compliance to infection prevention and control protocols at school and at home.

*Implications of all the available evidence:* This dataset could inform COVID-19 risk-based public health response strategies targeted for school settings. Future response strategies to epidemic waves shall consider elements of promoting a healthy lifestyle for children at school and at home. Public health policy could ultimately benefit from the inclusion of the human exposome methodological framework and its tools towards the improved identification of susceptible sub-population groups and to facilitate the deployment of site-tailored public health measures; this may be particularly relevant for children and their potential to spread the disease to vulnerable groups.

## 1. Introduction

By October 1, 2020, 1755 cases of coronavirus disease (COVID-19) caused by the severe acute respiratory syndrome coronavirus 2 (SARS-CoV-2) and 22 associated deaths had been reported in Cyprus (1). The first two COVID-19 cases were reported on March 10, 2020 and non-pharmacological interventions (NPI) were implemented to restrict the spread of the virus (1). School closures were among the first measures (March 13) followed by closures of dining and recreation areas (March 16) (2). Stay-at-home orders, also widely referred to as lockdown went in effect on March 24. As the daily reported cases started decreasing, the gradual easing of measures was introduced in early May. Schools re-opened on May 21, following a strict protocol that included among others, a rotation of students with a maximum of 12 allowed in class and a defined break area for each class (3). Recommendations were also made for the implementation of personal measures including limitations in the number of contacts (physical distancing) and reinforcement of personal hygiene habits.

The implemented NPI measures required changes in routine behaviours of primary school children, including those of attending classes face to face, commuting to school, conducting extracurricular activities and meeting with friends. Within the methodological framework of the exposome, these routine behaviours as well as the context in which they take place, the associated environmental exposures, personal and contextual parameters define the exposome profile of children (i.e. the totality of environmental exposures during one’s lifetime) (4,5). Given that routine activities are associated with a suite of exposures and, thus, an exposome profile, compliance to the NPI measures is expected to have led to changes in the exposome profile of primary school children. The comprehensive assessment of the totality of changes in lifestyle and behaviours could be conducted by implementing the methodological framework of the human exposome and through the description of changes in exposome profiles between different time frames.

So far, there have been limited data about the magnitude and extent of possible changes in children’s lifestyle and behaviour profile due to the implemented NPI measures for the COVID-19 pandemic. Studies on changes of children’s lifestyle in Spain, Italy, Canada U.S., and China during the COVID-19 confinement period showed that parameters such as physical activity, screen time, diet as well as sleep time were affected (6–11).

We implemented a national survey to study changes in the children’s exposome upon primary schools re-opening (right after the population-wide lockdown) and compared it against the children’s exposome during the pre-lockdown period in Cyprus. The objectives of this study were: (i) to describe the children’s environment, individual behaviour and lifestyle at school and at home (exposome profiles) before and after the lockdown in Cyprus by adopting the methodological context of the human exposome, and (ii) to assess the children’s compliance to the COVID-19 NPI protocols and recommendations with special focus on personal hygiene and physical distancing at both school and home settings.

## 2. Methods

### 2.1. Study design and population

The Exposome@School | COVID-19 national survey was conducted online. Information about the survey and links to the questionnaire were forwarded to parents of primary school children via primary school’s administrations, during the re-opening of schools following the lockdown in the Republic of Cyprus (June 2020). Public schools and private schools teaching in Greek language (330 and 6, respectively) located in all government-controlled areas of the Republic of Cyprus were contacted, and 253 of them agreed to distribute the survey to their children’s parents. Eligible participants were children attending a primary school.

### 2.2. Data collection and ethics

Data was collected from June 1 until July 17, 2020 using REDCap hosted on a server at the Cyprus University of Technology (12). The survey questionnaire was available in Greek and parents were asked to respond on behalf of the children. The study was approved by the Cyprus National Bioethics Committee (EEBK/EΠ/2020.01.113) and the Ministry of Education, Culture, Youth and Sport (21.11.06.10). Parents were informed that the data collection was anonymous and that, if they wished to, they could withdraw from the study at any time while completing the questionnaire.

The study included questions about the children’s lifestyle and behaviours for two periods (Fig 1): (a) pre-lockdown, i.e. before March 2020 and (b) post-lockdown when schools reopened (May 21-June 26, 2020). The whole set of NPI measures implemented by the Republic of Cyprus during the pandemic period can be found in Figure S1. In this analysis, we included components from all three exposome domains, i.e. the general external domain (socio-economic status), the specific external domain (lifestyle and COVID-19 related parameters) and the internal domain (background characteristics, intrinsic properties, medical history) (Table 1).

**Table 1.** Exposome domains and their specific components/variables included in the Exposome@School|COVID-19 survey. The number of variables is based on the main questions of the questionnaire and not on sub-questions.

| Exposome domain | Study group of components | Variables | # variables |
| --- | --- | --- | --- |
| General external | Socio-economic status | Parents' educational level | 2 |
| Specific external | Lifestyle (children) | Exercise, diet, screen time, digital communication | 34 |
| Specific external | Lifestyle (children) – related with NPI measures | Personal hygiene, number of contacts, time spent at home weekdays and weekend | 22 |
| Specific external | Lifestyle (parents) | Parents' smoking and household cleaning activities frequency | 10 |
| Internal external | Anthropometrics | Weight, height | 2 |
| Internal external | Medical history | Chronic disease, vaccination | 4 |
| Internal external | Background characteristics | Age, sex, place of birth, years living in Cyprus, city of residence, municipality, postal code, school name | 8 |

**Figure 1.**
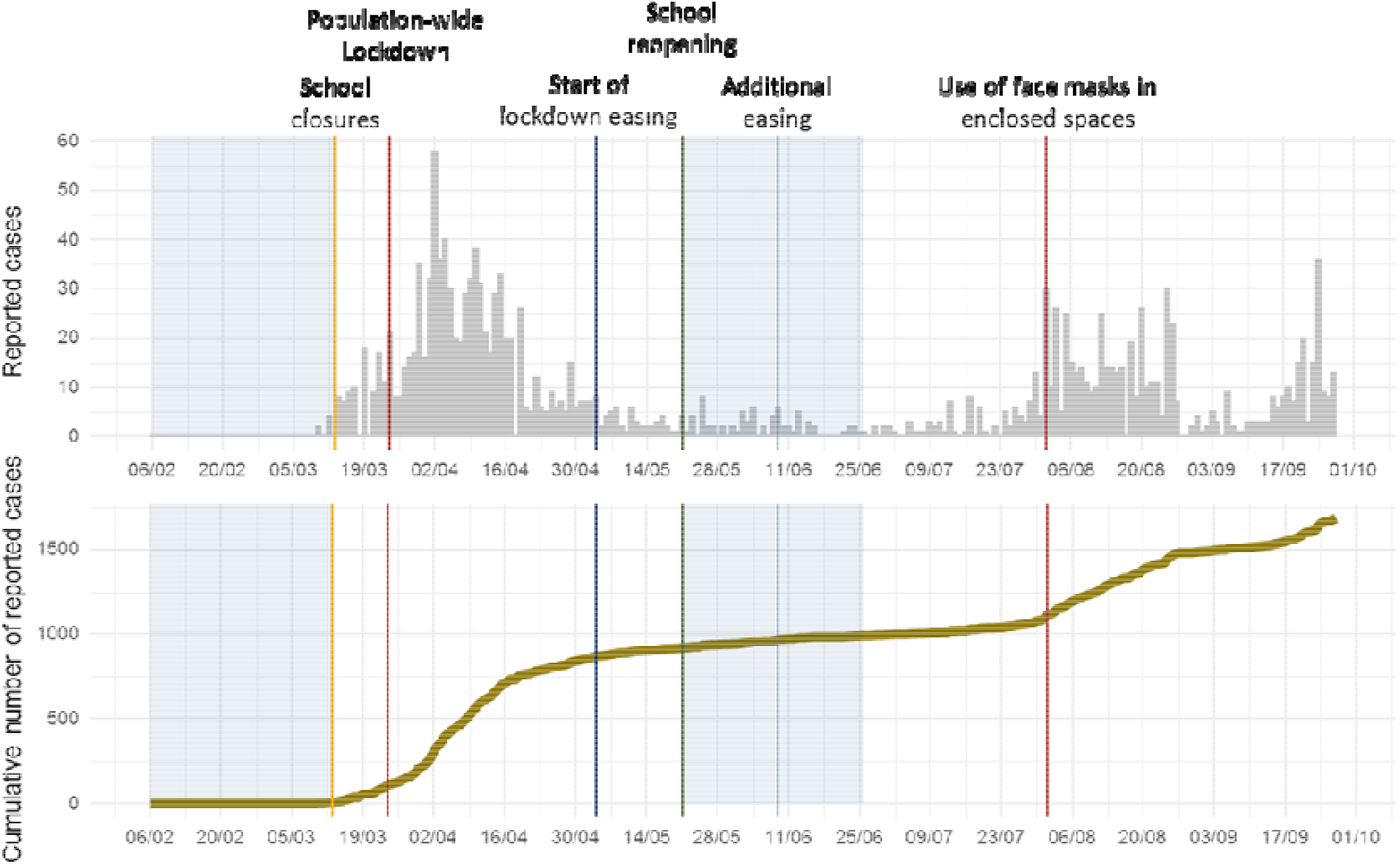
Epidemiological curves for COVID-19 reported cases (top) and cumulative number of cases (bottom) in Cyprus. NPI measures are denoted with vertical lines. The blue shaded areas indicate the two study periods for which primary school children’s lifestyle is described in our study.

More specifically, the online survey consisted of questions related to demographic and general characteristics of children (e.g., age, sex, health status, height, weight, city, returned or not to school) and their parents (educational level) as well as questions about the children’s lifestyle/behaviour (physical activity, diet, digital communication, screen time, personal hygiene, number of contacts and hours spent at home) and parents’ habits (smoking and household cleaning activities) for the two study periods (pre- and post-lockdown). Questions from four validated questionnaires and one diary were used: i) Diary of contacts (13): a contact was defined as either a two-way conversation with three or more words in the physical presence of another person or physical skin-to-skin contact (e.g. handshake, hug and kiss) (13), ii) European Health Interview Survey for 2014 based on the Cyprus Statistical Services (14), iii) European Urban Health Indicators survey (15,16), iv) Questionnaire on hand hygiene and cleaning of home areas (17), and v) Questionnaire on physical activity for children PAQ-C (18–20), where we used 3 out of 9 items included in the original questionnaire, so a spare time activity score could be calculated by taking the mean of all activities (e.g. basketball, cycling, ballet).

### 2.3. Statistical analysis

#### 2.3.1. Description of the children’s exposome profile

Frequencies and percentages were used for the description of categorical variables and means (standard deviations) or medians (interquartile ranges) for the continuous variables, depending on the distribution of the variable. In all categorical variables the “I don’t know/I don’t want to answer” responses were re-coded to “missing”. Variables were compared using the non-parametric Wilcoxon test for continuous characteristics and chi-square test for categorical characteristics. For continuous variables with pre- and post-lockdown values, the percent change was calculated as [(post – pre)/pre)*100] for use in the analysis.

#### 2.3.2. Exposome-wide association study

We performed an exposome-wide association study (ExWAS) to agnostically assess associations of demographic, lifestyle and behavioural parameters with the degree of compliance to COVID-19 recommended measures (Tables S1-S4). In a correlation analysis, all variables were used as continuous with categorical variables being assigned a score. Spearman correlation coefficients were calculated and results visualized with a circo-plot.

Four indicators of compliance to COVID-19 protocols were used as binary outcomes in separate logistic regression models, adjusted for age, sex and parents’ educational level: (i) increase in the hours staying at home during weekdays, (ii) decrease in the number of vulnerable contacts at home during weekdays, (iii) decrease in the number of contacts at school, and (iv) increase of hand washing frequency using antiseptic or soap. People belonging in the vulnerable group were those over 60 years, pregnant, or having a chronic disease. These indicators were set to 0 or 1 based on the responses for the pre-, and post-lockdown period for the number of hours staying at home (1 = post > pre), the number of contacts at school (1 = post < pre), the number of vulnerable contacts at home during weekdays (1 = post < pre) and the hand washing frequency using antiseptic or soap (1 = post>pre and 1 = post > 7 times/day & pre > 7 times/day). For the compliance indicator of decreased number of contacts at school, we used only the data from children who returned to school following the re-opening of schools (n=1331).

Categorical variables were grouped so that they would have a max of 4 levels (2-4 levels) in order to decrease the number of tests in the regression models. The p-values of all model parameters that were used for inference (i.e., excluding the intercept, age, sex and parents’ educational level coefficients) were summarized and adjusted for Benjamini–Hoechberg false discovery rate (FDR). Only parameters with FDR-adjusted p-value 0.05 were considered statistically significant.

All analyses were conducted in R 4.0.2 with RStudio 1.3.1093 (21,22). The input data, scripts, output, and questionnaire are available in the Supplementary Material.

## 3. Results

### 3.1. Study population characteristics

A total of 1509 primary school children from more than 180 schools all over Cyprus were included in the survey. There were 2708 entries who weren’t complete and they weren’t included in the analysis (parents who visited the questionnaire link but didn’t complete the survey). Participating children had a mean age of 9.6 years (SD: 1.7), a balanced sex distribution (48% females), and the majority of them were born in Cyprus (96%) (Table 2). The reported parents’ educational level was relatively high with about half of them holding at least one university degree (57% of mothers and 44% of fathers). Most parents completed the survey for one child (97%), 42 parents for two children (2.9%) and 1 for three children (0.1%).

**Table 2.**
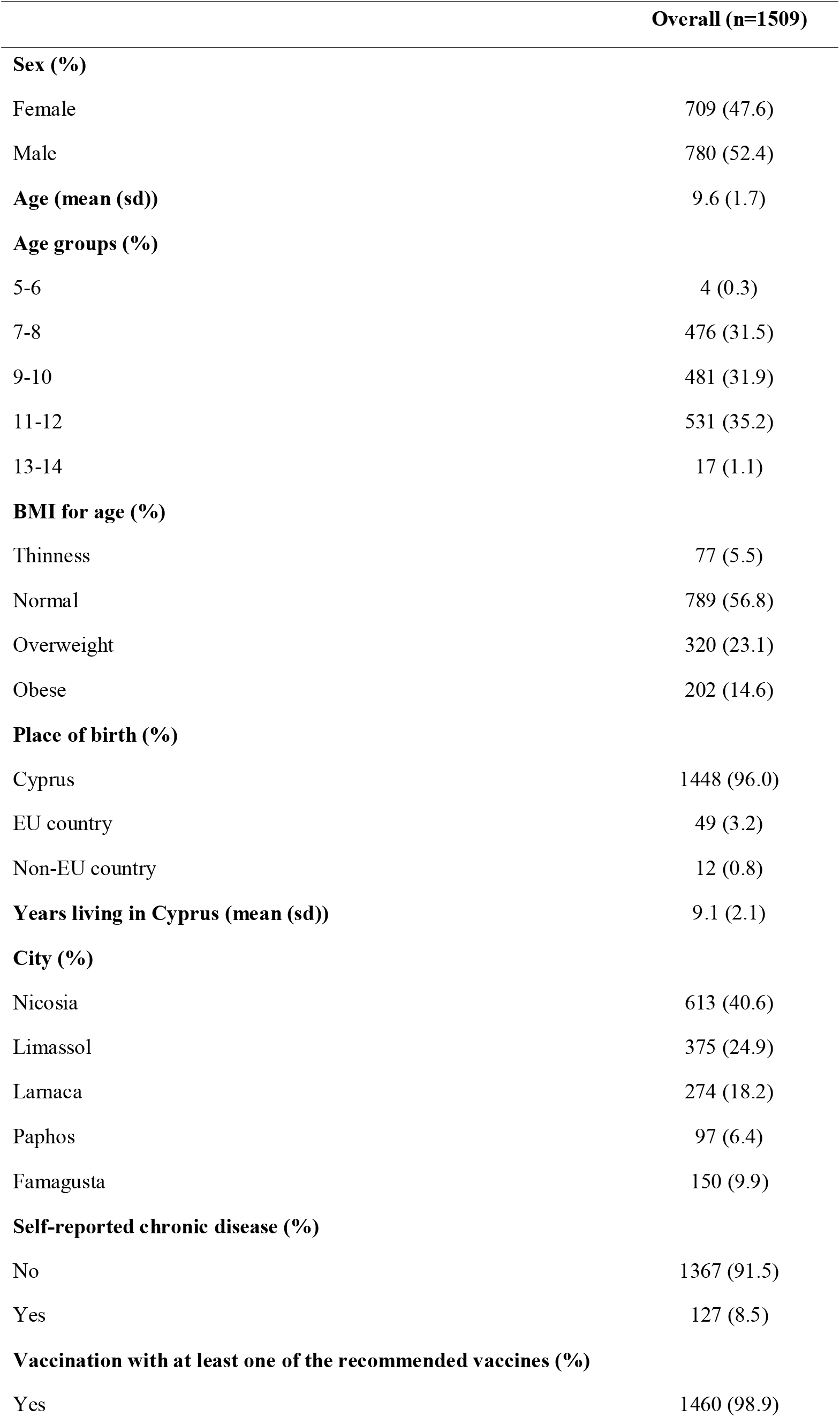
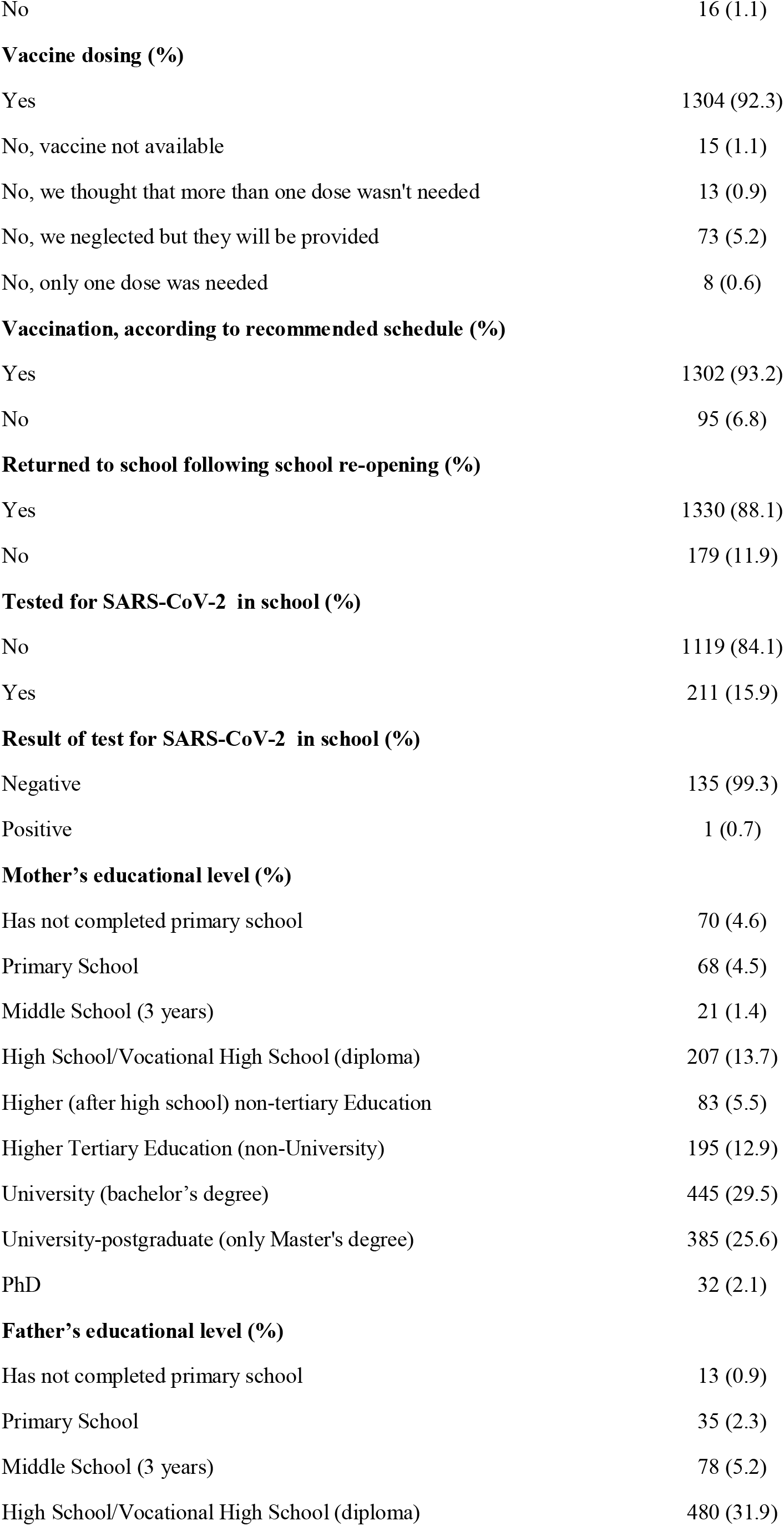
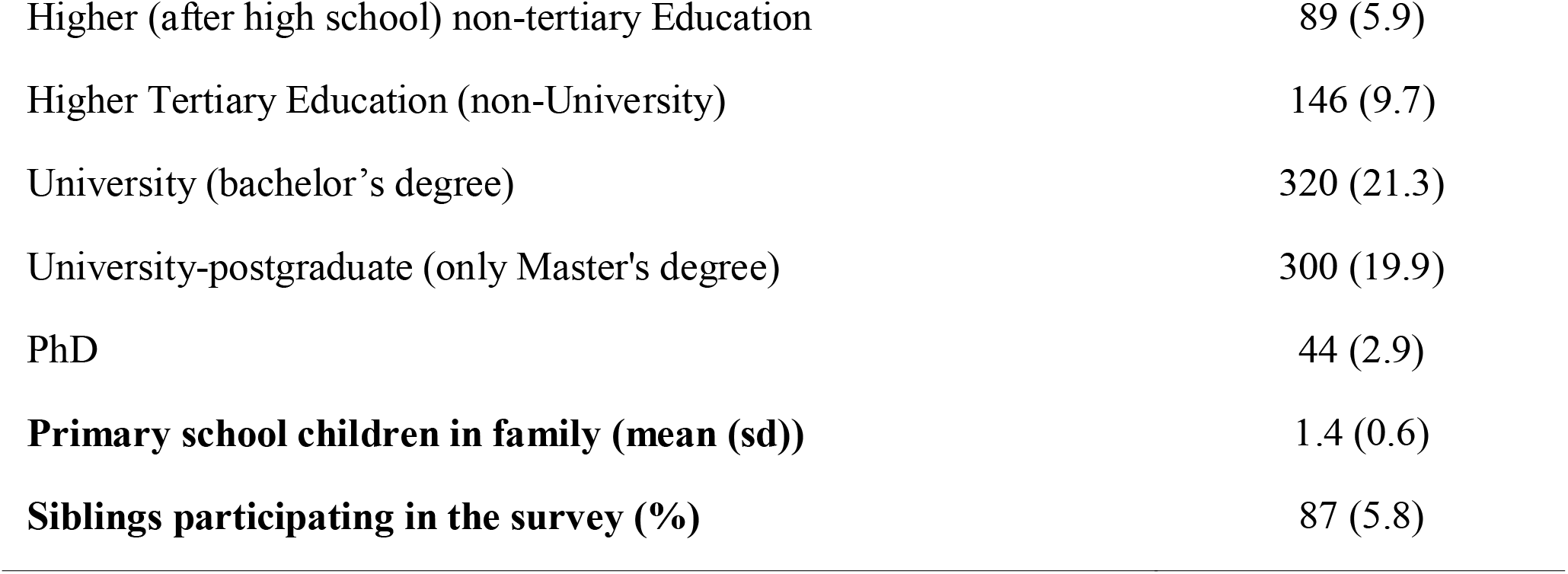
Demographics and general characteristics of participating children

|  | **Overall (n=1509)** |
| --- | --- |
| **Sex (%)** |  |
| Female | 709 (47.6) |
| Male | 780 (52.4) |
| **Age (mean (sd))** | 9.6 (1.7) |
| **Age groups (%)** |  |
| 5-6 | 4 (0.3) |
| 7-8 | 476 (31.5) |
| 9-10 | 481 (31.9) |
| 11-12 | 531 (35.2) |
| 13-14 | 17 (1.1) |
| **BMI for age (%)** |  |
| Thinness | 77 (5.5) |
| Normal | 789 (56.8) |
| Overweight | 320 (23.1) |
| Obese | 202 (14.6) |
| **Place of birth (%)** |  |
| Cyprus | 1448 (96.0) |
| EU country | 49 (3.2) |
| Non-EU country | 12 (0.8) |
| **Years living in Cyprus (mean (sd))** | 9.1 (2.1) |
| **City (%)** |  |
| Nicosia | 613 (40.6) |
| Limassol | 375 (24.9) |
| Larnaca | 274 (18.2) |
| Paphos | 97 (6.4) |
| Famagusta | 150 (9.9) |
| **Self-reported chronic disease (%)** |  |
| No | 1367 (91.5) |
| Yes | 127 (8.5) |
| **Vaccination with at least one of the recommended vaccines (%)** |  |
| Yes | 1460 (98.9) |
| No | 16 (1.1) |
| **Vaccine dosing (%)** |  |
| Yes | 1304 (92.3) |
| No, vaccine not available | 15 (1.1) |
| No, we thought that more than one dose wasn't needed | 13 (0.9) |
| No, we neglected but they will be provided | 73 (5.2) |
| No, only one dose was needed | 8 (0.6) |
| **Vaccination, according to recommended schedule (%)** |  |
| Yes | 1302 (93.2) |
| No | 95 (6.8) |
| **Returned to school following school re-opening (%)** |  |
| Yes | 1330 (88.1) |
| No | 179 (11.9) |
| **Tested for SARS-CoV-2 in school (%)** |  |
| No | 1119 (84.1) |
| Yes | 211 (15.9) |
| **Result of test for SARS-CoV-2 in school (%)** |  |
| Negative | 135 (99.3) |
| Positive | 1 (0.7) |
| **Mother's educational level (%)** |  |
| Has not completed primary school | 70 (4.6) |
| Primary School | 68 (4.5) |
| Middle School (3 years) | 21 (1.4) |
| High School/Vocational High School (diploma) | 207 (13.7) |
| Higher (after high school) non-tertiary Education | 83 (5.5) |
| Higher Tertiary Education (non-University) | 195 (12.9) |
| University (bachelor's degree) | 445 (29.5) |
| University-postgraduate (only Master's degree) | 385 (25.6) |
| PhD | 32 (2.1) |
| **Father's educational level (%)** |  |
| Has not completed primary school | 13 (0.9) |
| Primary School | 35 (2.3) |
| Middle School (3 years) | 78 (5.2) |
| High School/Vocational High School (diploma) | 480 (31.9) |
| Higher (after high school) non-tertiary Education | 89 (5.9) |
| Higher Tertiary Education (non-University) | 146 (9.7) |
| University (bachelor's degree) | 320 (21.3) |
| University-postgraduate (only Master's degree) | 300 (19.9) |
| PhD | 44 (2.9) |
| **Primary school children in family (mean (sd))** | **1.4 (0.6)** |
| **Siblings participating in the survey (%)** | **87 (5.8)** |

Most children’s health status was reported as healthy (92%) while for 8%, a chronic disease was reported by their parents; the most frequently reported diseases were allergies (34%) and asthma (33%) (Table S5). The majority of children were vaccinated according to the recommended vaccination schedule and received all recommended vaccine doses (93% and 92%, respectively). About half of the children had a normal BMI for their age (57%), while 23% were overweight and 15% were obese. We observed higher obesity levels for boys compared to girls (18% and 10%, respectively, Table S6).

A small portion of children did not go to school following the re-opening of schools on May 21 (12%). Out of the 1331 children who returned to school, 211 (16%) were tested for SARS-CoV-2 with a diagnostic test (PCR) in random sampling taking place in school premises; this testing campaign was initiated by the Cyprus Ministry of Health and only one child was tested positive (0.5%).

### 3.2. Children’s exposome profile in the pre- and post-lockdown periods

#### 3.2.1. Diet

Overall, children didn’t change their eating habits between the pre- and post-lockdown periods, with the exception of the consumption of sugary food items, breakfast and ready-made food. In effect, about half of the children consumed fruits daily (54% for both pre- and post-lockdown periods), whereas a third of them consumed vegetables daily (34% and 31%, pre- and post-lockdown, respectively) (Figure S2). The weekly meat consumption frequency was relatively high, with half of the children eating meat 2-3 times per week and about one third of them, eating meat 4-6 times per week in both study periods (Figure S3). Regarding legumes consumption, half of the children consumed legumes 2-3 times per week and one third of them, once a week, while the fish consumption frequency was lower, with about half of children eating fish once a week (46% for both periods) (Figure S3). About one third of the children ate snacks (such as crisps, bakery items, salted nuts) 2-3 times per week and about one quarter of them, once a week in both the pre- and post-lockdown periods (Figure S4).

The consumption of food items that contained sugar was high in both periods, but even higher in the post-lockdown period with 37% and 26% of the children eating sugary items every day and 4-6 times per week, respectively, in the post-lockdown period, when compared to 33% and 19% for the pre-lockdown period (*p<0*.*001*) (Figure S4). Similarly, the proportion of children eating breakfast daily increased in the post-lockdown period (80% compared to 76%, *p=0*.*002*) (Figure S5). On the other hand, following the re-opening of schools, children ate ready-made food less frequently with 20% of children never consuming ready-made food in the post-lockdown period, compared to 6% in the pre-lockdown period (*p<0*.*001*) (Figure S6).

#### 3.2.2. Physical activity

Following the re-opening of schools, children’s physical activity decreased both in school and in their free time compared to the period before lockdown (p<0.001). The spare time activity score, which is based on the weekly frequency of activities children engage with during their free-time, was significantly reduced in the post-lockdown period (median [Q1, Q3]: 1.38 [1.25, 1.62]) vs 1.5 [1.38, 1.75] in the pre-lockdown period, suggesting that the children’s physical activity was reduced following the lockdown (Figure S7). About half of the children engaged with activities that involved little physical effort during their free time (51%) and were sitting down during school break (46%) in the post-lockdown period compared to 29% and 9%, respectively for the pre-lockdown period (Figures S8-S9).

#### 3.2.3. Digital communication and screen time

Daily communication of children with friends and family using telephone or internet increased in the post-lockdown period (46% and 45%, respectively compared to 32% and 34% for the pre-lockdown period, *p<0*.*001*) (Figure S10), possibly because of a decrease in face-to-face gatherings. Screen time in the form of watching TV or using a computer, mobile phone, tablet, and game console also increased in the post-lockdown period (*p<0*.*001*), with screen time being 1-3 hours/day for 62% of children, and 4-7 hours/day for 25% of children (Figure S11).

#### 3.2.4. Personal hygiene habits

Children increased the frequency of hand hygiene with antiseptic products and hand washing with soap in the post-lockdown period, with about half of the children washing their hands with soap 4-7 times/day (51% vs 30% in the pre-lockdown period) and using hand antiseptic 1-3 times/day (44% vs 19% in the pre-lockdown period, *p<0*.*001*) (Figure S12).

#### 3.2.5. Number of contacts and number of hours staying at home

Children decreased the number of physical contacts during school re-opening after the population-wide lockdown (p<0.001) (Figure 2). Specifically, the median number of contacts during weekdays was about halved in school, home and elsewhere (10, 4 and 4, respectively, when compared to 24, 6 and 10, in the pre-lockdown period) (Figure S13). Similar decreases in contacts were observed during weekends (Figure S13). Children of all ages (5-14 years) decreased their average number of contacts in all settings during both weekends and weekdays, with children of younger age (6 years old) having a low number of vulnerable and total contacts at home in both pre- and post-lockdown periods (Figure 2).

**Figure 2.**
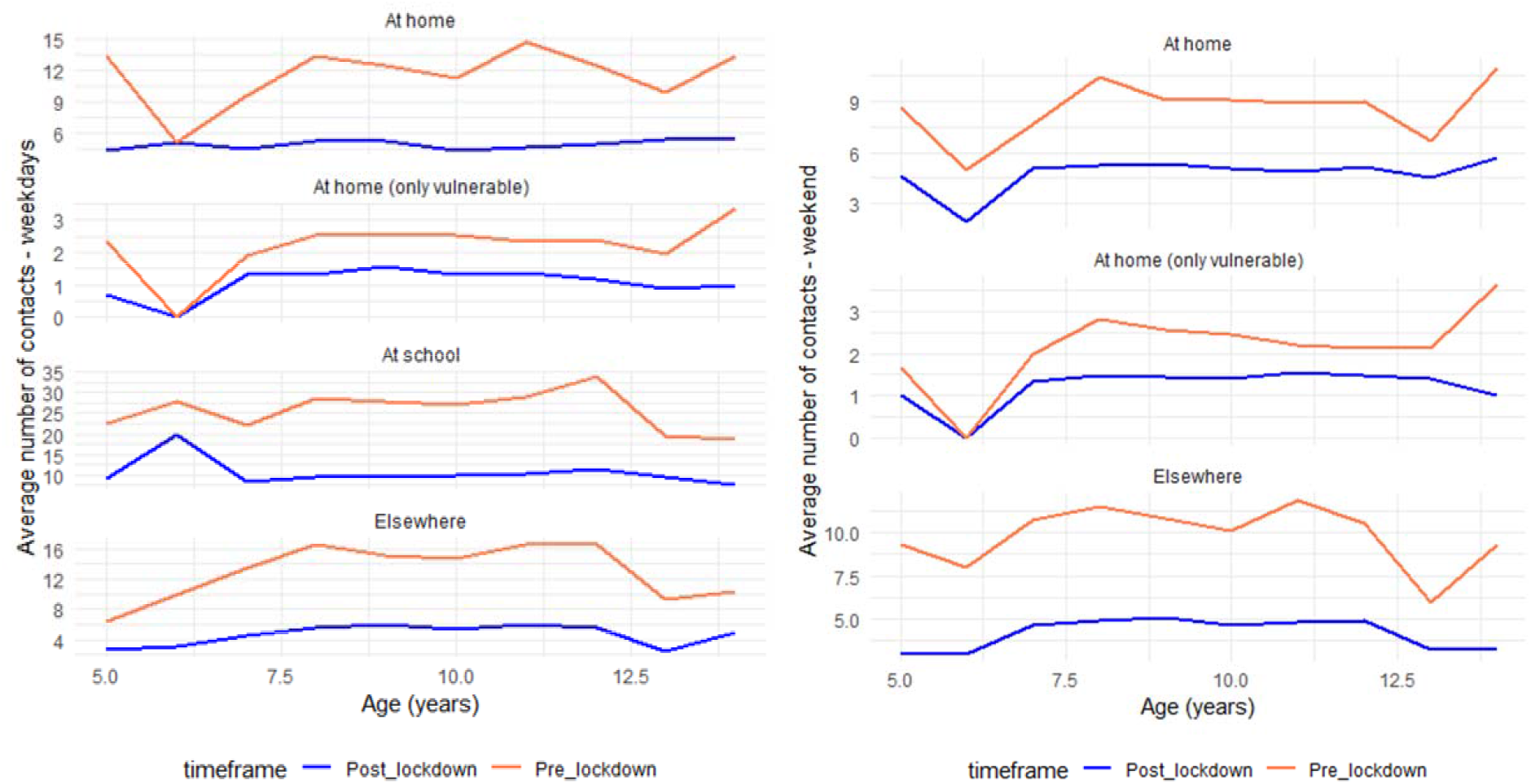
Average number of contacts at home (separately for vulnerable contacts), at school and elsewhere by age for the 2 periods: (a) following the re-opening of schools (post-lockdown) and (b) before the lockdown (pre-lockdown) during weekdays and weekends.

Significant (*p<0*.*001*) decreases were also observed in the number of vulnerable group contacts at home, for both weekends and weekdays (median [Q1, Q3]: 2[1, 3] in the pre-lockdown period and 1[0, 2] in the post-lockdown period) (Figures S13-S14). It is important to note that, before the lockdown, the percentage of children who were coming in contact at their home with at least one person belonging to the vulnerable groups (i.e., those > 60 years old, pregnant, or presence of chronic diseases) was higher compared to the period following the lockdown, for both weekends and weekdays (Table S7).

The median change (%) in the number of contacts during weekdays was negative for contacts at home, school and elsewhere, but it was zero for vulnerable contacts at home (−30 [−67, 0], −60 [−76, −47], −60 [−81, −30] and 0 [−60, 0], respectively) (Figure S15). Similar changes were observed for contacts during weekends (Figure S16).

Similarly to the decrease in contacts, children stayed more hours at home following the re-opening of schools, both in weekends and weekdays (median [Q1, Q3]: 20 [12, 24] and 15 [10, 20], respectively, when compared with 15 [10, 19] and 12 [7, 15] in the pre-lockdown period) (Figure S17). The median change (%) in the hours staying at home was higher for weekdays compared to weekends (31 [0, 67] and 20 [0. 60], respectively) (Figure S18).

#### 3.2.6. Parents’ smoking habits and household cleaning activities frequency

Parents’ smoking habits didn’t change between the two study periods with most parents not smoking in the house (about 70%) and about 26% of them smoking in the house daily (median [Q1, Q3]: 10 [5, 15] cigarettes per day) in both periods (Figure S19). As parents reported, the frequency of all household cleaning activities with the use of disinfectants or antiseptic products increased in the post-lockdown period, with the majority of parents cleaning the kitchen 7 times/week (72% compared to 51% in the pre-lockdown period); about half of them reported cleaning the bathroom 7 times/week (43% compared to 29%) and about one third of them mopping and cleaning other surfaces 7 times/week (33% and 32%, compared to 21% and 22%, accordingly) (Figure S20).

### 3.3. Exposome-wide associations (ExWAS)

#### 3.3.1. Correlations among exposome variables

A circo plot for all exposome variables adjusted for age, sex, mother’s and father’s educational level did not show any unexpected patterns of correlation among the variables (Fig. 3). Notable positive correlations (r ≥ 0.7) were observed for dietary variables (fish, vegetables, sugar, fruits, meat, chips and legumes) among the pre- and post-lockdown study periods (Table S8).

**Figure 3.**
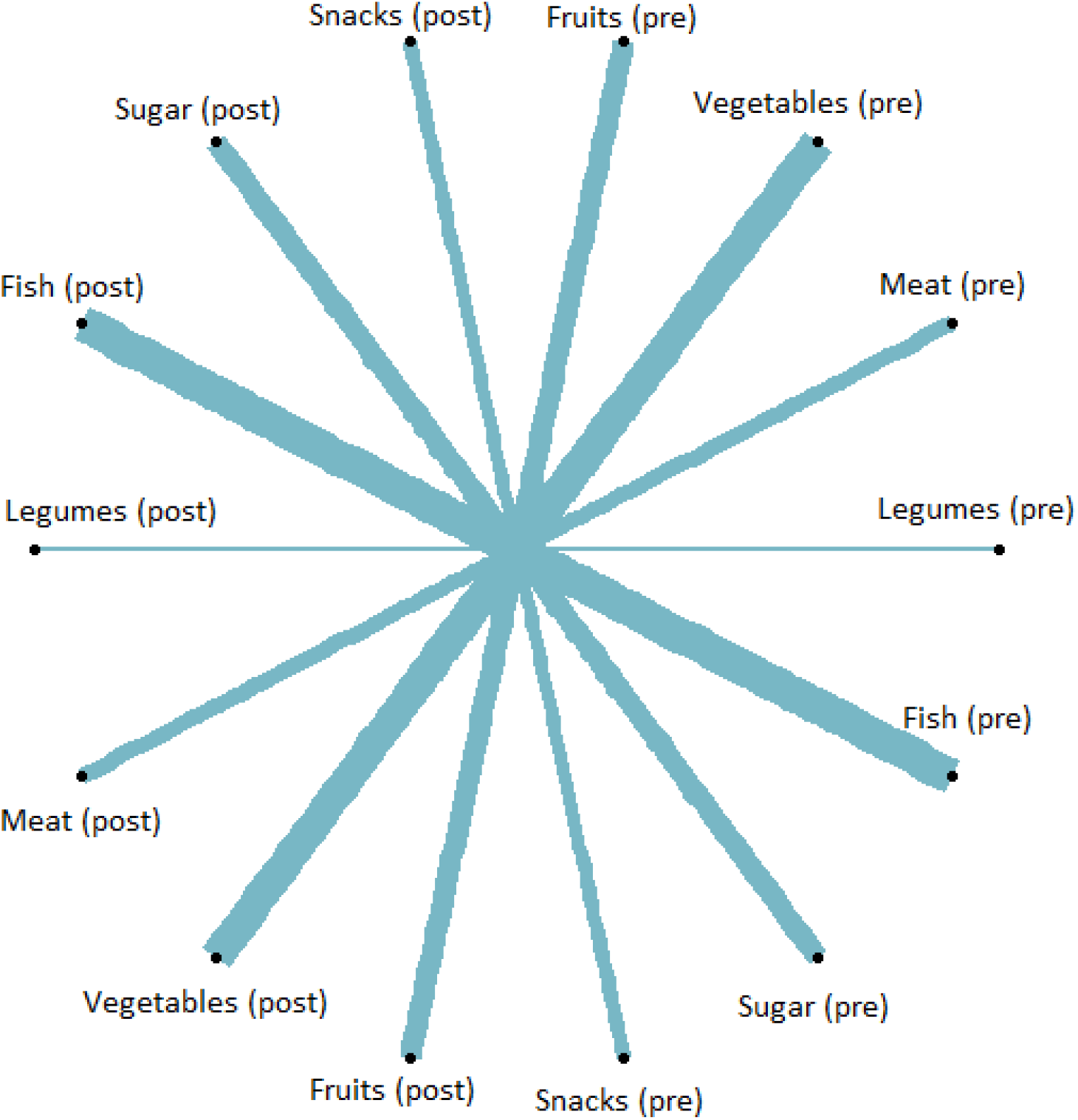
Circo plot for exposome variables adjusted for age, sex, mother’s and father’s educational level. Only correlations with |r| ≥ 0.7 are shown. Only positive correlations were observed for |r| ≥ 0.7. “Post” refers to the post-lockdown period and “pre” to the pre-lockdown period. The width of the line indicates the level of correlation (larger width indicates higher r).

#### 3.3.2. Assessment of children’s compliance to COVID-19 recommendations and hygiene protocols

Based on three out of four compliance indicators, the children’s compliance to COVID-19 measures following the lockdown was relatively high (73-85%), with the exception of the indicator of reduced number of contacts belonging in the vulnerable groups at home during weekdays, for which only about half of the children (48%) decreased the number of vulnerable contacts at home (Table 3). No significant differences were observed for the compliance indicators between boys and girls.

**Table 3.** Summary tables for compliance indicators

| **Outcome** | **Overall**<br>**N (%)** | **Males**<br>**N (%)** | **Females**<br>**N (%)** |
| --- | --- | --- | --- |
| Increase in hours staying at home (n=1509) | 1096 (72.9) | 562 (72.4) | 515 (72.9) |
| Decrease in number of vulnerable contacts at home<br>- weekdays (n=1190) | 573 (48.2) | 308 (48.4) | 257 (48.2) |
| Decrease in number of contacts at school (n=1330) | 1100 (85.1) | 569 (85.1) | 516 (84.9) |
| Increase in hand washing using soap or antiseptic<br>(n=1509) | 1099 (74.4) | 549 (71.5) | 532 (77.2) |

#### 3.3.3. ExWAS associations with indicators of compliance to COVID-19 protocols

In the ExWAS models of compliance to COVID-19 measures for which the outcome was the increase in hours staying at home during weekdays, six parameters had an FDR-corrected p-value < 0.05. In effect, sitting, standing or walking during the school break, consuming meat 4-7 times/week, and not returning to school in the post-lockdown period were positively associated with increased time spent at home. Living in Paphos (smaller town compared to the capital of Nicosia), number of days elapsed from school re-opening and use of hand antiseptic ≥ 4 times/day in the pre-lockdown period were negatively associated with this compliance indicator (Fig 4).

**Figure 4.**
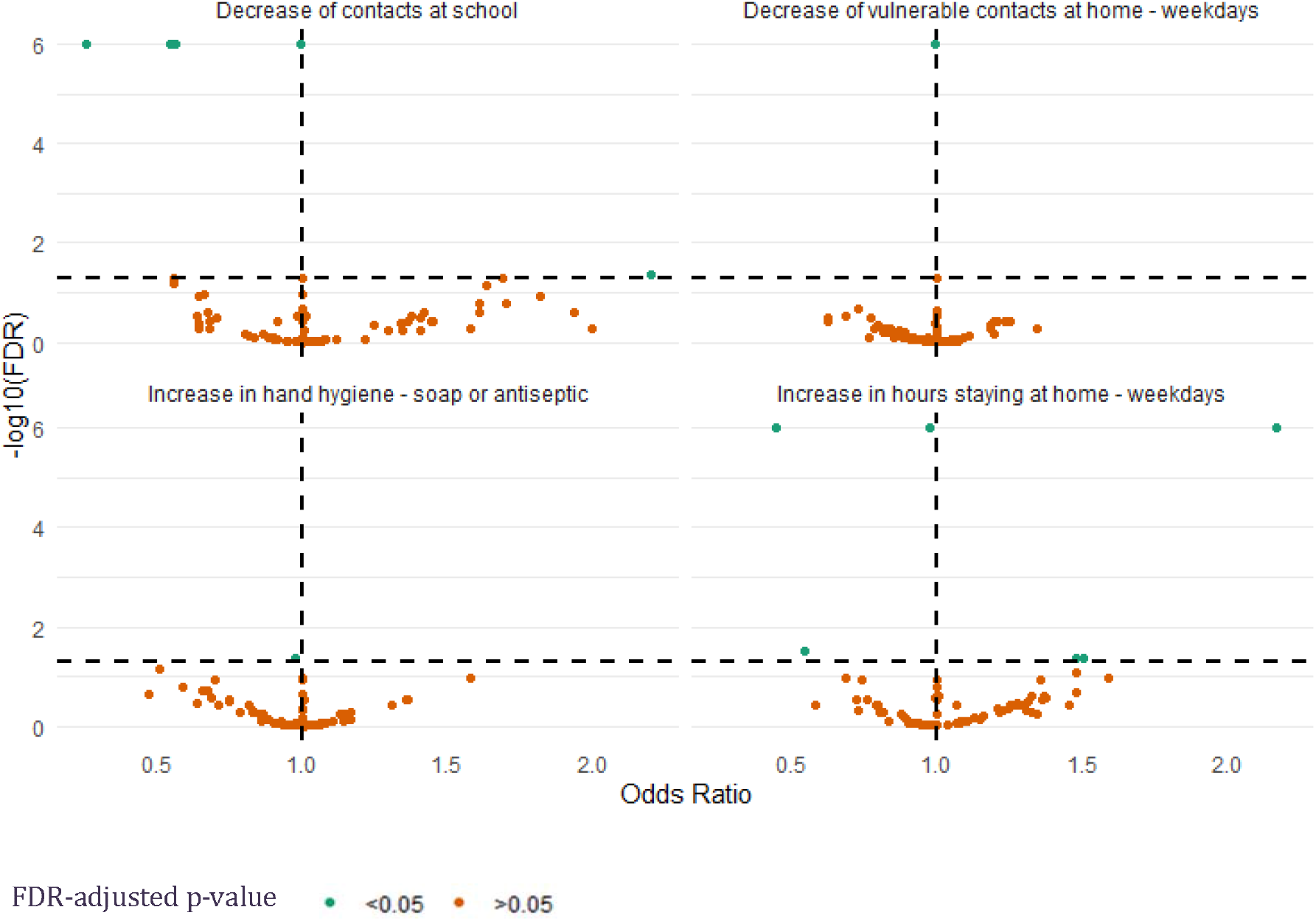
Odds ratios vs −log10 (FDR-adjusted p-value) for ExWAS models of compliance indicators. Compliance indicators from top left to bottom right: Decrease in the number of contacts at school, decrease in the number of vulnerable contacts at home during weekdays, increase in hand hygiene using soap or antiseptic and increase in hours staying at home during weekdays.

The percent change in the total number of contacts at home during weekdays and of vulnerable contacts at home during weekends were the negatively associated with the indicator of reduced number of vulnerable contacts at home during weekdays (FDR-adjusted p-value < 0.05).

The only variable positively associated with the compliance indicator of reduced number of contacts at school was never communicating with friends in the pre-lockdown period, whereas < 4 times/week sugar consumption before and after the lockdown, use of hand antiseptic ≥ 4 times/day in the pre-lockdown period and change (%) in the number of contacts outside of home and school were negatively associated with this compliance indicator (FDR-adjusted p-value < 0.05). The days elapsed since school re-opening that survey submission took place was the only predictor that was negatively associated with the compliance indicator of increasing hand hygiene using soap or hand antiseptic. All significant parameters with FDR-adjusted p-value ≤ 0.05 can be found in Table S9.

## 4. Discussion

The pandemic and its impact on lifestyle and behaviours can be better understood using comprehensive frameworks such as the one of the human exposome which has been previously used in the assessment of chronic health outcomes due to environmental exposures (23). In this national survey, we employed the exposome concept and its tools to evaluate the impact of the population-wide lockdown and other restrictive measures on lifestyle and behaviours of children. A representative sample was obtained from the majority of schools located in the territories of the Republic of Cyprus. The study also described children’s compliance to post-lockdown NPI measures and health protocols during school re-openings period (May-June 2020). The impact of the COVID-19 pandemic and its associated NPI measures (lockdown, school closures) on children’s exposome profile was evident by assessing changes in relevant exposome parameters, such as physical activity, diet, digital communication, screen time, personal hygiene habits, number of contacts and hours staying at home as well as their parents’ frequency of household cleaning activities. Specifically, the changes observed in the children’s exposome (behaviours and living environment) within a few months after the lockdown compared to the period before the introduction of NPI measures in March were the following: reduced physical activity, increased consumption of sugary food items, decreased consumption of ready-made food, increased frequency of screen time and digital communication with friends and family, decreased number of contacts at home, school and elsewhere, increased time spent at home, increased frequency of hand washing and antiseptic use and higher frequency of passive exposure to cleaning products through the increased frequency of household cleaning activities (24). Several of these changes in exposome parameters were observed in factors/parameters already linked with population health trends such as increasing obesity (25) and include inadequate daily consumption of sugary items and sedentary lifestyle including lower physical activity and higher screen time.

The degree of children’s compliance to NPI measures (assessed with either increasing time spent at home, or decreasing number of contacts at school, or decreasing number of vulnerable contacts at home, or increasing frequency of personal hygiene) was associated with certain lifestyle habits. These habits/behaviours were: higher consumption of sugar and meat, or not doing a lot of physical activities during school time in the post-lockdown period, or using less hand antiseptics and never communicating with friends in the pre-lockdown period. Such habits and lifestyle changes might be presumably driven by recommendations to stay at home and reduce contacts at school, and limiting face-to-face extracurricular activities. On the other hand, living in the smaller city of Paphos (with population of about 36000) was associated with a lower degree of compliance when compared with children living in Nicosia (capital city of Cyprus). This lower degree of compliance could be attributed to easier access to green/blue space and more social relationships for smaller in population size urban settings. Moreover, as days from the re-opening of schools elapsed, the degree of compliance was decreasing, presumably due to gradual relaxation of measures or lower adherence to the measures. Reducing contact with vulnerable individuals at home was the metric that children followed the least among all compliance indicators. This may be important to consider in future public health response measures for better protecting vulnerable population groups during epidemics.

Other studies showed that the COVID-19 confinement may have affected specific parameters in children’s lifestyle e.g. decreased physical activity and increased sedentary behaviour (6–11), increased sleep time (7,9) and diet changes (7,10). These studies were mainly focused on the period during COVID-19 strict restrictions (lockdown and school closures), whereas our study describes the children’s lifestyle in the period following the gradual relaxation of measures and re-opening of schools in Cyprus, using the exposome approach.

It is necessary to examine whether these exposome changes persist in the longer-term and to assess the impact of children’s sedentary behaviour and sugary food consumption on their later in life odds of developing chronic diseases. The fact that the majority of children conformed to a sedentary lifestyle in combination with the higher consumption of sugar following the re-opening of schools needs to be reassessed in future prospective studies and should inform future policy making of promoting healthy lifestyle for children as part of a comprehensive response to epidemic waves. The use of exposome tools will be important to identify susceptible sub-population groups and to facilitate the deployment of site-tailored public health measures (26) promoting a healthier lifestyle during periods in which physical distancing is necessary.

Strengths of this study are its relatively large sample size, being representative of the Cyprus children’s population. This is also the first study demonstrating the application of the exposome’s concept to a realistic COVID-19 pandemic setting where comprehensive understanding of primary school children’s behaviours took place. Limitations of this study include the possible recall bias of the participating parents, the cross-sectional study design and the fact that parents completed the questionnaire on behalf of their children.

Overall, our results on the description of the profile of children following and before the lockdown are important as they can be used as a reference for future exposome studies in primary school children. Detailed and agnostic description of the children’s exposome helps in weighing in known and possibly unknown effects of NPI measures, and thereby better informing decision making for pandemic response. Future studies could further benefit from broadening the number and type of exposome variables considered and by including biospecimen in their study design so that children’s internal exposome may be better captured (biomarkers of effect, -omics platforms).

The NPI measures implemented were deemed necessary to reduce the SARS-CoV-2 transmission. However, as our results show, they had unintended consequences, such as modifying the exposome profile of primary school children and potentially altering their later in life chronic disease risk. Hence, it is important that the impacts of any NPI measure is considered in its totality prior to its implementation and strategies are developed to minimize any risk and avoid any harm. The study’s findings could inform health policy guidelines towards decreasing the negative impacts of a future pandemic and its associated NPI measures.

## Supporting information

SI section

Deidentified data and script

## Data Availability

Deidentified data is publicly available with this submission

## Acknowledgments

Makris K.C. acknowledges the partial funding support by the EXPOSOGAS project, H2020 research and innovation programme under grant agreement #810995

## Tables and Figures – Main manuscript

### Checklist for Reporting Results of Internet E-Surveys (CHERRIES)

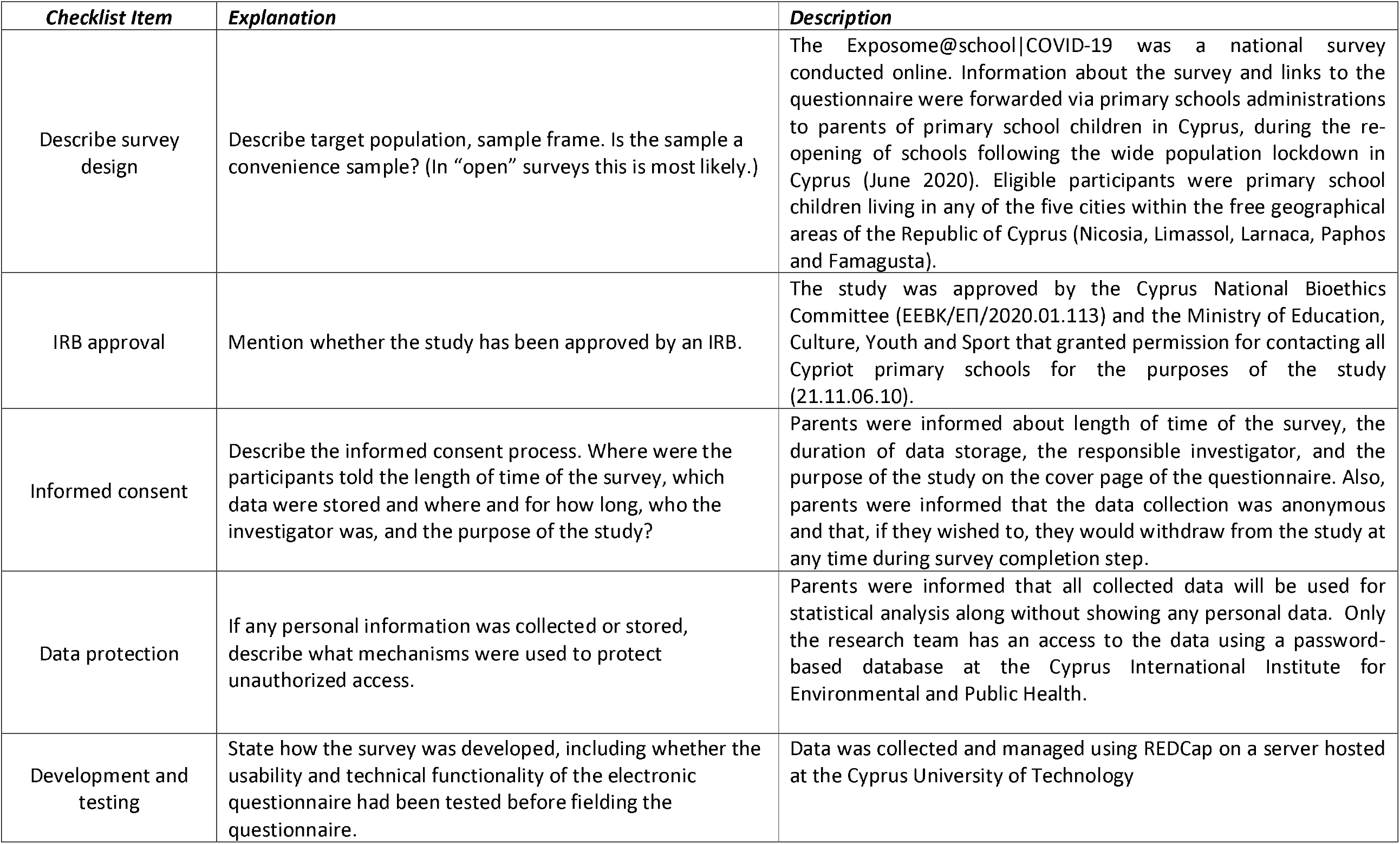
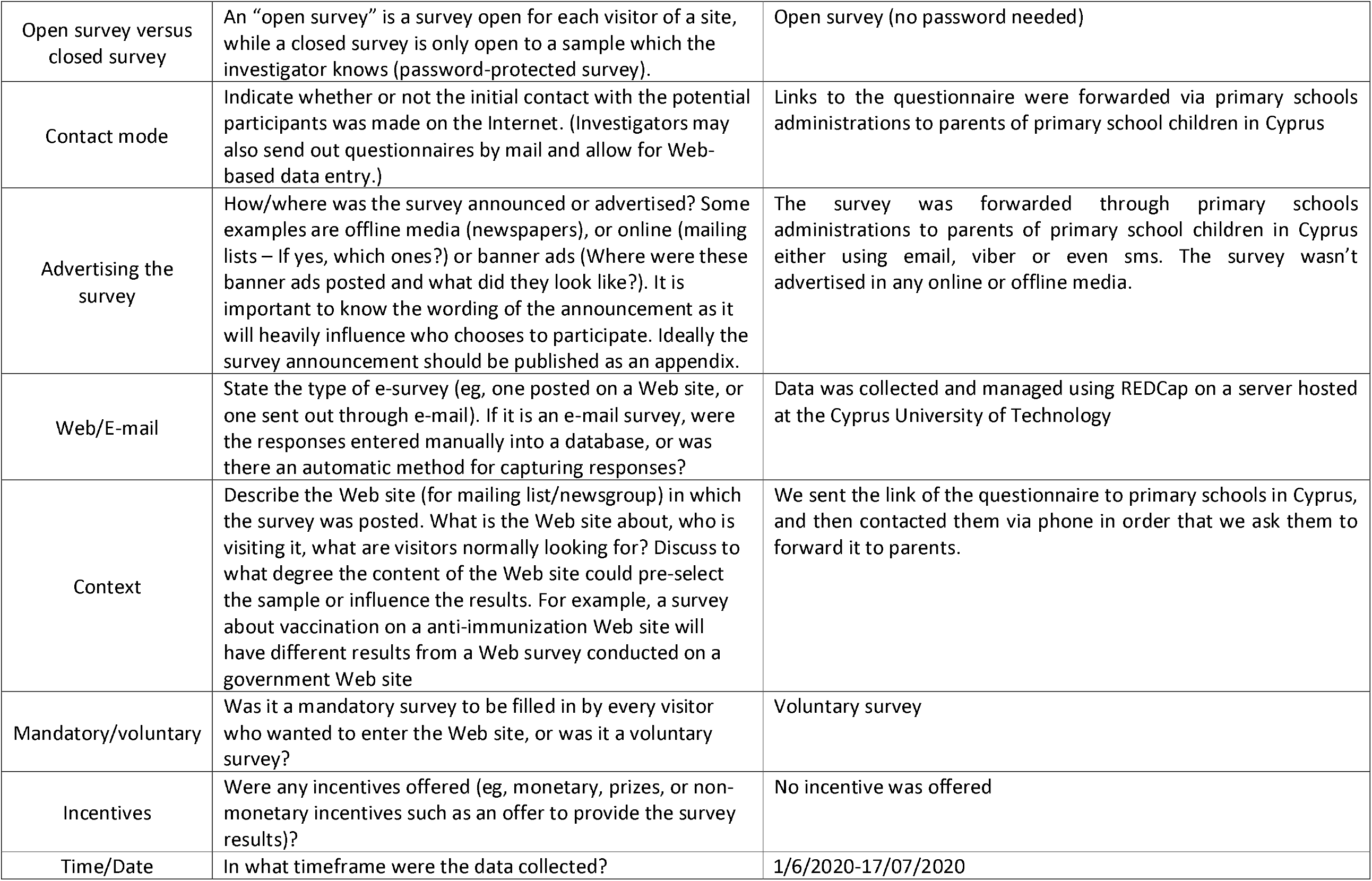
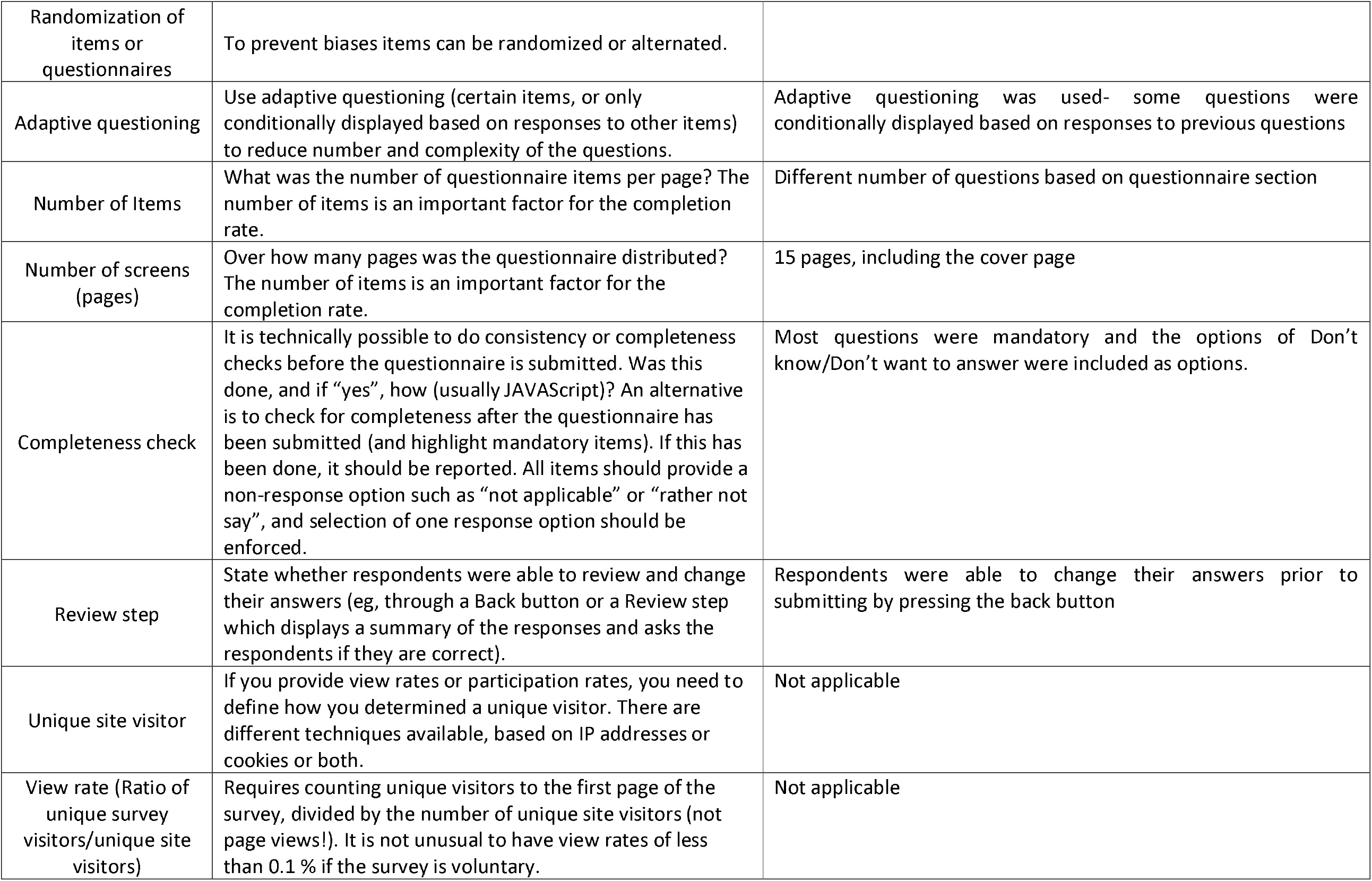
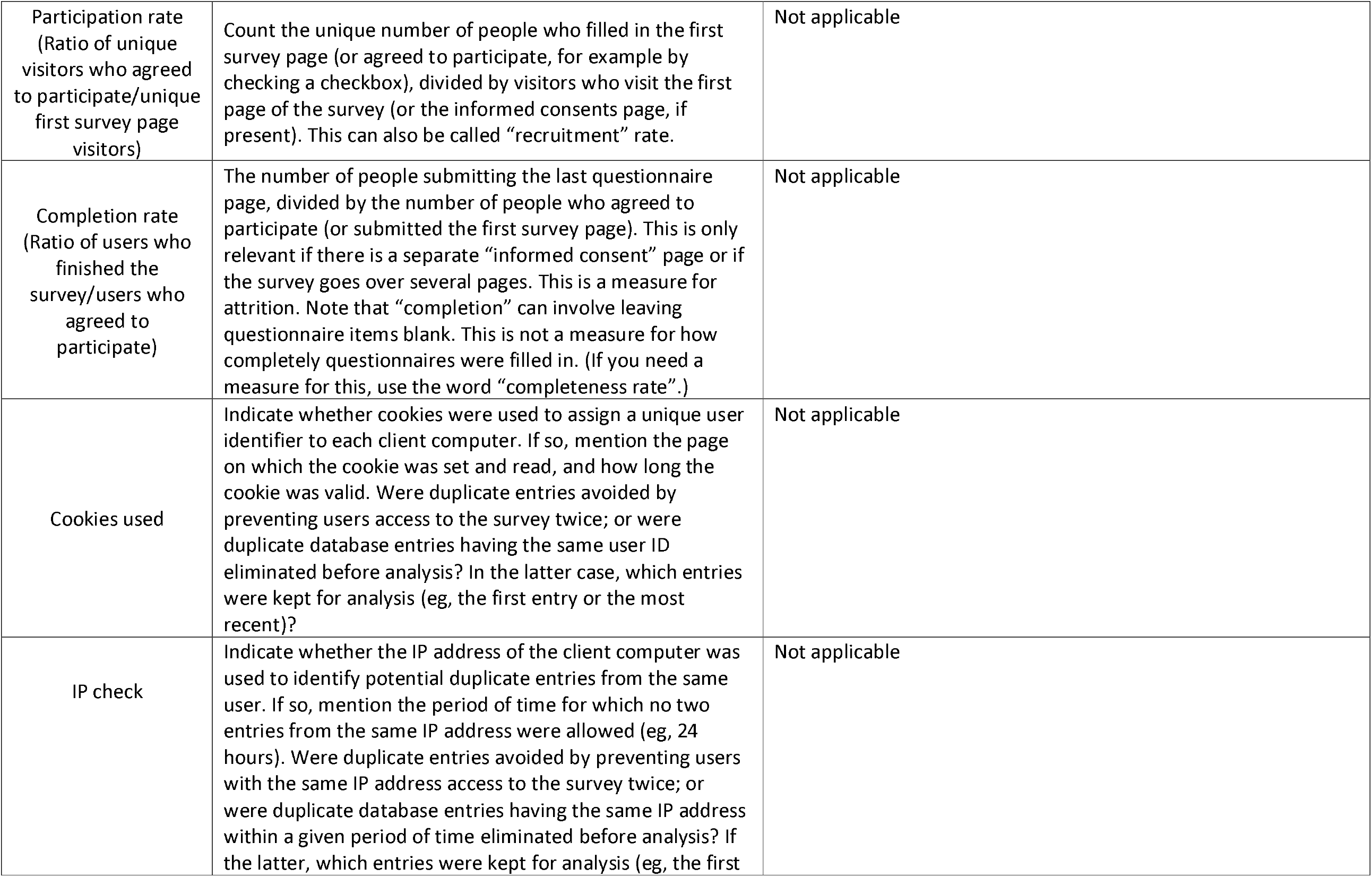
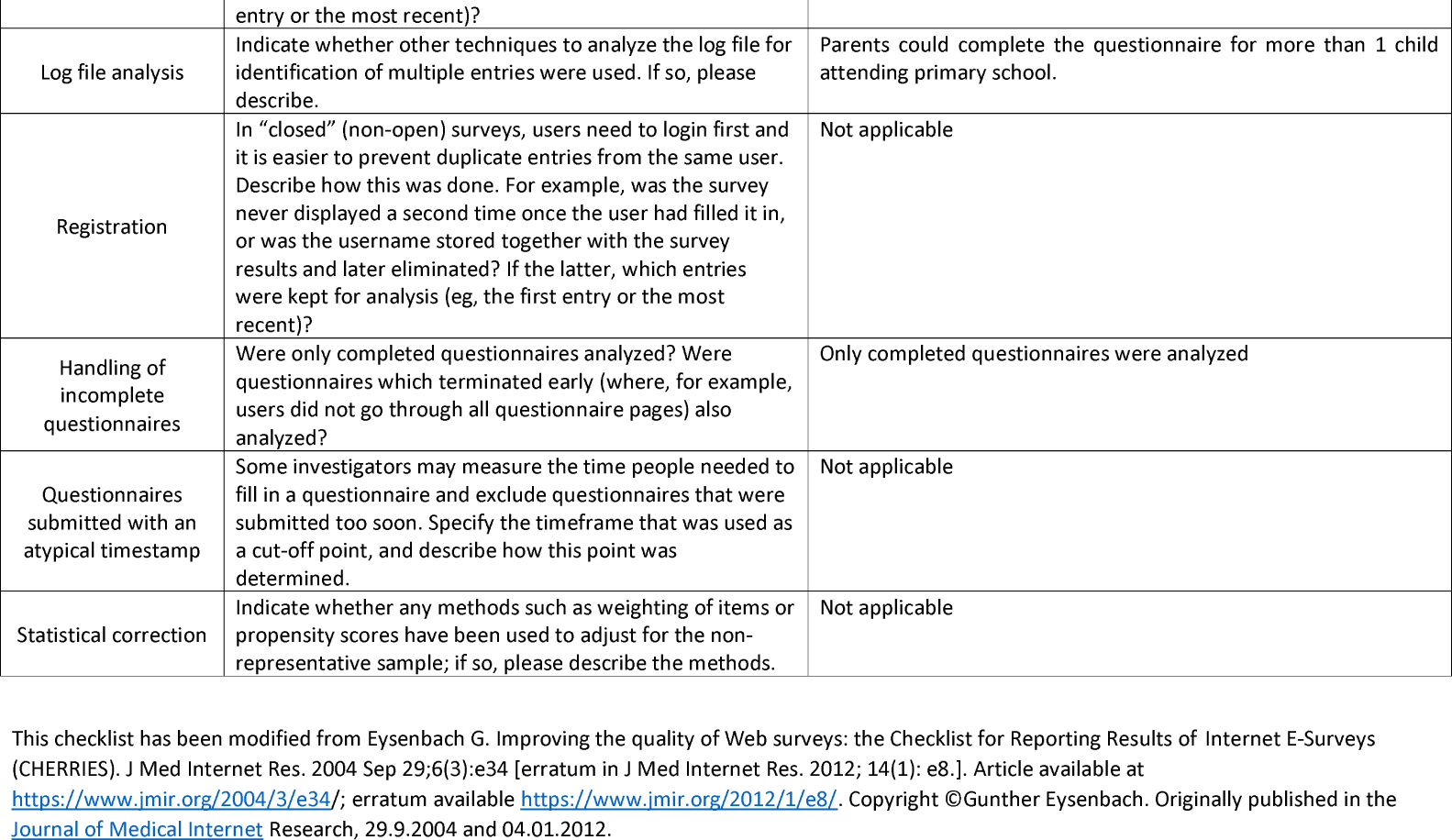
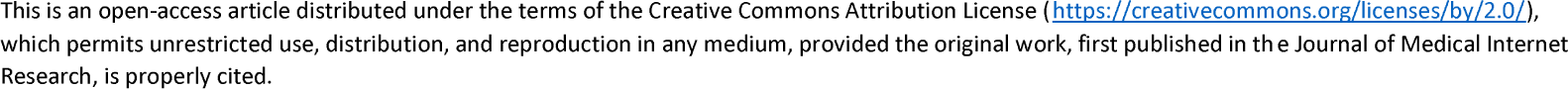

## References

1. World Health Organization. WHO Coronavirus Disease (COVID-19) Dashboard - Cyprus. 2020 [cited 2020 Feb 10]. Available from: https://covid19.who.int/region/euro/country/cy

2. Press Information Office. CORONAVIRUS COVID-19 - CYPRUS [Internet]. 2020 [cited 2020 May 9]. Available from: https://www.pio.gov.cy/coronavirus/

3. Ministry of Education, Culture, Sports and Youth. Ενημερωνóμαστε για τον Κορωνοϊó (Covid-19) [Internet]. 2020 [cited 2020 Dec 8]. Available from: http://www.moec.gov.cy/covid_19.html

4. Miller GW, Jones DP. The Nature of Nurture: Refining the Definition of the Exposome. Toxicological Sciences [Internet]. 2014 Jan [cited 2020 Oct 8];137(1):1–2. Available from: https://academic.oup.com/toxsci/article/1647257/The

5. Wild CP. Complementing the Genome with an “Exposome”: The Outstanding Challenge of Environmental Exposure Measurement in Molecular Epidemiology. Cancer Epidemiology Biomarkers & Prevention [Internet]. 2005 Aug 1 [cited 2020 Apr 8];14(8):1847–50. Available from: http://cebp.aacrjournals.org/cgi/doi/10.1158/1055-9965.EPI-05-0456

6. Dunton GF, Do B, Wang SD. Early effects of the COVID-19 pandemic on physical activity and sedentary behavior in children living in the U.S. BMC Public Health [Internet]. 2020 Dec [cited 2020 Oct 13];20(1):1351. Available from: https://bmcpublichealth.biomedcentral.com/articles/10.1186/s12889-020-09429-3

7. López-Bueno R, López-Sánchez GF, Casajús JA, Calatayud J, Gil-Salmerón A, Grabovac I, et al. Health-Related Behaviors Among School-Aged Children and Adolescents During the Spanish Covid-19 Confinement. Front Pediatr [Internet]. 2020 Sep 11 [cited 2020 Oct 13];8:573. Available from: https://www.frontiersin.org/article/10.3389/fped.2020.00573/fulls

8. Medrano M, Cadenas-Sanchez C, Oses M, Arenaza L, Amasene M, Labayen I. Changes in lifestyle behaviours during the COVID-19 confinement in Spanish children: A longitudinal analysis from the MUGI project. Pediatric Obesity [Internet]. 2020 Sep 24 [cited 2020 Oct 13]; Available from: https://onlinelibrary.wiley.com/doi/10.1111/ijpo.12731

9. Moore SA, Faulkner G, Rhodes RE, Brussoni M, Chulak-Bozzer T, Ferguson LJ, et al. Impact of the COVID-19 virus outbreak on movement and play behaviours of Canadian children and youth: a national survey. Int J Behav Nutr Phys Act [Internet]. 2020 Dec [cited 2020 Oct 13];17(1):85. Available from: https://ijbnpa.biomedcentral.com/articles/10.1186/s12966-020-00987-8

10. Pietrobelli A, Pecoraro L, Ferruzzi A, Heo M, Faith M, Zoller T, et al. Effects of COVID-19 Lockdown on Lifestyle Behaviors in Children with Obesity Living in Verona, Italy: A Longitudinal Study. Obesity [Internet]. 2020 Aug [cited 2020 Oct 13];28(8):1382–5. Available from: https://onlinelibrary.wiley.com/doi/abs/10.1002/oby.22861

11. Xiang M, Zhang Z, Kuwahara K. Impact of COVID-19 pandemic on children and adolescents’ lifestyle behavior larger than expected. Progress in Cardiovascular Diseases [Internet]. 2020 Jul [cited 2020 Oct 19];63(4):531–2. Available from: https://linkinghub.elsevier.com/retrieve/pii/S0033062020300967

12. Harris PA, Taylor R, Thielke R, Payne J, Gonzalez N, Conde JG. Research electronic data capture (REDCap)—A metadata-driven methodology and workflow process for providing translational research informatics support. Journal of Biomedical Informatics [Internet]. 2009 Apr [cited 2020 Oct 8];42(2):377–81. Available from: https://linkinghub.elsevier.com/retrieve/pii/S1532046408001226

13. Mossong J, Hens N, Jit M, Beutels P, Auranen K, Mikolajczyk R, et al. Social Contacts and Mixing Patterns Relevant to the Spread of Infectious Diseases. Riley S, editor. PLoS Med [Internet]. 2008 Mar 25 [cited 2020 Oct 8];5(3):e74. Available from: https://dx.plos.org/10.1371/journal.pmed.0050074

14. Republic of Cyprus, Ministry of Finance. European Health Interview Survey [Internet]. 2014 [cited 2020 Apr 14]. Available from: http://www.mof.gov.cy/mof/cystat/statistics.nsf/All/4DE6CF465BC4DDCAC225806800372D29?OpenDocument&sub=3&sel=1&e=&print.

15. European Urban Health Indications System Part 2. EURO-URHIS [Internet]. 2012. Available from: https://web.archive.org/web/20190308170137//http:results.urhis.eu/Default.aspx?Code=YY&Group=B

16. Pope D, Puzzolo E, Birt C, Guha J, Higgerson J, Patterson L, et al. Collecting standardised urban health indicator data at an individual level for adults living in urban areas: methodology from EURO-URHIS 2. Eur J Public Health [Internet]. 2016 Jan 7 [cited 2020 Oct 8];ckv220. Available from: https://academic.oup.com/eurpub/article-lookup/doi/10.1093/eurpub/ckv220

17. Ioannou S, Ioannou S, Andrianou XD, Charisiadis P, Christophi CA, Yiasoumi G, et al. Occupational exposures to disinfectants and pre-diabetes status among active nurses in Cyprus. Scand J Work Environ Health [Internet]. 2019 Sep [cited 2020 Oct 20];45(5):505–13. Available from: http://www.sjweh.fi/show_abstract.php?abstract_id=3804

18. Chortiatinou S, Risvas G, Zampelas A. «Eat healthy - live like scout», a nutrition behaviour change program in scouts, aged 12-15 years. Clinical Nutrition ESPEN [Internet]. 2016 Jun [cited 2020 Oct 8];13:e58. Available from: https://linkinghub.elsevier.com/retrieve/pii/S2405457716301164

19. Kowalski KC, Crocker PRE, Donen RM. The Physical Activity Questionnaire for Older Children (PAQ-C) and Adolescents (PAQ-A) Manual. Canada: College of Kinesiology, University of Saskatchewan, 1–38. [Internet]. Canada: College of Kinesiology, University of Saskatchewan; 2004 p. 1–38. Available from: https://www.prismsports.org/UserFiles/file/PAQ_manual_ScoringandPDF.pdf

20. Kowalski KC, Crocker PRE, Faulkner RA. Validation of the Physical Activity Questionnaire for Older Children. Pediatric Exercise Science [Internet]. 1997 May [cited 2020 Oct 8];9(2):174–86. Available from: https://journals.humankinetics.com/view/journals/pes/9/2/article-p174.xml

21. R Core Team. R: A Language and Environment for Statistical Computing [Internet]. 2017. Available from: http://www.r-project.org/

22. RStudio Team. RStudio: Integrated Development Environment for R [Internet]. 2015. Available from: http://www.rstudio.com/

23. Haddad N, Andrianou XD, Makris KC. A Scoping Review on the Characteristics of Human Exposome Studies. Curr Pollution Rep [Internet]. 2019 Dec [cited 2020 Apr 14];5(4):378–93. Available from: http://link.springer.com/10.1007/s40726-019-00130-7

24. Andra SS, Charisiadis P, Karakitsios S, Sarigiannis DA, Makris KC. Passive exposures of children to volatile trihalomethanes during domestic cleaning activities of their parents. Environmental Research [Internet]. 2015 Jan [cited 2020 Oct 20];136:187–95. Available from: https://linkinghub.elsevier.com/retrieve/pii/S0013935114003818

25. WHO. Childhood Obesity Surveillance Initiative - HIGHLIGHTS 2015-17 [Internet]. 2018 [cited 2020 Jul 25]. Available from: https://www.euro.who.int/__data/assets/pdf_file/0006/372426/WH14_COSI_factsheets_v2.pdf

26. Andrianou XD, Pronk A, Galea K, Stierum R, Loh M, Riccardo F, et al. Exposome-based public health interventions for infectious diseases in urban settings. Under review. 2020;

