## Supplementary material for "Exposome changes in primary school children following the wide population non-pharmacological interventions implemented due to COVID-19 in Cyprus: a national survey": SI section

Table S1 Variables used in the model with outcome being increase in hours at home during weekdays (n=54)

| BMI_sds | sugar_now |
| --- | --- |
| days_from_reopening | sugar_before |
| change_cleaning_timesweek_total | chips_now |
| change_cig_no_cigsperday | chips_before |
| change_spareactivity | breakfast_now |
| change_contact_no_wd_home | breakfast_before |
| change_contact_no_wd_school | supplements_now |
| change_contact_no_wd_elsewhere | supplements_before |
| change_contact_no_wd_vuln_home | fast_food_now |
| change_contact_no_we_home | fast_food_before |
| change_contact_no_we_elsewhere | net_friends_now |
| change_contact_no_we_vuln_home | net_friends_before |
| change_hours_hm_we | net_family_now |
| free_time_now | net_family_before |
| free_time_before | tv_pc_now |
| break_now | tv_pc_before |
| break_before | hands_soap_now |
| fruits_now | hands_soap_before |
| fruits_before | hands_anticeptic_now |
| veggies_now | hands_anticeptic_before |
| veggies_before | city |
| meat_now | chronic_disease |
| meat_before | vaccine_doses |
| leggumes_now | no_vaccination |
| leggumes_before | attend_school |
| fish_now | consent |
| fish_before | num_child |

Table S2 Variables used in the model with outcome being decrease in the number of vulnerable contacts at home during weekdays (n=54)

| BMI_sds | sugar_now |
| --- | --- |
| days_from_reopening | sugar_before |
| change_cleaning_timesweek_total | chips_now |
| change_cig_no_cigsperday | chips_before |
| change_spareactivity | breakfast_now |
| change_contact_no_wd_home | breakfast_before |
| change_contact_no_wd_school | supplements_now |
| change_contact_no_wd_elsewhere | supplements_before |
| change_contact_no_we_home | fast_food_now |
| change_contact_no_we_elsewhere | fast_food_before |
| change_contact_no_we_vuln_home | net_friends_now |
| change_hours_hm_wd | net_friends_before |
| change_hours_hm_we | net_family_now |
| free_time_now | net_family_before |
| free_time_before | tv_pc_now |
| break_now | tv_pc_before |
| break_before | hands_soap_now |
| fruits_now | hands_soap_before |
| fruits_before | hands_anticeptic_now |
| veggies_now | hands_anticeptic_before |
| veggies_before | city |
| meat_now | chronic_disease |
| meat_before | vaccine_doses |
| leggumes_now | no_vaccination |
| leggumes_before | attend_school |
| fish_now | consent |
| fish_before | num_child |

Table S3 Variables used in the model with outcome being decrease in the number of contacts at school (n=52)

| BMI_sds | fish_before |
| --- | --- |
| days_from_reopening | sugar_now |
| change_cleaning_timesweek_total | sugar_before |
| change_cig_no_cigsperday | chips_now |
| change_spareactivity | chips_before |
| change_contact_no_wd_home | breakfast_now |
| change_contact_no_wd_elsewhere | breakfast_before |
| change_contact_no_wd_vuln_home | supplements_now |
| change_contact_no_we_home | supplements_before |
| change_contact_no_we_elsewhere | fast_food_now |
| change_contact_no_we_vuln_home | fast_food_before |
| change_hours_hm_wd | net_friends_now |
| change_hours_hm_we | net_friends_before |
| free_time_now | net_family_now |
| free_time_before | net_family_before |
| break_now | tv_pc_now |
| break_before | tv_pc_before |
| fruits_now | hands_soap_now |
| fruits_before | hands_soap_before |
| veggies_now | hands_anticeptic_now |
| veggies_before | hands_anticeptic_before |
| meat_now | city |
| meat_before | chronic_disease |
| leggumes_now | vaccine_doses |
| leggumes_before | no_vaccination |
| fish_now | num_child |

Table S4 Variables used in the model with outcome being increase of hand washing frequency using antiseptic or soap (n=55)

| BMI_sds | sugar_now |
| --- | --- |
| days_from_reopening | sugar_before |
| change_cleaning_timesweek_total | chips_now |
| change_cig_no_cigsperday | chips_before |
| change_spareactivity | breakfast_now |
| change_contact_no_wd_home | breakfast_before |
| change_contact_no_wd_school | supplements_now |
| change_contact_no_wd_elsewhere | supplements_before |
| change_contact_no_wd_vuln_home | fast_food_now |
| change_contact_no_we_home | fast_food_before |
| change_contact_no_we_elsewhere | net_friends_now |
| change_contact_no_we_vuln_home | net_friends_before |
| change_hours_hm_wd | net_family_now |
| change_hours_hm_we | net_family_before |
| free_time_now | tv_pc_now |
| free_time_before | tv_pc_before |
| break_now | hands_soap_now |
| break_before | hands_soap_before |
| fruits_now | hands_anticeptic_now |
| fruits_before | hands_anticeptic_before |
| veggies_now | city |
| veggies_before | chronic_disease |
| meat_now | vaccine_doses |
| meat_before | no_vaccination |
| leggumes_now | attend_school |
| leggumes_before | consent |
| fish_now | num_child |
| fish_before |  |

Table S5 Chronic diseases for children as reported by their parents (n=122)

| **Chronic disease** | **N (%)** |
| --- | --- |
| Allergies | 41 (33.6) |
| Asthma | 40 (32.8) |
| Bronchitis | 9 (7.4) |
| Asthma & allergies | 4 (3.3) |
| Diabetes | 4 (3.3) |
| Allergies & bronchitis | 2 (1.6) |
| Epilepsy | 2 (1.6) |
| Hemoglobinopathy | 2 (1.6) |
| Mitral valve prolapse | 2 (1.6) |
| Emphysema | 1 (0.8) |
| Long QT | 1 (0.8) |
| Frequent infections | 1 (0.8) |
| Bronchitis & asthma | 1 (0.8) |
| Ulcerative colitis | 1 (0.8) |
| G6PD_lack | 1 (0.8) |
| Heart disease | 1 (0.8) |
| Celiac disease | 1 (0.8) |
| Mediterannean fever | 1 (0.8) |
| Right lung rupture | 1 (0.8) |
| Stigma | 1 (0.8) |
| Congenital heart disease | 1 (0.8) |
| Olier syndrome | 1 (0.8) |
| Hypothyroidism | 1 (0.8) |
| Atopic dermatitis | 1 (0.8) |
| More than 2 chronic diseases | 1 (0.8) |

Table S6 BMI for age by sex. p-value based on Wilcoxon test.

|  | **Males**  **(n=779)** | **Females**  **(n=709)** | **p-value** |
| --- | --- | --- | --- |
| **BMI_for_age (%)** |  |  | <0.001 |
| Thinness | 37 ( 5.1) | 40 ( 6.0) |  |
| Normal | 387 (53.6) | 402 (60.4) |  |
| Overweight | 165 (22.9) | 155 (23.3) |  |
| Obese | 133 (18.4) | 69 (10.4) |  |

Table S7 Number (%) of primary school children who were in contact at home with at least 1 person belonging in the vulnerable groups in the pre- and post-lockdown periods in weekends and weekdays. People in vulnerable groups were defined as people over 60 years old, pregnant women, people with chronic diseases.

|  | **Pre-lockdown N (%)** | **Post-lockdown N (%)** |
| --- | --- | --- |
| **weekdays** | 1155 (76.5) | 948 (62.8) |
| **weekends** | 1174 (77.9) | 958 (63.4) |

Table S8 Correlation analysis results. Adjusted for age, sex and parents’ educational level. The total number of variables used were 55. Only correlations with r ≥ 0.7 are shown (n=7).

| **From variable** | **To variable** | **r** |
| --- | --- | --- |
| Fish consumption (pre-lockdown) | Fish consumption (post -lockdown) | 0.82 |
| Vegetables consumption (pre-lockdown) | Vegetables consumption (post-lockdown) | 0.82 |
| Sugar consumption (pre-lockdown) | Sugar consumption (post -lockdown) | 0.79 |
| Fruits consumption (pre-lockdown) | Fruits consumption (post -lockdown) | 0.79 |
| Meat consumption (pre-lockdown) | Meat consumption (post -lockdown) | 0.77 |
| Snacks consumption (pre-lockdown) | Snacks consumption (post -lockdown) | 0.76 |
| Legumes consumption (pre-lockdown) | Legumes consumption (post -lockdown) | 0.73 |

Table S9 Parameters with FDR adjusted p-value ≤ 0.05

| **Variable** | **Term** | **Estimate** | **Std error** | **Statistic** | **p-value** | **Lower CI** | **Upper CI** | **Outcome** | **FDR-adjusted p-value** |
| --- | --- | --- | --- | --- | --- | --- | --- | --- | --- |
| Days from reopening |  | 0.972 | 0.007 | -3.881 | 0 | 0.958 | 0.986 | Increase in hours staying at home - weekdays | <0.001 |
| Hands antiseptic pre-lockdown | ≥4 times per day | 0.543 | 0.192 | -3.179 | 0.001 | 0.373 | 0.794 | Increase in hours staying at home - weekdays | 0.031 |
| Break post-lockdown | Sitting, standing, walking | 1.509 | 0.133 | 3.082 | 0.002 | 1.161 | 1.959 | Increase in hours staying at home - weekdays | 0.043 |
| Meat post-lockdown | 4-7 times per week | 1.476 | 0.124 | 3.142 | 0.002 | 1.159 | 1.885 | Increase in hours staying at home - weekdays | 0.043 |
| City | Paphos | 0.442 | 0.23 | -3.551 | 0 | 0.283 | 0.696 | Increase in hours staying at home - weekdays | <0.001 |
| Returned or not to school | Stays at home | 2.167 | 0.218 | 3.554 | 0 | 1.436 | 3.38 | Increase in hours staying at home - weekdays | <0.001 |
| % Change contacts no home - weekdays |  | 0.989 | 0.001 | -7.516 | 0 | 0.986 | 0.992 | Decrease of vulnerable contacts at home - weekdays | <0.001 |
| % Change vulnerable contacts no home - weekends |  | 0.989 | 0.001 | -7.764 | 0 | 0.986 | 0.992 | Decrease of vulnerable contacts at home - weekdays | <0.001 |
| % Change contacts no elsewhere -weekdays |  | 0.993 | 0.001 | -6.54 | 0 | 0.991 | 0.995 | Decrease in the number of contacts at school) | <0.001 |
| Sugar pre-lockdown | <4 times per week | 0.545 | 0.163 | -3.717 | 0 | 0.395 | 0.749 | Decrease in the number of contacts at school) | <0.001 |
| Digital communication friends pre-lockdown | Never | 2.203 | 0.254 | 3.11 | 0.002 | 1.352 | 3.67 | Decrease in the number of contacts at school | 0.043 |
| Hands antiseptic pre-lockdown | ≥4 times per day | 0.254 | 0.222 | -6.166 | 0 | 0.165 | 0.395 | Decrease in the number of contacts at school | <0.001 |
| Sugar post-lockdown | <4 times per week | 0.564 | 0.162 | -3.548 | 0 | 0.411 | 0.774 | Decrease in the number of contacts at school | <0.001 |
| Days from reopening |  | 0.977 | 0.008 | -3.158 | 0.002 | 0.962 | 0.991 | Increase in hand hygiene - soap or antiseptic | 0.043 |

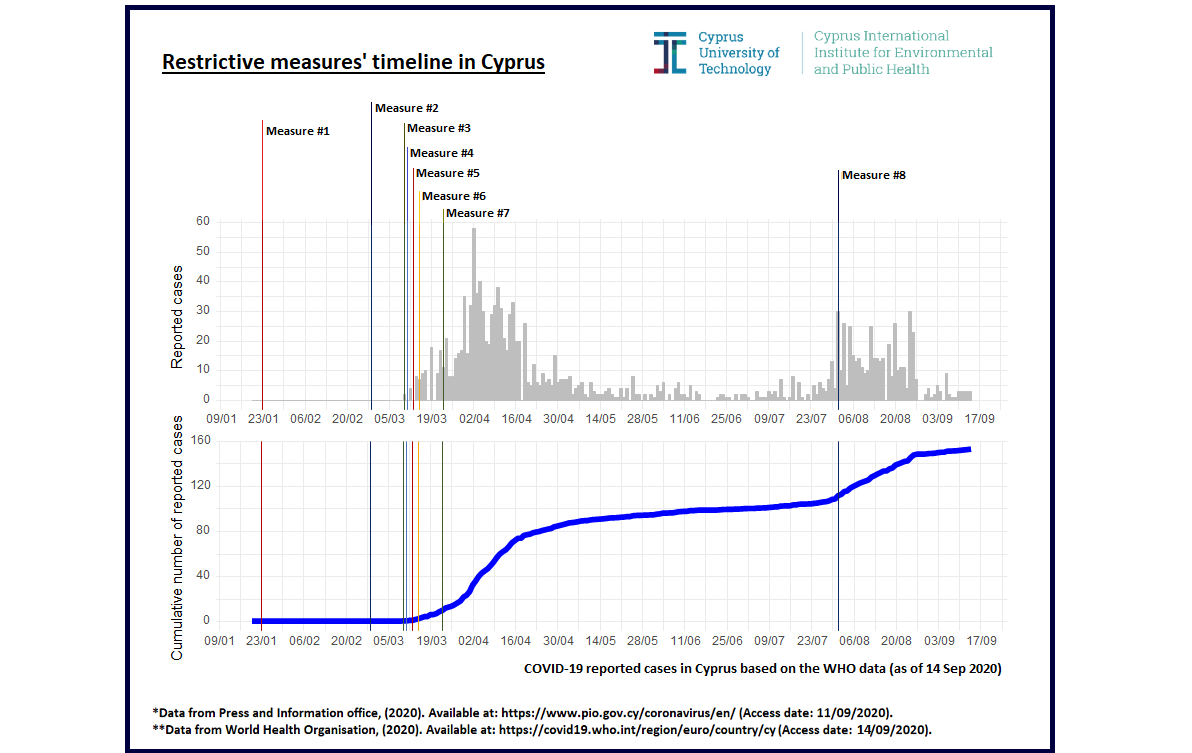

**Figure S1** Restrictive measures timeline in Cyprus.

**Measure #1**^1,2^: Strict control measures on people arriving in Cyprus from China.

**Measure #2**:

- Activation of the National Crisis Management Centre^2^
- Closure of 4 crossing points for more effective control of the crossings^2^
- Registration of government accommodation or campsites which in case of a pandemic would be necessary to isolate suspected cases^2,3^

**Measure #3^4^**:

- Closure of Nicosia schools at all levels, private and public for three days
- Ban on events and other mass indoor gatherings of more than 75 people until 31 March 2020
- Cancellation of mass events, rallies, parades and concerts in public places
- Football and other matches will normally take place without the presence of spectators

**Measure #4^5^**:

- Closure of all schools across Cyprus at all stages of education from March 13 to March 20, 2020.
- Suspended operation of all Ministry educational programs, afternoon and evening, of the 2 Special Schools, public and private, as well as tutorials across Cyprus from March 13 to 20, 2020.
- Suspended operation in higher education institutions, public and private, throughout Cyprus, until Sunday, March 22, 2020.

**Measure #5^2^:**

- From 1 a.m. of 15 March and for a period of 15 days in the first phase, the prohibition of entry to any citizen - regardless of nationality - who does not fall into the following categories:

• Cypriot citizens.

• Legal residents in the Republic of Cyprus.

• European nationals or third-country nationals working in the Republic.

• Nationals of countries who are in a designated diplomatic service or mission under bilateral or international conventions.

• Individual cases of European nationals or third-country nationals for unavoidable professional obligations, provided that the relevant authorization has been obtained from the competent Ministry.

• European or third country citizens attending educational institutions in the Republic of Cyprus.

- As regards the crossing points between the free and the occupied areas, entry will be permitted only to persons falling into the following categories:

• Cypriot citizens, Greek Cypriots and Turkish Cypriots.

• Legal residents in the free areas.

• Nationals of countries who are in a designated diplomatic service or mission under bilateral or international conventions.

- The suspension of classes in public and private educational institutions of the country is extended until 10 April.

**Measure #6:**

- Entry to the Republic of Cyprus between 18.00 on 16/3/2020 and 30/4/2020, will only be granted to individuals (see above), provided that, upon arrival, they are able to submit a medical certificate, issued no more than 4 days before, showing that they have been tested for coronavirus by certified medical centers in their country of origin^6^.
- All travellers returning to Cyprus from abroad, regardless of their country of origin, will be placed under a 14-day compulsory quarantine at accommodation facilities designated by the Republic of Cyprus^6^.
- Suspension of private business operations^6^.
- From March 16, 2020 from 6 am and for a period of four weeks, the following undertakings shall be closed^7^:

▪ Shopping malls and department stores

▪ Cafes, bars and all caterers with the exception of those that offer or have the ability to offer delivery services

▪ Entertainment centers

▪ Cinemas, theatres and auditoriums

▪ Libraries

▪ Museums, archaeological and historical sites

▪ Gaming agencies, casinos, etc.

▪ Sports facilities, sports clubs, cultural clubs and clubs

▪ Theme parks (amusement parks etc)

▪ Hairdressers, beauty salons.

- Retail businesses that do not fall into the category of suspended business can continue their business by ensuring that there are no more than five employees in the workplace^7^.
- All other companies that do not fall under the above categories can continue operating as they will strictly adhere to the hygiene and protection of personnel and premises^7^.

**Measure #7^8^:**

- Unnecessary movements are prohibited from 6 pm of March 24 2020 to 6am of 13 April 2020, with the exception of the following cases:

(i) Transfer to and from workplaces as well as for purposes work,
(ii) Absolutely necessary visits to government agencies, its services wider public sector and local government and transfers for purchase or supply of essential goods / services to and from businesses / services whose operation has not been suspended and if it is impossible to deliver them
(iii) Visiting a doctor or donating or going to a pharmacy,
(iv) Going to a bank, if the electronically transaction it is not possible,
(v) Trafficking to assist our relatives and / or fellow citizens who are unable to serve themselves or in groups that have to self-protective or self-restricted and / or in places compulsory restriction (quarantine),
(vi) First and second-degree relatives who attend ceremonies such as funerals, weddings and baptisms should not exceeding 10 persons.
(vii) movements for physical activity or for the needs of the pet, as long as they do not exceed two persons and are confined to adjacent once residence areas

provided that, all persons whose movement is permitted as above, carry with them an ID card or passport with a supplement evidence when requested by the competent authorities.

- From 6 pm on March 24, 2020 until 6:00 am of 13 April, people are not allowed to visit the following sites:

(i)	Parks,

(ii)	Playgrounds.

(iii)	Open sports grounds;

(iv)	Public meeting places, including squares, dams, excursions, beaches, marinas (It is understood that the operation of the above companies is also suspended).

- From 6 pm on March 24, 2020 until 6 am of 13 April 2020:

(i)	Any form of public market is suspended, or street sale and bargains,

(ii)	works on site other than those relating to projects are suspended public utility, for which a license will be issued by him Minister for Transport, Communication and Works,

(iii)	Citizens are prohibited from attending religious sites worship, such as churches, mosques and other places of worship, and

(iv)	Τhe custom of "labratzia" is forbidden and thus, any woods or other materials already assembled outside churches would remove. Local authorities in cooperation with the Police are the main responsible for enforcing the ban.

(v)	Closure of all retail businesses with exception^9^:

• Disability and orthopedic companies and laboratories.

• Medical and industrial gas companies and laboratories; and machinery.

• Optical companies and laboratories.

• The companies selling hearing aids.

• Car and motorcycle garages.

• Tire dealers.

• Bicycle trading and repair companies.

• Cleaners.

• Postal and transport service companies (courier).

• Pet and feed businesses or veterinary medicines.

• The companies of telecommunications providers, to the extent that relates to payment services, account renewal, repair and replacement of mobile devices.

• Companies that produce pesticides, fertilizers, medicines and agricultural materials or equipment.

• Businesses for the sale of cars and motor vehicles.

• Funeral offices.

• Nurseries and florists.

**Measure #8^10^:**

The use of a mask bacame mandatory by the general population as well as by employees:

1. The use of a mask is mandatory by citizens, 6 years and older, at the following establishments / venues:

- Supermarkets / bakeries
- Department stores / retail stores / shopping malls
- Churches - Visits to hospitals / clinics / nursing homes / other health care institutions
- Pharmacies
- Organizations/Departments of the public and wider public sector and private businesses which receive/serve customers (e.g. banks, citizen service centers, post offices, Cyprus Electricity Authority, etc).
- Betting shops
- Elevators

2. The use of a mask is mandatory for employees in customer service positions, at the following establishments / venues / businesses:

- Public transport (e.g. bus drivers)
- Indoor areas of private companies, Departments of the public and wider public sector which serve the public, such as citizen service centers, post offices, couriers, places for the payment of utility bills of organizations (e.g. EAC, CYTA, local government, etc)
- Persons who work at customer service in retail businesses.

Indicatively:

▪ Tills at supermarkets, bakeries, pharmacies, public markets, retail stores, etc.

▪ Customer service at other places such as butcheries, fish markets, fruit markets, kiosks, mini markets, retail trade businesses (e.g. clothing and shoe stores, cosmetic stores, and so forth), etc.

- Delivery persons / distributors
- Construction workers: people working in construction must use disposable masks when traveling by car (the number of persons in a car must not exceed 2 people) and when using an elevator.
- Any other business or establishment, where the use of a mask is foreseen according to the relevant protocols of the Ministry of Health

1: Press and Information Office (PIO), 2020. Available at: <https://www.pio.gov.cy/coronavirus/press/23012020-17.pdf> (Accessed date: 21/03/2020).

2: Press and Information Office (PIO), 2020. Available at: <https://www.pio.gov.cy/coronavirus/press/13032020_17.pdf> (Accessed date: 21/03/2020).

3: Press and Information Office (PIO), 2020. Available at: <https://www.pio.gov.cy/coronavirus/press/27022020-28.pdf> (Accessed date: 21/03/2020).

4: Press and Information Office (PIO), 2020. Available at: <https://www.pio.gov.cy/coronavirus/press/10032020_6.pdf> (Accessed date: 21/03/2020).

5: Press and Information Office (PIO), 2020. Available at: <https://www.pio.gov.cy/coronavirus/press/11032020_5.pdf> (Accessed date: 21/03/2020).

6: Press and Information Office (PIO), 2020. Available at: <https://www.pio.gov.cy/coronavirus/press/15032020_5.pdf> (Accessed date: 21/03/2020).

7: Press and Information Office (PIO), 2020. Available at: <https://www.pio.gov.cy/coronavirus/press/15032020_9.pdf> (Accessed date: 21/03/2020).

8: Press and Information Office (PIO), 2020. Available at: <https://www.pio.gov.cy/coronavirus/press/23032020_12.pdf> (Accessed date: 24/03/2020).

9: Press and Information Office (PIO), 2020. Available at: <https://www.pio.gov.cy/coronavirus/diat/10.pdf> (Accessed date: 24/03/2020).

10: Press and Information Office (PIO), 2020. Available at: <https://www.pio.gov.cy/coronavirus/en/press/03082020_2.pdf> (Accessed date: 05/08/2020)

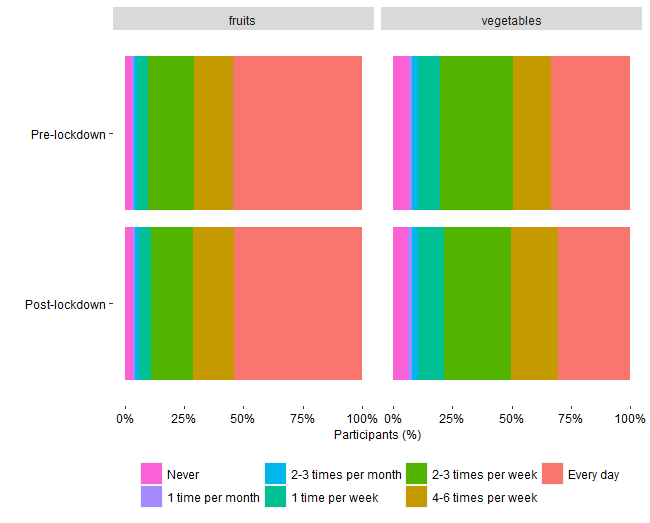

Figure S2 Consumption frequency of fruits and vegetables by primary school children (n=1509) before the lockdown (pre-lockdown) and following the re-opening of schools (post-lockdown).

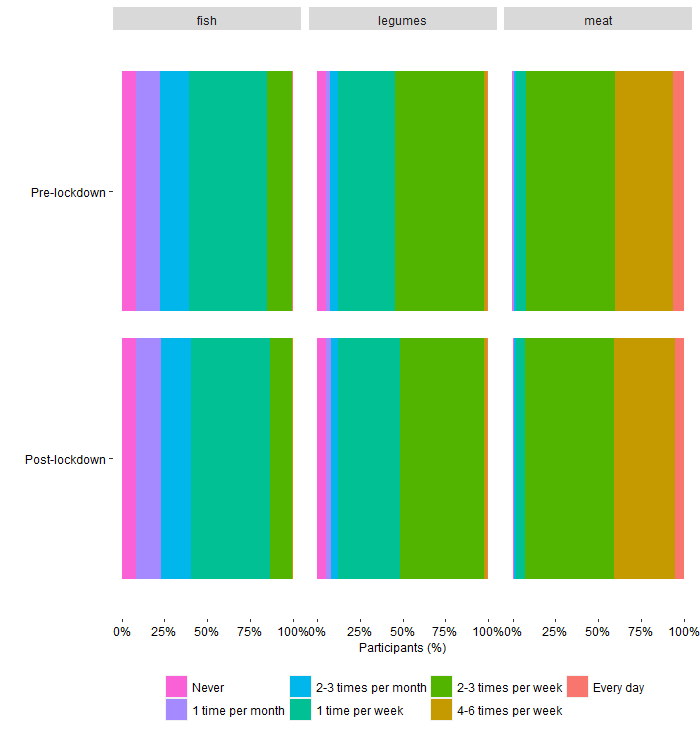

Figure S3 Consumption frequency of fish, legumes and meat by primary school children (n=1509) before the lockdown (pre-lockdown) and following the re-opening of schools (post-lockdown).

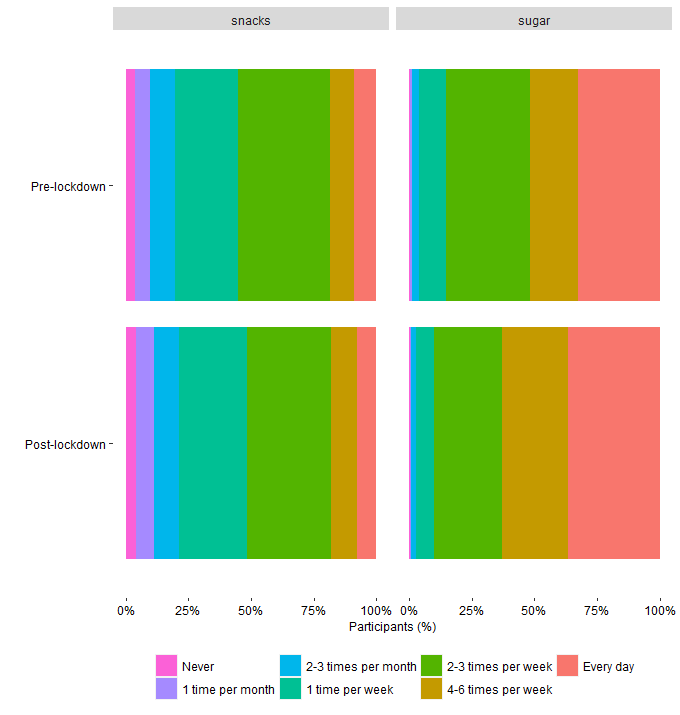

Figure S4 Consumption frequency of snacks and sugary food items by primary school children (n=1509) before the lockdown (pre-lockdown) and following the re-opening of schools (post-lockdown).

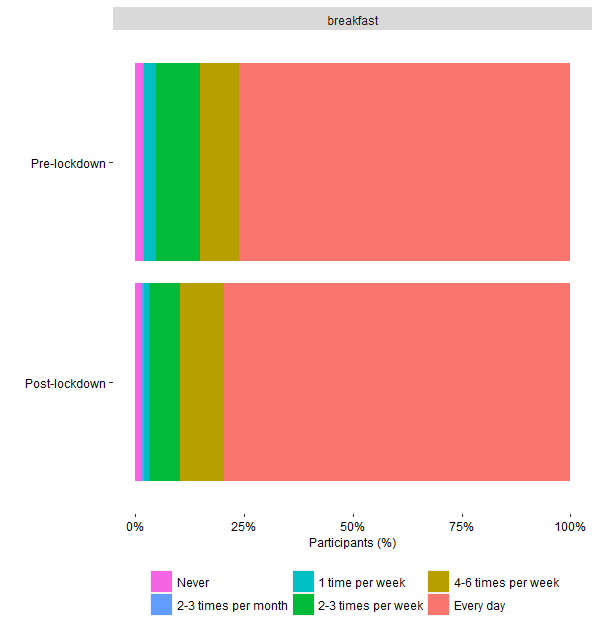

Figure S5 Consumption frequency of breakfast by primary school children (n=1509) before the lockdown (pre-lockdown) and following the re-opening of schools (post-lockdown).

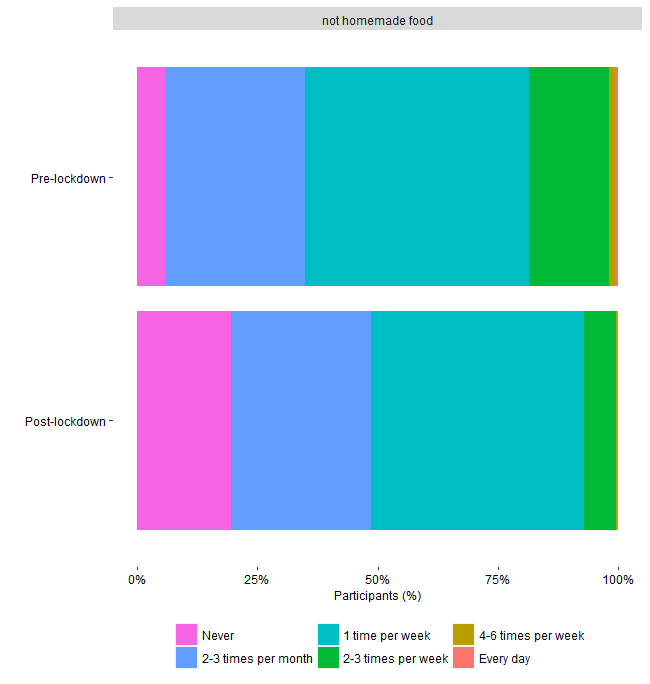

Figure S6 Consumption frequency of ready-made food by primary school children (n=1509) before the lockdown (pre-lockdown) and following the re-opening of schools (post-lockdown).

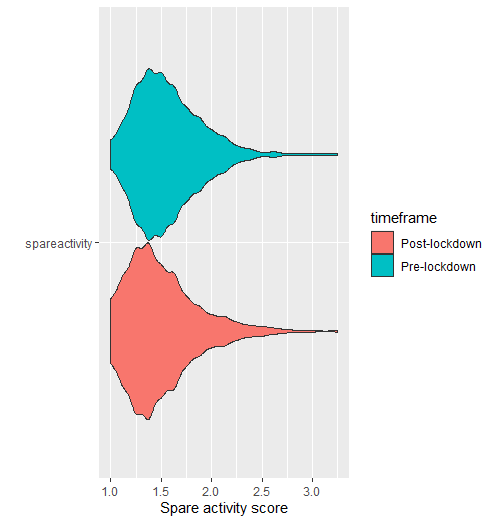

Figure S7 Spare time activity score for primary school children (n=1509) before the lockdown (pre-lockdown) and following the re-opening of schools (post-lockdown).

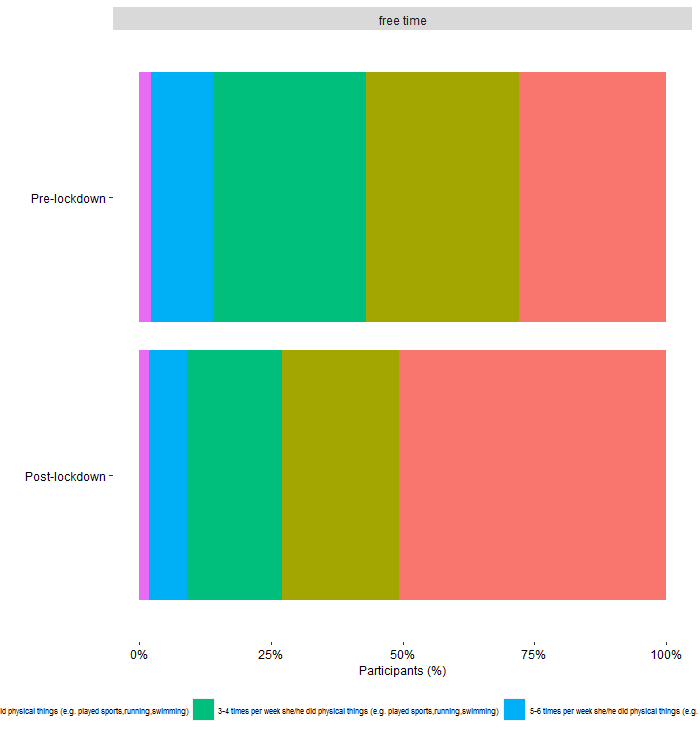

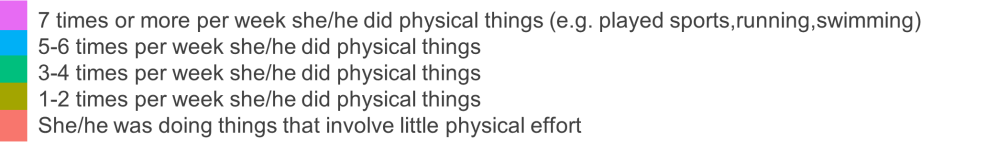

Figure S8 Free time activity for primary school children (n=1509) before the lockdown (pre-lockdown) and following the re-opening of schools (post-lockdown).

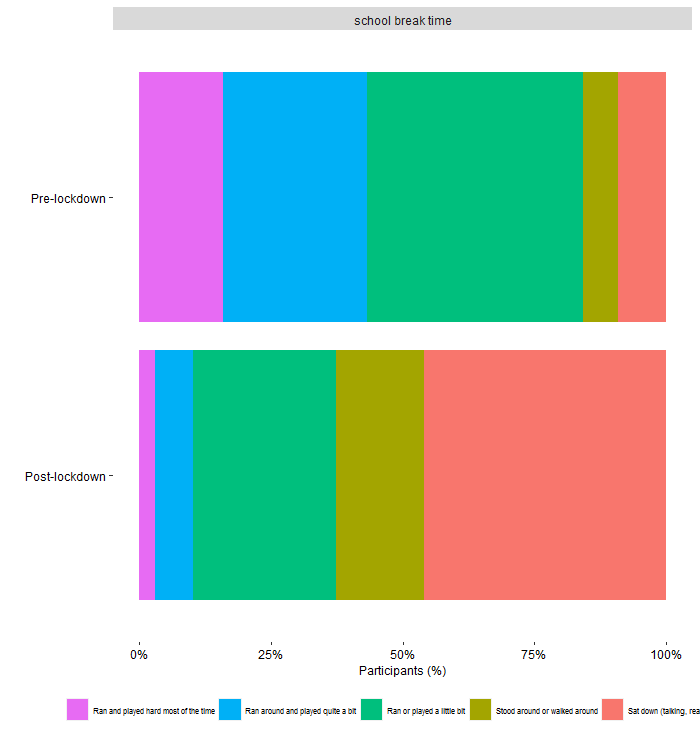

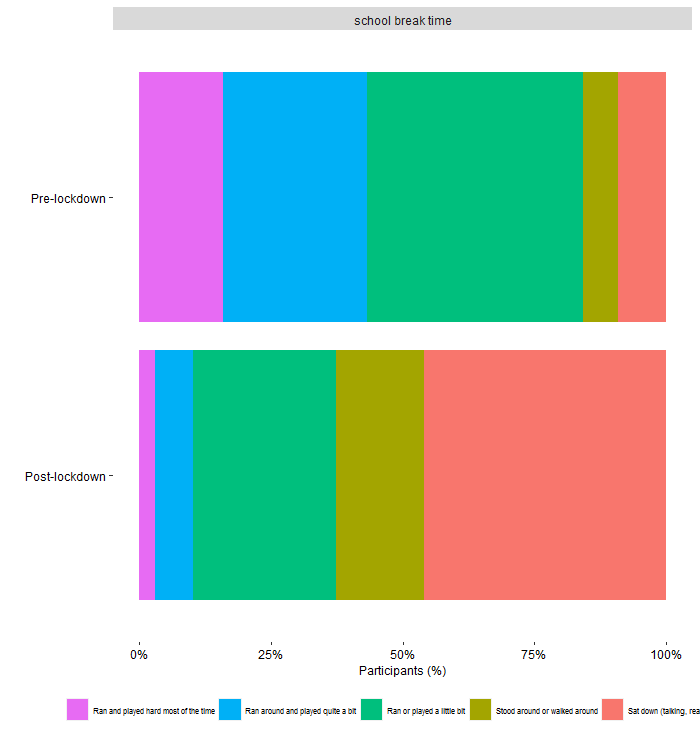

Ran and played hard most of the time

Ran around and played quite a bit

Ran or played a little

Stood or walked around

Sat down

Figure S9 School break time activity for primary school children (n=1509) before the lockdown (pre-lockdown) and following the re-opening of schools (post-lockdown).

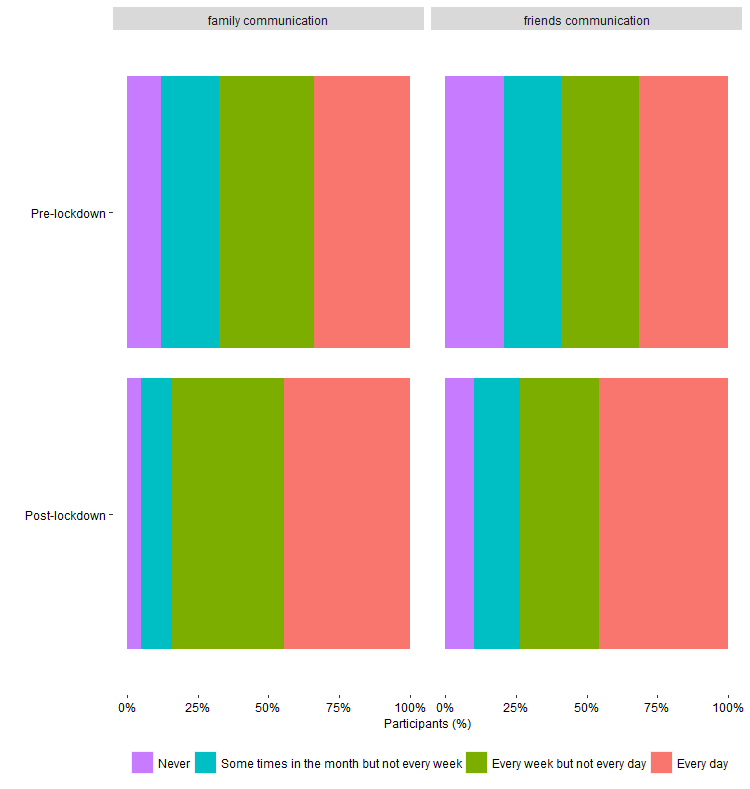

Figure S10 Communication frequency with family and friends for primary school children (n=1509) before the lockdown (pre-lockdown) and following the re-opening of schools (post-lockdown).

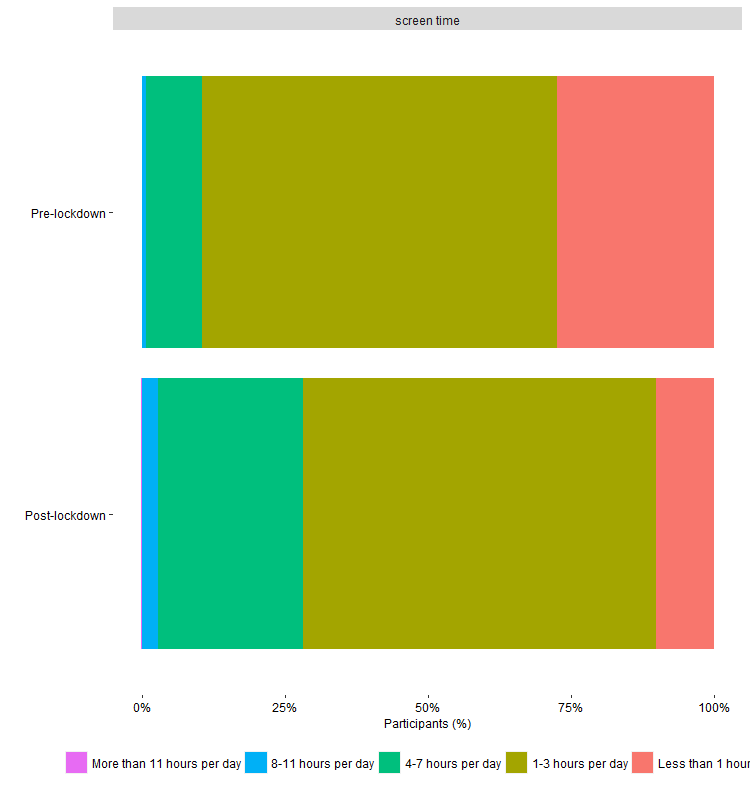

Figure S11 Screen time frequency with family and friends for primary school children (n=1509) before the lockdown (pre-lockdown) and following the re-opening of schools (post-lockdown).

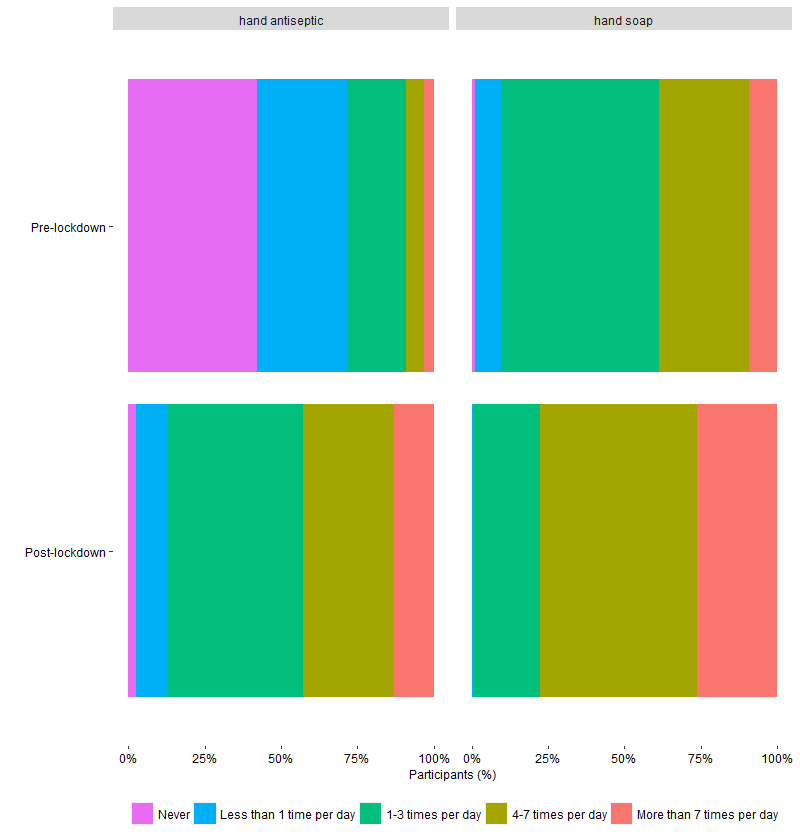

Figure S12 Hand hygiene frequency (hand antiseptic and hand-washing with soap) for primary school children (n=1509) before the lockdown (pre-lockdown) and following the re-opening of schools (post-lockdown).

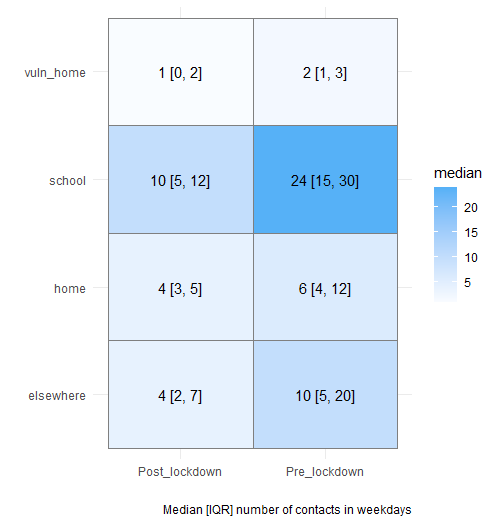

Figure S13 Median number of contacts at home (separately for vulnerable groups), school and elsewhere, during weekdays, for primary school children (n=1509) before the lockdown (pre-lockdown) and following the re-opening of schools (post-lockdown).

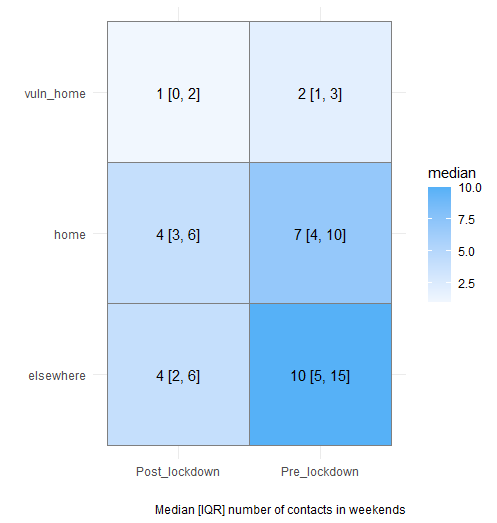

Figure S14 Median number of contacts at home (separately for vulnerable groups) and elsewhere, during weekends, for primary school children (n=1509) before the lockdown (pre-lockdown) and following the re-opening of schools (post-lockdown).

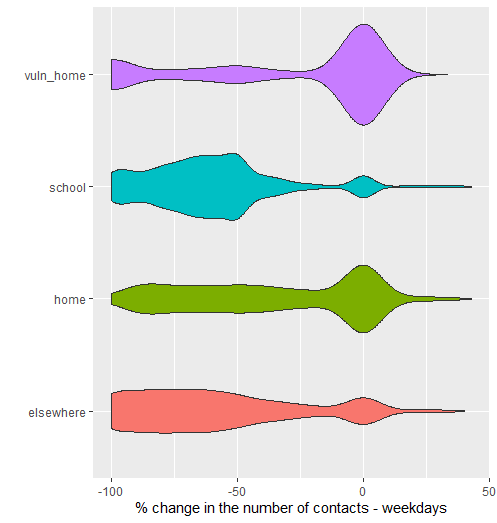

Figure S15 Change (%) in the number of contacts at home (separately for vulnerable groups), school and elsewhere, during weekdays, for primary school children (n=1509) following the re-opening of schools (post-lockdown) and compared with the period before the lockdown (pre-lockdown).

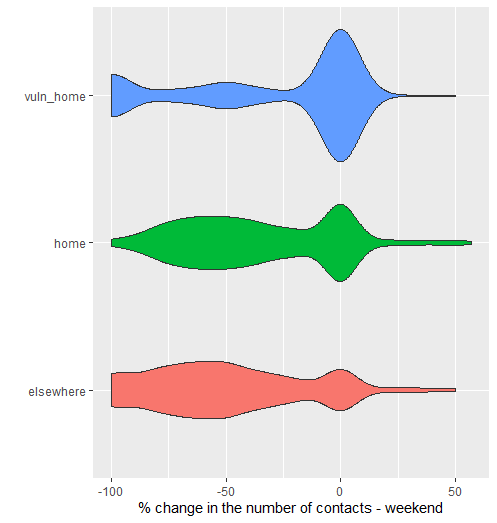

Figure S16 Change (%) in the number of contacts at home (separately for vulnerable groups) and elsewhere, during weekends, for primary school children (n=1509) following the re-opening of schools (post-lockdown) and compared with the period before the lockdown (pre-lockdown).

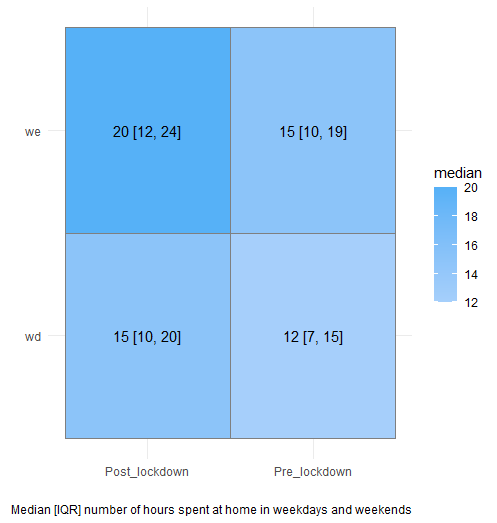

Figure S17 Median number of hours staying at home, during weekends and weekdays, for primary school children (n=1509) before the lockdown (pre-lockdown) and following the re-opening of schools (post-lockdown).

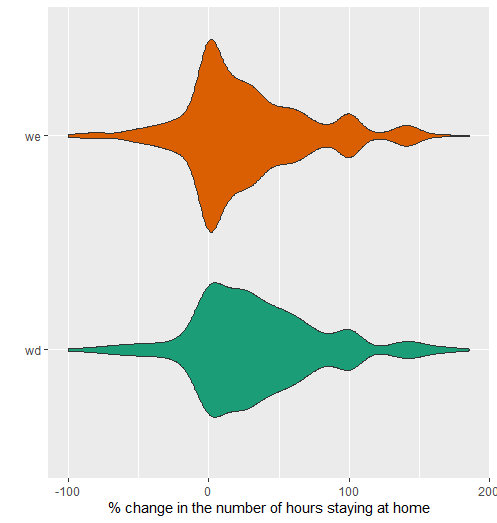

Figure S18 Change (%) in the number of hours staying at home, during weekends and weekdays, for primary school children (n=1509) following the re-opening of schools (post-lockdown) and compared with the period before the lockdown (pre-lockdown).

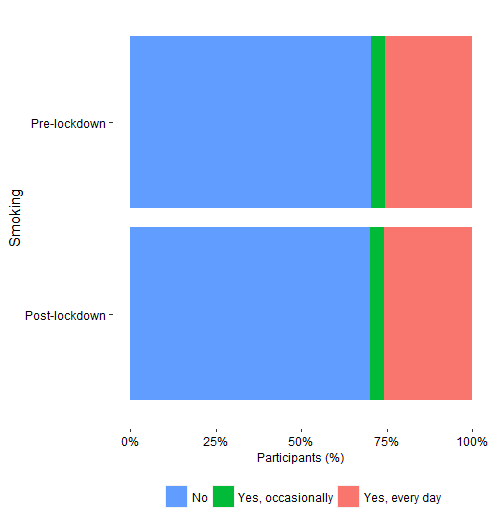

Figure S19 Parents’ smoking frequency before the lockdown (pre-lockdown) and following the re-opening of schools (post-lockdown).

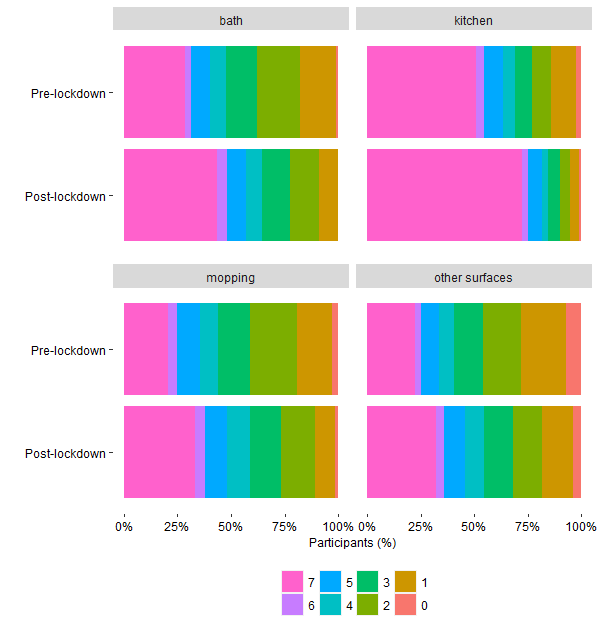

Figure S20 Parents’ cleaning frequency of home areas (times/week) before the lockdown (pre-lockdown) and following the re-opening of schools (post-lockdown).
