## Supplementary material for "Exposome changes in primary school children following the wide population non-pharmacological interventions implemented due to COVID-19 in Cyprus: a national survey": Deidentified data and script: data_prep_expo_school_.html

Exposome@school


### Exposome@school

The script was prepared for the data preparation for the statistical analysis of the data collected from the online survey Exposome at school (01/06/20 - 17/07/20).

From the raw data, variables on postal code, municipality and school were removed for protection of personal data.

### Distributions - raw

- Με πόσα άτομα έρχεται σε επαφή το παιδί σας από Δευτέρα μέχρι Παρασκευή, κατά μέσο όρο, ΑΥΤΗ ΤΗΝ ΠΕΡΙΟΔΟ, με τη σταδιακή άρση των περιοριστικών μέτρων;
  - the question refered to avergae no of contacts at home from Monday to Friday and not to the average no of contacts per day during weekdays....this applies also for the before period - so this could be interpreted with the average no of contacts for one day of the weekdays or the average no of contacts for all weekdays
- Κατά μέσο όρο, πόσες ώρες βρίσκεται στο σπίτι το παιδί σας το σαββατοκύριακο ΑΥΤΗ ΤΗΝ ΠΕΡΙΟΔΟ, με τη σταδιακή άρση των περιοριστικών μέτρων;
  - the question refered to average hours at home in weekend and not to the average number of hours per day during weekdays....this applies also for the before period
  - so this could be interpreted with the average no of hours at home for one of the two weekend days or the average no of hours at home for both days of the weekend
- Με πόσα άτομα έρχεται σε επαφή το παιδί σας από Δευτέρα μέχρι Παρασκευή, κατά μέσο όρο, ΑΥΤΗ ΤΗΝ ΠΕΡΙΟΔΟ, με τη σταδιακή άρση των περιοριστικών μέτρων;
  - the question refered to avergae no of contacts at home from Monday to Friday and not to the average no of contacts per day during weekdays....this applies also for the before period - so this could be interpreted with the average no of contacts for one day of the weekdays or the average no of contacts for all weekdays
- Κατά μέσο όρο, πόσες ώρες βρίσκεται στο σπίτι το παιδί σας το σαββατοκύριακο ΑΥΤΗ ΤΗΝ ΠΕΡΙΟΔΟ, με τη σταδιακή άρση των περιοριστικών μέτρων;
  - the question refered to average hours at home in weekend and not to the average number of hours per day during weekdays....this applies also for the before period
  - so this could be interpreted with the average no of hours at home for one of the two weekend days or the average no of hours at home for both days of the weekend

### Distributions - imputed and computed variables

### Session information

```
## R version 4.0.1 (2020-06-06)
## Platform: i386-w64-mingw32/i386 (32-bit)
## Running under: Windows 7 (build 7600)
## 
## Matrix products: default
## 
## locale:
## [1] LC_COLLATE=Greek_Greece.1253  LC_CTYPE=Greek_Greece.1253   
## [3] LC_MONETARY=Greek_Greece.1253 LC_NUMERIC=C                 
## [5] LC_TIME=Greek_Greece.1253    
## 
## attached base packages:
## [1] stats     graphics  grDevices utils     datasets  methods   base     
## 
## other attached packages:
##  [1] missMDA_1.17      PCAmixdata_3.1    factoextra_1.0.7  FactoMineR_2.3   
##  [5] ggpubr_0.4.0      glue_1.4.1        childsds_0.7.4    codebook_0.9.2   
##  [9] rmdformats_0.3.7  naniar_0.5.1      chron_2.3-55      haven_2.3.1      
## [13] inspectdf_0.0.8   NADA_1.6-1.1      FSA_0.8.30        scales_1.1.1     
## [17] reshape_0.8.8     texreg_1.37.4     janitor_2.0.1     Rcpp_1.0.4.6     
## [21] eeptools_1.2.4    lubridate_1.7.9   forcats_0.5.0     stringr_1.4.0    
## [25] dplyr_1.0.0       purrr_0.3.4       tidyverse_1.3.0   sjPlot_2.8.4     
## [29] psych_1.9.12.31   readr_1.3.1       stargazer_5.2.2   compare_0.2-6    
## [33] tableone_0.11.1   Hmisc_4.4-0       ggplot2_3.3.1     Formula_1.2-3    
## [37] survival_3.1-12   lattice_0.20-41   tibble_3.0.1      knitr_1.29       
## [41] tidyr_1.1.0       data.table_1.12.8 dtplyr_1.0.1      plyr_1.8.6       
## [45] readxl_1.3.1     
## 
## loaded via a namespace (and not attached):
##   [1] backports_1.1.7      sp_1.4-2             splines_4.0.1       
##   [4] TH.data_1.0-10       digest_0.6.25        foreach_1.5.0       
##   [7] htmltools_0.5.0      fansi_0.4.1          magrittr_1.5        
##  [10] checkmate_2.0.0      doParallel_1.0.15    cluster_2.1.0       
##  [13] openxlsx_4.1.5       ggfittext_0.9.0      modelr_0.1.8        
##  [16] sandwich_2.5-1       prettyunits_1.1.1    jpeg_0.1-8.1        
##  [19] colorspace_1.4-1     ggrepel_0.8.2        blob_1.2.1          
##  [22] rvest_0.3.5          mitools_2.4          xfun_0.15           
##  [25] crayon_1.3.4         jsonlite_1.6.1       lme4_1.1-23         
##  [28] iterators_1.0.12     zoo_1.8-8            gtable_0.3.0        
##  [31] emmeans_1.4.8        sjstats_0.18.0       sjmisc_2.8.5        
##  [34] car_3.0-8            abind_1.4-5          mvtnorm_1.1-1       
##  [37] DBI_1.1.0            rstatix_0.6.0        ggeffects_0.15.0    
##  [40] xtable_1.8-4         progress_1.2.2       performance_0.4.7   
##  [43] htmlTable_2.0.1      tmvnsim_1.0-2        flashClust_1.01-2   
##  [46] foreign_0.8-80       survey_4.0           vcd_1.4-7           
##  [49] htmlwidgets_1.5.1    httr_1.4.1           RColorBrewer_1.1-2  
##  [52] acepack_1.4.1        ellipsis_0.3.1       mice_3.9.0          
##  [55] pkgconfig_2.0.3      nnet_7.3-14          dbplyr_1.4.4        
##  [58] tidyselect_1.1.0     rlang_0.4.6          effectsize_0.3.1    
##  [61] munsell_0.5.0        cellranger_1.1.0     tools_4.0.1         
##  [64] cli_2.0.2            generics_0.0.2       sjlabelled_1.1.6    
##  [67] broom_0.5.6          evaluate_0.14        arm_1.11-1          
##  [70] yaml_2.2.1           fs_1.4.1             zip_2.0.4           
##  [73] visdat_0.5.3         nlme_3.1-148         leaps_3.1           
##  [76] xml2_1.3.2           compiler_4.0.1       rstudioapi_0.11     
##  [79] curl_4.3             png_0.1-7            ggsignif_0.6.0      
##  [82] reprex_0.3.0         statmod_1.4.34       stringi_1.4.6       
##  [85] parameters_0.8.0     Matrix_1.2-18        nloptr_1.2.2.1      
##  [88] vctrs_0.3.1          pillar_1.4.4         lifecycle_0.2.0     
##  [91] lmtest_0.9-37        estimability_1.3     maptools_1.0-1      
##  [94] insight_0.8.5        R6_2.4.1             latticeExtra_0.6-29 
##  [97] bookdown_0.20        rio_0.5.16           gridExtra_2.3       
## [100] codetools_0.2-16     boot_1.3-25          MASS_7.3-51.6       
## [103] assertthat_0.2.1     withr_2.2.0          mnormt_2.0.0        
## [106] multcomp_1.4-13      bayestestR_0.7.0     parallel_4.0.1      
## [109] hms_0.5.3            labelled_2.5.0       grid_4.0.1          
## [112] rpart_4.1-15         coda_0.19-3          minqa_1.2.4         
## [115] rmarkdown_2.3        snakecase_0.11.0     carData_3.0-4       
## [118] gamlss.dist_5.1-6    scatterplot3d_0.3-41 base64enc_0.1-3
```

×

- Distributions - raw
- Distributions - imputed and computed variables
- Session information
