## Supplementary material for "Exposome changes in primary school children following the wide population non-pharmacological interventions implemented due to COVID-19 in Cyprus: a national survey": Deidentified data and script: data_prep_expo_school_.Rmd

````
---
title: "Exposome@school"
output:
  rmdformats::html_clean
geometry: margin=1.5cm
editor_options: 
  chunk_output_type: console
---

The script was prepared for the data preparation for the statistical analysis of the data collected from the online survey Exposome at school (01/06/20 - 17/07/20).

From the raw data, variables on postal code, municipality and school were removed for protection of personal data.

```{r setup, include=FALSE}
knitr::opts_chunk$set(echo = TRUE)
```

```{r libraries, include=FALSE}

## Prepare workspace & install libraries
rm(list = ls(all = TRUE))

ipak <- function(pkg){
  new.pkg <- pkg[!(pkg %in% installed.packages()[, "Package"])]
  if (length(new.pkg)) 
    install.packages(new.pkg, dependencies = TRUE)
  sapply(pkg, require, character.only = TRUE)
}

# usage
packages <- c("readxl","plyr","dtplyr","data.table","tidyr","knitr",
              "tibble","Hmisc","tableone","compare","stargazer",
              "readr","psych","ggplot2","sjPlot","tidyverse","lubridate",
              "eeptools", "Rcpp", "janitor", "stringr","dplyr",
              "texreg","reshape","scales","FSA","NADA","inspectdf",
              "haven", "sjPlot", "forcats","chron","Hmisc","naniar",
              "rmdformats","codebook","childsds","glue","ggpubr","knitr",
              "FactoMineR", "factoextra","PCAmixdata","missMDA")

ipak(packages)

Sys.setlocale(category = "LC_ALL", locale = "Greek")


```


```{r data_reading, include=FALSE}

# The data file ".xls" was downloaded from REDCap on 20/07/20.  
# contains 4315 observations and 608 variables. 
# File has been saved as a csv file: "Exposomeschool_DATA_2020-07-20_1233_.csv"

# read data 
exsch_raw <- read.csv("rawdata/Exposomeschool_DATA_2020-07-20_1233_.csv", header=T, sep=",", encoding="UTF-8")  %>%
  clean_names ()

exsch_clean_a <- exsch_raw  %>%
  # include only completed questionnaires 
  filter(exposomeschool_complete == "2") %>%
  # convert characters to time variables
  mutate(submission_time = as.POSIXct(exposomeschool_timestamp, format="%Y-%m-%d %H:%M:%S")) %>%
  # rename ID variable
  dplyr::rename(ID = names(.)[1]) %>%
  # exclude test responses: before study 1/6/20 and IDs 2241 & 4309 
  filter(submission_time >=as.POSIXct("2020-06-01 22:25:26"),
         ID != "2241", ID != "4309") %>%
  # delete colunms with all NA 
  # but doesn't delete empty cols or cols with only zero values
  remove_empty("cols") #%>%
    # delete variables that are not useful for this analysis
  #dplyr::select(-c(covid_19_V2, covid_19_2, covid_19_3,exposomeschool_timestamp,
                   # remove empty columns and cols with only zero values
  #                exposomeschool4_timestamp,
  #                 exposomeschool4_complete,exposomeschool5_complete))

# str(exsch_clean_a, list.len=ncol(exsch_clean_a))

# create new variable for siblings (2 or 3 children in the same family) or not (1)
exsch_clean_a$sibling<-
   ifelse(
  (exsch_clean_a$exposomeschool2_complete==2 & exsch_clean_a$exposomeschool3_complete==0), 
  2, 
  ifelse(
  (exsch_clean_a$exposomeschool2_complete==2 & exsch_clean_a$exposomeschool3_complete==2), 
  3,  
  1))

exsch_clean_a$sibling<-as.factor(exsch_clean_a$sibling)
# summary(exsch_clean_a$sibling) 
# 1425 single kids, 42 x 2-siblings, 1 x 3-siblings > total=1512

# subset kid no 1 (for single kids and siblings) and select columns
exsch_singlekids <- exsch_clean_a  %>%
  dplyr::select(-contains(c("2","3"))) %>%
  dplyr::select(-"exposomeschool_complete")%>%
  # rename variables to make them same with variables names for siblings 
  rename(c("other_place_persons_wd_now"="otherplace_person_wd_now",
           "home_hours_weekend"="home_hours_we_now",
           "other_place_persons_we_now"="otherplace_person_we_now",
           "home_persons_before"="home_persons_wd_before",
           "otherplace_person_we_before"="other_place_persons_before"))

# str(exsch_singlekids, list.len=ncol(exsch_singlekids))

# subset kid no 2 (only for siblings 2 or 3) 
exsch_siblings<-exsch_clean_a  %>%
  filter(exposomeschool2_complete == "2" | exposomeschool3_complete == "2") %>%
dplyr::select(contains("2"), c("ID","num_child",
                  "smoking_now","cigs_per_day_now","cigs_per_week_now",
                  "bath_clean_anticept_now","product_bath",
                  "kitchen_anticeptic_now","product_kitchen",               
                  "mopping_anticeptic_now","product_mopping",              
                  "other_cleaning_now","product_other",
                  "smoking_before","cigs_per_day_before","cigs_per_week_before",
                  "bath_clean_anticept_before","product_bath_before",
                  "kitchen_anticeptic_before","product_kitchen_before",
                  "mopping_anticeptic_before","product_mopping_before",
                  "other_cleaning_before","product_other_before",
                  "city",
                  "sibling","submission_time")) %>%
    # rename variables so that variables have same names and can be rbinded 
 rename_at(vars(matches("2")), ~ str_remove (., "12"))  %>% 
    # delete variables that are not needed
   dplyr::select(-c("exposomeschool2_complete","exposomeschool2_timestamp")) %>%
      # rename variables so that you have same names and rbind them
  rename(c("vulnerable_weekdays"="vulnerable_wd_now",
           "home_persons_now11"="home_persons_now_we",
           "home_persons_before"="home_persons_we_before"))

# str(exsch_siblings, list.len=ncol(exsch_siblings))

# 1 variable missing "reason_sick_before" doesn't exist in 43 kids

# subset kid no 3 - only 1
exsch_siblings3<-exsch_clean_a  %>%
  filter(exposomeschool3_complete == "2") %>%
dplyr::select((contains("3")), c("ID","num_child",
                  "smoking_now","cigs_per_day_now","cigs_per_week_now",
                  "bath_clean_anticept_now","product_bath",
                  "kitchen_anticeptic_now","product_kitchen",               
                  "mopping_anticeptic_now","product_mopping",              
                  "other_cleaning_now","product_other",
                  "smoking_before","cigs_per_day_before","cigs_per_week_before",
                  "bath_clean_anticept_before","product_bath_before",
                  "kitchen_anticeptic_before","product_kitchen_before",
                  "mopping_anticeptic_before","product_mopping_before",
                  "other_cleaning_before","product_other_before",
                  "city",
                  "sibling","submission_time")) %>%
  # rename variables so that variables have same names and can be rbinded
 rename_at(vars(matches("3")), ~ str_remove (., "13"))  %>%
  # delete variables that are not needed
  dplyr::select(-c("exposomeschool3_complete","exposomeschool3_timestamp")) %>%
  # rename variables so that you have same names and rbind them
   rename(c("vulnerable_weekdays"="vulnerable_wd_now",
           "home_persons_now11"="home_persons_now_we",
           "home_persons_before"="home_persons_we_before"))

# 5 variables missing 
# "disease","reason_sick_before","reason_sick_now","where","test_covid" dont exist # in kid no 3
  
# str(exsch_siblings3, list.len=ncol(exsch_siblings3))
          
# rbind the 3 dataframes - exsch_singlekids, exsch_siblings, exsch_siblings3
df <- do.call("rbind.fill", list(exsch_singlekids, exsch_siblings, exsch_siblings3))

# str(df, list.len=ncol(df))

## check eligibility criteria - primary school children - age range: 5-14 years old

# check summary of year of birth variable
# summary(df$year_of_birth)
# in a few cases people added the birthdate (e.g. 26062013) instead of only birth year 
# so need to keep only last 4 digits for year variable

df_el<- df %>% 
# first create new year of birth variable and convert it to character
  mutate(year_of_birth_c=as.character(year_of_birth)) %>% 
# condition if birth year is larger than 2015 then keep only last 4 digits from birth year 
# character variable, else keep the birth year as is  
  mutate(birth_year=as.numeric(ifelse((year_of_birth>2015), 
  substr(year_of_birth_c, start=5, stop=nchar(year_of_birth_c)),
  year_of_birth)))  %>% 
# keep only eligible participants - only participants with years of birth 2006-2015
   filter(birth_year < 2016, birth_year > 2005)  %>% 
# delete year of birth variables not needed
dplyr::select(-c(year_of_birth_c, year_of_birth)) %>% 
# calculate age variable  
  mutate(age = 2020 - birth_year) %>%
# rename variables to make them more informative
  rename(c("weekend_before"="home_hours_we_before"))

# str(df_el, list.len=ncol(df_el))

# convert variables from integers into factors
df_el[, c("ID", "chronic_disease", "vaccine", "vaccine_doses","no_vaccination",
          "walking_now", "cycling_now","running_now", "swimming_now",
          "ballet_now", "ball_now","karate_now","other_activity_now",
          "free_time_now","break_now","sickness_now","breakfast_now",
          "fast_food_now","supplements_now","fruits_now","veggies_now",
          "meat_now","leggumes_now","fish_now","sugar_now","chips_now",
          "smoking_now","net_friends_now","net_family_now","tv_pc_now",
          "hands_soap_now","hands_anticeptic_now","walking_before",
          "cycling_before","running_before","swimming_before",
          "ballet_before","ball_before","karate_before","other_activity_before",
          "free_time_before","break_before","sickness_before",
          "breakfast_before","fast_food_before","supplements_before",
          "fruits_before","veggies_before","meat_before","leggumes_before",
          "fish_before","sugar_before","chips_before","smoking_before",
          "net_friends_before","net_family_before","tv_pc_before",
          "hands_soap_before","hands_anticeptic_before","sex",
          "place_of_birth","city","mother_education","father_education",
          "attend_school","consent","test_covid")]<-             
  lapply(df_el[, c("ID", "chronic_disease", "vaccine","vaccine_doses",
          "no_vaccination","walking_now", "cycling_now","running_now", "swimming_now",
          "ballet_now", "ball_now","karate_now","other_activity_now",
          "free_time_now","break_now","sickness_now","breakfast_now",
          "fast_food_now","supplements_now","fruits_now","veggies_now",
          "meat_now","leggumes_now","fish_now","sugar_now","chips_now",
          "smoking_now","net_friends_now","net_family_now","tv_pc_now",
          "hands_soap_now","hands_anticeptic_now","walking_before",
          "cycling_before","running_before","swimming_before",
          "ballet_before","ball_before","karate_before","other_activity_before",
          "free_time_before","break_before","sickness_before",
          "breakfast_before","fast_food_before","supplements_before",
          "fruits_before","veggies_before","meat_before","leggumes_before",
          "fish_before","sugar_before","chips_before","smoking_before",
          "net_friends_before","net_family_before","tv_pc_before",
          "hands_soap_before","hands_anticeptic_before","sex",
          "place_of_birth","city","mother_education","father_education",
          "attend_school","consent","test_covid")], factor)

# str(df_el, list.len=ncol(df_el))

# revalue variables levels to their actual names based on codebook

df_el <- df_el %>% 
 mutate_at(c("chronic_disease", "vaccine","no_vaccination",
             "sickness_now","supplements_now", 
             # for supplements_before the levels are 1,2,4,5
             "sickness_before"),
           .funs = function(t) revalue(x = t,
    replace = c(
    "1" = "Yes", 
    "2" = "No",
    "3" = "Don't know", 
    "4" = "Don't want to answer"))) %>% 
 mutate_at(c("walking_now", "cycling_now","running_now", "swimming_now",
             "ballet_now", "ball_now","karate_now","other_activity_now",
             "walking_before","cycling_before","running_before","swimming_before",
             "ballet_before","ball_before","karate_before","other_activity_before"),
           .funs = function(t) revalue(x = t,
    replace = c(
    "1" = "No",
    "2" = "1-2 times per week",
    "3" = "3-4 times per week",
    "4" = "5-6 times per week",
    "5" = "7 times or more per week",
    "6" = "Don't know",
    "7" = "Don't want to answer"))) %>%
  mutate_at(c("free_time_now","free_time_before"), 
                          .funs = function(t) revalue(x = t,
    replace = c(
    "1" = "She/he was doing things that involve little physical effort",  
    "2" = "1-2 times per week she/he did physical things (e.g. played sports,running,swimming)",
    "3" = "3-4 times per week she/he did physical things (e.g. played sports,running,swimming)",
    "4" = "5-6 times per week she/he did physical things (e.g. played sports,running,swimming)",
    "5" = "7 times or more per week she/he did physical things (e.g. played sports,running,swimming)",
    "6" = "Don't know",
    "7" = "Don't want to answer"))) %>%
  mutate_at(c("breakfast_now","breakfast_before",
              "fast_food_now","fast_food_before"), 
                          .funs = function(t) revalue(x = t,
    replace = c(
    "1" = "Every day",
    "2" = "4-6 times per week",
    "3" = "2-3 times per week",
    "4" = "1 time per week",
    "5" = "2-3 times per month",
    "6" = "Never",
    "7" = "Don't know",
    "8" = "Don't want to answer"))) %>%
  mutate_at(c("fruits_before","veggies_before","meat_before",
              "leggumes_before", "fish_before","sugar_before",
              "chips_before","fruits_now","veggies_now","meat_now",
              "leggumes_now","fish_now","sugar_now","chips_now"), 
                          .funs = function(t) revalue(x = t,
    replace = c(
    "1" = "Every day",
    "2" = "4-6 times per week",
    "3" = "2-3 times per week",
    "4" = "1 time per week",
    "5" = "2-3 times per month",
    "6" = "1 time per month",
    "7" = "Never",
    "8" = "Don't know",
    "9" = "Don't want to answer"))) %>%
  mutate_at(c("smoking_now","smoking_before"), 
                          .funs = function(t) revalue(x = t,
    replace = c(
    "1" = "Yes, every day",
    "2" = "Yes, occasionally",
    "3" = "No",
    "4" = "Don't know/don't remember/not sure",
    "5" = "Don't want to answer"))) %>%
  mutate_at(c("net_friends_now","net_family_now",
              "net_friends_before","net_family_before"), 
                          .funs = function(t) revalue(x = t,
    replace = c(
    "1" = "Every day",
    "2" = "Every week but not every day",
    "3" = "Some times in the month but not every week",
    "4" = "Never",
    "5" = "Don't know",
    "6" = "Don't want to answer"))) %>%
  mutate_at(c("tv_pc_now","tv_pc_before"), 
                          .funs = function(t) revalue(x = t,
    replace = c(
    "1" = "Less than 1 hour per day",
    "2" = "1-3 hours per day",
    "3" = "4-7 hours per day",
    "4" = "8-11 hours per day",
    "5" = "More than 11 hours per day",
    "6" = "Don't know",
    "7" = "Don't want to answer"))) %>%
  mutate_at(c("hands_soap_now","hands_anticeptic_now",
              "hands_soap_before","hands_anticeptic_before"), 
                          .funs = function(t) revalue(x = t,
    replace = c(
    "1" = "Never",
    "2" = "Less than 1 time per day",
    "3" = "1-3 times per day",
    "4" = "4-7 times per day",
    "5" = "More than 7 times per day",
    "6" = "Don't know",
    "7" = "Don't want to answer"))) %>%
  mutate_at(c("mother_education","father_education"), 
                          .funs = function(t) revalue(x = t,
    replace = c(
    "1" = "Has never been to school",
    "2" = "Has not completed primary school",
    "3" = "Primary School",
    "4" = "Middle School (3 years)",
    "5" = "High School/Vocational High School (diploma)",
    "6" = "Higher (after high school) non-tertiary Education",
    "7" = "Higher Tertiary Education (non-Univesity)",
    "8" = "University (Bachelor degree)",
    "9" = "University-Postgraduate (only Master's degree)",
    "10" = "PhD",
    "11" = "Don't know/Don't want to answer")))

df_el$break_now <- revalue (df_el$break_now,
                c(
    "1" = "Sat down (talking, reading, eating)",
    "2" = "Stood around or walked around",
    "3" = "Ran or played a little bit",
    "4" = "Ran around and played quite a bit",
    "5" = "Ran and played hard most of the time",
    "6" = "Don't know",
    "7" = "Don't want to answer"))            

df_el$break_before <- revalue (df_el$break_before,
                c(
    "1" = "Sat down (talking, reading)",
    "2" = "Stood around or walked around",
    "3" = "Ran or played a little bit",
    "4" = "Ran around and played quite a bit",
    "5" = "Ran and played hard most of the time",
    "6" = "Eating",
    "7" = "Don't know",
    "8" = "Don't want to answer")) 

#### The variables of break for now and before have different levels
# The before variable has the extra option "eating" whereas for break_now "eating" is included in the first category "Sat down (talking, reading, eating)" - transformed break_before: the eating category was added in the first category sat down so that before and after are the same

# add the option "Eating" in the first category "Sat down" so that break_before & break_now have same levels
df_el$break_before <- revalue (df_el$break_before,
                c(
    "Sat down (talking, reading)" = "Sat down (talking, reading, eating)",
    "Eating" = "Sat down (talking, reading, eating)"))

df_el$vaccine_doses <- revalue(df_el$vaccine_doses,
# 1, Ναι | 2, Όχι γιατί το εμβόλιο δεν ήταν διαθέσιμο | 3, Όχι γιατί θεωρήσαμε ότι δεν χρειαζεται # | 4, Όχι γιατί το αμελήσαμε αλλά θα τις κάνει | 5, Δεν χρειαζόταν πάνω από μία δόσεις στο/στα  # εμβόλιο/εμβόλια που έκανε | 6, Δεν γνωρίζω | 7, Δεν απαντώ
  c("1" = "Yes",
    "2" = "No, vaccine not available",
    "3" = "No, we thought that more than one dose wasn't needed",
    "4" = "No, we neglected but they will be provided",
    "5" = "No, only one dose was needed",
    "6" = "Don't know",
    "7" = "Don't want to answer"))

df_el$supplements_before <- revalue(df_el$supplements_before,
  c("1" = "Yes",
    "2" = "No",
    "4" = "Don't know",
    "5" = "Don't want to answer"))

df_el$sex <- revalue(df_el$sex, 
                    c("1"="Male", 
                      "2"="Female",
                      "3"="Don't want to answer"))

df_el$place_of_birth <- revalue(df_el$place_of_birth, 
                    c("1"="Cyprus", 
                      "2"="EU country",
                      "3"="Non-EU country"))

df_el$city <- revalue(df_el$city, 
                    c("1"="Limassol", 
                      "2"="Paphos",
                      "3"="Larnaca",
                      "4"="Nicosia",
                      "5"="Famagusta"))

df_el$attend_school <- revalue(df_el$attend_school, 
                    c("1"="Goes to school", 
                      "2"="Stays at home"))

df_el$consent <- revalue(df_el$consent, 
                    c("1"="Yes", 
                      "2"="No"))

df_el$test_covid <- revalue(df_el$test_covid, 
                    c("1"="Negative", 
                      "2"="Positive",
                      "3"="Don't know",
                      "4"="Don't want to answer"))

# str(df_el, list.len=ncol(df_el))
```


# Distributions - raw

```{r distributions, echo=FALSE}

# check distributions using QQ plots and summaries

# inspect variables using package inspect df functions

# inspect_cat(df_el)   ##error
# inspect_imb(df_el, df2 = NULL, include_na = FALSE)   ##error

# summary(df_el$age)
qqnorm(df_el$age,main="age")

# summary(df_el$num_child)
# Min. 1st Qu.  Median    Mean 3rd Qu.    Max.    NA's 
# 0.000   1.000   1.000   1.367   2.000   5.000      12
qqnorm(df_el$num_child,main="children_no_primary_sch")
# non-normal

# summary(df_el$chronic_disease)
# summary(df_el$vaccine)
# summary(df_el$vaccine_doses)
# summary(df_el$no_vaccination)

# summary(df_el$walking_now)
# summary(df_el$cycling_now)
# summary(df_el$running_now)
# summary(df_el$swimming_now)
# summary(df_el$ballet_now)
# summary(df_el$ball_now)
# summary(df_el$karate_now)
# summary(df_el$other_activity_now)

# summary(df_el$free_time_now)
# summary(df_el$break_now)
# summary(df_el$sickness_now)

# summary(df_el$breakfast_now)
# summary(df_el$fast_food_now)
# summary(df_el$supplements_now)
# summary(df_el$fruits_now)
# summary(df_el$fruits_per_day_now)
# Min. 1st Qu.  Median    Mean 3rd Qu.    Max.    NA's 
# 0.000   1.000   2.000   1.962   2.000  15.000      54
qqnorm(df_el$fruits_per_day_now,main="fruit_portions_perday_now")
# non-normal

# summary(df_el$veggies_now)
# summary(df_el$veggies_per_day_now)
#  Min. 1st Qu.  Median    Mean 3rd Qu.    Max.    NA's 
# 0.000   1.000   1.000   1.681   2.000  13.000     100 
qqnorm(df_el$veggies_per_day_now,main="veggies_portions_perday_now")
# non-normal

# summary(df_el$meat_now)
# summary(df_el$meat_per_day_now)
#  Min.  1st Qu.   Median     Mean  3rd Qu.     Max.     NA's 
#  0.00     1.00     1.00    13.82     2.00 16790.00       11
qqnorm(df_el$meat_per_day_now,main="meat_portions_perday_now")
# non-normal
# sum(df_el$meat_per_day_now>20,na.rm=TRUE)
# 14

# summary(df_el$leggumes_now)
# summary(df_el$leggumes_per_day_now)
# Min. 1st Qu.  Median    Mean 3rd Qu.    Max.    NA's 
# 0.000   1.000   1.000   2.327   1.000 180.000      82
qqnorm(df_el$leggumes_per_day_now,main="leggumes_portions_perday_now")
# non-normal
# sum(df_el$leggumes_per_day_now>20,na.rm=TRUE)
# 12

# summary(df_el$fish_now)
# summary(df_el$fish_per_day)
# Min. 1st Qu.  Median    Mean 3rd Qu.    Max.    NA's 
# 0.000   1.000   1.000   1.939   1.000 180.000     131
qqnorm(df_el$fish_per_day,main="fish_portions_perday_now")
# non-normal
# sum(df_el$fish_per_day>20,na.rm=TRUE)
# 10

# summary(df_el$sugar_now)
# summary(df_el$sugar_per_day_now)
# Min. 1st Qu.  Median    Mean 3rd Qu.    Max.    NA's 
# 0.000   1.000   1.000   2.156   2.000 200.000      11
qqnorm(df_el$sugar_per_day_now,main="sugar_portions_perday_now")
# non-normal
# sum(df_el$sugar_per_day_now>20,na.rm=TRUE)
# 5

# summary(df_el$chips_now)
# summary(df_el$chips_per_day_now)
# Min. 1st Qu.  Median    Mean 3rd Qu.    Max.    NA's 
# 0.000   1.000   1.000   1.259   1.000  90.000      66 
qqnorm(df_el$chips_per_day_now,main="chips_portions_perday_now")
# non-normal
# sum(df_el$chips_per_day_now>20,na.rm=TRUE)
# 1

# summary(df_el$smoking_now)
# summary(df_el$cigs_per_day_now)
qqnorm(df_el$cigs_per_day_now,main="cigarettes_no_perday_now")
# non-normal

# summary(df_el$cigs_per_week_now)
qqnorm(df_el$cigs_per_week_now,main="cigarettes_no_perweek_now")
# non-normal

# summary(df_el$net_friends_now)
# summary(df_el$hours_friends_now)
#   Min. 1st Qu.  Median    Mean 3rd Qu.    Max.    NA's 
#  0.000   1.000   1.000   1.977   2.000  30.000     825 
qqnorm(df_el$hours_friends_now,main="friends_communic_hours_perday_now")
# non-normal
# sum(df_el$hours_friends_now>24,na.rm=TRUE)
# 2

# summary(df_el$net_family_now)
# summary(df_el$hours_family_now)
qqnorm(df_el$hours_family_now,main="family_communic_hours_perday_now")
# non-normal

# summary(df_el$tv_pc_now)

# summary(df_el$hands_soap_now)
# summary(df_el$hands_anticeptic_now)

# summary(df_el$bath_clean_anticept_now)
# Min. 1st Qu.  Median    Mean 3rd Qu.    Max. 
# 0.000   3.000   5.000   4.865   7.000  47.000
qqnorm(df_el$bath_clean_anticept_now,main="bath_clean_days_perweek_now")
# non-normal
# sum(df_el$bath_clean_anticept_now>7,na.rm=TRUE)
# 6

# summary(df_el$kitchen_anticeptic_now)
# Min. 1st Qu.  Median    Mean 3rd Qu.    Max. 
# 0.000   6.000   7.000   6.036   7.000  30.000
qqnorm(df_el$kitchen_anticeptic_now,main="kitchen_clean_days_perweek_now")
# non-normal
# sum(df_el$kitchen_anticeptic_now>7,na.rm=TRUE)
# 10

# summary(df_el$mopping_anticeptic_now)
# Min. 1st Qu.  Median    Mean 3rd Qu.    Max. 
# 0.000   2.000   4.000   4.492   7.000  66.000
qqnorm(df_el$mopping_anticeptic_now,main="mopping_days_perweek_now")
# non-normal
# sum(df_el$mopping_anticeptic_now>7,na.rm=TRUE)
# 7

# summary(df_el$other_cleaning_now)
# Min. 1st Qu.  Median    Mean 3rd Qu.    Max. 
# 0.00    2.00    4.00    4.16    7.00   20.00
qqnorm(df_el$other_cleaning_now,main="othersurfaces_clean_days_perweek_now")
# non-normal
# sum(df_el$other_cleaning_now>7,na.rm=TRUE)
# 2

# summary(df_el$home_hours_wd)
# Min. 1st Qu.  Median    Mean   3rd Qu.  Max. 
# 0.00   10.00   15.50   16.33   20.00 1510.00
qqnorm(df_el$home_hours_wd,main="stayinghome_hoursperday_weekdays_now")
# non-normal
# sum(df_el$home_hours_wd>120,na.rm=TRUE) #24*5=120
# 11

# summary(df_el$home_persons_wd_now)
# Min. 1st Qu.  Median    Mean   3rd Qu.  Max. 
# 0.000   3.000   4.000   4.788   5.000  51.000
qqnorm(df_el$home_persons_wd_now,main="contacts_home_weekdays_now")
# non-normal
# sum(df_el$home_persons_wd_now>20,na.rm=TRUE)
# 10

# summary(df_el$vulnerable_wd_now)
#  Min. 1st Qu.  Median    Mean   3rd Qu. Max. 
# 0.000   0.000   1.000   1.331   2.000  27.000
qqnorm(df_el$vulnerable_wd_now,main="vulner_contacts_home_weekdays_now")
# non-normal
# sum(df_el$vulnerable_wd_now>20,na.rm=TRUE)
# 1

# summary(df_el$school_persons_now)
# Min. 1st Qu.  Median    Mean   3rd Qu.  Max.    NA's 
# 0.000   5.000  10.000   9.846  12.000 150.000      39
qqnorm(df_el$school_persons_now,main="contacts_school_weekdays_now")
# non-normal

# summary(df_el$otherplace_person_wd_now)
# Min. 1st Qu.  Median    Mean   3rd Qu.  Max. 
# 0.000   2.000   4.000   5.834   7.000 524.000
qqnorm(df_el$otherplace_person_wd_now,main="contacts_other_weekdays_now")
# non-normal
# sum(df_el$otherplace_person_wd_now>100,na.rm=TRUE)
# 1

# summary(df_el$home_hours_we_now)
# Min. 1st Qu.  Median    Mean 3rd Qu.    Max. 
# 0.00   12.00   20.00   19.37   24.00  307.00
qqnorm(df_el$home_hours_we_now,main="stayinghome_weekend_now")
# non-normal
# sum(df_el$home_hours_we_now>48,na.rm=TRUE)
# 6

# summary(df_el$home_persons_now_we)
# Min. 1st Qu.  Median    Mean 3rd Qu.    Max. 
# 0.000   3.000   4.000   5.197   6.000 150.000
qqnorm(df_el$home_persons_now_we,main="contacts_home_weekend_now")
# non-normal
# sum(df_el$home_persons_now_we>100,na.rm=TRUE)
# 1

# summary(df_el$vulnerable_weekend_now)
# Min. 1st Qu.  Median    Mean 3rd Qu.    Max. 
# 0.000   0.000   1.000   1.447   2.000  50.000
qqnorm(df_el$vulnerable_weekend_now,main="vulner_contacts_home_weekend_now")
# non-normal
# sum(df_el$vulnerable_weekend_now>20,na.rm=TRUE)
# 1

# summary(df_el$otherplace_person_we_now)
#  Min. 1st Qu.  Median    Mean 3rd Qu.    Max. 
#  0.000   2.000   4.000   4.813   6.000  30.000
qqnorm(df_el$otherplace_person_we_now,main="contacts_other_weekend_now")
# non-normal

# summary(df_el$walking_before)
# summary(df_el$cycling_before)
# summary(df_el$running_before)
# summary(df_el$swimming_before)
# summary(df_el$ballet_before)
# summary(df_el$ball_before)
# summary(df_el$karate_before)
# summary(df_el$other_activity_before)

# summary(df_el$free_time_before)
# summary(df_el$break_before)
# summary(df_el$sickness_before)

# summary(df_el$breakfast_before)
# summary(df_el$fast_food_before)
# summary(df_el$supplements_before)
# summary(df_el$fruits_before)
# summary(df_el$fruits_per_day_before)
# Min. 1st Qu.  Median    Mean 3rd Qu.    Max.    NA's 
# 0.000   1.000   1.000   1.953   2.000  20.000      50 
qqnorm(df_el$fruits_per_day_before,main="fruit_portions_perday_before")
# non-normal

# summary(df_el$veggies_before)
# summary(df_el$veggies_per_day_before)
# Min. 1st Qu.  Median    Mean 3rd Qu.    Max.    NA's 
# 0.000   1.000   1.000   1.695   2.000  30.000     104
qqnorm(df_el$veggies_per_day_before,main="veggies_portions_perday_before")
# non-normal
# sum(df_el$veggies_per_day_before>20,na.rm=TRUE)
# 2

# summary(df_el$meat_before)
# summary(df_el$meat_per_day_before)
# Min. 1st Qu.  Median    Mean 3rd Qu.    Max.    NA's 
# 0.00    1.00    1.00    2.28    2.00  270.00       9
qqnorm(df_el$meat_per_day_before,main="meat_portions_perday_before")
# non-normal
# sum(df_el$meat_per_day_before>20,na.rm=TRUE)
# 8

# summary(df_el$leggumes_before)
# summary(df_el$leggumes_per_day_before)
# Min. 1st Qu.  Median    Mean 3rd Qu.    Max.    NA's 
# 0.000   1.000   1.000   2.033   1.000 270.000      82
qqnorm(df_el$leggumes_per_day_before,main="leggumes_portions_perday_before")
# non-normal
# sum(df_el$leggumes_per_day_before>20,na.rm=TRUE)
# 8

# summary(df_el$fish_before)
# summary(df_el$fish_per_day_before)
# Min. 1st Qu.  Median    Mean 3rd Qu.    Max.    NA's 
# 0.000   1.000   1.000   1.715   1.000 180.000     131
qqnorm(df_el$fish_per_day_before,main="fish_portions_perday_before")
# non-normal
# sum(df_el$fish_per_day_before>20,na.rm=TRUE)
# 7

# summary(df_el$sugar_before)
# summary(df_el$sugar_per_day_before)
# Min. 1st Qu.  Median    Mean 3rd Qu.    Max.    NA's 
#  0.000   1.000   1.000   1.737   2.000  45.000      10 
qqnorm(df_el$sugar_per_day_before,main="sugar_portions_perday_before")
# non-normal
# sum(df_el$sugar_per_day_before>20,na.rm=TRUE)
# 2

# summary(df_el$chips_before)
# summary(df_el$chips_per_day_before)
#   Min. 1st Qu.  Median    Mean 3rd Qu.    Max.    NA's 
#  0.000   1.000   1.000   1.218   1.000  20.000      61 
qqnorm(df_el$chips_per_day_before,main="chips_portions_perday_before")
# non-normal

# summary(df_el$smoking_before)
# summary(df_el$cigs_per_day_before)
qqnorm(df_el$cigs_per_day_before,main="cigarettes_no_perday_before")
# non-normal

# summary(df_el$cigs_per_week_before)
qqnorm(df_el$cigs_per_week_before,main="cigarettes_no_perweek_before")
# non-normal

# summary(df_el$net_friends_before)
# summary(df_el$hours_friends_before)
qqnorm(df_el$hours_friends_before,main="friends_communic_hours_perweek_before")
# non-normal

# summary(df_el$net_family_before)
# summary(df_el$hours_family_before)
qqnorm(df_el$hours_family_before,main="family_communic_hours_perweek_before")
# non-normal

# summary(df_el$tv_pc_before)

# summary(df_el$hands_soap_before)
# summary(df_el$hands_anticeptic_before)

# summary(df_el$bath_clean_anticept_before)
# Min. 1st Qu.  Median    Mean 3rd Qu.    Max. 
# 0.000   2.000   3.000   3.956   7.000  36.000
qqnorm(df_el$bath_clean_anticept_before,main="bath_clean_days_perweek_before")
# non-normal
# sum(df_el$bath_clean_anticept_before>7,na.rm=TRUE)
# 3

# summary(df_el$kitchen_anticeptic_before)
# Min. 1st Qu.  Median    Mean 3rd Qu.    Max. 
# 0.000   3.000   7.000   4.998   7.000  20.000
qqnorm(df_el$kitchen_anticeptic_before,main="kitchen_clean_days_perweek_before")
# non-normal
# sum(df_el$kitchen_anticeptic_before>7,na.rm=TRUE)
# 3

# summary(df_el$mopping_anticeptic_before)
# Min. 1st Qu.  Median    Mean 3rd Qu.    Max. 
# 0.000   2.000   3.000   3.632   5.000  20.000
qqnorm(df_el$mopping_anticeptic_before,main="mopping_days_perweek_before")
# non-normal
# sum(df_el$mopping_anticeptic_before>7,na.rm=TRUE)
# 2

# summary(df_el$other_cleaning_before)
# Min. 1st Qu.  Median    Mean 3rd Qu.    Max. 
# 0.000   1.000   3.000   3.413   6.000   8.000
qqnorm(df_el$other_cleaning_before,main="othersurfaces_clean_days_perweek_before")
# non-normal
# sum(df_el$other_cleaning_before>7,na.rm=TRUE)
# 1

# summary(df_el$home_hours_wd_before)
#   Min. 1st Qu.  Median    Mean 3rd Qu.    Max. 
#   0.00    7.00   12.00   14.82   15.00 5015.00
qqnorm(df_el$home_hours_wd_before,main="stayinghome_hoursperday_weekdays_before")
# non-normal
# sum(df_el$home_hours_wd_before>24,na.rm=TRUE)
# 11

# summary(df_el$home_persons_wd_before)
#  Min. 1st Qu.  Median    Mean 3rd Qu.    Max. 
#  0.00    4.00    6.00   12.32   12.00  400.00
qqnorm(df_el$home_persons_wd_before,main="contacts_home_weekdays_before")
# non-normal
# sum(df_el$home_persons_wd_before>100,na.rm=TRUE)
# 6

# summary(df_el$vulnerable_weekdays_before)
# Min. 1st Qu.  Median    Mean 3rd Qu.    Max. 
# 0.000   1.000   2.000   2.663   3.000 430.000
qqnorm(df_el$vulnerable_weekdays_before,main="vulner_contacts_home_weekdays_before")
# non-normal
# sum(df_el$vulnerable_weekdays_before>100,na.rm=TRUE)
# 1

# summary(df_el$school_persons_before)
# Min.   1st Qu.    Median      Mean   3rd Qu.      Max. 
# 0.000e+00 1.500e+01 2.400e+01 3.679e+09 3.000e+01 5.556e+12
qqnorm(df_el$school_persons_before,main="contacts_school_weekdays_before")
# non-normal
# sum(df_el$school_persons_before>300,na.rm=TRUE)
# 5

# summary(df_el$otherplace_person_wd_before)
# Min. 1st Qu.  Median    Mean 3rd Qu.    Max. 
# 0.00    5.00   10.00   16.15   20.00  412.00
qqnorm(df_el$otherplace_person_wd_before,main="contacts_other_weekdays_before")
# non-normal
# sum(df_el$otherplace_person_wd_before>200,na.rm=TRUE)
# 1

# summary(df_el$home_hours_we_before)
# Min. 1st Qu.  Median    Mean 3rd Qu.    Max. 
# 0.00   10.00   15.00   17.18   19.00 2415.00
qqnorm(df_el$home_hours_we_before,main="stayinghome_hoursperday_weekend_before")
# non-normal
# sum(df_el$home_hours_we_before>48,na.rm=TRUE)
# 8

# summary(df_el$home_persons_we_before)
#  Min. 1st Qu.  Median    Mean 3rd Qu.    Max.    NA's 
#  0.000   4.000   7.000   9.048  10.000 105.000      44 
qqnorm(df_el$home_persons_we_before,main="contacts_home_weekend_before")
# non-normal
# sum(df_el$home_persons_we_before>100,na.rm=TRUE)
# 1

# summary(df_el$vulnerable_weekend_before)
# Min. 1st Qu.  Median    Mean 3rd Qu.    Max. 
# 0.000   1.000   2.000   2.751   3.000 390.000
qqnorm(df_el$vulnerable_weekend_before,main="vulner_contacts_home_weekend_before")
# non-normal
# sum(df_el$vulnerable_weekend_before>100,na.rm=TRUE)
# 2

# summary(df_el$other_place_persons_before)
#   Min.   1st Qu.    Median      Mean   3rd Qu.      Max. 
# 0.000e+00 5.000e+00 1.000e+01 3.701e+11 1.500e+01 5.589e+14
qqnorm(df_el$other_place_persons_before,main="contacts_other_weekend_before")
# non-normal
# sum(df_el$other_place_persons_before>200,na.rm=TRUE)
# 2

# summary(df_el$sex)
# summary(df_el$place_of_birth)

# summary(df_el$city)
# summary(df_el$years_living_in_cyprus)
#  Min. 1st Qu.  Median    Mean 3rd Qu.    Max. 
#   0.00    8.00    9.00   15.85   11.00 8011.00 
qqnorm(df_el$years_living_in_cyprus,main="years_living_in_Cyprus")
# non-normal
# sum(df_el$years_living_in_cyprus>14,na.rm=TRUE)
# 2

# summary(df_el$birth_year)

# summary(df_el$mother_education)
# summary(df_el$father_education)

# summary(df_el$height)
# Min. 1st Qu.  Median    Mean 3rd Qu.    Max.    NA's 
#    0.0   126.0   138.0   130.9   150.0   500.0      81 
qqnorm(df_el$height,main="height")
# non-normal
# sum(df_el$height<80,na.rm=TRUE)
# 2
# sum(df_el$height==0,na.rm=TRUE)
# 1
# sum(df_el$height>200,na.rm=TRUE)
# 1

# summary(df_el$weight)
#  Min. 1st Qu.  Median    Mean 3rd Qu.    Max.    NA's 
#  0.00   27.00   33.00   35.59   40.00  230.00      62
qqnorm(df_el$weight,main="weight")
# non-normal
# sum(df_el$weight>200,na.rm=TRUE)
# 1
# sum(df_el$weight==0,na.rm=TRUE)
# 1

# summary(df_el$attend_school)
# summary(df_el$consent)
# summary(df_el$test_covid)

```

# Distributions - imputed and computed variables

```{r distributions_imp, echo=FALSE, message = FALSE}

# replace all don't know, don't want to answer to NA
df_imp <- df_el %>% 
 mutate_at(c("chronic_disease","vaccine","no_vaccination","vaccine_doses",
             "sickness_now","supplements_now", 
             "supplements_before","sickness_before",
             "walking_now", "cycling_now","running_now","swimming_now",
             "ballet_now", "ball_now","karate_now","other_activity_now",
             "walking_before","cycling_before","running_before",
             "swimming_before","ballet_before","ball_before",
             "karate_before","other_activity_before",
             "free_time_now","free_time_before",
             "breakfast_now","breakfast_before",
             "fast_food_now","fast_food_before",
             "fruits_before","veggies_before","meat_before",
             "leggumes_before", "fish_before","sugar_before",
             "chips_before","fruits_now","veggies_now","meat_now",
             "leggumes_now","fish_now","sugar_now","chips_now",
             "smoking_now","smoking_before",
             "net_friends_now","net_family_now",
             "net_friends_before","net_family_before",
             "tv_pc_now","tv_pc_before",
             "hands_soap_now","hands_anticeptic_now",
             "hands_soap_before","hands_anticeptic_before",
             "mother_education","father_education","sex","test_covid",
             "break_now","break_before"),
           .funs = function(t) revalue(x = t,
    replace = c(
    "Don't know" = NA, 
    "Don't want to answer" = NA,
    "Don't know/don't remember/not sure" = NA,
    "Don't know/Don't want to answer" = NA)))  %>% 
# replace extreme values with NA
  replace_with_na_at(c("other_place_persons_before","otherplace_person_wd_before",
                       "otherplace_person_wd_now","home_persons_now_we",
                       "vulnerable_weekdays_before","vulnerable_weekend_before"),
                     condition = ~.x > 100) %>%
  replace_with_na_at(c("school_persons_before"),
                     condition = ~.x > 300) %>% 
  replace_with_na_at(c("home_hours_we_now","home_hours_we_before"), # 24*2 = 48
                     condition = ~.x > 48) %>% 
  replace_with_na_at(c("bath_clean_anticept_now","bath_clean_anticept_before",
                       "kitchen_anticeptic_now","kitchen_anticeptic_before",
                       "mopping_anticeptic_now","mopping_anticeptic_before",
                       "other_cleaning_now","other_cleaning_before"),
                     condition = ~.x > 7) %>% 
  replace_with_na_at(c("home_hours_wd",
                       "home_hours_wd_before"), # 24*5 = 120
                     condition = ~.x > 120) %>% 
  replace_with_na_at(c("meat_per_day_now","meat_per_day_before",
                       "leggumes_per_day_now","leggumes_per_day_before",
                       "fish_per_day","fish_per_day_before",
                       "sugar_per_day_now","sugar_per_day_before",
                       "chips_per_day_now","veggies_per_day_before"),
                     condition = ~.x > 20) %>% 
  replace_with_na_at("hours_friends_now",
                     condition = ~.x > 24) %>% 
  replace_with_na_at("num_child",
                     condition = ~.x == 0) %>% 
  replace_with_na_at("years_living_in_cyprus",
                     condition = ~.x > 14) %>% 
  replace_with_na_at(c("height","weight"),
                     condition = ~.x > 200) %>% 
  replace_with_na_at(c("height","weight"),
                     condition = ~.x == 0) %>% 
  replace_with_na_at(c("height"),
                     condition = ~.x == 70) %>% 
# in a few cases people added the height in m (e.g. 1.56) instead of cm 
# so for these cases need to multiply *100
# if height is smaller than 2 then multiply *100, else keep the height as is  
  mutate(heightcm=ifelse((height < 2), 
         height * 100,
         height)) 

# check again distributions of variables for which extreme values have been replaced with NA

hist(df_imp$other_place_persons_before,main="contacts_other_weekend_before")
# non-normal
# summary(df_imp$other_place_persons_before)
# Min. 1st Qu.  Median    Mean 3rd Qu.    Max.    NA's 
# 0.00    5.00   10.00   10.93   15.00  100.00       2
hist(df_imp$otherplace_person_wd_before,main="contacts_other_weekday_before")
# non-normal
# summary(df_imp$otherplace_person_wd_before)
# Min. 1st Qu.  Median    Mean 3rd Qu.    Max.    NA's 
# 0.00    5.00   10.00   15.48   20.00  100.00       5
hist(df_imp$otherplace_person_wd_now,main="contacts_other_weekend_now")
# non-normal
# summary(df_imp$otherplace_person_wd_now)
# Min. 1st Qu.  Median    Mean 3rd Qu.    Max.    NA's 
# 0.00    2.00    4.00    5.49    7.00  100.00       1
hist(df_imp$home_persons_now_we,main="contacts_home_weekend_now")
# non-normal
# summary(df_imp$home_persons_now_we)
# Min. 1st Qu.  Median    Mean 3rd Qu.    Max.    NA's 
# 0.000   3.000   4.000   5.101   6.000  64.000       1 
hist(df_imp$vulnerable_weekdays_before,main="vuln_contacts_home_weekdays_before")
# non-normal
# summary(df_imp$vulnerable_weekdays_before)
# Min. 1st Qu.  Median    Mean 3rd Qu.    Max.    NA's 
# 0.00    1.00    2.00    2.38    3.00   40.00       1
hist(df_imp$vulnerable_weekend_before,main="vuln_contacts_home_weekend_before")
# non-normal
# summary(df_imp$vulnerable_weekend_before)
# Min. 1st Qu.  Median    Mean 3rd Qu.    Max.    NA's 
# 0.000   1.000   2.000   2.357   3.000  50.000       2
hist(df_imp$school_persons_before,main="contacts_school_weekday_before")
# non-normal
# summary(df_imp$school_persons_before)
# Min. 1st Qu.  Median    Mean 3rd Qu.    Max.    NA's 
# 0.0    15.0    24.0    28.1    30.0   300.0       5
hist(df_imp$home_hours_we_now,main="stayinghome_weekend_now")
# non-normal
# summary(df_imp$home_hours_we_now)
# Min. 1st Qu.  Median    Mean 3rd Qu.    Max.    NA's 
# 0.00   12.00   20.00   18.66   24.00   48.00       6
hist(df_imp$home_hours_we_before,main="stayinghome_weekend_before")
# non-normal
# summary(df_imp$home_hours_we_before)
# Min. 1st Qu.  Median    Mean 3rd Qu.    Max.    NA's 
# 0.00   10.00   15.00   14.87   19.00   48.00       8
hist(df_imp$bath_clean_anticept_now,main="bath_clean_days_perweek_now")
# non-normal
# summary(df_imp$bath_clean_anticept_now)
# Min. 1st Qu.  Median    Mean 3rd Qu.    Max.    NA's 
# 0.000   3.000   5.000   4.812   7.000   7.000       6
hist(df_imp$bath_clean_anticept_before,main="bath_clean_days_perweek_before")
# non-normal
# summary(df_imp$bath_clean_anticept_before)
# Min. 1st Qu.  Median    Mean 3rd Qu.    Max.    NA's 
# 0.000   2.000   3.000   3.913   7.000   7.000       3
hist(df_imp$kitchen_anticeptic_now,main="kitchen_clean_days_perweek_now")
# non-normal
# summary(df_imp$kitchen_anticeptic_now)
# Min. 1st Qu.  Median    Mean 3rd Qu.    Max.    NA's 
# 0.000   6.000   7.000   5.971   7.000   7.000      10
hist(df_imp$kitchen_anticeptic_before,main="kitchen_clean_days_perweek_before")
# non-normal
# summary(df_imp$kitchen_anticeptic_before)
# Min. 1st Qu.  Median    Mean 3rd Qu.    Max.    NA's 
# 0.000   3.000   7.000   4.981   7.000   7.000       3 
hist(df_imp$mopping_anticeptic_now,main="mopping_days_perweek_now")
# non-normal
# summary(df_imp$mopping_anticeptic_now)
# Min. 1st Qu.  Median    Mean 3rd Qu.    Max.    NA's 
# 0.000   2.000   4.000   4.393   7.000   7.000       7
hist(df_imp$mopping_anticeptic_before,main="mopping_days_perweek_before")
# non-normal
# summary(df_imp$mopping_anticeptic_before)
# Min. 1st Qu.  Median    Mean 3rd Qu.    Max.    NA's 
# 0.000   2.000   3.000   3.617   5.000   7.000       2
hist(df_imp$other_cleaning_now,main="other_cleaning_days_perweek_now")
# non-normal
# summary(df_imp$other_cleaning_now)
# Min. 1st Qu.  Median    Mean 3rd Qu.    Max.    NA's 
# 0.000   2.000   4.000   4.142   7.000   7.000       2
hist(df_imp$other_cleaning_before,main="other_cleaning_days_perweek_before")
# non-normal
# summary(df_imp$other_cleaning_before)
# Min. 1st Qu.  Median    Mean 3rd Qu.    Max.    NA's 
# 0.00    1.00    3.00    3.41    6.00    7.00       1
hist(df_imp$hours_friends_now,main="friends_communic_hours_perday_now")
# non-normal
# summary(df_imp$hours_friends_now)
# Min. 1st Qu.  Median    Mean 3rd Qu.    Max.    NA's 
# 0.000   1.000   1.000   1.895   2.000  18.000     827
hist(df_imp$home_hours_wd,main="stayinghome_weekdays_now")
# non-normal
# summary(df_imp$home_hours_wd)
# Min. 1st Qu.  Median    Mean 3rd Qu.    Max.    NA's 
# 0.00   10.00   15.00   14.86   20.00   24.00      11
hist(df_imp$home_hours_wd_before,main="stayinghome_weekdays_before")
# non-normal
# summary(df_imp$home_hours_wd_before)
# Min. 1st Qu.  Median    Mean 3rd Qu.    Max.    NA's 
# 0.00    7.00   12.00   11.09   15.00   24.00      11
hist(df_imp$meat_per_day_now,main="meat_portions_per_day_now")
# non-normal
# summary(df_imp$meat_per_day_now)
# Min. 1st Qu.  Median    Mean 3rd Qu.    Max.    NA's 
# 0.000   1.000   1.000   1.599   2.000  20.000      25 
hist(df_imp$meat_per_day_before,main="meat_portions_per_day_before")
# non-normal
# summary(df_imp$meat_per_day_before)
# Min. 1st Qu.  Median    Mean 3rd Qu.    Max.    NA's 
# 0.000   1.000   1.000   1.616   2.000  20.000      17
hist(df_imp$leggumes_per_day_before,main="leggumes_portions_per_day_before")
# non-normal
# summary(df_imp$leggumes_per_day_before)
# Min. 1st Qu.  Median    Mean 3rd Qu.    Max.    NA's 
# 0.000   1.000   1.000   1.305   1.000  20.000      90
hist(df_imp$leggumes_per_day_now,main="leggumes_per_day_now")
# non-normal
# summary(df_imp$leggumes_per_day_now)
# Min. 1st Qu.  Median    Mean 3rd Qu.    Max.    NA's 
# 0.000   1.000   1.000   1.287   1.000  20.000      94
hist(df_imp$leggumes_per_day_before,main="leggumes_per_day_before")
# non-normal
# summary(df_imp$leggumes_per_day_before)
# Min. 1st Qu.  Median    Mean 3rd Qu.    Max.    NA's 
# 0.000   1.000   1.000   1.305   1.000  20.000      90
hist(df_imp$fish_per_day,main="fish_per_day_now")
# non-normal
# summary(df_imp$fish_per_day)
# Min. 1st Qu.  Median    Mean 3rd Qu.    Max.    NA's 
# 0.000   1.000   1.000   1.157   1.000  20.000     141
hist(df_imp$fish_per_day_before,main="fish_per_day_before")
# non-normal
# summary(df_imp$fish_per_day_before)
# Min. 1st Qu.  Median    Mean 3rd Qu.    Max.    NA's 
# 0.000   1.000   1.000   1.192   1.000  20.000     138
hist(df_imp$sugar_per_day_now,main="sugar_per_day_now")
# non-normal
# summary(df_imp$sugar_per_day_now)
# Min. 1st Qu.  Median    Mean 3rd Qu.    Max.    NA's 
# 0.000   1.000   1.000   1.886   2.000  20.000      16
hist(df_imp$sugar_per_day_before,main="sugar_per_day_before")
# non-normal
# summary(df_imp$sugar_per_day_before)
# Min. 1st Qu.  Median    Mean 3rd Qu.    Max.    NA's 
# 0.000   1.000   1.000   1.689   2.000  20.000      12
hist(df_imp$chips_per_day_now,main="chips_per_day_now")
# non-normal
# summary(df_imp$chips_per_day_now)
# Min. 1st Qu.  Median    Mean 3rd Qu.    Max.    NA's 
# 0.000   1.000   1.000   1.197   1.000  20.000   67
hist(df_imp$veggies_per_day_before,main="veggies_per_day_before")
# non-normal
# summary(df_imp$veggies_per_day_before)
# Min. 1st Qu.  Median    Mean 3rd Qu.    Max.    NA's 
# 0.000   1.000   1.000   1.661   2.000  15.000     106  
hist(df_imp$num_child,main="number of primary school children in family")
# non-normal
# summary(df_imp$num_child)
# Min. 1st Qu.  Median    Mean 3rd Qu.    Max.    NA's 
# 1.000   1.000   1.000   1.368   2.000   5.000      13 
hist(df_imp$years_living_in_cyprus,main="years living in Cyprus")
# non-normal
# summary(df_imp$years_living_in_cyprus)
# Min. 1st Qu.  Median    Mean 3rd Qu.    Max.    NA's 
# 0.000   8.000   9.000   9.112  11.000  14.000       5
hist(df_imp$heightcm,main="height")
# non-normal
# summary(df_imp$heightcm)
# Min. 1st Qu.  Median    Mean 3rd Qu.    Max.    NA's 
# 95.0   130.0   140.0   138.9   150.0   200.0     84
hist(df_imp$weight,main="weight")
# non-normal
# summary(df_imp$weight)
# Min. 1st Qu.  Median    Mean 3rd Qu.    Max.    NA's 
# 16.00   27.00   33.00   35.48   40.00  131.00      64

# calculate BMI based on formula: weight/height^2
df_imp$BMI<-df_imp$weight/((df_imp$heightcm/100)^2)
# summary(df_imp$BMI)
# Min. 1st Qu.  Median    Mean 3rd Qu.    Max.    NA's 
# 8.958  15.556  17.585  18.172  20.000  77.515      96
# qqnorm(df_imp$BMI)

# replace extreme minimum and maximum values for BMI (<10 & > 40)
df_imp <- df_imp %>% 
  replace_with_na_at("BMI",
                     condition = ~.x < 10) %>%
  replace_with_na_at("BMI",
                     condition = ~.x > 40)

# summary(df_imp$BMI)
# Min. 1st Qu.  Median    Mean 3rd Qu.    Max.    NA's 
# 10.24   15.58   17.59   18.13   20.00   37.26     101
qqnorm(df_imp$BMI,main="BMI")

# calculate BMI SDS based on WHO 2007 growth references
df_imp <- df_imp %>% 
  mutate(BMI_sds = sds(BMI,
                       age = age,
                       sex = sex, 
                       male = "Male", female = "Female",
                       ref = who2007.ref,
                       item = "bmi",
                       type = "SDS")) %>%
# create categories based on WHO cut-off points for 
# thinness, normal, overweight and obese.
# <-2: Thinness, 1>-2: Normal, >1: Overweight, >2: Obese  
  mutate(BMI_for_age = cut(BMI_sds,
                       breaks=c(-Inf, -2, 1, 2, Inf),
                       labels=c("Thinness","Normal","Overweight","Obese"))) %>%
# create variables for all cleaning activities by summarizing times per week for all cleaning activities - before and now
  rowwise() %>%
  mutate(all_cleaning_now=sum(bath_clean_anticept_now,
                            kitchen_anticeptic_now,
                            mopping_anticeptic_now,
                            other_cleaning_now, na.rm=TRUE)) %>%
  mutate(all_cleaning_before=sum(bath_clean_anticept_before,
                               kitchen_anticeptic_before,
                               mopping_anticeptic_before,
                               other_cleaning_before, na.rm=TRUE))

# check distributions of computed variables
qqnorm(df_imp$BMI_sds,main="BMI_sds")

# summary(df_imp$BMI_for_age)

qqnorm(df_imp$all_cleaning_now,main="all_cleaning_now")
# summary(df_imp$all_cleaning_now)
qqnorm(df_imp$all_cleaning_before,main="all_cleaning_before")
# summary(df_imp$all_cleaning_before)

# run codebook to get an overview of the variables (addin)

# save dataframe
saveRDS(df_imp, file="produced_data/df_imp.rds")

```


# Session information

```{r session, echo=FALSE}
sessionInfo()
```
````
