## Supplementary material for "Exposome changes in primary school children following the wide population non-pharmacological interventions implemented due to COVID-19 in Cyprus: a national survey": Deidentified data and script: ms_analysis_expo_school_201016.html

Exposome@school


### Exposome@school

The script was prepared for the statistical analysis of the data collected from the online survey Exposome@school (01/06/20 - 17/07/20).

### Table 1 - Demographic & other characteristics of participants

Demographic & other characteristics of participants

|  | Overall |
| --- | --- |
| n | 1509 |
| sex = Female (%) | 709 (47.6) |
| age (mean (SD)) | 9.60 (1.74) |
| age\_groups = 10-14 (%) | 784 (52.0) |
| age\_groups\_1 (%) |  |
| 5-6 | 4 ( 0.3) |
| 7-8 | 476 (31.5) |
| 9-10 | 481 (31.9) |
| 11-12 | 531 (35.2) |
| 13-14 | 17 ( 1.1) |
| age\_groups\_2 = 10-12 (%) | 767 (51.5) |
| BMI\_for\_age (%) |  |
| Thinness | 77 ( 5.5) |
| Normal | 789 (56.8) |
| Overweight | 320 (23.1) |
| Obese | 202 (14.6) |
| place\_of\_birth (%) |  |
| Cyprus | 1448 (96.0) |
| EU country | 49 ( 3.2) |
| Non-EU country | 12 ( 0.8) |
| years\_living\_in\_cyprus (mean (SD)) | 9.11 (2.09) |
| city (%) |  |
| Limassol | 375 (24.9) |
| Paphos | 97 ( 6.4) |
| Larnaca | 274 (18.2) |
| Nicosia | 613 (40.6) |
| Famagusta | 150 ( 9.9) |
| chronic\_disease = No (%) | 1367 (91.5) |
| vaccine = No (%) | 16 ( 1.1) |
| vaccine\_doses (%) |  |
| Yes | 1304 (92.3) |
| No, vaccine not available | 15 ( 1.1) |
| No, we thought that more than one dose wasn't needed | 13 ( 0.9) |
| No, we neglected but they will be provided | 73 ( 5.2) |
| No, only one dose was needed | 8 ( 0.6) |
| no\_vaccination = No (%) | 1302 (93.2) |
| attend\_school = Stays at home (%) | 179 (11.9) |
| consent = No (%) | 1119 (84.1) |
| test\_covid = Positive (%) | 1 ( 0.7) |
| mother\_education (%) |  |
| Has not completed primary school | 70 ( 4.6) |
| Primary School | 68 ( 4.5) |
| Middle School (3 years) | 21 ( 1.4) |
| High School/Vocational High School (diploma) | 207 (13.7) |
| Higher (after high school) non-tertiary Education | 83 ( 5.5) |
| Higher Tertiary Education (non-Univesity) | 195 (12.9) |
| University (Bachelor degree) | 445 (29.5) |
| University-Postgraduate (only Master's degree) | 385 (25.6) |
| PhD | 32 ( 2.1) |
| father\_education (%) |  |
| Has not completed primary school | 13 ( 0.9) |
| Primary School | 35 ( 2.3) |
| Middle School (3 years) | 78 ( 5.2) |
| High School/Vocational High School (diploma) | 480 (31.9) |
| Higher (after high school) non-tertiary Education | 89 ( 5.9) |
| Higher Tertiary Education (non-Univesity) | 146 ( 9.7) |
| University (Bachelor degree) | 320 (21.3) |
| University-Postgraduate (only Master's degree) | 300 (19.9) |
| PhD | 44 ( 2.9) |
| num\_child (mean (SD)) | 1.37 (0.55) |
| sibling (%) |  |
| 1 | 1422 (94.2) |
| 2 | 84 ( 5.6) |
| 3 | 3 ( 0.2) |

BMI by sex

|  | Male | Female | p | test |
| --- | --- | --- | --- | --- |
| n | 779 | 709 |  |  |
| BMI\_for\_age (%) |  |  | <0.001 |  |
| Thinness | 37 ( 5.1) | 40 ( 6.0) |  |  |
| Normal | 387 (53.6) | 402 (60.4) |  |  |
| Overweight | 165 (22.9) | 155 (23.3) |  |  |
| Obese | 133 (18.4) | 69 (10.4) |  |  |

BMI by sex for 9-year old children

|  | Male | Female | p | test |
| --- | --- | --- | --- | --- |
| n | 131 | 109 |  |  |
| BMI\_for\_age (%) |  |  | 0.889 |  |
| Thinness | 7 ( 5.9) | 5 ( 4.8) |  |  |
| Normal | 67 (56.8) | 65 (61.9) |  |  |
| Overweight | 24 (20.3) | 19 (18.1) |  |  |
| Obese | 20 (16.9) | 16 (15.2) |  |  |

Chronic diseases

|  | Overall |
| --- | --- |
| n | 1509 |
| chrdisease (%) |  |
| atopic\_dermatitis | 1 ( 0.8) |
| allergies | 35 (28.7) |
| asthma | 39 (32.0) |
| asthma\_allergies | 5 ( 4.1) |
| emphysema | 4 ( 3.3) |
| long\_qt | 1 ( 0.8) |
| frequent\_infections | 1 ( 0.8) |
| hemoglobinopathy | 5 ( 4.1) |
| allergies\_bronchitis | 2 ( 1.6) |
| bronchitis | 9 ( 7.4) |
| epilepsy | 2 ( 1.6) |
| diabetes | 5 ( 4.1) |
| ulcerative\_colitis | 1 ( 0.8) |
| g6pd\_lack | 1 ( 0.8) |
| heart\_disease | 1 ( 0.8) |
| celiac\_disease | 1 ( 0.8) |
| mediterannean\_fever | 1 ( 0.8) |
| mitral\_valve\_prolapse | 1 ( 0.8) |
| right\_lung\_rupture | 1 ( 0.8) |
| stigma | 1 ( 0.8) |
| congenital\_heart\_disease | 1 ( 0.8) |
| congenital\_heart\_disease\_down\_syndrome\_dandy\_walker\_syndrome\_poor\_hearing\_gluten\_sensitivity | 1 ( 0.8) |
| olier\_syndrome | 1 ( 0.8) |
| hypothyroidism | 1 ( 0.8) |
| bronchitis\_asthma | 1 ( 0.8) |

- No of children who come in contact with at least 1 person belonging in vulnerable groups at home:
  - in weekdays during the study period: (false, true, NA) logical, 562, 947
  - in weekdays before the pandemic: logical, 354, 1154, 1
  - in weekends during the study period: logical, 552, 957
  - in weekends before the pandemic: logical, 334, 1173, 2

Compared to the population census data, 2011 (https://www.mof.gov.cy/mof/cystat/statistics.nsf/populationcondition\_22main\_en/populationcondition\_22main\_en?OpenForm&sub=2&sel=2), our survey participants

- higher level of parents education (57% & 44% vs 20% with at least uni degree)
- similar town distribution : Nicosia (41% vs 39%), Limassol (25% vs 28%), Larnaka (18% vs 17%), Paphos (6% vs 11%) and Ammochostos (10% vs 6%)
- Similar Overweight and obesity prevalence in Cyprus (9-year olds) based on WHO definition (%) – COSI 2015-2017 (https://www.euro.who.int/\_\_data/assets/pdf\_file/0006/372426/WH14\_COSI\_factsheets\_v2.pdf?ua=1)
  - boys: 21% obese 43% overweight (including obese)
  - girls: 19% obese, 43% overweight (including obese)

### Descriptives

#### Diet

N [%] consumption frequency for fruits before and during the measures

|  | Post-lockdown | Pre-lockdown | p | test |
| --- | --- | --- | --- | --- |
| n | 1509 | 1509 |  |  |
| value (%) |  |  | 0.758 |  |
| Every day | 814 (54.1) | 819 (54.3) |  |  |
| 4-6 times per week | 255 (17.0) | 246 (16.3) |  |  |
| 2-3 times per week | 272 (18.1) | 293 (19.4) |  |  |
| 1 time per week | 74 ( 4.9) | 73 ( 4.8) |  |  |
| 2-3 times per month | 23 ( 1.5) | 18 ( 1.2) |  |  |
| 1 time per month | 17 ( 1.1) | 10 ( 0.7) |  |  |
| Never | 49 ( 3.3) | 48 ( 3.2) |  |  |

N [%] consumption frequency for vegetables before and during the measures

|  | Post-lockdown | Pre-lockdown | p | test |
| --- | --- | --- | --- | --- |
| n | 1509 | 1509 |  |  |
| value (%) |  |  | 0.144 |  |
| Every day | 462 (30.7) | 504 (33.5) |  |  |
| 4-6 times per week | 296 (19.7) | 243 (16.1) |  |  |
| 2-3 times per week | 428 (28.4) | 459 (30.5) |  |  |
| 1 time per week | 159 (10.6) | 142 ( 9.4) |  |  |
| 2-3 times per month | 42 ( 2.8) | 40 ( 2.7) |  |  |
| 1 time per month | 22 ( 1.5) | 18 ( 1.2) |  |  |
| Never | 97 ( 6.4) | 100 ( 6.6) |  |  |

N [%] consumption frequency for meat before and during the measures

|  | Post-lockdown | Pre-lockdown | p | test |
| --- | --- | --- | --- | --- |
| n | 1509 | 1509 |  |  |
| value (%) |  |  | 0.500 |  |
| Every day | 79 ( 5.2) | 96 ( 6.4) |  |  |
| 4-6 times per week | 531 (35.2) | 506 (33.6) |  |  |
| 2-3 times per week | 789 (52.4) | 788 (52.3) |  |  |
| 1 time per week | 77 ( 5.1) | 83 ( 5.5) |  |  |
| 2-3 times per month | 17 ( 1.1) | 25 ( 1.7) |  |  |
| 1 time per month | 5 ( 0.3) | 2 ( 0.1) |  |  |
| Never | 9 ( 0.6) | 8 ( 0.5) |  |  |

N [%] consumption frequency for legumes before and during the measures

|  | Post-lockdown | Pre-lockdown | p | test |
| --- | --- | --- | --- | --- |
| n | 1509 | 1509 |  |  |
| value (%) |  |  | 0.828 |  |
| Every day | 8 ( 0.5) | 7 ( 0.5) |  |  |
| 4-6 times per week | 33 ( 2.2) | 31 ( 2.1) |  |  |
| 2-3 times per week | 737 (48.9) | 781 (51.9) |  |  |
| 1 time per week | 541 (35.9) | 503 (33.4) |  |  |
| 2-3 times per month | 67 ( 4.4) | 66 ( 4.4) |  |  |
| 1 time per month | 41 ( 2.7) | 39 ( 2.6) |  |  |
| Never | 79 ( 5.2) | 79 ( 5.2) |  |  |

N [%] consumption frequency for fish before and during the measures

|  | Post-lockdown | Pre-lockdown | p | test |
| --- | --- | --- | --- | --- |
| n | 1509 | 1509 |  |  |
| value (%) |  |  | 0.721 |  |
| Every day | 0 ( 0.0) | 2 ( 0.1) |  |  |
| 4-6 times per week | 1 ( 0.1) | 1 ( 0.1) |  |  |
| 2-3 times per week | 200 (13.3) | 223 (14.9) |  |  |
| 1 time per week | 694 (46.2) | 687 (45.8) |  |  |
| 2-3 times per month | 259 (17.2) | 252 (16.8) |  |  |
| 1 time per month | 224 (14.9) | 213 (14.2) |  |  |
| Never | 125 ( 8.3) | 123 ( 8.2) |  |  |

N [%] consumption frequency for sugar before and during the measures

|  | Post-lockdown | Pre-lockdown | p | test |
| --- | --- | --- | --- | --- |
| n | 1509 | 1509 |  |  |
| value (%) |  |  | <0.001 |  |
| Every day | 550 (36.5) | 490 (32.6) |  |  |
| 4-6 times per week | 394 (26.2) | 289 (19.2) |  |  |
| 2-3 times per week | 412 (27.4) | 504 (33.5) |  |  |
| 1 time per week | 105 ( 7.0) | 163 (10.8) |  |  |
| 2-3 times per month | 29 ( 1.9) | 39 ( 2.6) |  |  |
| 1 time per month | 8 ( 0.5) | 14 ( 0.9) |  |  |
| Never | 7 ( 0.5) | 6 ( 0.4) |  |  |

N [%] consumption frequency for snacks before and during the measures

|  | Post-lockdown | Pre-lockdown | p | test |
| --- | --- | --- | --- | --- |
| n | 1509 | 1509 |  |  |
| value (%) |  |  | 0.338 |  |
| Every day | 119 ( 7.9) | 133 ( 8.8) |  |  |
| 4-6 times per week | 155 (10.3) | 144 ( 9.6) |  |  |
| 2-3 times per week | 502 (33.3) | 553 (36.7) |  |  |
| 1 time per week | 408 (27.1) | 378 (25.1) |  |  |
| 2-3 times per month | 150 (10.0) | 150 (10.0) |  |  |
| 1 time per month | 109 ( 7.2) | 90 ( 6.0) |  |  |
| Never | 63 ( 4.2) | 58 ( 3.9) |  |  |

N [%] consumption frequency for not homemade food before and during the measures

|  | Post-lockdown | Pre-lockdown | p | test |
| --- | --- | --- | --- | --- |
| n | 1509 | 1509 |  |  |
| value (%) |  |  | <0.001 |  |
| Every day | 4 ( 0.3) | 9 ( 0.6) |  |  |
| 4-6 times per week | 3 ( 0.2) | 19 ( 1.3) |  |  |
| 2-3 times per week | 100 ( 6.6) | 251 (16.7) |  |  |
| 1 time per week | 665 (44.2) | 698 (46.4) |  |  |
| 2-3 times per month | 439 (29.2) | 441 (29.3) |  |  |
| Never | 295 (19.6) | 86 ( 5.7) |  |  |

N [%] consumption frequency for breakfast before and during the measures

|  | Post-lockdown | Pre-lockdown | p | test |
| --- | --- | --- | --- | --- |
| n | 1509 | 1509 |  |  |
| value (%) |  |  | 0.002 |  |
| Every day | 1200 (79.5) | 1147 (76.1) |  |  |
| 4-6 times per week | 152 (10.1) | 137 ( 9.1) |  |  |
| 2-3 times per week | 107 ( 7.1) | 152 (10.1) |  |  |
| 1 time per week | 20 ( 1.3) | 39 ( 2.6) |  |  |
| 2-3 times per month | 9 ( 0.6) | 4 ( 0.3) |  |  |
| Never | 21 ( 1.4) | 28 ( 1.9) |  |  |

N [%] consumption frequency for supplements before and during the measures

|  | Post-lockdown | Pre-lockdown | p | test |
| --- | --- | --- | --- | --- |
| n | 1509 | 1509 |  |  |
| value = No (%) | 1238 (82.3) | 1203 (80.1) | 0.141 |  |

#### Physical activity

```
## Warning: Removed 74 rows containing non-finite values (stat_ydensity).
```

Spare activity score of participants - median [IQR]

|  | Post\_lockdown | Pre\_lockdown | p | test |
| --- | --- | --- | --- | --- |
| n | 1509 | 1509 |  |  |
| value (median [IQR]) | 1.38 [1.25, 1.62] | 1.50 [1.38, 1.75] | <0.001 | nonnorm |

Walking N [%] frequency before and during the measures

|  | Post-lockdown | Pre-lockdown | p | test |
| --- | --- | --- | --- | --- |
| n | 1509 | 1509 |  |  |
| value (%) |  |  | <0.001 |  |
| No | 726 (48.3) | 808 (53.8) |  |  |
| 1-2 times per week | 497 (33.0) | 374 (24.9) |  |  |
| 3-4 times per week | 195 (13.0) | 211 (14.0) |  |  |
| 5-6 times per week | 60 ( 4.0) | 81 ( 5.4) |  |  |
| 7 times or more per week | 26 ( 1.7) | 29 ( 1.9) |  |  |

Cycling N [%] frequency before and during the measures

|  | Post-lockdown | Pre-lockdown | p | test |
| --- | --- | --- | --- | --- |
| n | 1509 | 1509 |  |  |
| value (%) |  |  | 0.059 |  |
| No | 497 (32.9) | 496 (32.9) |  |  |
| 1-2 times per week | 520 (34.5) | 547 (36.3) |  |  |
| 3-4 times per week | 273 (18.1) | 287 (19.1) |  |  |
| 5-6 times per week | 127 ( 8.4) | 118 ( 7.8) |  |  |
| 7 times or more per week | 92 ( 6.1) | 58 ( 3.9) |  |  |

Running N [%] frequency before and during the measures

|  | Post-lockdown | Pre-lockdown | p | test |
| --- | --- | --- | --- | --- |
| n | 1509 | 1509 |  |  |
| value (%) |  |  | 0.597 |  |
| No | 1010 (67.2) | 1015 (67.6) |  |  |
| 1-2 times per week | 327 (21.8) | 302 (20.1) |  |  |
| 3-4 times per week | 110 ( 7.3) | 129 ( 8.6) |  |  |
| 5-6 times per week | 40 ( 2.7) | 43 ( 2.9) |  |  |
| 7 times or more per week | 15 ( 1.0) | 13 ( 0.9) |  |  |

Ballet N [%] frequency before and during the measures

|  | Post-lockdown | Pre-lockdown | p | test |
| --- | --- | --- | --- | --- |
| n | 1509 | 1509 |  |  |
| value (%) |  |  | <0.001 |  |
| No | 1206 (80.0) | 912 (60.6) |  |  |
| 1-2 times per week | 230 (15.3) | 445 (29.5) |  |  |
| 3-4 times per week | 50 ( 3.3) | 119 ( 7.9) |  |  |
| 5-6 times per week | 17 ( 1.1) | 24 ( 1.6) |  |  |
| 7 times or more per week | 5 ( 0.3) | 6 ( 0.4) |  |  |

Football N [%] frequency before and during the measures

|  | Post-lockdown | Pre-lockdown | p | test |
| --- | --- | --- | --- | --- |
| n | 1509 | 1509 |  |  |
| value (%) |  |  | <0.001 |  |
| No | 1063 (70.5) | 933 (61.9) |  |  |
| 1-2 times per week | 263 (17.5) | 268 (17.8) |  |  |
| 3-4 times per week | 140 ( 9.3) | 229 (15.2) |  |  |
| 5-6 times per week | 29 ( 1.9) | 69 ( 4.6) |  |  |
| 7 times or more per week | 12 ( 0.8) | 8 ( 0.5) |  |  |

Karate N [%] frequency before and during the measures

|  | Post-lockdown | Pre-lockdown | p | test |
| --- | --- | --- | --- | --- |
| n | 1509 | 1509 |  |  |
| value (%) |  |  | <0.001 |  |
| No | 1364 (90.6) | 1221 (81.1) |  |  |
| 1-2 times per week | 105 ( 7.0) | 192 (12.7) |  |  |
| 3-4 times per week | 32 ( 2.1) | 81 ( 5.4) |  |  |
| 5-6 times per week | 3 ( 0.2) | 10 ( 0.7) |  |  |
| 7 times or more per week | 2 ( 0.1) | 2 ( 0.1) |  |  |

Swimming N [%] frequency before and during the measures

|  | Post-lockdown | Pre-lockdown | p | test |
| --- | --- | --- | --- | --- |
| n | 1509 | 1509 |  |  |
| value (%) |  |  | <0.001 |  |
| No | 940 (62.3) | 1086 (72.3) |  |  |
| 1-2 times per week | 452 (30.0) | 330 (22.0) |  |  |
| 3-4 times per week | 77 ( 5.1) | 62 ( 4.1) |  |  |
| 5-6 times per week | 34 ( 2.3) | 17 ( 1.1) |  |  |
| 7 times or more per week | 6 ( 0.4) | 8 ( 0.5) |  |  |

Other activity N [%] frequency before and during the measures

|  | Post-lockdown | Pre-lockdown | p | test |
| --- | --- | --- | --- | --- |
| n | 1509 | 1509 |  |  |
| value (%) |  |  | <0.001 |  |
| No | 1248 (83.8) | 1089 (73.1) |  |  |
| 1-2 times per week | 168 (11.3) | 290 (19.5) |  |  |
| 3-4 times per week | 54 ( 3.6) | 78 ( 5.2) |  |  |
| 5-6 times per week | 11 ( 0.7) | 26 ( 1.7) |  |  |
| 7 times or more per week | 8 ( 0.5) | 6 ( 0.4) |  |  |

Free time N [%] frequency before and during the measures

|  | Post-lockdown | Pre-lockdown | p | test |
| --- | --- | --- | --- | --- |
| n | 1509 | 1509 |  |  |
| value (%) |  |  | <0.001 |  |
| She/he was doing things that involve little physical effort | 750 (50.7) | 415 (27.9) |  |  |
| 1-2 times per week she/he did physical things (e.g. played sports,running,swimming) | 328 (22.2) | 432 (29.0) |  |  |
| 3-4 times per week she/he did physical things (e.g. played sports,running,swimming) | 268 (18.1) | 433 (29.1) |  |  |
| 5-6 times per week she/he did physical things (e.g. played sports,running,swimming) | 105 ( 7.1) | 174 (11.7) |  |  |
| 7 times or more per week she/he did physical things (e.g. played sports,running,swimming) | 28 ( 1.9) | 34 ( 2.3) |  |  |

School break time N [%] frequency before and during the measures

|  | Post-lockdown | Pre-lockdown | p | test |
| --- | --- | --- | --- | --- |
| n | 1509 | 1509 |  |  |
| value (%) |  |  | <0.001 |  |
| Sat down (talking, reading, eating) | 600 (46.0) | 133 ( 9.0) |  |  |
| Stood around or walked around | 217 (16.7) | 101 ( 6.8) |  |  |
| Ran or played a little bit | 355 (27.2) | 606 (41.0) |  |  |
| Ran around and played quite a bit | 92 ( 7.1) | 404 (27.3) |  |  |
| Ran and played hard most of the time | 39 ( 3.0) | 235 (15.9) |  |  |

Sickness N [%] frequency before and during the measures

|  | Post-lockdown | Pre-lockdown | p | test |
| --- | --- | --- | --- | --- |
| n | 1509 | 1509 |  |  |
| value = No (%) | 1483 (98.5) | 1474 (98.3) | 0.654 |  |

Change in spare activity score - median [IQR]

spareactivity

0 [-15.4, 8.3]

#### Parents smoking in the house

Smoking n [%] frequency before and during the measures

|  | Post-lockdown | Pre-lockdown | p | test |
| --- | --- | --- | --- | --- |
| n | 1509 | 1509 |  |  |
| value (%) |  |  | 0.969 |  |
| Yes, every day | 386 (25.7) | 381 (25.4) |  |  |
| Yes, occasionally | 63 ( 4.2) | 62 ( 4.1) |  |  |
| No | 1051 (70.1) | 1058 (70.5) |  |  |

Cigarettes no per day - median [IQR]

|  | Post-lockdown | Pre-lockdown | p | test |
| --- | --- | --- | --- | --- |
| n | 1509 | 1509 |  |  |
| value (median [IQR]) | 10.00 [5.00, 15.00] | 10.00 [5.00, 15.00] | 0.850 | nonnorm |

Cigarettes no per week - median [IQR]

|  | Post-lockdown | Pre-lockdown | p | test |
| --- | --- | --- | --- | --- |
| n | 1509 | 1509 |  |  |
| value (median [IQR]) | 5.00 [2.00, 10.00] | 6.00 [3.00, 10.00] | 0.287 | nonnorm |

Change in cig no per day/week - median [IQR] (both non-smokers and smokers)

| cigsperday | cigsperweek |
| --- | --- |
| 0 [0, 0] | 0 [0, 0] |

#### Communication and screen time

```
## Warning in rm(sum_cig_change, sum_cig_change_s, sel_smoking_db): object
## 'sum_cig_change_s' not found
```

Friends communication N [%] frequency before and during the measures

|  | Post-lockdown | Pre-lockdown | p | test |
| --- | --- | --- | --- | --- |
| n | 1509 | 1509 |  |  |
| value (%) |  |  | <0.001 |  |
| Every day | 684 (45.6) | 474 (31.6) |  |  |
| Every week but not every day | 421 (28.0) | 412 (27.5) |  |  |
| Some times in the month but not every week | 245 (16.3) | 304 (20.3) |  |  |
| Never | 151 (10.1) | 310 (20.7) |  |  |

Family communication N [%] frequency before and during the measures

|  | Post-lockdown | Pre-lockdown | p | test |
| --- | --- | --- | --- | --- |
| n | 1509 | 1509 |  |  |
| value (%) |  |  | <0.001 |  |
| Every day | 668 (44.5) | 506 (33.8) |  |  |
| Every week but not every day | 595 (39.7) | 506 (33.8) |  |  |
| Some times in the month but not every week | 165 (11.0) | 303 (20.2) |  |  |
| Never | 72 ( 4.8) | 182 (12.2) |  |  |

Screen time N [%] frequency before and during the measures

|  | Post-lockdown | Pre-lockdown | p | test |
| --- | --- | --- | --- | --- |
| n | 1509 | 1509 |  |  |
| value (%) |  |  | <0.001 |  |
| Less than 1 hour per day | 150 (10.0) | 412 (27.4) |  |  |
| 1-3 hours per day | 928 (61.7) | 934 (62.0) |  |  |
| 4-7 hours per day | 381 (25.3) | 148 ( 9.8) |  |  |
| 8-11 hours per day | 42 ( 2.8) | 12 ( 0.8) |  |  |
| More than 11 hours per day | 2 ( 0.1) | 0 ( 0.0) |  |  |

Friends communication hours per day - median [IQR]

|  | before | now | p | test |
| --- | --- | --- | --- | --- |
| n | 1509 | 1509 |  |  |
| value (median [IQR]) | 1.00 [1.00, 2.00] | 1.00 [1.00, 1.00] | 0.394 | nonnorm |

Family communication hours per day - median [IQR]

|  | before | now | p | test |
| --- | --- | --- | --- | --- |
| n | 1509 | 1509 |  |  |
| value (median [IQR]) | 1.00 [1.00, 2.00] | 1.00 [1.00, 2.00] | 0.024 | nonnorm |

#### Children personal hygiene

Antiseptic use for hands N [%] frequency before and during the measures

|  | Post-lockdown | Pre-lockdown | p | test |
| --- | --- | --- | --- | --- |
| n | 1509 | 1509 |  |  |
| value (%) |  |  | <0.001 |  |
| Less than 1 time per day | 155 (10.4) | 438 (29.4) |  |  |
| 1-3 times per day | 660 (44.3) | 289 (19.4) |  |  |
| 4-7 times per day | 443 (29.7) | 86 ( 5.8) |  |  |
| More than 7 times per day | 195 (13.1) | 48 ( 3.2) |  |  |
| Never | 38 ( 2.5) | 627 (42.1) |  |  |

Hand washing with soap N [%] frequency before and during the measures

|  | Post-lockdown | Pre-lockdown | p | test |
| --- | --- | --- | --- | --- |
| n | 1509 | 1509 |  |  |
| value (%) |  |  | <0.001 |  |
| Less than 1 time per day | 10 ( 0.7) | 129 ( 8.6) |  |  |
| 1-3 times per day | 327 (21.9) | 777 (51.8) |  |  |
| 4-7 times per day | 768 (51.3) | 443 (29.6) |  |  |
| More than 7 times per day | 391 (26.1) | 135 ( 9.0) |  |  |
| Never | 0 ( 0.0) | 15 ( 1.0) |  |  |

#### Home cleaning activities

```
## Warning: Removed 34 rows containing non-finite values (stat_ydensity).
```

Kitchen cleaning N [%] frequency before and during the measures

|  | Post-lockdown | Pre-lockdown | p | test |
| --- | --- | --- | --- | --- |
| n | 1509 | 1509 |  |  |
| value (%) |  |  | <0.001 |  |
| 0 | 16 ( 1.1) | 38 ( 2.5) |  |  |
| 1 | 62 ( 4.1) | 175 (11.6) |  |  |
| 2 | 69 ( 4.6) | 135 ( 9.0) |  |  |
| 3 | 89 ( 5.9) | 117 ( 7.8) |  |  |
| 4 | 44 ( 2.9) | 84 ( 5.6) |  |  |
| 5 | 93 ( 6.2) | 135 ( 9.0) |  |  |
| 6 | 41 ( 2.7) | 57 ( 3.8) |  |  |
| 7 | 1085 (72.4) | 765 (50.8) |  |  |

Bathroom cleaning N [%] frequency before and during the measures

|  | Post-lockdown | Pre-lockdown | p | test |
| --- | --- | --- | --- | --- |
| n | 1509 | 1509 |  |  |
| value (%) |  |  | <0.001 |  |
| 0 | 5 ( 0.3) | 18 ( 1.2) |  |  |
| 1 | 130 ( 8.6) | 249 (16.5) |  |  |
| 2 | 204 (13.6) | 303 (20.1) |  |  |
| 3 | 196 (13.0) | 214 (14.2) |  |  |
| 4 | 109 ( 7.3) | 116 ( 7.7) |  |  |
| 5 | 136 ( 9.0) | 133 ( 8.8) |  |  |
| 6 | 70 ( 4.7) | 43 ( 2.9) |  |  |
| 7 | 653 (43.4) | 430 (28.6) |  |  |

Mopping N [%] frequency before and during the measures

|  | Post-lockdown | Pre-lockdown | p | test |
| --- | --- | --- | --- | --- |
| n | 1509 | 1509 |  |  |
| value (%) |  |  | <0.001 |  |
| 0 | 25 ( 1.7) | 41 ( 2.7) |  |  |
| 1 | 134 ( 8.9) | 251 (16.7) |  |  |
| 2 | 241 (16.0) | 327 (21.7) |  |  |
| 3 | 215 (14.3) | 227 (15.1) |  |  |
| 4 | 163 (10.9) | 128 ( 8.5) |  |  |
| 5 | 157 (10.5) | 158 (10.5) |  |  |
| 6 | 69 ( 4.6) | 61 ( 4.0) |  |  |
| 7 | 498 (33.2) | 314 (20.8) |  |  |

Other surfaces cleaning N [%] frequency before and during the measures

|  | Post-lockdown | Pre-lockdown | p | test |
| --- | --- | --- | --- | --- |
| n | 1509 | 1509 |  |  |
| value (%) |  |  | <0.001 |  |
| 0 | 59 ( 3.9) | 107 ( 7.1) |  |  |
| 1 | 215 (14.3) | 315 (20.9) |  |  |
| 2 | 208 (13.8) | 269 (17.8) |  |  |
| 3 | 204 (13.5) | 203 (13.5) |  |  |
| 4 | 131 ( 8.7) | 105 ( 7.0) |  |  |
| 5 | 151 (10.0) | 128 ( 8.5) |  |  |
| 6 | 51 ( 3.4) | 44 ( 2.9) |  |  |
| 7 | 488 (32.4) | 337 (22.3) |  |  |

Kitchen cleaning times per week - median [IQR]

|  | Post\_lockdown | Pre\_lockdown | p | test |
| --- | --- | --- | --- | --- |
| n | 1509 | 1509 |  |  |
| value (median [IQR]) | 7.00 [6.00, 7.00] | 7.00 [3.00, 7.00] | <0.001 | nonnorm |

Bathroom cleaning times per week - median [IQR]

|  | Post\_lockdown | Pre\_lockdown | p | test |
| --- | --- | --- | --- | --- |
| n | 1509 | 1509 |  |  |
| value (median [IQR]) | 5.00 [3.00, 7.00] | 3.00 [2.00, 7.00] | <0.001 | nonnorm |

Mopping times per week - median [IQR]

|  | Post\_lockdown | Pre\_lockdown | p | test |
| --- | --- | --- | --- | --- |
| n | 1509 | 1509 |  |  |
| value (median [IQR]) | 4.00 [2.00, 7.00] | 3.00 [2.00, 5.00] | <0.001 | nonnorm |

Other surfaces cleaning times per week - median [IQR]

|  | Post\_lockdown | Pre\_lockdown | p | test |
| --- | --- | --- | --- | --- |
| n | 1509 | 1509 |  |  |
| value (median [IQR]) | 4.00 [2.00, 7.00] | 3.00 [1.00, 6.00] | <0.001 | nonnorm |

All house cleaning times per week - median [IQR]

|  | Post\_lockdown | Pre\_lockdown | p | test |
| --- | --- | --- | --- | --- |
| n | 1509 | 1509 |  |  |
| value (median [IQR]) | 20.00 [14.00, 26.00] | 16.00 [10.00, 22.00] | <0.001 | nonnorm |

Change in cleaning activities times per week at home - median [IQR]

| bath | kitchen | mopping | other surfaces | total |
| --- | --- | --- | --- | --- |
| 0 [0, 66.7] | 0 [0, 50] | 0 [0, 50] | 0 [0, 72.9] | 15.8 [0, 50] |

#### Contacts at home, school and elsewhere and hours at home

```
## `summarise()` regrouping output by 'age', 'timeframe' (override with `.groups` argument)
```

```
## `summarise()` regrouping output by 'age', 'timeframe' (override with `.groups` argument)
```

Change in contacts number at home (including vulnerable people), school and elsewhere in weekdays - median [IQR]

| elsewhere | home | school | vuln\_home |
| --- | --- | --- | --- |
| -60 [-80.6, -29.7] | -30 [-66.7, 0] | -60 [-76, -46.7] | 0 [-60, 0] |

Change in contacts number at home (including vulnerable people) and elsewhere in weekends - median [IQR]

| elsewhere | home | vuln\_home |
| --- | --- | --- |
| -50 [-75, -20] | -33.3 [-60, 0] | 0 [-50, 0] |

Contacts at home on weekdays - median [IQR]

|  | Post\_lockdown | Pre\_lockdown | p | test |
| --- | --- | --- | --- | --- |
| n | 1509 | 1509 |  |  |
| value (median [IQR]) | 4.00 [3.00, 5.00] | 6.00 [4.00, 12.00] | <0.001 | nonnorm |

Contacts at home on weekends - median [IQR]

|  | Post\_lockdown | Pre\_lockdown | p | test |
| --- | --- | --- | --- | --- |
| n | 1509 | 1509 |  |  |
| value (median [IQR]) | 4.00 [3.00, 6.00] | 7.00 [4.00, 10.00] | <0.001 | nonnorm |

Vulnerable contacts at home on weekdays - median [IQR]

|  | Post\_lockdown | Pre\_lockdown | p | test |
| --- | --- | --- | --- | --- |
| n | 1509 | 1509 |  |  |
| value (median [IQR]) | 1.00 [0.00, 2.00] | 2.00 [1.00, 3.00] | <0.001 | nonnorm |

Vulnerable contacts at home on weekends - median [IQR]

|  | Post\_lockdown | Pre\_lockdown | p | test |
| --- | --- | --- | --- | --- |
| n | 1509 | 1509 |  |  |
| value (median [IQR]) | 1.00 [0.00, 2.00] | 2.00 [1.00, 3.00] | <0.001 | nonnorm |

Contacts at school - median [IQR]

|  | Post\_lockdown | Pre\_lockdown | p | test |
| --- | --- | --- | --- | --- |
| n | 1509 | 1509 |  |  |
| value (median [IQR]) | 10.00 [5.00, 12.00] | 24.00 [15.00, 30.00] | <0.001 | nonnorm |

Contacts elsewhere on weekdays - median [IQR]

|  | Post\_lockdown | Pre\_lockdown | p | test |
| --- | --- | --- | --- | --- |
| n | 1509 | 1509 |  |  |
| value (median [IQR]) | 4.00 [2.00, 7.00] | 10.00 [5.00, 20.00] | <0.001 | nonnorm |

Contacts elsewhere on weekends - median [IQR]

|  | Post\_lockdown | Pre\_lockdown | p | test |
| --- | --- | --- | --- | --- |
| n | 1509 | 1509 |  |  |
| value (median [IQR]) | 4.00 [2.00, 6.00] | 10.00 [5.00, 15.00] | <0.001 | nonnorm |

#### Hours at home

Change in time spent at home in weekends and weekdays - median [IQR]

| wd | we |
| --- | --- |
| 31.2 [0, 66.7] | 20 [0, 60] |

Hours at home on weekdays - median [IQR]

|  | Post\_lockdown | Pre\_lockdown | p | test |
| --- | --- | --- | --- | --- |
| n | 1509 | 1509 |  |  |
| value (median [IQR]) | 15.00 [10.00, 20.00] | 12.00 [7.00, 15.00] | <0.001 | nonnorm |

Hours at home on weekends - median [IQR]

|  | Post\_lockdown | Pre\_lockdown | p | test |
| --- | --- | --- | --- | --- |
| n | 1509 | 1509 |  |  |
| value (median [IQR]) | 20.00 [12.00, 24.00] | 15.00 [10.00, 19.00] | <0.001 | nonnorm |

### Correlations

##### Continuous and ordinal variables

```
## The following `from` values were not present in `x`: Every day
```

```
## The following `from` values were not present in `x`: More than 11 hours per day
```

```
## The following `from` values were not present in `x`: Never
```

```
## 
## Correlation method: 'spearman'
## Missing treated using: 'pairwise.complete.obs'
```

Correlations - > abs(value)>=(0.7)

Partial Correlations - > abs(value)>=(0.7) (adjusted for age, sex, mother's and father's educational level)

### Compliance binary (1=increase in hours at home for weekdays) - regression analysis

#### Multivariate - adj. age, sex and parents' educational level

```
## Warning: attributes are not identical across measure variables; they will be
## dropped
```

N % of compliance - increased number of hours at home

|  | Overall |
| --- | --- |
| n | 1509 |
| compliance = 1 (%) | 1096 (72.9) |

N % of compliance - increased number of hours at home

|  | Male | Female | p | test |
| --- | --- | --- | --- | --- |
| n | 779 | 709 |  |  |
| compliance = 1 (%) | 562 (72.4) | 515 (72.9) | 0.867 |  |

### Compliance binary (1=decrease in the number of vulnerable contacts at home during weekdays - regression analysis

#### Multivariate - adj. age, sex and parents' educational level

```
## Warning: attributes are not identical across measure variables; they will be
## dropped
```

N % of compliance - decreased number of vulnerables contacts at home - weekdays

|  | Overall |
| --- | --- |
| n | 1190 |
| compliance = 1 (%) | 573 (48.2) |

N % of compliance - decreased number of vulnerables contacts at home - weekdays

|  | Male | Female | p | test |
| --- | --- | --- | --- | --- |
| n | 637 | 534 |  |  |
| compliance = 1 (%) | 308 (48.4) | 257 (48.2) | 1.000 |  |

### Compliance (1=decrease in the number of contacts at school) - regression analysis

#### Multivariate - adj. age, sex and parents' educational level

N % of compliance - decreased number of contacts at school

|  | Overall |
| --- | --- |
| n | 1330 |
| compliance = 1 (%) | 1100 (85.1) |

N % of compliance - decreased number of contacts at school

|  | Male | Female | p | test |
| --- | --- | --- | --- | --- |
| n | 687 | 627 |  |  |
| compliance = 1 (%) | 569 (85.1) | 516 (84.9) | 0.989 |  |

### Compliance binary (1=increase in frequency of hand washing with soap or antiseptic use) - regression analysis

#### Multivariate - adj. age, sex and parents' educational level

```
## Warning: attributes are not identical across measure variables; they will be
## dropped
```

N % of compliance - increase in hand washing with soap or antiseptic use

|  | Overall |
| --- | --- |
| n | 1509 |
| compliance\_hand = 1 (%) | 1099 (74.4) |

N % of compliance - increase in hand washing with soap or antiseptic use

|  | Male | Female | p | test |
| --- | --- | --- | --- | --- |
| n | 779 | 709 |  |  |
| compliance\_hand = 1 (%) | 549 (71.5) | 532 (77.2) | 0.015 |  |

### FDR for all models

### Session information

```
## R version 4.0.1 (2020-06-06)
## Platform: i386-w64-mingw32/i386 (32-bit)
## Running under: Windows 7 (build 7600)
## 
## Matrix products: default
## 
## locale:
## [1] LC_COLLATE=Greek_Greece.1253  LC_CTYPE=Greek_Greece.1253   
## [3] LC_MONETARY=Greek_Greece.1253 LC_NUMERIC=C                 
## [5] LC_TIME=Greek_Greece.1253    
## 
## attached base packages:
## [1] stats     graphics  grDevices utils     datasets  methods   base     
## 
## other attached packages:
##  [1] RColorBrewer_1.1-2 stringi_1.4.6      wesanderson_0.3.6  rvg_0.2.5         
##  [5] officer_0.3.12     ggraph_2.0.3       igraph_1.2.5       corrr_0.4.2       
##  [9] broom_0.5.6        timelineR_1.0.0    corrplot_0.84      missMDA_1.17      
## [13] PCAmixdata_3.1     factoextra_1.0.7   FactoMineR_2.3     ggpubr_0.4.0      
## [17] glue_1.4.1         childsds_0.7.4     codebook_0.9.2     rmdformats_0.3.7  
## [21] naniar_0.5.1       chron_2.3-55       haven_2.3.1        inspectdf_0.0.8   
## [25] NADA_1.6-1.1       FSA_0.8.30         scales_1.1.1       reshape_0.8.8     
## [29] texreg_1.37.4      janitor_2.0.1      Rcpp_1.0.4.6       eeptools_1.2.4    
## [33] lubridate_1.7.9    forcats_0.5.0      stringr_1.4.0      dplyr_1.0.0       
## [37] purrr_0.3.4        tidyverse_1.3.0    sjPlot_2.8.4       psych_1.9.12.31   
## [41] readr_1.3.1        stargazer_5.2.2    compare_0.2-6      tableone_0.11.1   
## [45] Hmisc_4.4-0        ggplot2_3.3.1      Formula_1.2-3      survival_3.1-12   
## [49] lattice_0.20-41    tibble_3.0.1       knitr_1.29         tidyr_1.1.0       
## [53] data.table_1.12.8  dtplyr_1.0.1       plyr_1.8.6         readxl_1.3.1      
## 
## loaded via a namespace (and not attached):
##   [1] tidyselect_1.1.0     lme4_1.1-23          htmlwidgets_1.5.1   
##   [4] grid_4.0.1           maptools_1.0-1       munsell_0.5.0       
##   [7] codetools_0.2-16     effectsize_0.3.1     DT_0.14             
##  [10] statmod_1.4.34       withr_2.2.0          colorspace_1.4-1    
##  [13] highr_0.8            uuid_0.1-4           rstudioapi_0.11     
##  [16] leaps_3.1            ggsignif_0.6.0       vcd_1.4-7           
##  [19] labeling_0.3         emmeans_1.4.8        polyclip_1.10-0     
##  [22] mnormt_2.0.0         farver_2.0.3         coda_0.19-3         
##  [25] vctrs_0.3.1          generics_0.0.2       TH.data_1.0-10      
##  [28] lambda.r_1.2.4       xfun_0.15            R6_2.4.1            
##  [31] doParallel_1.0.15    graphlayouts_0.7.0   arm_1.11-1          
##  [34] assertthat_0.2.1     multcomp_1.4-13      nnet_7.3-14         
##  [37] gtable_0.3.0         tidygraph_1.2.0      sandwich_2.5-1      
##  [40] rlang_0.4.6          systemfonts_0.2.3    scatterplot3d_0.3-41
##  [43] splines_4.0.1        rstatix_0.6.0        acepack_1.4.1       
##  [46] checkmate_2.0.0      reshape2_1.4.4       yaml_2.2.1          
##  [49] abind_1.4-5          modelr_0.1.8         crosstalk_1.1.0.1   
##  [52] backports_1.1.7      tools_4.0.1          bookdown_0.20       
##  [55] ellipsis_0.3.1       base64enc_0.1-3      progress_1.2.2      
##  [58] prettyunits_1.1.1    rpart_4.1-15         viridis_0.5.1       
##  [61] zoo_1.8-8            ggrepel_0.8.2        cluster_2.1.0       
##  [64] fs_1.4.1             survey_4.0           magrittr_1.5        
##  [67] futile.options_1.0.1 openxlsx_4.1.5       lmtest_0.9-37       
##  [70] reprex_0.3.0         tmvnsim_1.0-2        mvtnorm_1.1-1       
##  [73] sjmisc_2.8.5         hms_0.5.3            evaluate_0.14       
##  [76] xtable_1.8-4         rio_0.5.16           sjstats_0.18.0      
##  [79] jpeg_0.1-8.1         gridExtra_2.3        ggeffects_0.15.0    
##  [82] compiler_4.0.1       mice_3.9.0           crayon_1.3.4        
##  [85] minqa_1.2.4          htmltools_0.5.0      visdat_0.5.3        
##  [88] DBI_1.1.0            tweenr_1.0.1         formatR_1.7         
##  [91] sjlabelled_1.1.6     dbplyr_1.4.4         MASS_7.3-51.6       
##  [94] boot_1.3-25          Matrix_1.2-18        car_3.0-8           
##  [97] cli_2.0.2            mitools_2.4          parallel_4.0.1      
## [100] insight_0.8.5        pkgconfig_2.0.3      flashClust_1.01-2   
## [103] foreign_0.8-80       sp_1.4-2             xml2_1.3.2          
## [106] foreach_1.5.0        estimability_1.3     rvest_0.3.5         
## [109] snakecase_0.11.0     digest_0.6.25        parameters_0.8.0    
## [112] rmarkdown_2.3        cellranger_1.1.0     htmlTable_2.0.1     
## [115] gdtools_0.2.2        curl_4.3             nloptr_1.2.2.1      
## [118] lifecycle_0.2.0      nlme_3.1-148         jsonlite_1.6.1      
## [121] carData_3.0-4        futile.logger_1.4.3  viridisLite_0.3.0   
## [124] fansi_0.4.1          labelled_2.5.0       pillar_1.4.4        
## [127] httr_1.4.1           bayestestR_0.7.0     zip_2.0.4           
## [130] png_0.1-7            iterators_1.0.12     class_7.3-17        
## [133] ggforce_0.3.2        performance_0.4.7    blob_1.2.1          
## [136] ggfittext_0.9.0      latticeExtra_0.6-29  e1071_1.7-3
```

×

- Table 1 - Demographic & other characteristics of participants
- Descriptives
  - Diet
  - Physical activity
  - Parents smoking in the house
  - Communication and screen time
  - Children personal hygiene
  - Home cleaning activities
  - Contacts at home, school and elsewhere and hours at home
  - Hours at home
- Correlations
- Compliance binary (1=increase in hours at home for weekdays) - regression analysis
  - Multivariate - adj. age, sex and parents' educational level
- Compliance binary (1=decrease in the number of vulnerable contacts at home during weekdays - regression analysis
  - Multivariate - adj. age, sex and parents' educational level
- Compliance (1=decrease in the number of contacts at school) - regression analysis
  - Multivariate - adj. age, sex and parents' educational level
- Compliance binary (1=increase in frequency of hand washing with soap or antiseptic use) - regression analysis
  - Multivariate - adj. age, sex and parents' educational level
- FDR for all models
- Session information
