## Supplementary material for "Exposome changes in primary school children following the wide population non-pharmacological interventions implemented due to COVID-19 in Cyprus: a national survey": Deidentified data and script: ms_analysis_expo_school_201016.Rmd

````
---
title: "Exposome@school"
output:
  rmdformats::html_clean
geometry: margin=1.5cm
editor_options: 
  chunk_output_type: console
---

The script was prepared for the statistical analysis of the data collected from the online survey Exposome@school (01/06/20 - 17/07/20).


```{r setup, include=FALSE}
knitr::opts_chunk$set(echo = TRUE)
```

```{r libraries, include=FALSE}

## Prepare workspace & install libraries
rm(list = ls(all = TRUE))

ipak <- function(pkg){
  new.pkg <- pkg[!(pkg %in% installed.packages()[, "Package"])]
  if (length(new.pkg)) 
    install.packages(new.pkg, dependencies = TRUE)
  sapply(pkg, require, character.only = TRUE)
}

# usage
packages <- c("readxl","plyr","dtplyr","data.table","tidyr","knitr",
              "tibble","Hmisc","tableone","compare","stargazer",
              "readr","psych","ggplot2","sjPlot","tidyverse","lubridate",
              "eeptools", "Rcpp", "janitor", "stringr","dplyr",
              "texreg","reshape","scales","FSA","NADA","inspectdf",
              "haven", "sjPlot", "forcats","chron","Hmisc","naniar",
              "rmdformats","codebook","childsds","glue","ggpubr","knitr",
              "FactoMineR", "factoextra","PCAmixdata","missMDA","corrplot",
              "timelineR","glue", "broom","corrr","igraph","ggraph",
              "officer","rvg","wesanderson","stringi")

ipak(packages)

```


```{r table_1, echo=FALSE}

df_imp<-readRDS("produced_data/df_imp.rds") %>% 
  as.data.frame() %>%
  # create age groups
mutate(age_groups = cut(age,
                       breaks=c(-Inf, 9, Inf),
                       labels=c("5-9","10-14"))) %>% 
mutate(age_groups_1 = cut(age,
                       breaks=c(-Inf, 6, 8, 10, 12, Inf),
                       labels=c("5-6","7-8","9-10","11-12","13-14"))) %>% 
mutate(age_groups_2 = cut(age,
                       breaks=c(-Inf, 5, 9, 12, Inf),
                       labels=c(NA,"6-9","10-12",NA))) 
# str(df_imp, list.len=ncol(df_imp))

# rename levels of variables so that they have latin characters and be grouped to the same names
# first transform to factors
df_imp <- df_imp %>%
  mutate(chrdisease=as.factor(disease))

# levels(df_imp$chrdisease) - 87 levels

levels(df_imp$chrdisease) <- 
  c(NA,"atopic_dermatitis","allergy",NA, #4
    NA,NA,"allergies from environmental polluted air quality","asthma", #8
    "asthma_allergies","asthma","emphysema","long_qt", #12        
    "allergy_dust","frequent_infections","bronchial_asthma","hemoglobinopathy",#16
    "hemoglobinopathy","allergy_infected_insects","allergies","allergies", #20
    "allergy","allergy","allergy","allergy_mites_olive_flowers", #24
 "allergy_pollen_plants","allergy_dust","allergy_dust","allergy_dust_flowers", #28
    "allergy_nuts","allergies","allergies","allergies",  #32
    "allergies","allergies","allergies_bronchitis","allergies_asthma", #36
    "allergies_bronchitis","allergic_asthma","allergic_asthma_dust","asthma", #40
    "asthma","asthma","asthma","asthma", #44
    "allergies_asthma","allergies_asthma","asthmatic_bronchitis","asthmatic_bronchitis", #48
    "asthma","asthma","asthma","epileptic_seizures", #52
    "bronchial","bronchial_asthma","bronchitis","bronchitis", #56
    "bronchial_asthma","diabetes","diabetes_type_i","diabetes_type_i", #60
    NA,"ulcerative_colitis","g6pd_lack","epilepsy", #64
    "heart_disease","celiac_disease","mediterannean_fever","bronchial_asthma", #68
    "asthma",NA,"asthma","asthma", #72
    "asthma","asthma","bronchial_asthma","mitral_valve_prolapse", #76
    "right_lung_rupture","stigma","congenital_heart_disease","congenital_heart_disease_down_syndrome_dandy_walker_syndrome_poor_hearing_gluten_sensitivity", #80
    "olier_syndrome","hypothyroidism","bronchitis","bronchitis", #84
    "bronchitis","bronchitis","bronchitis_asthma") #88

# chronicdiseases<-df_imp %>%
#  select(c("disease","chrdisease"))
# levels(df_imp$chrdisease) - 41 levels

# rename levels - group of diseases
df_imp <-df_imp %>%
 mutate_at(.vars = "chrdisease", 
            .funs = function(t) revalue(x = t,
            replace = c("allergy"="allergies",
                        "allergies from environmental polluted air quality"="allergies",
                        "allergy_dust"="allergies",
                        "bronchial_asthma"="asthma",
                        "allergy_infected_insects"="allergies",
                        "allergy_mites_olive_flowers"="allergies",
                        "allergy_pollen_plants"="allergies",
                        "allergy_dust_flowers"="allergies",
                        "allergy_nuts"="allergies",
                        "allergies_asthma"="asthma_allergies",
                        "allergic_asthma"="asthma",
                        "allergic_asthma_dust"="asthma",
                        "asthmatic_bronchitis"="bronchitis",
                        "epileptic_seizures"="epilepsy",
                        "bronchial"="bronchitis",
                        "diabetes_type_i"="diabetes"))) 

# levels(df_imp$chrdisease) - 25 levels

# exclude participant that has high school as school name 
df_imp<- df_imp %>%
   filter(ID != "1429" | is.na(ID)) %>%
  droplevels

# select variables for table 1
sel_demog <- 
  c("sex","age","age_groups","age_groups_1","age_groups_2",
    "BMI_for_age","place_of_birth",
    "years_living_in_cyprus","city",
    "chronic_disease","vaccine","vaccine_doses","no_vaccination",
    "attend_school","consent","test_covid",
    "mother_education","father_education","num_child","sibling")

categorvar_demog<-
  c("sex","age_groups","age_groups_1","age_groups_2",
    "chronic_disease","vaccine","vaccine_doses",
    "no_vaccination","attend_school","consent","test_covid","BMI_for_age",
    "mother_education","father_education","city","place_of_birth","sibling")

# only 9-year olds to compare with COSI
df_nine<-df_imp  %>%
  filter(age==9)
```

# Table 1 - Demographic & other characteristics of participants

`r kable(print(CreateTableOne(sel_demog, data=df_imp, factorVars=categorvar_demog)), caption="Demographic & other characteristics of participants") `

`r kable(print(CreateTableOne("BMI_for_age", data=df_imp, factorVars=categorvar_demog, strata="sex",testNonNormal = kruskal.test, smd=F), nonnormal="value"), caption="BMI by sex") `

`r kable(print(CreateTableOne("BMI_for_age", data=df_nine, factorVars=categorvar_demog, strata="sex", testNonNormal = kruskal.test, smd=F), nonnormal="value"), caption="BMI by sex for 9-year old children") `

`r kable(print(CreateTableOne("chrdisease", data=df_imp, factorVars="chrdisease")), caption="Chronic diseases") `

- No of children who come in contact with at least 1 person belonging in vulnerable groups at home: 
    + in weekdays during the study period: (false, true, NA)
`r summary(df_imp$vulnerable_wd_now>0)`
    + in weekdays before the pandemic:
`r summary(df_imp$vulnerable_weekdays_before>0)`
    + in weekends during the study period:
`r summary(df_imp$vulnerable_weekend_now>0)`
    + in weekends before the pandemic:
`r summary(df_imp$vulnerable_weekend_before>0)`

Compared to the population census data, 2011 (https://www.mof.gov.cy/mof/cystat/statistics.nsf/populationcondition_22main_en/populationcondition_22main_en?OpenForm&sub=2&sel=2), 
our survey participants 

- higher level of parents education (57% & 44% vs 20% with at least uni degree)

- similar town distribution : Nicosia (41% vs 39%), Limassol (25% vs 28%), Larnaka (18% vs 17%), Paphos (6% vs 11%) and Ammochostos (10% vs 6%)

- Similar Overweight and obesity prevalence in Cyprus (9-year olds) based on WHO definition (%) – COSI 2015-2017 (https://www.euro.who.int/__data/assets/pdf_file/0006/372426/WH14_COSI_factsheets_v2.pdf?ua=1)
    + boys: 21% obese
            43% overweight (including obese) 
    + girls: 19% obese, 
             43% overweight (including obese) 

# Descriptives

## Diet  

```{r diet_descriptives, echo=FALSE}

rm(df_nine)

# add child_id variable
df_imp$child_ID <- 1:nrow(df_imp)

sel_diet<-c("fruits_now","fruits_before",
            "veggies_now","veggies_before",
            "meat_now","meat_before",
            "leggumes_now","leggumes_before",
            "fish_now","fish_before",
            "sugar_now","sugar_before",
            "chips_now","chips_before")

# selection of variables
diet_db <- df_imp %>%
  dplyr::select(child_ID, all_of(sel_diet))

# transform dataframe into long dataset
diet_long <- diet_db %>%
  pivot_longer(cols=-child_ID) %>%
  mutate(timeframe=case_when(str_detect(string = name, "before")~"Pre-lockdown",
                                 TRUE~"Post-lockdown"),
         name=case_when(str_detect(name, "veggies")~"vegetables",
                        str_detect(name, "fruits")~"fruits",
                        str_detect(name, "meat")~"meat",
                        str_detect(name, "leggumes")~"legumes",
                        str_detect(name, "fish")~"fish",
                        str_detect(name, "sugar")~"sugar",
                        str_detect(name, "chips")~"snacks"))

# calculate percentages
diet_perc <- diet_long %>%
  group_by(value, name, timeframe) %>%
  summarise(N = length(child_ID), .groups="drop") %>%
  filter(!is.na(value)) %>%
  group_by(timeframe, name) %>%
  mutate(
    SumN = sum(N),
    Proportion = proportions(N),
    Perc = round(((N / sum(N)) * 100), digits = 1))

# ggplots
diet_perc %>% 
  filter(name=="vegetables"|name=="fruits") %>% 
ggplot(aes(x = timeframe, y = Perc, fill = value)) +
   geom_col(position = "fill") +
  scale_y_continuous(labels = scales::percent) +
  facet_wrap(~name)+
  ylab('Participants (%)') +
  xlab("") + coord_flip() +
     guides(fill = guide_legend(reverse = TRUE))+
  theme(axis.text.x = element_text(vjust = 1, hjust=0.5,colour = "black",size = 9),
        legend.title = element_blank(),
        legend.position="bottom",
        legend.text = element_text(size=9),
        legend.spacing.x = unit(0.1, 'cm'),
        axis.title.x = element_text(size=9),
        axis.text.y = element_text(colour = "black", size = 9),
        panel.grid.major = element_blank(),panel.grid.minor = element_blank(),
        panel.border = element_blank(),panel.background = element_blank())

diet_perc %>% 
  filter(name=="meat"|name=="legumes"|name=="fish") %>% 
ggplot(aes(x = timeframe, y = Perc, fill = value)) +
   geom_col(position = "fill") +
  scale_fill_discrete(na.translate=FALSE) +
  scale_y_continuous(labels = scales::percent) +
  facet_wrap(~name)+
  ylab('Participants (%)') +
  xlab("") + coord_flip() +
       guides(fill = guide_legend(reverse = TRUE))+
  theme(axis.text.x = element_text(vjust = 1, hjust=0.5,colour = "black",size =9),
        legend.title = element_blank(),
        legend.position="bottom",
        legend.text = element_text(size=9),
        legend.spacing.x = unit(0.1, 'cm'),
        axis.title.x = element_text(size=9),
        axis.text.y = element_text(colour = "black", size = 9),
        panel.grid.major = element_blank(),panel.grid.minor = element_blank(),
        panel.border = element_blank(),panel.background = element_blank())

diet_perc %>% 
  filter(name=="sugar"|name=="snacks") %>% 
ggplot(aes(x = timeframe, y = Perc, fill = value)) +
   geom_col(position = "fill") +
  scale_y_continuous(labels = scales::percent) +
    scale_fill_discrete(na.translate=FALSE) +
  facet_wrap(~name)+
  ylab('Participants (%)') +
  xlab("") + coord_flip() +
       guides(fill = guide_legend(reverse = TRUE))+
 theme(axis.text.x = element_text(vjust = 1, hjust=0.5,colour = "black",size = 9),
        legend.title = element_blank(),
        legend.position="bottom",
        legend.text = element_text(size=9),
        legend.spacing.x = unit(0.1, 'cm'),
        axis.title.x = element_text(size=9),
        axis.text.y = element_text(colour = "black", size = 9),
        panel.grid.major = element_blank(),panel.grid.minor = element_blank(),
        panel.border = element_blank(),panel.background = element_blank())

sel_diet_2<-c("breakfast_now","breakfast_before",
            "supplements_now","supplements_before",
            "fast_food_now","fast_food_before")

# selection of variables
diet_db_2 <- df_imp %>%
  dplyr::select(child_ID, all_of(sel_diet_2))

# transform dataframe into long dataset
diet_long_2 <- diet_db_2 %>%
  pivot_longer(cols=-child_ID) %>%
  mutate(timeframe=case_when(str_detect(string = name, "before")~"Pre-lockdown",
                                 TRUE~"Post-lockdown"),
         name=case_when(str_detect(name, "breakfast")~"breakfast",
                        str_detect(name, "supplements")~"supplements",
                        str_detect(name, "fast_food")~"not homemade food"))

# calculate percentages
diet_perc_2 <- diet_long_2 %>%
  group_by(value, name, timeframe) %>%
  summarise(N = length(child_ID), .groups="drop") %>%
  filter(!is.na(value)) %>%
  group_by(timeframe, name) %>%
  mutate(
    SumN = sum(N),
    Proportion = proportions(N),
    Perc = round(((N / sum(N)) * 100), digits = 1))

# ggplots
diet_perc_2 %>%
  filter(name=="not homemade food") %>%
ggplot(aes(x = timeframe, y = Perc, fill = value)) +
   geom_col(position = "fill") +
  scale_y_continuous(labels = scales::percent) +
  scale_fill_discrete(na.translate=FALSE) +
  facet_wrap(~name)+
  ylab('Participants (%)') +
  xlab("") + coord_flip() +
       guides(fill = guide_legend(reverse = TRUE))+
  theme(axis.text.x = element_text(vjust = 1, hjust=0.5,colour = "black",size = 9),
        legend.title = element_blank(),
        legend.position="bottom",
        legend.text = element_text(size=9),
        legend.spacing.x = unit(0.1, 'cm'),
        axis.title.x = element_text(size=9),
        axis.text.y = element_text(colour = "black", size = 9),
        panel.grid.major = element_blank(),panel.grid.minor = element_blank(),
        panel.border = element_blank(),panel.background = element_blank())

diet_perc_2 %>%
  filter(name=="breakfast") %>%
ggplot(aes(x = timeframe, y = Perc, fill = value)) +
   geom_col(position = "fill") +
  scale_y_continuous(labels = scales::percent) +
      scale_fill_discrete(na.translate=FALSE) +
  facet_wrap(~name)+
  ylab('Participants (%)') +
  xlab("") + coord_flip() +
     guides(fill = guide_legend(reverse = TRUE))+
  theme(axis.text.x = element_text(vjust = 1, hjust=0.5,colour = "black",size = 9),
        legend.title = element_blank(),
        legend.position="bottom",
        legend.text = element_text(size=9),
        legend.spacing.x = unit(0.1, 'cm'),
        axis.title.x = element_text(size=9),
        axis.text.y = element_text(colour = "black", size = 9),
        panel.grid.major = element_blank(),panel.grid.minor = element_blank(),
        panel.border = element_blank(),panel.background = element_blank())

diet_perc_2 %>%
  filter(name=="supplements") %>%
ggplot(aes(x = timeframe, y = Perc, fill = value)) +
   geom_col(position = "fill") +
  scale_y_continuous(labels = scales::percent) +
      scale_fill_discrete(na.translate=FALSE) +
  facet_wrap(~name)+
  ylab('Participants (%)') +
  xlab("") + coord_flip() +
     guides(fill = guide_legend(reverse = TRUE))+
  theme(axis.text.x = element_text(vjust = 1, hjust=0.5,colour = "black",size = 9),
        legend.title = element_blank(),
        legend.position="bottom",
        legend.text = element_text(size=9),
        legend.spacing.x = unit(0.1, 'cm'),
        axis.title.x = element_text(size=9),
        axis.text.y = element_text(colour = "black", size = 9),
        panel.grid.major = element_blank(),panel.grid.minor = element_blank(),
        panel.border = element_blank(),panel.background = element_blank())

# summary tables - data prep
onlyfruits<-diet_long %>% filter(name=="fruits") %>% droplevels(.)
onlyveg<-diet_long %>% filter(name=="vegetables") %>% droplevels(.)
onlymeat<-diet_long %>% filter(name=="meat") %>% droplevels(.)
onlylegumes<-diet_long %>% filter(name=="legumes") %>% droplevels(.)
onlyfish<-diet_long %>% filter(name=="fish") %>% droplevels(.)
onlysug<-diet_long %>% filter(name=="sugar") %>% droplevels(.)
onlysnacks<-diet_long %>% filter(name=="snacks") %>% droplevels(.)
onlyfastfood<-diet_long_2 %>% filter(name=="not homemade food") %>% droplevels(.)
onlybreakfast<-diet_long_2 %>% filter(name=="breakfast") %>% droplevels(.)
onlysup<-diet_long_2 %>% filter(name=="supplements") %>% droplevels(.)

```

`r kable(print(CreateCatTable(vars="value", strata ="timeframe", data=onlyfruits)), caption="N [%] consumption frequency for fruits before and during the measures") `
`r kable(print(CreateCatTable(vars="value", strata ="timeframe", data=onlyveg)), caption="N [%] consumption frequency for vegetables before and during the measures")`
`r kable(print(CreateCatTable(vars="value", strata ="timeframe", data=onlymeat)), caption="N [%] consumption frequency for meat before and during the measures")`
`r kable(print(CreateCatTable(vars="value", strata ="timeframe", data=onlylegumes)), caption="N [%] consumption frequency for legumes before and during the measures")`
`r kable(print(CreateCatTable(vars="value", strata ="timeframe", data=onlyfish)), caption="N [%] consumption frequency for fish before and during the measures")`
`r kable(print(CreateCatTable(vars="value", strata ="timeframe", data=onlysug)), caption="N [%] consumption frequency for sugar before and during the measures")`
`r kable(print(CreateCatTable(vars="value", strata ="timeframe", data=onlysnacks)), caption="N [%] consumption frequency for snacks before and during the measures")`
`r kable(print(CreateCatTable(vars="value", strata ="timeframe", data=onlyfastfood)), caption="N [%] consumption frequency for not homemade food before and during the measures")`
`r kable(print(CreateCatTable(vars="value", strata ="timeframe", data=onlybreakfast)), caption="N [%] consumption frequency for breakfast before and during the measures")`
`r kable(print(CreateCatTable(vars="value", strata ="timeframe", data=onlysup)), caption="N [%] consumption frequency for supplements before and during the measures")`

## Physical activity 

```{r activity_descriptives, echo=FALSE}

rm(list=ls(pattern="^diet"))
rm(list=ls(pattern="only"))

sel_physical_act <- 
  c("walking_now","walking_before",
    "cycling_now","cycling_before", 
    "running_now","running_before",
    "swimming_now","swimming_before",
    "ballet_now","ballet_before",
    "ball_now","ball_before",
    "karate_now","karate_before",
    "other_activity_now","other_activity_before")

# selection of variables
phys_db <- df_imp %>%
  dplyr::select(child_ID, all_of(sel_physical_act))

# transform dataframe into long dataset
phys_long <- phys_db %>%
  pivot_longer(cols=-child_ID) %>%
  mutate(timeframe=case_when(str_detect(string = name, "before")~"Pre-lockdown",
                                 TRUE~"Post-lockdown"),
         name=case_when(str_detect(name, "walking")~"walking",
                        str_detect(name, "cycling")~"cycling",
                        str_detect(name, "running")~"running",
                        str_detect(name, "swimming")~"swimming",
                        str_detect(name, "ballet")~"ballet",
                        str_detect(name, "ball")~"football",
                        str_detect(name, "karate")~"karate",
                        str_detect(name, "other_")~"other activity"))

# calculate percentages
phys_perc <- phys_long %>%
  group_by(value, name, timeframe) %>%
  summarise(N = length(child_ID), .groups="drop") %>%
  filter(!is.na(value)) %>%
  group_by(timeframe, name) %>%
  mutate(
    SumN = sum(N),
    Proportion = proportions(N),
    Perc = round(((N / sum(N)) * 100), digits = 1))

# ggplot
ggplot(phys_perc, aes(x = timeframe, y = Perc, fill = value)) +
   geom_col(position = "fill") +
  scale_y_continuous(labels = scales::percent) +
  facet_wrap(~name)+
  ylab('Participants (%)') +
  xlab("") + coord_flip() +
       guides(fill = guide_legend(reverse = TRUE))+
  theme(axis.text.x = element_text(vjust = 1, hjust=0.5,colour = "black", size =9),
        legend.title = element_blank(),
        legend.position="bottom",
        legend.text = element_text(size=9),
        legend.spacing.x = unit(0.1, 'cm'),
        axis.title.x = element_text(size=9),
        axis.text.y = element_text(colour = "black", size = 9),
        panel.grid.major = element_blank(),panel.grid.minor = element_blank(),
        panel.border = element_blank(),panel.background = element_blank())

# create variable for spare activity score based on PAQ-C scoring rules to get one score for spare time activity for before and now
# Item 1 (Spare time activity): Take the mean of all activities (“no” activity being a 1, “7 times or more” being a 5) on the activity checklist to form a composite score for item 1.

df_num <- df_imp %>% 
               mutate_at(c("walking_now","walking_before",
                            "cycling_now","cycling_before", 
                            "running_now","running_before",
                            "swimming_now","swimming_before",
                            "ballet_now","ballet_before",
                            "ball_now","ball_before",
                            "karate_now","karate_before",
                            "other_activity_now","other_activity_before"), 
                .funs = function(t) 
                  as.numeric(as.character(revalue(x = t,
                  replace = c(
                               "No" = "1",
                               "1-2 times per week" = "2",
                               "3-4 times per week" = "3",
                               "5-6 times per week"= "4",
                               "7 times or more per week"= "5"
                               ))))) 

# Average items to form spare activity score for now and before
df_num <- df_num %>%
  dplyr::mutate(spare_act_now = rowMeans(
                              dplyr::select(., 
                                            dplyr::one_of(
                                  c("walking_now",
                            "cycling_now",
                            "running_now",
                            "swimming_now",
                            "ballet_now",
                            "ball_now",
                            "karate_now",
                            "other_activity_now")))))

df_num <- df_num %>% 
 dplyr::mutate(spare_act_bef = rowMeans(
                              dplyr::select(., 
                                            dplyr::one_of(
                                  c("walking_before",
                            "cycling_before",
                            "running_before",
                            "swimming_before",
                            "ballet_before",
                            "ball_before",
                            "karate_before",
                            "other_activity_before")))))

# select variables
spareact_db <- df_num %>%
  dplyr::select(child_ID, spare_act_bef, spare_act_now)  

# transform to long dataset
spareact_long <- spareact_db %>%
  pivot_longer(cols=-child_ID) %>%
  mutate(timeframe=case_when(str_detect(string = name, "_bef")~"Pre_lockdown",
                                 TRUE~"Post_lockdown"),
         name=case_when(str_detect(name, "spare")~"spareactivity"))

# plot
spareact_long %>%
ggplot() +
  geom_violin(aes(x=name, y=value, fill=timeframe))+
  ylab("Spare activity score") +
  xlab("") +
  coord_flip()

# percent change 
spareact_change <-spareact_long %>%
  dplyr::select(child_ID, timeframe, name, value) %>%
  pivot_wider(id_cols = c("child_ID", "name"), names_from="timeframe", values_from="value") %>%
  mutate(change=case_when(Post_lockdown==0 & Pre_lockdown==0 ~ 0,
                          TRUE~((Post_lockdown-Pre_lockdown)/Pre_lockdown)*100)) 

# percent change summary
sum_spareact_change <-spareact_change  %>%
  group_by(name) %>%
  summarise(median=median(change, na.rm = TRUE),
            p25=quantile(change, probs=0.25, na.rm=TRUE),
            p75=quantile(change, probs=0.75, na.rm=TRUE), .groups="drop") %>%
  mutate_at(.vars = c("median", "p25", "p75"), .funs =~ round(., digits=1)) %>%
  mutate(median_iqr=glue('{median} [{p25}, {p75}]')) %>%
  pivot_wider(id_cols=name, names_from=name, values_from=median_iqr) 

sel_phys_2<-c("free_time_now","free_time_before",
          "break_now","break_before",
          "sickness_now","sickness_before")

# selection of variables
phys_db_2 <- df_imp %>%
  dplyr::select(child_ID, all_of(sel_phys_2))

# transform dataframe into long dataset
phys_long_2 <- phys_db_2 %>%
  pivot_longer(cols=-child_ID) %>%
  mutate(timeframe=case_when(str_detect(string = name, "before")~"Pre-lockdown",
                                 TRUE~"Post-lockdown"),
         name=case_when(str_detect(name, "free")~"free time",
                        str_detect(name, "break")~"school break time",
                        str_detect(name, "sickness")~"sickness"))

# calculate percentages
phys_perc_2 <- phys_long_2 %>%
  group_by(value, name, timeframe) %>%
  summarise(N = length(child_ID), .groups="drop") %>%
  filter(!is.na(value)) %>%
  group_by(timeframe, name) %>%
  mutate(
    SumN = sum(N),
    Proportion = proportions(N),
    Perc = round(((N / sum(N)) * 100), digits = 1))

# ggplots

phys_perc_2  %>%
  filter(name=="free time")  %>%
ggplot(aes(x = timeframe, y = Perc, fill = value)) +
   geom_col(position = "fill") +
  scale_y_continuous(labels = scales::percent) +
    scale_fill_discrete(na.translate=FALSE) +
  facet_wrap(~name)+
  ylab('Participants (%)') +
  xlab("") + coord_flip() +
       guides(fill = guide_legend(reverse = TRUE))+
  theme(axis.text.x = element_text(vjust = 1, hjust=0.5,colour = "black", size =9),
        legend.title = element_blank(),
        legend.position="bottom",
        legend.text = element_text(size=6),
        legend.spacing.x = unit(0.1, 'cm'),
        axis.title.x = element_text(size=9),
        axis.text.y = element_text(colour = "black", size = 9),
        panel.grid.major = element_blank(),panel.grid.minor = element_blank(),
        panel.border = element_blank(),panel.background = element_blank())

phys_perc_2  %>%
  filter(name=="school break time")  %>%
ggplot(aes(x = timeframe, y = Perc, fill = value)) +
   geom_col(position = "fill") +
  scale_y_continuous(labels = scales::percent) +
    scale_fill_discrete(na.translate=FALSE) +
  facet_wrap(~name)+
  ylab('Participants (%)') +
  xlab("") + coord_flip() +
       guides(fill = guide_legend(reverse = TRUE))+
theme(axis.text.x = element_text(vjust = 1, hjust=0.5,colour = "black", size =9),
        legend.title = element_blank(),
        legend.position="bottom",
        legend.text = element_text(size=6),
        legend.spacing.x = unit(0.1, 'cm'),
        axis.title.x = element_text(size=9),
        axis.text.y = element_text(colour = "black", size = 9),
        panel.grid.major = element_blank(),panel.grid.minor = element_blank(),
        panel.border = element_blank(),panel.background = element_blank())

phys_perc_2  %>%
  filter(name=="sickness")  %>%
ggplot(aes(x = timeframe, y = Perc, fill = value)) +
   geom_col(position = "fill") +
  scale_y_continuous(labels = scales::percent) +
    scale_fill_discrete(na.translate=FALSE) +
  facet_wrap(~name)+
  ylab('Participants (%)') +
  xlab("") + coord_flip() +
  theme(axis.text.x = element_text(vjust = 1, hjust=0.5,colour ="black", size=9),
        legend.title = element_blank(),
        legend.position="bottom",
        legend.text = element_text(size=9),
        legend.spacing.x = unit(0.1, 'cm'),
        axis.title.x = element_text(size=9),
        axis.text.y = element_text(colour = "black", size = 9),
        panel.grid.major = element_blank(),panel.grid.minor = element_blank(),
        panel.border = element_blank(),panel.background = element_blank())

# summary tables - data prep
onlywalk<-phys_long %>% filter(name=="walking") %>% droplevels(.)
onlycyc<-phys_long %>% filter(name=="cycling") %>% droplevels(.)
onlyrun<-phys_long %>% filter(name=="running") %>% droplevels(.)
onlyballet<-phys_long %>% filter(name=="ballet") %>% droplevels(.)
onlyfootball<-phys_long %>% filter(name=="football") %>% droplevels(.)
onlykarate<-phys_long %>% filter(name=="karate") %>% droplevels(.)
onlyswim<-phys_long %>% filter(name=="swimming") %>% droplevels(.)
onlyotheract<-phys_long %>% filter(name=="other activity") %>% droplevels(.)
onlyfreetime<-phys_long_2 %>% filter(name=="free time") %>% droplevels(.)
onlyschbreak<-phys_long_2 %>% filter(name=="school break time") %>% droplevels(.)
onlysick<-phys_long_2 %>% filter(name=="sickness") %>% droplevels(.)

```

`r kable(print(CreateTableOne("value", data=spareact_long, strata="timeframe", testNonNormal = kruskal.test, smd=F), nonnormal="value"), caption="Spare activity score of participants - median [IQR]") `

`r kable(print(CreateCatTable(vars="value", strata ="timeframe", data=onlywalk)), caption="Walking N [%] frequency before and during the measures") `
`r kable(print(CreateCatTable(vars="value", strata ="timeframe", data=onlycyc)), caption="Cycling N [%] frequency before and during the measures") `
`r kable(print(CreateCatTable(vars="value", strata ="timeframe", data=onlyrun)), caption="Running N [%] frequency before and during the measures") `
`r kable(print(CreateCatTable(vars="value", strata ="timeframe", data=onlyballet)), caption="Ballet N [%] frequency before and during the measures") `
`r kable(print(CreateCatTable(vars="value", strata ="timeframe", data=onlyfootball)), caption="Football N [%] frequency before and during the measures") `
`r kable(print(CreateCatTable(vars="value", strata ="timeframe", data=onlykarate)), caption="Karate N [%] frequency before and during the measures") `
`r kable(print(CreateCatTable(vars="value", strata ="timeframe", data=onlyswim)), caption="Swimming N [%] frequency before and during the measures") `
`r kable(print(CreateCatTable(vars="value", strata ="timeframe", data=onlyotheract)), caption="Other activity N [%] frequency before and during the measures") `
`r kable(print(CreateCatTable(vars="value", strata ="timeframe", data=onlyfreetime)), caption="Free time N [%] frequency before and during the measures") `
`r kable(print(CreateCatTable(vars="value", strata ="timeframe", data=onlyschbreak)), caption="School break time N [%] frequency before and during the measures") `
`r kable(print(CreateCatTable(vars="value", strata ="timeframe", data=onlysick)), caption="Sickness N [%] frequency before and during the measures") `

`r kable(sum_spareact_change, caption="Change in spare activity score - median [IQR]")`

## Parents smoking in the house 

```{r smoking_descriptives, echo=FALSE}

rm(list=ls(pattern="only"))
rm(list=ls(pattern="^phys"))
rm(spareact_db,spareact_long,sum_spareact_change)
   
# selection of variables
sel_smoking_db <- df_imp %>%
  dplyr::select(child_ID, c("smoking_now","smoking_before"))

# transform dataframe into long dataset
smoking_long <- sel_smoking_db %>%
  pivot_longer(cols=-child_ID) %>%
  mutate(timeframe=case_when(str_detect(string = name, "before")~"Pre-lockdown",
                                 TRUE~"Post-lockdown"),
         name=case_when(str_detect(name, "smoking")~"smoking"))

# calculate percentages
smoking_perc <- smoking_long %>%
  group_by(value, timeframe) %>%
  summarise(N = length(child_ID), .groups="drop") %>%
  filter(!is.na(value)) %>%
  group_by(timeframe) %>%
  mutate(
    SumN = sum(N),
    Proportion = proportions(N),
    Perc = round(((N / sum(N)) * 100), digits = 1))

# ggplot
ggplot(smoking_perc, aes(x = timeframe, y = Perc, fill = value)) +
   geom_col(position = "fill") +
  scale_y_continuous(labels = scales::percent) +
  ylab('Participants (%)') +
  xlab("Smoking") + coord_flip() +
         guides(fill = guide_legend(reverse = TRUE))+
theme(axis.text.x = element_text(vjust = 1, hjust=0.5,colour = "black", size =9),
        legend.title = element_blank(),
        legend.position="bottom",
        legend.text = element_text(size=9),
        legend.spacing.x = unit(0.1, 'cm'),
        axis.title.x = element_text(size=9),
        axis.text.y = element_text(colour = "black", size = 9),
        panel.grid.major = element_blank(),panel.grid.minor = element_blank(),
        panel.border = element_blank(),panel.background = element_blank())

sel_smoking_no <- 
  c("cigs_per_day_before","cigs_per_week_before",
    "cigs_per_day_now","cigs_per_week_now")

# selection of variables
smoking_db_2 <- df_imp %>%
  dplyr::select(child_ID, all_of(sel_smoking_no)) 

# transform dataframe into long dataset
smoking_long_2 <- smoking_db_2 %>%
  pivot_longer(cols=-child_ID) %>%
  mutate(timeframe=case_when(str_detect(string = name, "b")~"Pre-lockdown",
                                 TRUE~"Post-lockdown"),
         name=case_when(str_detect(name, "_day")~"cigsperday",
                        str_detect(name, "_week")~"cigsperweek"))

# plot
smoking_long_2 %>%
  filter(value<quantile(value, probs=0.99, na.rm=TRUE))   %>%
ggplot() +
  geom_violin(aes(x=name, y=value, fill=timeframe))+
  ylab("") +
  xlab("") +
  coord_flip()

# summary tables - data prep
onlycigsday <- smoking_long_2 %>% filter(name=="cigsperday") %>% droplevels(.)
onlycigsweek <- smoking_long_2 %>% filter(name=="cigsperweek") %>% droplevels(.)

# replace NA with 0 for use in correlation and regression
smoking_db_3 <- smoking_db_2 %>%
  mutate(cig_day_b=case_when(is.na(cigs_per_day_before)~0,
         TRUE~as.numeric(cigs_per_day_before))) %>%
  mutate(cig_day_n=case_when(is.na(cigs_per_day_now)~0,
         TRUE~as.numeric(cigs_per_day_now))) %>%
  mutate(cig_week_b=case_when(is.na(cigs_per_week_before)~0,
         TRUE~as.numeric(cigs_per_day_before))) %>%
  mutate(cig_week_n=case_when(is.na(cigs_per_week_now)~0,
         TRUE~as.numeric(cigs_per_week_now)))

# transform dataframe into long dataset
smoking_long_3 <- smoking_db_3 %>%
  select(-c("cigs_per_day_before","cigs_per_week_before",
    "cigs_per_day_now","cigs_per_week_now")) %>%
  pivot_longer(cols=-child_ID) %>%
  mutate(timeframe=case_when(str_detect(string = name, "b")~"Pre_lockdown",
                                 TRUE~"Post_lockdown"),
         name=case_when(str_detect(name, "_day")~"cigsperday",
                        str_detect(name, "_week")~"cigsperweek"))

# percent change 
cig_change <-smoking_long_3 %>%
  dplyr::select(child_ID, timeframe, name, value) %>%
  pivot_wider(id_cols = c("child_ID", "name"), names_from="timeframe", values_from="value") %>%
  mutate(change=case_when(Post_lockdown==0 & Pre_lockdown==0 ~ 0,
                          TRUE~((Post_lockdown-Pre_lockdown)/Pre_lockdown)*100)) 

# percent change summary
sum_cig_change <-cig_change  %>%
  group_by(name) %>%
  summarise(median=median(change, na.rm = TRUE),
            p25=quantile(change, probs=0.25, na.rm=TRUE),
            p75=quantile(change, probs=0.75, na.rm=TRUE), .groups="drop") %>%
  mutate_at(.vars = c("median", "p25", "p75"), .funs =~ round(., digits=1)) %>%
  mutate(median_iqr=glue('{median} [{p25}, {p75}]')) %>%
  pivot_wider(id_cols=name, names_from=name, values_from=median_iqr) 
```

`r kable(print(CreateCatTable("value", strata ="timeframe", data=smoking_long)), caption="Smoking n [%] frequency before and during the measures") `

`r kable(print(CreateTableOne("value", data=onlycigsday, strata="timeframe", testNonNormal = kruskal.test, smd=F), nonnormal="value"), caption="Cigarettes no per day - median [IQR]") `

`r kable(print(CreateTableOne("value", data=onlycigsweek, strata="timeframe", testNonNormal = kruskal.test, smd=F), nonnormal="value"), caption="Cigarettes no per week - median [IQR]") `

`r kable(sum_cig_change, caption="Change in cig no per day/week - median [IQR] (both non-smokers and smokers)")`

## Communication and screen time 

```{r communic_descriptives, echo=FALSE}

rm(list=ls(pattern="only"))
rm(list=ls(pattern="^smoking"))
rm(sum_cig_change, sum_cig_change_s, sel_smoking_db)

sel_comm_screen<-c("net_friends_now","net_friends_before",
               "net_family_now","net_family_before",
               "tv_pc_now","tv_pc_before")

# selection of variables
comm_db <- df_imp %>%
  dplyr::select(child_ID, all_of(sel_comm_screen))

# transform dataframe into long dataset
comm_long <- comm_db %>%
  pivot_longer(cols=-child_ID) %>%
  mutate(timeframe=case_when(str_detect(string = name, "before")~"Pre-lockdown",
                                 TRUE~"Post-lockdown"),
         name=case_when(str_detect(name, "friends")~"friends communication",
                        str_detect(name, "family")~"family communication",
                        str_detect(name, "tv")~"screen time"))

# calculate percentages
comm_perc <- comm_long %>%
  group_by(value, name, timeframe) %>%
 summarise(N = length(child_ID), .groups="drop") %>%
  filter(!is.na(value)) %>%
  group_by(timeframe, name) %>%
  mutate(
    SumN = sum(N),
    Proportion = proportions(N),
    Perc = round(((N / sum(N)) * 100), digits = 1))

# ggplot
comm_perc %>%
  filter(name != "screen time") %>%
ggplot(aes(x = timeframe, y = Perc, fill = value)) +
   geom_col(position = "fill") +
  scale_y_continuous(labels = scales::percent) +
  facet_wrap(~name)+
  ylab('Participants (%)') +
  xlab("") + coord_flip() +
       guides(fill = guide_legend(reverse = TRUE))+
theme(axis.text.x = element_text(vjust = 1, hjust=0.5,colour = "black", size =9),
        legend.title = element_blank(),
        legend.position="bottom",
        legend.text = element_text(size=9),
        legend.spacing.x = unit(0.1, 'cm'),
        axis.title.x = element_text(size=9),
        axis.text.y = element_text(colour = "black", size = 9),
        panel.grid.major = element_blank(),panel.grid.minor = element_blank(),
        panel.border = element_blank(),panel.background = element_blank())

comm_perc %>%
  filter(name == "screen time") %>%
ggplot(aes(x = timeframe, y = Perc, fill = value)) +
   geom_col(position = "fill") +
  scale_y_continuous(labels = scales::percent) +
  facet_wrap(~name)+
  ylab('Participants (%)') +
  xlab("") + coord_flip() +
       guides(fill = guide_legend(reverse = TRUE))+
theme(axis.text.x = element_text(vjust = 1, hjust=0.5,colour = "black", size =9),
        legend.title = element_blank(),
        legend.position="bottom",
        legend.text = element_text(size=9),
        legend.spacing.x = unit(0.1, 'cm'),
        axis.title.x = element_text(size=9),
        axis.text.y = element_text(colour = "black", size = 9),
        panel.grid.major = element_blank(),panel.grid.minor = element_blank(),
        panel.border = element_blank(),panel.background = element_blank())

sel_comm_screen_2 <- 
  c("hours_friends_now","hours_friends_before",
    "hours_family_now","hours_family_before")

# selection of variables
comm_db_2 <- df_imp %>%
  dplyr::select(child_ID, all_of(sel_comm_screen_2))

# transform dataframe into long dataset
comm_long_2 <- comm_db_2 %>%
  pivot_longer(cols=-child_ID) %>%
  mutate(timeframe=case_when(str_detect(string = name, "before")~"before",
                                 TRUE~"now"),
         name=case_when(str_detect(name, "friends")~"friends",
                        str_detect(name, "family")~"family"))

# plot
comm_long_2 %>%
  filter(value<quantile(value, probs=0.95, na.rm=TRUE))   %>%
ggplot() +
  geom_violin(aes(x=name, y=value, fill=timeframe))+
  ylab("Hours per day") +
  xlab("") +
  coord_flip()

# summary tables - data prep
onlyfriend<-comm_long %>% filter(name=="friends communication") %>% droplevels(.)
onlyfamily<-comm_long %>% filter(name=="family communication") %>% droplevels(.)
onlyscreen<-comm_long %>% filter(name=="screen time") %>% droplevels(.)
onlyfriendh<-comm_long_2 %>% filter(name=="family") %>% droplevels(.)
onlyfamilyh<-comm_long_2 %>% filter(name=="friends") %>% droplevels(.)
```

`r kable(print(CreateCatTable(vars="value", strata ="timeframe", data=onlyfriend)), caption="Friends communication N [%] frequency before and during the measures") `
`r kable(print(CreateCatTable(vars="value", strata ="timeframe", data=onlyfamily)), caption="Family communication N [%] frequency before and during the measures") `
`r kable(print(CreateCatTable(vars="value", strata ="timeframe", data=onlyscreen)), caption="Screen time N [%] frequency before and during the measures") `

`r kable(print(CreateTableOne("value", data=onlyfriendh, strata="timeframe", testNonNormal = kruskal.test, smd=F), nonnormal="value"), caption="Friends communication hours per day - median [IQR]") `
`r kable(print(CreateTableOne("value", data=onlyfamilyh, strata="timeframe", testNonNormal = kruskal.test, smd=F), nonnormal="value"), caption="Family communication hours per day - median [IQR]") `

    
## Children personal hygiene 

```{r hygiene_descriptives, echo=FALSE}

rm(list=ls(pattern="only"))
rm(list=ls(pattern="^comm"))

sel_hygiene<-c("hands_soap_now","hands_soap_before",
               "hands_anticeptic_now","hands_anticeptic_before")

# selection of variables
hyg_db <- df_imp %>%
  dplyr::select(child_ID, all_of(sel_hygiene))


# transform dataframe into long dataset
hyg_long <- hyg_db %>%
  pivot_longer(cols=-child_ID) %>%
  mutate(timeframe=case_when(str_detect(string = name, "before")~"Pre-lockdown",
                                 TRUE~"Post-lockdown"),
         name=case_when(str_detect(name, "soap")~"hand soap",
                        str_detect(name, "anticeptic")~"hand antiseptic")) 

# calculate percentages
hyg_perc <- hyg_long %>%
  group_by(value, name, timeframe) %>%
  summarise(N = length(child_ID), .groups="drop") %>%
  filter(!is.na(value)) %>%
  group_by(timeframe, name) %>%
  mutate(
    SumN = sum(N),
    Proportion = proportions(N),
    Perc = round(((N / sum(N)) * 100), digits = 1)) %>%
mutate_at(.vars = c("value"), 
            .funs = list(~fct_relevel(., "More than 7 times per day", "4-7 times per day", "1-3 times per day","Less than 1 time per day","Never")))

# ggplot

ggplot(hyg_perc, aes(x = timeframe, y = Perc, fill = value)) +
   geom_col(position = "fill") +
  scale_y_continuous(labels = scales::percent) +
  facet_wrap(~name)+
  ylab('Participants (%)') +
  xlab("") + coord_flip() +
         guides(fill = guide_legend(reverse = TRUE))+
theme(axis.text.x = element_text(vjust = 1, hjust=0.5,colour = "black", size =9),
        legend.title = element_blank(),
        legend.position="bottom",
        legend.text = element_text(size=9),
        legend.spacing.x = unit(0.1, 'cm'),
        axis.title.x = element_text(size=9),
        axis.text.y = element_text(colour = "black", size = 9),
        panel.grid.major = element_blank(),panel.grid.minor = element_blank(),
        panel.border = element_blank(),panel.background = element_blank())

# summary tables - data prep
onlyantisep<-hyg_long %>% filter(name=="hand antiseptic") %>% droplevels(.)
onlysoap<-hyg_long %>% filter(name=="hand soap") %>% droplevels(.)

```

`r kable(print(CreateCatTable(vars="value", strata ="timeframe", data=onlyantisep)), caption="Antiseptic use for hands N [%] frequency before and during the measures") `
`r kable(print(CreateCatTable(vars="value", strata ="timeframe", data=onlysoap)), caption="Hand washing with soap N [%] frequency before and during the measures") `

## Home cleaning activities

```{r cleaning_descriptives, echo=FALSE}

rm(list=ls(pattern="only"))
rm(list=ls(pattern="^hyg"))

sel_cleaning <- 
  c("bath_clean_anticept_now","bath_clean_anticept_before",
    "kitchen_anticeptic_now","kitchen_anticeptic_before",
    "mopping_anticeptic_now", "mopping_anticeptic_before",
    "other_cleaning_now","other_cleaning_before",
    "all_cleaning_now","all_cleaning_before")

# select variables
cleaning_db <- df_imp %>%
  dplyr::select(child_ID, all_of(sel_cleaning))  

# transform to long dataset
cleaning_long <- cleaning_db %>%
  pivot_longer(cols=-child_ID) %>%
  mutate(timeframe=case_when(str_detect(string = name, "_b")~"Pre_lockdown",
                                 TRUE~"Post_lockdown"),
         name=case_when(str_detect(name, "bath")~"bath",
                             str_detect(name, "kitchen")~"kitchen",
                             str_detect(name, "mopping")~"mopping",
                             str_detect(name, "other")~"other surfaces",
                            str_detect(name, "all")~"total"))

# plots
cleaning_long %>%
  filter(!name == "total") %>%
ggplot() +
  geom_violin(aes(x=name, y=value, fill=timeframe))+
  ylab("Cleaning - times per week") +
  xlab("") +
  coord_flip()

cleaning_long %>%
filter(name == "total") %>%
  ggplot() +
  geom_violin(aes(x=name, y=value, fill=timeframe))+
  ylab("Cleaning - times per week") +
  xlab("") +
  coord_flip()

# percent change 
cleaning_change <-cleaning_long %>%
  dplyr::select(child_ID, timeframe, name, value) %>%
  pivot_wider(id_cols = c("child_ID", "name"), names_from="timeframe", values_from="value") %>%
  mutate(change=case_when(Post_lockdown==0 & Pre_lockdown==0 ~ 0,
                          TRUE~((Post_lockdown-Pre_lockdown)/Pre_lockdown)*100)) 
 
# percent change summary
sum_cleaning_change <-cleaning_change  %>%
  group_by(name) %>%
  summarise(median=median(change, na.rm = TRUE),
            p25=quantile(change, probs=0.25, na.rm=TRUE),
            p75=quantile(change, probs=0.75, na.rm=TRUE), .groups="drop") %>%
  mutate_at(.vars = c("median", "p25", "p75"), .funs =~ round(., digits=1)) %>%
  mutate(median_iqr=glue('{median} [{p25}, {p75}]')) %>%
  pivot_wider(id_cols=name, names_from=name, values_from=median_iqr)

# convert variables from integers into factors
df_num[, c("bath_clean_anticept_now","bath_clean_anticept_before",
    "kitchen_anticeptic_now","kitchen_anticeptic_before",
    "mopping_anticeptic_now", "mopping_anticeptic_before",
    "other_cleaning_now","other_cleaning_before")]<-             
  lapply(df_imp[, c("bath_clean_anticept_now","bath_clean_anticept_before",
    "kitchen_anticeptic_now","kitchen_anticeptic_before",
    "mopping_anticeptic_now", "mopping_anticeptic_before",
    "other_cleaning_now","other_cleaning_before")], factor)

# select variables
cleaning_db_2 <- df_num %>%
  dplyr::select(child_ID, all_of(sel_cleaning), -c(all_cleaning_now,all_cleaning_before))  

# transform to long dataset
cleaning_long_2 <- cleaning_db_2 %>%
  pivot_longer(cols=-child_ID) %>%
  mutate(timeframe=case_when(str_detect(string = name, "_b")~"Pre-lockdown",
                                 TRUE~"Post-lockdown"),
         name=case_when(str_detect(name, "bath")~"bath",
                             str_detect(name, "kitchen")~"kitchen",
                             str_detect(name, "mopping")~"mopping",
                             str_detect(name, "other")~"other surfaces"))

# calculate percentages
clea_perc <- cleaning_long_2 %>%
  group_by(value, name, timeframe) %>%
  summarise(N = length(child_ID), .groups="drop") %>%
  filter(!is.na(value)) %>%
  group_by(timeframe, name) %>%
  mutate(
    SumN = sum(N),
    Proportion = proportions(N),
    Perc = round(((N / sum(N)) * 100), digits = 1))

# ggplot
ggplot(clea_perc, aes(x = timeframe, y = Perc, fill = value)) +
   geom_col(position = "fill") +
  scale_y_continuous(labels = scales::percent) +
  facet_wrap(~name)+
  ylab('Participants (%)') +
  xlab("") + coord_flip() +
       guides(fill = guide_legend(reverse = TRUE))+
theme(axis.text.x = element_text(vjust = 1, hjust=0.5,colour = "black",size = 9),
        legend.title = element_blank(),
        legend.position="bottom",
        legend.text = element_text(size=9),
        legend.spacing.x = unit(0.1, 'cm'),
        axis.title.x = element_text(size=9),
        axis.text.y = element_text(colour = "black", size = 9),
        panel.grid.major = element_blank(),panel.grid.minor = element_blank(),
        panel.border = element_blank(),panel.background = element_blank())

# summary tables - data prep
onlykitchen<-cleaning_long_2 %>% filter(name=="kitchen") %>% droplevels(.)
onlybath<-cleaning_long_2 %>% filter(name=="bath") %>% droplevels(.)
onlymop<-cleaning_long_2 %>% filter(name=="mopping") %>% droplevels(.)
onlyothersurf<-cleaning_long_2 %>% filter(name=="other surfaces") %>% droplevels(.)

onlykitchenn<-cleaning_long %>% filter(name=="kitchen") %>% droplevels(.)
onlybathn<-cleaning_long %>% filter(name=="bath") %>% droplevels(.)
onlymopn<-cleaning_long %>% filter(name=="mopping") %>% droplevels(.)
onlyothersurfn<-cleaning_long %>% filter(name=="other surfaces") %>% droplevels(.)
onlytotalclean<-cleaning_long %>% filter(name=="total") %>% droplevels(.)

```

`r kable(print(CreateCatTable(vars="value", strata ="timeframe", data=onlykitchen)), caption="Kitchen cleaning N [%] frequency before and during the measures") `
`r kable(print(CreateCatTable(vars="value", strata ="timeframe", data=onlybath)), caption="Bathroom cleaning N [%] frequency before and during the measures") `
`r kable(print(CreateCatTable(vars="value", strata ="timeframe", data=onlymop)), caption="Mopping N [%] frequency before and during the measures") `
`r kable(print(CreateCatTable(vars="value", strata ="timeframe", data=onlyothersurf)), caption="Other surfaces cleaning N [%] frequency before and during the measures") `

`r kable(print(CreateTableOne("value", data=onlykitchenn, strata="timeframe", testNonNormal = kruskal.test, smd=F), nonnormal="value"), caption="Kitchen cleaning times per week - median [IQR]") `
`r kable(print(CreateTableOne("value", data=onlybathn, strata="timeframe", testNonNormal = kruskal.test, smd=F), nonnormal="value"), caption="Bathroom cleaning times per week - median [IQR]") `
`r kable(print(CreateTableOne("value", data=onlymopn, strata="timeframe", testNonNormal = kruskal.test, smd=F), nonnormal="value"), caption="Mopping times per week - median [IQR]") `
`r kable(print(CreateTableOne("value", data=onlyothersurfn, strata="timeframe", testNonNormal = kruskal.test, smd=F), nonnormal="value"), caption="Other surfaces cleaning times per week - median [IQR]") `
`r kable(print(CreateTableOne("value", data=onlytotalclean, strata="timeframe", testNonNormal = kruskal.test, smd=F), nonnormal="value"), caption="All house cleaning times per week - median [IQR]") `

`r kable(sum_cleaning_change, caption="Change in cleaning activities times per week at home - median [IQR]")`


## Contacts at home, school and elsewhere and hours at home

```{r contacts, echo=FALSE}

rm(list=ls(pattern="only"))
rm(clea_perc, cleaning_db, cleaning_db_2, cleaning_long, cleaning_long_2, sum_cleaning_change)

sel_contacts<-c("home_persons_wd_now","home_persons_wd_before",
                "school_persons_now","school_persons_before",
                "otherplace_person_wd_now","otherplace_person_wd_before",
                "home_persons_now_we","home_persons_we_before",
                "otherplace_person_we_now","other_place_persons_before",
                "home_hours_wd","home_hours_wd_before",
                "home_hours_we_now","home_hours_we_before",
                "vulnerable_wd_now","vulnerable_weekdays_before",
                "vulnerable_weekend_now","vulnerable_weekend_before")

contacts_db <- df_imp %>%
  dplyr::select(all_of(sel_contacts), all_of(sel_demog), child_ID, submission_time)  %>%
 dplyr::rename(
   home_hours_wd_now="home_hours_wd",
   school_persons_wd_now="school_persons_now",
   school_persons_wd_before="school_persons_before",
   other_place_persons_we_before="other_place_persons_before",
   vulnerable_we_now="vulnerable_weekend_now",
   vulnerable_we_before="vulnerable_weekend_before",
   vulnerable_wd_before="vulnerable_weekdays_before") 
 
number_of_contacts_long <- contacts_db %>%
  dplyr::select(contains("before"), contains("now"),child_ID, age,
                - c("home_hours_wd_now","home_hours_wd_before",
                "home_hours_we_now","home_hours_we_before")) %>%
  pivot_longer(cols=-c("child_ID","age")) %>%
  mutate(timeframe=case_when(str_detect(string = name, "before")~"Pre_lockdown",
                                 TRUE~"Post_lockdown"),
         days=case_when(str_detect(name, "wd")~"wd",
                        str_detect(name, "we")~"we"),
         place=case_when(str_detect(name, "home")~"home",
                         str_detect(name, "vulnerable")~"vuln_home",
                         str_detect(name, "school")~"school",
                         str_detect(name, "other")~"elsewhere")) %>%
  mutate(n_contacts=value)  

# summary contacts weekends
sum_contacts_before_during_we <-  number_of_contacts_long %>%
  filter(days=="we") %>% droplevels(.) %>%
  group_by(place, timeframe) %>%
  summarise(median=median(n_contacts, na.rm = TRUE),
            p25=quantile(n_contacts, probs=0.25, na.rm=TRUE),
            p75=quantile(n_contacts, probs=0.75, na.rm=TRUE), .groups="drop") %>%
  mutate(median_iqr=glue('{median} [{p25}, {p75}]'))

# plot median [IQR] with colouring
ggplot(sum_contacts_before_during_we, aes(x=timeframe, y=place, fill=median))+
  geom_tile(color="grey50") +
  geom_text(aes(label=glue('{median} [{p25}, {p75}]'))) +
  scale_fill_gradient2(low = "#132B43", high = "#56B1F7") +
  theme_minimal() +
  labs(caption ="Median [IQR] number of contacts in weekends",
    y = "",
    x="") 

# summary contacts weekdays 
sum_contacts_before_during_wd <- number_of_contacts_long %>%
  filter(days=="wd") %>% droplevels(.) %>%
  group_by(place, timeframe) %>%
  summarise(median=median(n_contacts, na.rm = TRUE),
            p25=quantile(n_contacts, probs=0.25, na.rm=TRUE),
            p75=quantile(n_contacts, probs=0.75, na.rm=TRUE), .groups="drop") %>%
  mutate(median_iqr=glue("{median} [{p25}-{p75}]")) 

# plot median [IQR] with colouring
ggplot(sum_contacts_before_during_wd, aes(x=timeframe, y=place, fill=median))+
  geom_tile(color="grey50") +
  geom_text(aes(label=glue('{median} [{p25}, {p75}]'))) +
  scale_fill_gradient2(low = "#132B43", high = "#56B1F7") +
  theme_minimal() +
  labs(
    caption =  "Median [IQR] number of contacts in weekdays",
    y = "",
    x="") 

# percent change - weekdays
number_of_contacts_change_wd <-number_of_contacts_long %>%
    filter(days=="wd") %>% droplevels(.) %>%
  dplyr::select(child_ID, timeframe, place, n_contacts) %>%
  pivot_wider(id_cols = c("child_ID", "place"), names_from="timeframe", values_from="n_contacts") %>%
  mutate(change=case_when(Post_lockdown==0 & Pre_lockdown==0 ~ 0,
  TRUE~((Post_lockdown-Pre_lockdown)/Pre_lockdown)*100))

# percent change summary - weekdays
sum_number_of_contacts_change_wd <-number_of_contacts_change_wd  %>%
  group_by(place) %>%
  summarise(median=median(change, na.rm = TRUE),
            p25=quantile(change, probs=0.25, na.rm=TRUE),
            p75=quantile(change, probs=0.75, na.rm=TRUE), .groups="drop") %>%
  mutate_at(.vars = c("median", "p25", "p75"), .funs =~ round(., digits=1)) %>%
  mutate(median_iqr=glue('{median} [{p25}, {p75}]')) %>%
  pivot_wider(id_cols=place, names_from=place, values_from=median_iqr)

# percent change - weekend
number_of_contacts_change_we <-number_of_contacts_long %>%
    filter(days=="we") %>% droplevels(.) %>%
  dplyr::select(child_ID, timeframe, place, n_contacts) %>%
  pivot_wider(id_cols = c("child_ID", "place"), names_from="timeframe", values_from="n_contacts") %>%
  mutate(change=case_when(Post_lockdown==0 & Pre_lockdown==0 ~ 0,
                          TRUE~((Post_lockdown-Pre_lockdown)/Pre_lockdown)*100)) 
 
# percent change summary - weekend
sum_number_of_contacts_change_we <-number_of_contacts_change_we  %>%
  group_by(place) %>%
  summarise(median=median(change, na.rm = TRUE),
            p25=quantile(change, probs=0.25, na.rm=TRUE),
            p75=quantile(change, probs=0.75, na.rm=TRUE), .groups="drop") %>%
mutate_at(.vars = c("median", "p25", "p75"), .funs =~ round(., digits=1)) %>%
  mutate(median_iqr=glue('{median} [{p25}, {p75}]')) %>%
  pivot_wider(id_cols=place, names_from=place, values_from=median_iqr)

# plot of change - weekdays
number_of_contacts_change_wd %>%
  filter(change<quantile(number_of_contacts_change_wd$change, probs=0.95, na.rm=TRUE)) %>%
ggplot() +
  geom_violin(aes(x=place, y=change, fill = place))+
  ylab("% change in the number of contacts - weekdays") +
  xlab("") +
  theme(legend.position="none") + 
  coord_flip()

# plot of change - weekend
number_of_contacts_change_we %>%
  filter(change<quantile(number_of_contacts_change_we$change, probs=0.95, na.rm=TRUE)) %>%
ggplot() +
  geom_violin(aes(x=place, y=change, fill = place)) +
  ylab("% change in the number of contacts - weekend") +
  xlab("") +
  theme(legend.position="none") + 
  coord_flip()

## plot no of contacts and age - weekdays
number_of_contacts_long %>%
  filter(days=="wd") %>%
  #filter(n_contacts<quantile(number_of_contacts_long$n_contacts, probs=0.99, na.rm=TRUE)) %>%
  group_by(child_ID, age, timeframe, place) %>%
  filter(!is.na(value)) %>%
  summarize(sum_contacts=sum(n_contacts), .groups="drop") %>%
  group_by(age, timeframe, place) %>%
  summarize(average_cont=mean(sum_contacts)) %>% 
  mutate_at(.vars = c("place"), 
            .funs = list(~fct_relevel(., "home", "vuln_home", "school", 
                                       "elsewhere"))) %>%  
  mutate(place_labs=fct_recode(place, "At home"="home", "At school"="school", "Elsewhere"="elsewhere", "At home (only vulnerable)"="vuln_home")) %>% 
  ggplot() +
  geom_line(aes(x=age, y=average_cont, color=timeframe), size=1) +
  scale_color_manual(values=c("blue","coral")) +  
  facet_wrap(~place_labs, ncol = 1, scales = "free_y") +
  theme_minimal() +
    theme(legend.position="bottom") +
  labs(y="Average number of contacts - weekdays",
       x="Age (years)")
  
## plot no of contacts and age - weekend
number_of_contacts_long %>%
  filter(days=="we") %>%
  #filter(n_contacts<quantile(number_of_contacts_long$n_contacts, probs=0.99, na.rm=TRUE)) %>%
  group_by(child_ID, age, timeframe, place) %>%
  filter(!is.na(value)) %>%
  summarize(sum_contacts=sum(n_contacts), .groups="drop") %>%
  group_by(age, timeframe, place) %>%
  summarize(average_cont=mean(sum_contacts)) %>% 
  mutate_at(.vars = c("place"), 
            .funs = list(~fct_relevel(., "home", "vuln_home", 
                                       "elsewhere"))) %>%  
  mutate(place_labs=fct_recode(place, "At home"="home", "Elsewhere"="elsewhere", "At home (only vulnerable)"="vuln_home")) %>% 
  ggplot() +
  geom_line(aes(x=age, y=average_cont, color=timeframe), size=1) +
  scale_color_manual(values=c("blue","coral")) +  
  facet_wrap(~place_labs, ncol = 1, scales = "free_y") +
  theme_minimal() +
    theme(legend.position="bottom") +
  labs(y="Average number of contacts - weekend",
       x="Age (years)")

# summary tables - data prep
onlyhomecontactswd<-number_of_contacts_long %>% filter(name==c("home_persons_wd_before","home_persons_wd_now")) %>% droplevels(.)
onlyhomecontactswe<-number_of_contacts_long %>% filter(name==c("home_persons_now_we","home_persons_we_before")) %>% droplevels(.)
onlyvulnhomecontactswe<-number_of_contacts_long %>% filter(name==c("vulnerable_we_before","vulnerable_we_now")) %>% droplevels(.)
onlyvulnhomecontactswd<-number_of_contacts_long %>% filter(name==c("vulnerable_wd_now","vulnerable_wd_before")) %>% droplevels(.)
onlyschcontacts<-number_of_contacts_long %>% filter(name==c("school_persons_wd_now","school_persons_wd_before")) %>% droplevels(.)
onlyothercontactswd<-number_of_contacts_long %>% filter(name==c("otherplace_person_wd_before","otherplace_person_wd_now")) %>% droplevels(.)
onlyothercontactswe<-number_of_contacts_long %>% filter(name==c("other_place_persons_we_before","otherplace_person_we_now")) %>% droplevels(.)
```

`r kable(sum_number_of_contacts_change_wd, caption="Change in contacts number at home (including vulnerable people), school and elsewhere in weekdays - median [IQR]")`

`r kable(sum_number_of_contacts_change_we, caption="Change in contacts number at home (including vulnerable people) and elsewhere in weekends - median [IQR]")`

`r kable(print(CreateTableOne("value", data=onlyhomecontactswd, strata="timeframe", testNonNormal = kruskal.test, smd=F), nonnormal="value"), caption="Contacts at home on weekdays - median [IQR]") `

`r kable(print(CreateTableOne("value", data=onlyhomecontactswe, strata="timeframe", testNonNormal = kruskal.test, smd=F), nonnormal="value"), caption="Contacts at home on weekends - median [IQR]") `

`r kable(print(CreateTableOne("value", data=onlyvulnhomecontactswd, strata="timeframe", testNonNormal = kruskal.test, smd=F), nonnormal="value"), caption="Vulnerable contacts at home on weekdays - median [IQR]") `

`r kable(print(CreateTableOne("value", data=onlyvulnhomecontactswe, strata="timeframe", testNonNormal = kruskal.test, smd=F), nonnormal="value"), caption="Vulnerable contacts at home on weekends - median [IQR]") `

`r kable(print(CreateTableOne("value", data=onlyschcontacts, strata="timeframe", testNonNormal = kruskal.test, smd=F), nonnormal="value"), caption="Contacts at school - median [IQR]") `

`r kable(print(CreateTableOne("value", data=onlyothercontactswd, strata="timeframe", testNonNormal = kruskal.test, smd=F), nonnormal="value"), caption="Contacts elsewhere on weekdays - median [IQR]") `

`r kable(print(CreateTableOne("value", data=onlyothercontactswe, strata="timeframe", testNonNormal = kruskal.test, smd=F), nonnormal="value"), caption="Contacts elsewhere on weekends - median [IQR]") `

## Hours at home

```{r hours_home, echo=FALSE}

rm(list=ls(pattern="only"))

# hours spent at home - weekend and weekdays
time_spent_home_long <- contacts_db %>%
  dplyr::select(contains("home_hours"), child_ID) %>% 
  pivot_longer(cols=-child_ID) %>%
  mutate(timeframe=case_when(str_detect(string = name, "before")~"Pre_lockdown",
                             TRUE~"Post_lockdown"),
         days=case_when(str_detect(name, "wd")~"wd",
                        str_detect(name, "we")~"we"))

# summary hours spent at home - weekend and weekdays
sum_time_spent_home <- time_spent_home_long  %>%
  group_by(days, timeframe) %>%
  summarise(median=median(value, na.rm = TRUE),
            p25=quantile(value, probs=0.25, na.rm=TRUE),
            p75=quantile(value, probs=0.75, na.rm=TRUE), .groups="drop") %>%
  mutate(median_iqr=glue('{median} [{p25}-{p75}]'))

# plot median [IQR] with colouring
ggplot(sum_time_spent_home, aes(x=timeframe, y=days, fill=median))+
  geom_tile(color="grey50") +
  geom_text(aes(label=glue('{median} [{p25}, {p75}]'))) +
  scale_fill_gradient2(low = "#132B43", high = "#56B1F7") +
  theme_minimal() +
  labs(
    caption= "Median [IQR] number of hours spent at home in weekdays and weekends",
    y = "",
    x="") 

# hours change  
time_spent_home_change <-time_spent_home_long %>%
  dplyr::select(child_ID, timeframe, days, value) %>%
  pivot_wider(id_cols = c("child_ID", "days"), names_from=c("timeframe"), values_from="value") %>%
  mutate(change=case_when(Post_lockdown==0 & Pre_lockdown==0 ~ 0,
                          TRUE~((Post_lockdown-Pre_lockdown)/Pre_lockdown)*100)) 

sum_time_spent_home_change <-time_spent_home_change  %>%
  group_by(days) %>%
  summarise(median=median(change, na.rm = TRUE),
            p25=quantile(change, probs=0.25, na.rm=TRUE),
            p75=quantile(change, probs=0.75, na.rm=TRUE), .groups="drop") %>%
  mutate_at(.vars = c("median", "p25", "p75"), .funs =~ round(., digits=1)) %>%
  mutate(median_iqr=glue('{median} [{p25}, {p75}]')) %>%
  pivot_wider(id_cols=days, names_from=days, values_from=median_iqr)

time_spent_home_change %>%
  filter(change<quantile(time_spent_home_change$change, probs=0.95, na.rm=TRUE)) %>%
ggplot() +
  geom_violin(aes(x=days, y=change, fill=days)) +
  ylab("% change in the number of hours staying at home") +
  xlab("") +
  theme(legend.position="none") + 
  scale_fill_brewer(palette="Dark2") +
  # scale_fill_manual(values=c("#E69F00","#56B4E9")) +
  coord_flip()

# summary tables - data prep
onlyhomehourswd<-time_spent_home_long %>% filter(name==c("home_hours_wd_now","home_hours_wd_before")) %>% droplevels(.)
onlyhomehourswe<-time_spent_home_long %>% filter(name==c("home_hours_we_now","home_hours_we_before")) %>% droplevels(.)

```
`r kable(sum_time_spent_home_change, caption="Change in time spent at home in weekends and weekdays - median [IQR]")`

`r kable(print(CreateTableOne("value", data=onlyhomehourswd, strata="timeframe", testNonNormal = kruskal.test, smd=F), nonnormal="value"), caption="Hours at home on weekdays - median [IQR]") `

`r kable(print(CreateTableOne("value", data=onlyhomehourswe, strata="timeframe", testNonNormal = kruskal.test, smd=F), nonnormal="value"), caption="Hours at home on weekends - median [IQR]") `

# Correlations

### Continuous and ordinal variables 

```{r correlations, echo=FALSE, warning=FALSE}

# remove dataframes not needed
rm(list=ls(pattern="only"))
rm(contacts_db, number_of_contacts_long, 
   sum_contacts_before_during_wd,sum_contacts_before_during_we,
   sum_number_of_contacts_change_wd,sum_number_of_contacts_change_we,
   sum_time_spent_home,sum_time_spent_home_change,time_spent_home_long)
   
# use percent change for continuous variables
number_of_contacts_wd_change_corr<-number_of_contacts_change_wd %>%
  pivot_wider(id_cols="child_ID", names_from = "place", names_prefix="change_contact_no_wd_",values_from="change")

number_of_contacts_we_change_corr<-number_of_contacts_change_we %>%
  pivot_wider(id_cols="child_ID", names_from = "place", names_prefix="change_contact_no_we_",values_from="change")

cig_change_corr<-cig_change %>%
  pivot_wider(id_cols="child_ID", names_from = "name", names_prefix="change_cig_no_",values_from="change")

time_spent_home_change_corr<-time_spent_home_change %>%
  pivot_wider(id_cols="child_ID", names_from = "days", names_prefix="change_hours_hm_",values_from="change")

cleaning_change_corr<-cleaning_change %>%
  pivot_wider(id_cols="child_ID", names_from = "name", names_prefix="change_cleaning_timesweek_",values_from="change")

spareact_change_corr<-spareact_change %>%
  pivot_wider(id_cols="child_ID", names_from = "name", names_prefix="change_",values_from="change")

# str(df_num, list.len=ncol(df_num))

df_cor<- df_num %>%
  #create variable days from reopening 
   mutate(days_from_reopening=as.numeric(difftime(as.Date(submission_time), as.Date("2020-05-21"), units="days"))) %>%
  # replace levels with less levels
  mutate_at(c("free_time_now","free_time_before"), 
           .funs = function(t) revalue(x = t,
    replace = c(
    "She/he was doing things that involve little physical effort" = "little physical effort",
    "1-2 times per week she/he did physical things (e.g. played sports,running,swimming)" = "1-2 times per week",
    "3-4 times per week she/he did physical things (e.g. played sports,running,swimming)" = ">2 times per week",
    "5-6 times per week she/he did physical things (e.g. played sports,running,swimming)" = ">2 times per week",
    "7 times or more per week she/he did physical things (e.g. played sports,running,swimming)" = ">2 times per week"))) %>%
  mutate_at(c("breakfast_now","breakfast_before"),
           .funs = function(t) revalue(x = t,
            replace = c("Never"="<7 times per week",
                        "2-3 times per month"="<7 times per week",
                        "1 time per week"="<7 times per week",
                        "2-3 times per week"="<7 times per week",
                        "4-6 times per week"="<7 times per week",
                        "Every day"="7 times per week"
                        ))) %>%
  mutate_at(c("fast_food_now","fast_food_before"),
           .funs = function(t) revalue(x = t,
            replace = c("Never"="Never",
                        "2-3 times per month"="2-3 times per month",
                        "1 time per week"="1-7 times per week",
                        "2-3 times per week"="1-7 times per week",
                        "4-6 times per week"="1-7 times per week",
                        "Every day"="1-7 times per week"
                        ))) %>%
  mutate_at(c("meat_before","sugar_before","fruits_before","veggies_before",
              "meat_now","sugar_now","fruits_now","veggies_now"), 
            .funs = function(t) revalue(x = t,
    replace = c("Every day"= "4-7 times per week",
                "4-6 times per week" = "4-7 times per week",
                "2-3 times per week" = "<4 times per week",
                "1 time per week" = "<4 times per week",
                "2-3 times per month"="<4 times per week",
                "1 time per month" = "<4 times per week",
                "Never" = "<4 times per week"))) %>%
  mutate_at(c("chips_before","chips_now"), 
            .funs = function(t) revalue(x = t,
    replace = c("Every day"= "4-7 times per week",
                "4-6 times per week" = "4-7 times per week",
                "2-3 times per week" = "1-3 times per week",
                "1 time per week" = "1-3 times per week",
                "2-3 times per month"="<1 time per week",
                "1 time per month" = "<1 time per week",
                "Never" = "<1 time per week"))) %>%
  mutate_at(c("fish_before","fish_now"), 
            .funs = function(t) revalue(x = t,
    replace = c("Every day"= "2-7 times per week",
                "4-6 times per week" = "2-7 times per week",
                "2-3 times per week" = "2-7 times per week",
                "1 time per week" = "1 time per week",
                "2-3 times per month"="<1 time per week",
                "1 time per month" = "<1 time per week",
                "Never" = "<1 time per week"))) %>% 
 mutate_at(c("leggumes_before","leggumes_now"), 
            .funs = function(t) revalue(x = t,
    replace = c("Every day"= "2-7 times per week",
                "4-6 times per week" = "2-7 times per week",
                "2-3 times per week" = "2-7 times per week",
                "1 time per week" = "<2 times per week",
                "2-3 times per month"="<2 times per week",
                "1 time per month" = "<2 times per week",
                "Never" = "<2 times per week"))) %>% 
  mutate_at(c("break_now","break_before"), 
           .funs = function(t) revalue(x = t,
    replace = c(
    "Sat down (talking, reading, eating)" = "Sitting, standing, walking",
    "Stood around or walked around" = "Sitting, standing, walking",
    "Ran or played a little bit" = "Ran, played a little or quite a bit",
    "Ran around and played quite a bit" = "Ran, played a little or quite a bit"))) %>%
  mutate_at(c("vaccine_doses"), 
           .funs = function(t) revalue(x = t,
    replace = c(
    "No, vaccine not available" = "No",
    "No, we thought that more than one dose wasn't needed" = "No",
    "No, we neglected but they will be provided" = "No",
    "No, only one dose was needed" = "No"))) %>%
  mutate_at(c("tv_pc_before","tv_pc_now"), 
           .funs = function(t) revalue(x = t,
    replace = c(
    "1-3 hours per day" = "1-3 hours per day",
    "4-7 hours per day" = ">3 hours per day",
    "8-11 hours per day" = ">3 hours per day",
    "More than 11 hours per day" = ">3 hours per day"))) %>%
  mutate_at(c("hands_soap_before","hands_soap_now",
              "hands_anticeptic_before","hands_anticeptic_now"), 
           .funs = function(t) revalue(x = t,
    replace = c(
    "Never" = "<4 times per day",
    "Less than 1 time per day" = "<4 times per day",
    "1-3 times per day" = "<4 times per day",
    "4-7 times per day" = "4 and above times per day",
    "More than 7 times per day" = "4 and above times per day"))) %>%
  mutate_at(c("father_education","mother_education"), 
           .funs = function(t) revalue(x = t,
    replace = c(
    "Has not completed primary school" = "Noeduc_Prim_Middle_School",
    "Primary School" = "Noeduc_Prim_Middle_School",
    "Middle School (3 years)" = "Noeduc_Prim_Middle_School",
    "High School/Vocational High School (diploma)" = "High_Higher_Education",
    "Higher (after high school) non-tertiary Education" = "High_Higher_Education",
    "Higher Tertiary Education (non-Univesity)" = "High_Higher_Education",
    "University (Bachelor degree)" = "University",
    "University-Postgraduate (only Master's degree)" = "University",
    "PhD" = "University"))) %>%
  # relevel variables so that they start from lower frequency and increasing
  mutate_at(.vars = c("chips_before","chips_now"),
            .funs = list(~fct_relevel(., "<1 time per week",
                                      "1-3 times per week",
                                      "4-7 times per week")))  %>%
  mutate_at(.vars = c("fish_before","fish_now"),
            .funs = list(~fct_relevel(., "<1 time per week",
                                      "1 time per week",
                                      "2-7 times per week")))  %>%
  mutate_at(.vars = c("meat_before","meat_now","sugar_before","sugar_now",
                      "veggies_now","veggies_before","fruits_before","fruits_now"),
            .funs = list(~fct_relevel(., "<4 times per week",
                                      "4-7 times per week")))  %>%
  mutate_at(.vars = c("leggumes_before","leggumes_now"),
            .funs = list(~fct_relevel(., "<2 times per week",
                                      "2-7 times per week")))  %>%
  mutate_at(.vars = c("chronic_disease","vaccine",
                      "vaccine_doses","no_vaccination","consent",
                      "sickness_now","supplements_now",
                      "sickness_before","supplements_before"),
            .funs = list(~fct_relevel(., "No", "Yes")))  %>%
  mutate_at(.vars = c("breakfast_now","breakfast_before"),
            .funs = list(~fct_relevel(., 
                                      "<7 times per week", 
                                      "7 times per week")))  %>%
   mutate_at(.vars = c("fast_food_now","fast_food_before"),
            .funs = list(~fct_relevel(., "Never", 
                                      "2-3 times per month", 
                                      "1-7 times per week")))  %>%
  mutate_at(.vars = c("smoking_now","smoking_before"),
            .funs = list(~fct_relevel(., "No", "Yes, occasionally", 
                                      "Yes, every day")))  %>%
  mutate_at(.vars = c("net_friends_now","net_family_now",
              "net_friends_before","net_family_before"),
            .funs = list(~fct_relevel(., "Never", 
                                     "Some times in the month but not every week", 
                                     "Every week but not every day",
                                     "Every day"))) %>%
  left_join(., number_of_contacts_wd_change_corr, by="child_ID") %>%
  left_join(., number_of_contacts_we_change_corr, by="child_ID") %>%
  left_join(., time_spent_home_change_corr, by="child_ID") %>%
  left_join(., cig_change_corr, by="child_ID") %>%
  left_join(., cleaning_change_corr, by="child_ID") %>%
  left_join(., spareact_change_corr, by="child_ID") %>%
  ungroup()

# str(df_cor, list.len=ncol(df_cor))

# relevel raw variables
df_cor_1<- df_num %>%
  #create variable days from reopening 
   mutate(days_from_reopening=as.numeric(difftime(as.Date(submission_time), as.Date("2020-05-21"), units="days"))) %>%
 mutate_at(.vars = c("fruits_before","veggies_before","meat_before",
              "leggumes_before", "fish_before","sugar_before",
              "chips_before","fruits_now","veggies_now","meat_now",
              "leggumes_now","sugar_now","chips_now"),
            .funs = list(~fct_relevel(., "Never","1 time per month",
                                      "2-3 times per month","1 time per week",
                                      "2-3 times per week", "4-6 times per week",
                                      "Every day")))  %>%
   mutate_at(.vars = c("fish_now"),
            .funs = list(~fct_relevel(., "Never","1 time per month",
                                      "2-3 times per month","1 time per week",
                                      "2-3 times per week", "4-6 times per week")))  %>%
   mutate_at(c("vaccine_doses"), 
           .funs = function(t) revalue(x = t,
    replace = c(
    "No, vaccine not available" = "No",
    "No, we thought that more than one dose wasn't needed" = "No",
    "No, we neglected but they will be provided" = "No",
    "No, only one dose was needed" = "No"))) %>%
  mutate_at(.vars = c("chronic_disease","vaccine","vaccine_doses",
                      "no_vaccination","consent",
                      "sickness_now","supplements_now",
                      "sickness_before","supplements_before"),
            .funs = list(~fct_relevel(., "No", "Yes")))  %>%
  mutate_at(.vars = c("breakfast_now","breakfast_before",
                      "fast_food_now","fast_food_before"),
            .funs = list(~fct_relevel(., "Never","2-3 times per month", 
                                      "1 time per week", 
                                      "2-3 times per week",
                                      "4-6 times per week",
                                      "Every day")))  %>%
  mutate_at(.vars = c("smoking_now","smoking_before"),
            .funs = list(~fct_relevel(., "No", "Yes, occasionally", 
                                      "Yes, every day")))  %>%
  mutate_at(.vars = c("net_friends_now","net_family_now",
              "net_friends_before","net_family_before"),
            .funs = list(~fct_relevel(., "Never", 
                                     "Some times in the month but not every week", 
                                     "Every week but not every day",
                                     "Every day"))) %>%
  left_join(., number_of_contacts_wd_change_corr, by="child_ID") %>%
  left_join(., number_of_contacts_we_change_corr, by="child_ID") %>%
  left_join(., time_spent_home_change_corr, by="child_ID") %>%
  left_join(., cig_change_corr, by="child_ID") %>%
  left_join(., cleaning_change_corr, by="child_ID") %>%
  left_join(., spareact_change_corr, by="child_ID") %>%
  ungroup()

# prepare dataset with raw variables that will be included in correlations
df_corr_1 <- df_cor_1 %>%
  dplyr::select(BMI_sds, days_from_reopening,
                change_cleaning_timesweek_total,
                change_cig_no_cigsperday,
                change_spareactivity,
                starts_with("change_contact"),
                starts_with("change_hours"),
                all_of(sel_phys_2),
                all_of(sel_diet),all_of(sel_diet_2),
                all_of(sel_demog),
                -BMI_for_age, -contains("age_groups"),
                all_of(sel_comm_screen),
                all_of(sel_hygiene)) %>%
  mutate_if(is.factor, .funs=~as.numeric(.)) %>%
  mutate_if(is.numeric, list(~na_if(., Inf))) %>% 
  select(-sibling, -test_covid, -place_of_birth, -vaccine, -city, -consent, -years_living_in_cyprus, -sibling)

# check correlation with raw variables
cors_1<-df_corr_1 %>% 
  correlate(use = "pairwise.complete.obs", method = "spearman", diagonal = 1) %>%    # Create correlation data frame (cor_df)
  #column_to_rownames(var = "rowname") %>% 
  shave(upper=TRUE) %>% 
  pivot_longer(cols=-rowname, names_to="to") %>%
  filter(!is.na(value)) %>% 
  filter(value!=1) %>% 
  dplyr::rename(from="rowname") %>% 
  mutate(type=case_when(value<0~"negative",
                        value>0~"positive"),
         weight=abs(value*100)) %>% 
  filter(abs(value)>=(0.7)) 

# graph
graph_1 <- graph_from_data_frame(cors_1)
pal <- wesanderson::wes_palette("Zissou1", n = 5, type = "discrete")

ggraph(graph_1, layout = 'linear', circular=TRUE) + 
  geom_edge_arc(aes(color = type, width=weight)) + 
  geom_node_text(aes(label = name),family="serif")+
  scale_edge_color_manual(values = c(pal[4], pal[1])) +
  theme_void() + 
  theme(legend.position = 'none') 

# partial correlation
corpartial_1 <- partial.r(df_corr_1, x=c(1:40, 43:46,49:59), y = c(41,42,47,48), 
                        use="pairwise", method="spearman")

corpartial_1[upper.tri(corpartial_1)] <- NA

cors_p_1 <- as.data.frame(apply(corpartial_1, 2, function(x) ifelse (!is.na(x),x,NA))) %>% 
  rownames_to_column(var="rowname") %>% 
    pivot_longer(cols=-rowname, names_to="to") %>%
  filter(!is.na(value)) %>% 
  filter(value!=1) %>% 
  dplyr::rename(from="rowname") %>% 
  mutate(type=case_when(value<0~"negative",
                        value>0~"positive"),
         weight=abs(value*100)) %>% 
  filter(abs(value)>=(0.7)) 

graph_p_1 <- graph_from_data_frame(cors_p_1)

ggraph(graph_p_1, layout = 'linear', circular=TRUE) + 
  geom_edge_arc(aes(color = type, width=weight)) + 
  geom_node_text(aes(label = name),family="serif")+
  geom_node_point()+
  scale_edge_color_manual(values = c(pal[2], pal[1])) +
  theme_void() + 
  theme(legend.position = 'none') 

```

Correlations - > abs(value)>=(0.7)
`r DT::datatable(cors_1)`

Partial Correlations - > abs(value)>=(0.7) (adjusted for age, sex, mother's and father's educational level)
`r DT::datatable(cors_p_1)`

# Compliance binary (1=increase in hours at home for weekdays) - regression analysis

## Multivariate - adj. age, sex and parents' educational level

```{r compliance_hours, echo=FALSE}
rm(number_of_contacts_change_wd,number_of_contacts_change_we,
   cig_change,time_spent_home_change,cleaning_change,spareact_change, cig_change_corr,cleaning_change_corr,number_of_contacts_wd_change_corr,
   number_of_contacts_we_change_corr,time_spent_home_change_corr,
   spareact_change_corr)
rm(list=ls(pattern="_1"))

df_reg_all_wd <- df_cor %>%
  mutate(compliance=as.factor(case_when(
        home_hours_wd_before < home_hours_wd ~ 1,
        home_hours_wd_before >= home_hours_wd ~ 0,
        TRUE~NA_real_))) %>%
dplyr::select(child_ID, compliance, BMI_sds, days_from_reopening,
                change_cleaning_timesweek_total,
                change_cig_no_cigsperday,
                change_spareactivity,
                starts_with("change_contact"),
                starts_with("change_hours"), -change_hours_hm_wd,
                all_of(sel_phys_2),-sickness_now, -sickness_before,
                all_of(sel_diet), all_of(sel_diet_2),
                all_of(sel_comm_screen),
                all_of(sel_hygiene),
                all_of(sel_demog), 
                -BMI_for_age, -contains("age_groups"),
                -sibling, -test_covid, -place_of_birth, 
                -vaccine, -years_living_in_cyprus) %>%
  # rename variables so that reference group is the one with the largest freq
  mutate_at(.vars = c("break_now","break_before"), 
            .funs = function(t) revalue(x = t,
            replace = c("Ran and played hard most of the time"="zRan and played hard most of the time"))) %>%
  mutate_at(.vars = c("fruits_before","fruits_now","sugar_before","sugar_now"), 
            .funs = function(t) revalue(x = t,
            replace = c("<4 times per week"="z<4 times per week"))) %>%
    mutate_at(.vars = c("leggumes_before","leggumes_now"), 
            .funs = function(t) revalue(x = t,
            replace = c("<2 times per week"="z<2 times per week"))) %>%
    mutate_at(.vars = c("fish_before","fish_now","chips_before","chips_now"), 
            .funs = function(t) revalue(x = t,
            replace = c("<1 time per week"="z<1 time per week"))) %>%  mutate_at(.vars = c("breakfast_now","breakfast_before"), 
            .funs = function(t) revalue(x = t,
            replace = c("<7 times per week"="z<7 times per week"))) %>%
mutate_at(.vars = c("tv_pc_before","tv_pc_now"), 
            .funs = function(t) revalue(x = t,
            replace = c(">3 hours per day"="z>3 hours per day"))) %>%   mutate_at(.vars = c("city"), 
            .funs = function(t) revalue(x = t,
            replace = c("Famagusta"="zFamagusta",
                        "Larnaca"="zLarnaca",
                        "Limassol"="zLimassol"))) %>%
  mutate_at(.vars = c("vaccine_doses"), 
            .funs = function(t) revalue(x = t,
            replace = c("No"="zNo"))) %>%
  mutate_at(.vars = c("father_education","mother_education"), 
            .funs = function(t) revalue(x = t,
            replace = c("Noeduc_Prim_Middle_School"="zNoeduc_Prim_Middle_School",
                        "High_Higher_Education"="zHigh_Higher_Education")))

# str(df_reg_all_wd, list.len=ncol(df_reg_all_wd))

multi_het_corr_db_mod_COMPLIANCE_CONT_wd <- df_reg_all_wd %>% 
  select(child_ID, compliance, num_child,
         days_from_reopening, BMI_sds, 
         age, sex, mother_education, father_education,
         change_cleaning_timesweek_total,
         change_cig_no_cigsperday,
         change_spareactivity,
         starts_with("change_contact"))

multi_compliance_CONT_wd <- reshape2::melt(multi_het_corr_db_mod_COMPLIANCE_CONT_wd,
id.vars = c("child_ID", "compliance", "age", "sex", 
            "mother_education","father_education")) 

multi_compliance_CONT_RES_wd <- multi_compliance_CONT_wd %>%
  filter(!is.infinite(value)) %>% 
  filter(!is.nan(value)) %>% 
  filter(!is.na(value)) %>%
  filter(!is.na(variable)) %>% 
  group_by(variable) %>%
  do(tidy(glm(compliance ~ age+sex+mother_education+father_education+value, 
              data = ., family = binomial(link = "logit")),
    conf.int = TRUE, conf.level = 0.95, exponentiate = TRUE))

multi_het_corr_db_mod_COMPLIANCE_CAT_wd <- df_reg_all_wd %>% 
  select(ends_with("before"),
         ends_with("now"),
         age, sex, city, chronic_disease,
         vaccine_doses, no_vaccination, attend_school, consent,
         mother_education, father_education, compliance, child_ID)

multi_compliance_CAT_wd <- reshape2::melt(multi_het_corr_db_mod_COMPLIANCE_CAT_wd,
id.vars = c("child_ID", "compliance", "age", "sex",
            "mother_education","father_education")) 

multi_compliance_CAT_RES_wd <- multi_compliance_CAT_wd %>%
  filter(!is.infinite(value)) %>% 
  filter(!is.nan(value)) %>% 
  filter(!is.na(value)) %>%
  filter(!is.na(variable)) %>% 
  group_by(variable) %>%
  do(tidy(glm(compliance ~ age + sex + mother_education + father_education + value, data = ., family = binomial(link = "logit")),
    conf.int = TRUE, conf.level = 0.95, exponentiate = TRUE))

multi_compliance_RES_wd <- rbindlist(l=list(multi_compliance_CONT_RES_wd,
                                   multi_compliance_CAT_RES_wd), 
                                  use.names = TRUE, fill = TRUE)


multi_compliance_RES_final_wd<-multi_compliance_RES_wd %>%
  as.data.frame() %>%
  filter(term != "(Intercept)") %>%
  filter(term != "age") %>%
  filter(term != "sexFemale") %>%
  filter(term != "mother_educationzHigh_Higher_Education") %>%
  filter(term != "mother_educationUniversity") %>%
  filter(term != "father_educationzHigh_Higher_Education") %>%
  filter(term != "father_educationUniversity") %>%
  mutate_if(.predicate = is.numeric, .funs = ~round(., 3))

multi_compliance_RES_final_wd <- multi_compliance_RES_final_wd %>%
  mutate(Outcome="Increase in hours staying at home - weekdays")

model1_multivariate<-multi_compliance_RES_final_wd

```


`r kable(print(CreateTableOne("compliance", data=df_reg_all_wd, factorVars="compliance")), caption="N % of compliance - increased number of hours at home")`

`r kable(print(CreateTableOne("compliance", data=df_reg_all_wd, factorVars="compliance", strata="sex")), caption="N % of compliance - increased number of hours at home")`

`r DT::datatable(multi_compliance_RES_final_wd)`

# Compliance binary (1=decrease in the number of vulnerable contacts at home during weekdays - regression analysis

## Multivariate - adj. age, sex and parents' educational level

```{r compliance_vuln, echo=FALSE}

rm(df_reg_all_wd)
rm(list=ls(pattern="^multi"))

df_reg_conv <- df_cor %>%
  mutate(compliance=as.factor(case_when(
    vulnerable_wd_now < vulnerable_weekdays_before ~ 1,
    vulnerable_wd_now >= vulnerable_weekdays_before ~ 0,
    TRUE~NA_real_))) %>%
  filter(!(vulnerable_wd_now == 0 & vulnerable_weekdays_before == 0)) %>%
dplyr::select(child_ID, compliance, BMI_sds, days_from_reopening,
                change_cleaning_timesweek_total,
                change_cig_no_cigsperday,
                change_spareactivity,
                starts_with("change_contact"), -change_contact_no_wd_vuln_home,
                starts_with("change_hours"),
                all_of(sel_phys_2),-sickness_now, -sickness_before,
                all_of(sel_diet), all_of(sel_diet_2),
                all_of(sel_comm_screen),
                all_of(sel_hygiene),
                all_of(sel_demog),
                -BMI_for_age, -contains("age_groups"),
                -sibling, -test_covid, -place_of_birth, 
                -vaccine, -years_living_in_cyprus) %>%
# rename variables so that reference group is the one with the largest freq
  mutate_at(.vars = c("break_now","break_before"), 
            .funs = function(t) revalue(x = t,
            replace = c("Ran and played hard most of the time"="zRan and played hard most of the time"))) %>%
  mutate_at(.vars = c("fruits_before","fruits_now","sugar_before","sugar_now"), 
            .funs = function(t) revalue(x = t,
            replace = c("<4 times per week"="z<4 times per week"))) %>%
    mutate_at(.vars = c("leggumes_before","leggumes_now"), 
            .funs = function(t) revalue(x = t,
            replace = c("<2 times per week"="z<2 times per week"))) %>%
    mutate_at(.vars = c("fish_before","fish_now","chips_before","chips_now"), 
            .funs = function(t) revalue(x = t,
            replace = c("<1 time per week"="z<1 time per week"))) %>%  mutate_at(.vars = c("breakfast_now","breakfast_before"), 
            .funs = function(t) revalue(x = t,
            replace = c("<7 times per week"="z<7 times per week"))) %>%
mutate_at(.vars = c("tv_pc_before","tv_pc_now"), 
            .funs = function(t) revalue(x = t,
            replace = c(">3 hours per day"="z>3 hours per day"))) %>%   mutate_at(.vars = c("city"), 
            .funs = function(t) revalue(x = t,
            replace = c("Famagusta"="zFamagusta",
                        "Larnaca"="zLarnaca",
                        "Limassol"="zLimassol"))) %>%
  mutate_at(.vars = c("vaccine_doses"), 
            .funs = function(t) revalue(x = t,
            replace = c("No"="zNo"))) %>%
  mutate_at(.vars = c("father_education","mother_education"), 
            .funs = function(t) revalue(x = t,
            replace = c("Noeduc_Prim_Middle_School"="zNoeduc_Prim_Middle_School",
                        "High_Higher_Education"="zHigh_Higher_Education")))

# str(df_reg_conv, list.len=ncol(df_reg_conv))

multi_het_corr_db_mod_COMPLIANCE_CONT_conv <- df_reg_conv %>% 
  select(child_ID, compliance, num_child,
         days_from_reopening, BMI_sds, 
         age, sex, mother_education, father_education,
         change_cleaning_timesweek_total,
         change_cig_no_cigsperday,
         change_spareactivity,
         starts_with("change_contact"),
         starts_with("change_hours"))

multi_compliance_CONT_conv <- reshape2::melt(multi_het_corr_db_mod_COMPLIANCE_CONT_conv,
id.vars = c("child_ID", "compliance", "age", "sex", "mother_education", "father_education")) 

multi_compliance_CONT_RES_conv <- multi_compliance_CONT_conv %>%
  filter(!is.infinite(value)) %>% 
  filter(!is.nan(value)) %>% 
  filter(!is.na(value)) %>%
  filter(!is.na(variable)) %>% 
  group_by(variable) %>%
  do(tidy(glm(compliance ~ age+sex+mother_education+father_education+value, 
              data = ., 
              family = binomial(link = "logit")),
    conf.int = TRUE, conf.level = 0.95, exponentiate = TRUE))

multi_het_corr_db_mod_COMPLIANCE_CAT_conv <- df_reg_conv %>% 
  select(ends_with("before"),
         ends_with("now"),
         age, sex, mother_education, father_education,
         compliance, city, chronic_disease,
         vaccine_doses, no_vaccination, attend_school, consent,
         mother_education, father_education, child_ID)

multi_compliance_CAT_conv <- reshape2::melt(multi_het_corr_db_mod_COMPLIANCE_CAT_conv,
id.vars = c("child_ID", "compliance", "age", "sex", "mother_education", "father_education")) 

multi_compliance_CAT_RES_conv <- multi_compliance_CAT_conv %>%
  filter(!is.infinite(value)) %>% 
  filter(!is.nan(value)) %>% 
  filter(!is.na(value)) %>%
  filter(!is.na(variable)) %>% 
  group_by(variable) %>%
 do(tidy(glm(compliance ~ age+sex+mother_education+father_education+value, 
             data = ., 
              family = binomial(link = "logit")),
    conf.int = TRUE, conf.level = 0.95, exponentiate = TRUE))


multi_compliance_RES_conv <- rbindlist(l=list(multi_compliance_CONT_RES_conv,
                                   multi_compliance_CAT_RES_conv), 
                                   use.names = TRUE, fill = TRUE)


multi_compliance_RES_final_conv<-multi_compliance_RES_conv %>%
  as.data.frame() %>%
  filter(term != "(Intercept)") %>%
  filter(term != "age") %>%
  filter(term != "sexFemale") %>%
  filter(term != "mother_educationzHigh_Higher_Education") %>%
  filter(term != "mother_educationUniversity") %>%
  filter(term != "father_educationzHigh_Higher_Education") %>%
  filter(term != "father_educationUniversity") %>%
  mutate_if(.predicate = is.numeric, .funs = ~round(., 3))


multi_compliance_RES_final_conv <- multi_compliance_RES_final_conv %>%
  mutate(Outcome="Decrease of vulnerable contacts at home - weekdays")

model2_multivariate<-multi_compliance_RES_final_conv

```

`r kable(print(CreateTableOne("compliance", data=df_reg_conv, factorVars="compliance")), caption="N % of compliance - decreased number of vulnerables contacts at home - weekdays")`

`r kable(print(CreateTableOne("compliance", data=df_reg_conv, factorVars="compliance", strata="sex")), caption="N % of compliance - decreased number of vulnerables contacts at home - weekdays")`

`r DT::datatable(multi_compliance_RES_final_conv)`


# Compliance (1=decrease in the number of contacts at school) - regression analysis

## Multivariate - adj. age, sex and parents' educational level

```{r compliance_school, echo=FALSE, warning=FALSE}

rm(list=ls(pattern="^multi"))
rm(df_reg_conv)

df_reg_cons <- df_cor %>%
  filter(attend_school=="Goes to school") %>%
  mutate(compliance=as.factor(case_when(
    school_persons_now < school_persons_before ~ 1,
    school_persons_now >= school_persons_before ~ 0,
    TRUE~NA_real_))) %>%
dplyr::select(child_ID, compliance, BMI_sds, days_from_reopening,
                change_cleaning_timesweek_total,
                change_cig_no_cigsperday,
                change_spareactivity,
                starts_with("change_contact"),
                -change_contact_no_wd_school,
                starts_with("change_hours"),
                all_of(sel_phys_2),-sickness_now, -sickness_before,
                all_of(sel_diet), all_of(sel_diet_2),
                all_of(sel_comm_screen),
                all_of(sel_hygiene),
                all_of(sel_demog), -attend_school,-consent,
                -BMI_for_age, -contains("age_groups"),
                -sibling, -test_covid, -place_of_birth, 
                -vaccine, -years_living_in_cyprus) %>%
# rename variables so that reference group is the one with the largest freq
  mutate_at(.vars = c("break_now","break_before"), 
            .funs = function(t) revalue(x = t,
            replace = c("Ran and played hard most of the time"="zRan and played hard most of the time"))) %>%
  mutate_at(.vars = c("fruits_before","fruits_now","sugar_before","sugar_now"), 
            .funs = function(t) revalue(x = t,
            replace = c("<4 times per week"="z<4 times per week"))) %>%
    mutate_at(.vars = c("leggumes_before","leggumes_now"), 
            .funs = function(t) revalue(x = t,
            replace = c("<2 times per week"="z<2 times per week"))) %>%
    mutate_at(.vars = c("fish_before","fish_now","chips_before","chips_now"), 
            .funs = function(t) revalue(x = t,
            replace = c("<1 time per week"="z<1 time per week"))) %>%  mutate_at(.vars = c("breakfast_now","breakfast_before"), 
            .funs = function(t) revalue(x = t,
            replace = c("<7 times per week"="z<7 times per week"))) %>%
mutate_at(.vars = c("tv_pc_before","tv_pc_now"), 
            .funs = function(t) revalue(x = t,
            replace = c(">3 hours per day"="z>3 hours per day"))) %>%   mutate_at(.vars = c("city"), 
            .funs = function(t) revalue(x = t,
            replace = c("Famagusta"="zFamagusta",
                        "Larnaca"="zLarnaca",
                        "Limassol"="zLimassol"))) %>%
  mutate_at(.vars = c("vaccine_doses"), 
            .funs = function(t) revalue(x = t,
            replace = c("No"="zNo"))) %>%
  mutate_at(.vars = c("father_education","mother_education"), 
            .funs = function(t) revalue(x = t,
            replace = c("Noeduc_Prim_Middle_School"="zNoeduc_Prim_Middle_School",
                        "High_Higher_Education"="zHigh_Higher_Education")))

# str(df_reg_cons, list.len=ncol(df_reg_cons))

multi_het_corr_db_mod_COMPLIANCE_CONT_cons <- df_reg_cons %>% 
  select(child_ID, compliance, num_child,
         days_from_reopening, BMI_sds, 
         age, sex, mother_education, father_education,
         change_cleaning_timesweek_total,
         change_cig_no_cigsperday,
         change_spareactivity,
         starts_with("change_contact"),
         starts_with("change_hours"))

multi_compliance_CONT_cons <- reshape2::melt(multi_het_corr_db_mod_COMPLIANCE_CONT_cons,
id.vars = c("child_ID", "compliance", "age", "sex","mother_education","father_education" )) 

multi_compliance_CONT_RES_cons <- multi_compliance_CONT_cons %>%
  filter(!is.infinite(value)) %>% 
  filter(!is.nan(value)) %>% 
  filter(!is.na(value)) %>%
  filter(!is.na(variable)) %>% 
  group_by(variable) %>%
  do(tidy(glm(compliance ~ age+sex+mother_education+father_education+value, 
              data = ., 
              family = binomial(link = "logit")),
    conf.int = TRUE, conf.level = 0.95, exponentiate = TRUE))

multi_het_corr_db_mod_COMPLIANCE_CAT_cons <- df_reg_cons %>% 
  select(ends_with("before"),
         ends_with("now"),
         age, sex, city, chronic_disease,
         vaccine_doses, no_vaccination,
         mother_education, father_education, compliance, child_ID)


multi_compliance_CAT_cons <- reshape2::melt(multi_het_corr_db_mod_COMPLIANCE_CAT_cons,
id.vars = c("child_ID", "compliance", "age", "sex", "mother_education","father_education")) 

multi_compliance_CAT_RES_cons <- multi_compliance_CAT_cons %>%
  filter(!is.infinite(value)) %>% 
  filter(!is.nan(value)) %>% 
  filter(!is.na(value)) %>%
  filter(!is.na(variable)) %>% 
  group_by(variable) %>%
  do(tidy(glm(compliance ~ age + sex + mother_education+father_education+value, data = ., 
              family = binomial(link = "logit")),
    conf.int = TRUE, conf.level = 0.95, exponentiate = TRUE))

multi_compliance_RES_cons <- rbindlist(l=list(multi_compliance_CONT_RES_cons,
                                   multi_compliance_CAT_RES_cons), 
                                  use.names = TRUE, fill = TRUE)


multi_compliance_RES_final_cons<-multi_compliance_RES_cons %>%
  as.data.frame() %>%
  filter(term != "(Intercept)") %>%
  filter(term != "age") %>%
  filter(term != "sexFemale") %>%
  filter(term != "mother_educationzHigh_Higher_Education") %>%
  filter(term != "mother_educationUniversity") %>%
  filter(term != "father_educationzHigh_Higher_Education") %>%
  filter(term != "father_educationUniversity") %>%
  mutate_if(.predicate = is.numeric, .funs = ~round(., 3))

multi_compliance_RES_final_cons <- multi_compliance_RES_final_cons %>%
  mutate(Outcome="Decrease of contacts at school")

model3_multivariate<-multi_compliance_RES_final_cons

```

`r kable(print(CreateTableOne("compliance", data=df_reg_cons, factorVars="compliance")), caption="N % of compliance - decreased number of contacts at school")`

`r kable(print(CreateTableOne("compliance", data=df_reg_cons, factorVars="compliance", strata="sex")), caption="N % of compliance - decreased number of contacts at school")`

`r DT::datatable(multi_compliance_RES_final_cons)`

# Compliance binary (1=increase in frequency of hand washing with soap or antiseptic use) - regression analysis


## Multivariate - adj. age, sex and parents' educational level

```{r compliance_handwashing, echo=FALSE}

rm(list=ls(pattern="^multi"))
rm(df_reg_cons)

# create new variable for hand washing compliance (soap or antiseptic use)
df_num$compliance_hand<-as.factor(
   ifelse(
  (df_num$hands_soap_now=="1-3 times per day" & 
     df_num$hands_soap_before=="Less than 1 time per day") | 
    (df_num$hands_anticeptic_now=="1-3 times per day" & 
     df_num$hands_anticeptic_before=="Less than 1 time per day") , 
  1, 
  ifelse(
  (df_num$hands_soap_now=="4-7 times per day" & 
     df_num$hands_soap_before=="1-3 times per day") |
    (df_num$hands_anticeptic_now=="4-7 times per day" & 
     df_num$hands_anticeptic_before=="1-3 times per day"), 
  1,  
  ifelse(
  (df_num$hands_soap_now=="4-7 times per day" & 
     df_num$hands_soap_before=="Less than 1 time per day") |
    (df_num$hands_anticeptic_now=="4-7 times per day" & 
     df_num$hands_anticeptic_before=="Less than 1 time per day"), 
  1,
  ifelse(
  (df_num$hands_soap_now=="More than 7 times per day" & 
     df_num$hands_soap_before=="4-7 times per day") |
    (df_num$hands_anticeptic_now=="More than 7 times per day" & 
     df_num$hands_anticeptic_before=="4-7 times per day"), 
  1,  
  ifelse(
  (df_num$hands_soap_now=="More than 7 times per day" & 
     df_num$hands_soap_before=="1-3 times per day") |
    (df_num$hands_anticeptic_now=="More than 7 times per day" & 
     df_num$hands_anticeptic_before=="1-3 times per day"), 
  1, 
  ifelse(
  (df_num$hands_soap_now=="More than 7 times per day" & 
     df_num$hands_soap_before=="Less than 1 time per day") |
    (df_num$hands_anticeptic_now=="More than 7 times per day" & 
     df_num$hands_anticeptic_before=="Less than 1 time per day"), 
  1,
  ifelse(
  (df_num$hands_soap_now=="More than 7 times per day" & 
     df_num$hands_soap_before=="More than 7 times per day") |
    (df_num$hands_anticeptic_now=="More than 7 times per day" & 
     df_num$hands_anticeptic_before=="More than 7 times per day"), 
  1,
  0))))))))

#check <- df_num %>%
#  select(hands_soap_now, hands_soap_before, 
#hands_anticeptic_now, hands_anticeptic_before, compliance_hand)

df_reg_hand <- df_cor %>%
dplyr::select(child_ID, BMI_sds, days_from_reopening,
                change_cleaning_timesweek_total,
                change_cig_no_cigsperday,
                change_spareactivity,
                starts_with("change_contact"),
                starts_with("change_hours"),
                all_of(sel_phys_2),-sickness_now, -sickness_before,
                all_of(sel_diet), all_of(sel_diet_2),
                all_of(sel_comm_screen),
                all_of(sel_hygiene), 
                all_of(sel_demog),
                -BMI_for_age, -contains("age_groups"),
                -sibling, -test_covid, -place_of_birth, 
                -vaccine, -years_living_in_cyprus) %>%
  left_join(., df_num[, c("child_ID","compliance_hand")], by="child_ID") %>%
# rename variables so that reference group is the one with the largest freq
  mutate_at(.vars = c("break_now","break_before"), 
            .funs = function(t) revalue(x = t,
            replace = c("Ran and played hard most of the time"="zRan and played hard most of the time"))) %>%
  mutate_at(.vars = c("fruits_before","fruits_now","sugar_before","sugar_now"), 
            .funs = function(t) revalue(x = t,
            replace = c("<4 times per week"="z<4 times per week"))) %>%
    mutate_at(.vars = c("leggumes_before","leggumes_now"), 
            .funs = function(t) revalue(x = t,
            replace = c("<2 times per week"="z<2 times per week"))) %>%
    mutate_at(.vars = c("fish_before","fish_now","chips_before","chips_now"), 
            .funs = function(t) revalue(x = t,
            replace = c("<1 time per week"="z<1 time per week"))) %>%  mutate_at(.vars = c("breakfast_now","breakfast_before"), 
            .funs = function(t) revalue(x = t,
            replace = c("<7 times per week"="z<7 times per week"))) %>%
mutate_at(.vars = c("tv_pc_before","tv_pc_now"), 
            .funs = function(t) revalue(x = t,
            replace = c(">3 hours per day"="z>3 hours per day"))) %>%   mutate_at(.vars = c("city"), 
            .funs = function(t) revalue(x = t,
            replace = c("Famagusta"="zFamagusta",
                        "Larnaca"="zLarnaca",
                        "Limassol"="zLimassol"))) %>%
  mutate_at(.vars = c("vaccine_doses"), 
            .funs = function(t) revalue(x = t,
            replace = c("No"="zNo"))) %>%
  mutate_at(.vars = c("father_education","mother_education"), 
            .funs = function(t) revalue(x = t,
            replace = c("Noeduc_Prim_Middle_School"="zNoeduc_Prim_Middle_School",
                        "High_Higher_Education"="zHigh_Higher_Education")))
  
# str(df_reg_hand, list.len=ncol(df_reg_hand))

multi_het_corr_db_mod_COMPLIANCE_CONT_hw <- df_reg_hand %>% 
  select(child_ID, compliance_hand, num_child,
         days_from_reopening, BMI_sds, 
         age, sex, mother_education, father_education,
         change_cleaning_timesweek_total,
         change_cig_no_cigsperday,
         change_spareactivity,
         starts_with("change_contact"),
         starts_with("change_hours"))

multi_compliance_CONT_hw <- reshape2::melt(multi_het_corr_db_mod_COMPLIANCE_CONT_hw,
id.vars = c("child_ID", "compliance_hand", "age", "sex","mother_education", "father_education")) 

multi_compliance_CONT_RES_hw <- multi_compliance_CONT_hw %>%
  filter(!is.infinite(value)) %>% 
  filter(!is.nan(value)) %>% 
  filter(!is.na(value)) %>%
  filter(!is.na(variable)) %>% 
  group_by(variable) %>%
  do(tidy(glm(compliance_hand ~ age+sex+mother_education+father_education+value, 
              data = ., 
              family = binomial(link = "logit")),
    conf.int = TRUE, conf.level = 0.95, exponentiate = TRUE))

multi_het_corr_db_mod_COMPLIANCE_CAT_hw <- df_reg_hand %>% 
  select(ends_with("before"),-hands_soap_before,-hands_anticeptic_before,
         ends_with("now"),-hands_soap_now,-hands_anticeptic_now,
         age, sex, city, chronic_disease,
         vaccine_doses, no_vaccination, attend_school, consent,
         mother_education, father_education, compliance_hand, child_ID)


multi_compliance_CAT_hw <- reshape2::melt(multi_het_corr_db_mod_COMPLIANCE_CAT_hw,
id.vars = c("child_ID", "compliance_hand", "age", "sex", "mother_education", "father_education")) 

multi_compliance_CAT_RES_hw <- multi_compliance_CAT_hw %>%
  filter(!is.infinite(value)) %>% 
  filter(!is.nan(value)) %>% 
  filter(!is.na(value)) %>%
  filter(!is.na(variable)) %>% 
  group_by(variable) %>%
  do(tidy(glm(compliance_hand ~ age + sex + mother_education + father_education + value, data = ., 
              family = binomial(link = "logit")),
    conf.int = TRUE, conf.level = 0.95, exponentiate = TRUE))

multi_compliance_RES_hw <- rbindlist(l=list(multi_compliance_CONT_RES_hw,
                                   multi_compliance_CAT_RES_hw), 
                                  use.names = TRUE, fill = TRUE)


multi_compliance_RES_final_hw<-multi_compliance_RES_hw %>%
  as.data.frame() %>%
  filter(term != "(Intercept)") %>%
  filter(term != "age") %>%
  filter(term != "sexFemale") %>%
  filter(term != "mother_educationzHigh_Higher_Education") %>%
  filter(term != "mother_educationUniversity") %>%
  filter(term != "father_educationzHigh_Higher_Education") %>%
  filter(term != "father_educationUniversity") %>%
  mutate_if(.predicate = is.numeric, .funs = ~round(., 3))


multi_compliance_RES_final_hw <- multi_compliance_RES_final_hw %>%
  mutate(Outcome="Increase in hand hygiene - soap or antiseptic")


model4_multivariate<-multi_compliance_RES_final_hw

```

`r kable(print(CreateTableOne("compliance_hand", data=df_reg_hand, factorVars="compliance_hand")), caption="N % of compliance - increase in hand washing with soap or antiseptic use")`

`r kable(print(CreateTableOne("compliance_hand", data=df_reg_hand, factorVars="compliance_hand", strata="sex")), caption="N % of compliance - increase in hand washing with soap or antiseptic use")`

`r DT::datatable(multi_compliance_RES_final_hw)`

# FDR for all models

```{r echo=FALSE}
rm(list=ls(pattern="^multi"))
rm(df_reg_hand)
all_models_multi<-bind_rows(model1_multivariate, model2_multivariate, 
                            model3_multivariate,  model4_multivariate)

all_models_multi$FDR_overall <-p.adjust(all_models_multi$p.value,
    method = "fdr")  
library(RColorBrewer)

ggplot(all_models_multi, aes(x=variable, y=-log10(FDR_overall))) +
  geom_point(aes(color=Outcome), size = 1.25) +
  geom_hline(yintercept = -log10(0.05), color = "black", 
             linetype = "dashed", size=1) +
  facet_wrap(~Outcome, nrow = 4) +
  #  geom_text(hjust=0.1,vjust=0.2, size = 1) +
 # coord_flip()+
  scale_color_brewer(palette = "Dark2")+
  #geom_errorbar(aes(x= estimate, y=sig, ymin=conf.low, ymax=conf.high))+
  theme_minimal() +
  theme(
    legend.position = "bottom",
    panel.border = element_blank(),
    panel.grid.major.x = element_blank(),
    panel.grid.minor.x = element_blank(),
    strip.background = element_blank(),
  strip.text.x = element_blank(), axis.text.x = element_blank())+
  labs(x = NULL, y = "-log10(FDR)") +guides(color=guide_legend(nrow=2,byrow=TRUE))


# prepare for plot
all_models_multi <- all_models_multi %>%
  mutate(FDRimp_overall = ifelse(
  (FDR_overall == 0), 
  0.000001,
  FDR_overall)) %>%
  mutate(sig_overall=-log10(FDRimp_overall)) %>%
  mutate(FDRlevels_overall=cut(FDR_overall,
                       breaks=c(-Inf, 0.05,  Inf),
                       labels=c("<0.05",">0.05"))) 

# manhatan plot with threshold for fdr < 0.05 and odds ratio - all compliance models
all_models_multi  %>%
ggplot(aes(x=estimate, y=sig_overall)) +
  geom_point(aes(color=FDRlevels_overall), alpha = 1, size = 1.25) +
  geom_hline(yintercept = -log10(0.05), color = "black", 
             linetype = "dashed", size=1) +
  geom_vline(xintercept = 1, color = "black", 
             linetype = "dashed", size=1) +
    facet_wrap(~Outcome, nrow = 2, ncol =2) +
    scale_color_brewer(palette = "Dark2")+
  #  geom_text(hjust=0.1,vjust=0.2, size = 1) +
 # coord_flip()+
  #scale_color_manual(values = c("skyblue3", "deeppink4"))+
  labs(x = "Odds Ratio", y = "-log10(FDR)") +
  #geom_errorbar(aes(x= estimate, y=sig, ymin=conf.low, ymax=conf.high))+
  theme_minimal() +
  theme(
    legend.position = "top",
    panel.border = element_blank(),
    panel.grid.major.x = element_blank(),
    panel.grid.minor.x = element_blank())

# manhatan plot with threshold for fdr < 0.05 and odds ratio - separately 
all_models_multi  %>%
  filter(Outcome=="Increase in hours staying at home - weekdays") %>%
ggplot(aes(x=estimate, y=sig_overall)) +
  geom_point(aes(color=FDRlevels_overall), alpha = 1, size = 1.25) +
  geom_hline(yintercept = -log10(0.05), color = "black", 
             linetype = "dashed", size=1) +
  geom_vline(xintercept = 1, color = "black", 
             linetype = "dashed", size=1) +
  labs(x = "Odds Ratio", y = "-log10(FDR)") +
  theme_minimal() +
  theme(
    legend.position = "top",
    panel.border = element_blank(),
    panel.grid.major.x = element_blank(),
    panel.grid.minor.x = element_blank())

all_models_multi  %>%
  filter(Outcome=="Decrease of vulnerable contacts at home - weekdays") %>%
ggplot(aes(x=estimate, y=sig_overall)) +
  geom_point(aes(color=FDRlevels_overall), alpha = 1, size = 1.25) +
  geom_hline(yintercept = -log10(0.05), color = "dark blue", 
             linetype = "dashed", size=1) +
  geom_vline(xintercept = 1, color = "dark blue", 
             linetype = "dashed", size=1) +
  labs(x = "Odds Ratio", y = "-log10(FDR)") +
  theme_minimal() +
  theme(
    legend.position = "top",
    panel.border = element_blank(),
    panel.grid.major.x = element_blank(),
    panel.grid.minor.x = element_blank())

all_models_multi  %>%
  filter(Outcome=="Decrease of contacts at school") %>%
ggplot(aes(x=estimate, y=sig_overall)) +
  geom_point(aes(color=FDRlevels_overall), alpha = 1, size = 1.25) +
  geom_hline(yintercept = -log10(0.05), color = "dark blue", 
             linetype = "dashed", size=1) +
  geom_vline(xintercept = 1, color = "dark blue", 
             linetype = "dashed", size=1) +
  labs(x = "Odds Ratio", y = "-log10(FDR)") +
  theme_minimal() +
  theme(
    legend.position = "top",
    panel.border = element_blank(),
    panel.grid.major.x = element_blank(),
    panel.grid.minor.x = element_blank())

all_models_multi  %>%
  filter(Outcome=="Increase in hand hygiene - soap or antiseptic") %>%
ggplot(aes(x=estimate, y=sig_overall)) +
  geom_point(aes(color=FDRlevels_overall), alpha = 1, size = 1.25) +
  geom_hline(yintercept = -log10(0.05), color = "dark blue", 
             linetype = "dashed", size=1) +
  geom_vline(xintercept = 1, color = "dark blue", 
             linetype = "dashed", size=1) +
  labs(x = "Odds Ratio", y = "-log10(FDR)") +
  theme_minimal() +
  theme(
    legend.position = "top",
    panel.border = element_blank(),
    panel.grid.major.x = element_blank(),
    panel.grid.minor.x = element_blank())
```

`r DT::datatable(all_models_multi)`

`r DT::datatable(filter(all_models_multi, FDR_overall<=0.05))`


# Session information

```{r session, echo=FALSE}
sessionInfo()
```
````
