## Supplementary material for "Exposome changes in primary school children following the wide population non-pharmacological interventions implemented due to COVID-19 in Cyprus: a national survey": Deidentified data and script:

### Exposome@School

Καλώς Ήρθατε!

Παρακαλούμε συμπληρώστε το πιο κάτω ερωτηματολόγιο για το παιδί σας.

---

Συμπληρωματικά στοιχεία για τις τρέχουσες διαγνωστικές εξετάσεις στα σχολεία

Η 2η φάση αντιμετώπισης της πανδημίας COVID-19 περιλαμβάνει τη σταδιακή χαλάρωση των περιοριστικών μέτρων, ένα εξ' αυτών και η επαναλειτουργία των σχολείων. Η κυβέρνηση διενεργεί ένα πρόγραμμα διαγνωστικών εξετάσεων για τη νόσο COVID-19 με δειγματοληψία στους μαθητές και το προσωπικό των σχολείων. Συμπληρωματικά με τις δειγματοληψίες, σας ζητούνται στοιχεία για τον προσδιορισμό αλλαγών σε συμπεριφορές/πρακτικές των παιδιών στο σχολείο και στο σπίτι που θα βοηθήσουν να διαχειριστούμε το ρίσκο μετάδοσης του ιού όσο το δυνατόν καλύτερα.

Με την έγκριση του Υπουργείου Παιδείας, Πολιτισμού, Αθλητισμού και Νεολαίας (Αρ.Φακ.:21.11.06.10) διενεργείται έρευνα από το Διεθνές Ινστιτούτο Κύπρου για την Περιβαλλοντική και Δημόσια Υγεία του Τεχνολογικού Πανεπιστημίου Κύπρου σε συνεργασία με το Τμήμα Διατροφής του Υπουργείου Υγείας, η οποία έχει εγκριθεί από την Εθνική Επιτροπή Βιοηθικής Κύπρου (ΕΕΒΚ ΕΠ 2020.01.113).

Ποιος είναι ο κύριος στόχος της έρευνας;

Οι στόχοι της έρευνας περιλαμβάνουν την αξιολόγηση των εκθέσεων και συμπεριφορών των παιδιών αλλά και των αλλαγών στον τρόπο ζωής τους στο σπίτι και το σχολείο, ενώ θα περιγραφούν και παράγοντες που μπορεί να σχετίζονται με την ασθένεια COVID-19.

Ποιο το όφελος συμμετοχής σας (του παιδιού σας) στην έρευνα;

Συνεισφέρετε στη συλλογή σημαντικών στοιχείων συμπληρωματικών των διαγνωστικών εξετάσεων για την καλύτερη εφαρμογή προγραμμάτων δημόσιας υγείας που μειώνουν το ρίσκο μετάδοσης ιού στα σχολεία, και κατ'επέκταση στον κυπριακό πληθυσμό.

Σημαντικές πληροφορίες

• Το ερωτηματολόγιο συμπληρώνεται από γονείς για το παιδί τους ή παιδιά τους που είτε έλαβε/αν μέρος είτε όχι στο πρόγραμμα διαγνωστικών ελέγχων για τον νέο κορωνοϊό στα σχολεία.

• Ο συνολικός χρόνος συμπλήρωσης του ερωτηματολογίου κυμαίνεται μεταξύ 15- 20 λεπτών.

• Η συμπλήρωση του ερωτηματολογίου είναι εθελοντική. Κατά τη διάρκεια του ερωτηματολογίου, μπορείτε να αποσύρετε την συμμετοχή σας κλείνοντας απλά το παράθυρο. Τα στοιχεία σας αποθηκεύονται μόνο εάν επιλέξετε την εντολή "SUBMIT" που εμφανίζεται στο τέλος του ερωτηματολογίου.

Πώς θα χρησιμοποιηθούν τα δεδομένα σας;

• Όλα τα δεδομένα που θα συλλεχθούν θα χρησιμοποιηθούν για στατιστική επεξεργασία μαζί με τα αποτελέσματα των άλλων συμμετεχόντων στη μελέτη, χωρίς να φαίνονται τα προσωπικά σας στοιχεία.

• Η ανωνυμία σας (των παιδιών σας) θα διασφαλιστεί καθ' όλη τη διεξαγωγή της έρευνας και κατά τη δημοσιοποίηση των αποτελεσμάτων σε συνέδρια, επιστημονικά περιοδικά, κτλ.

• Η πλήρης βάση δεδομένων και ο κώδικας σύνδεσης των δεδομένων θα φυλάγεται σε ασφαλές μέρος με πρόσβαση σε αυτά μόνο η ερευνητική ομάδα.

•Τα δεδομένα που θα αποκτηθούν από την μελέτη θα αποθηκευτούν σε ασφαλή χώρο από το Διεθνές Ινστιτούτο Κύπρου για την Περιβαλλοντική και Δημόσια Υγεία για 10 χρόνια και μετά θα καταστραφούν.

Η συμμετοχή σας στη μελέτη είναι σημαντική, γιατί θα βοηθήσει όλους μας να μάθουμε περισσότερα για τις επιδράσεις του τρόπου ζωής των παιδιών στο ρίσκο λοίμωξης τους από τον νέο κορωνοϊό με απώτερο στόχο την ακόμη αποτελεσματικότερη προστασία τους στο σχολικό και ευρύτερο περιβάλλον.

Περισσότερες πληροφορίες για την ερευνητική δραστηριότητα του Ινστιτούτου μπορείτε να βρείτε στην ιστοσελίδα: https://www.cut.ac.cy/cii/.

Έχω διαβάσει τις πιο πάνω πληροφορίες και:*

◯ Συμφωνώ να σας παράσχω πληροφορίες που αφορούν το παιδί μου για να χρησιμοποιηθούν ΑΝΩΝΥΜΑ ΚΑΙ ΕΜΠΙΣΤΕΥΤΙΚΑ στη συγκεκριμένη επιστημονική έρευνα.

Πόσα παιδιά έχετε που φοιτούν στο δημοτικό σχολείο;

__________________________________

Παρακαλώ συμπληρώστε το ερωτηματολόγιο παρέχοντας πληροφορίες που αφορούν μόνο στο ένα από τα παιδιά σας. Αν θέλετε να συμπληρώσετε ξανά το ερωτηματολόγιο και για το άλλο σας παιδί, ακολουθείστε τις οδηγίες που θα εμφανιστούν στην οθόνη σας, μετά που θα έχετε πατήσει 'SUBMIT' στο τέλος του ερωτηματολογίου.

Παρακαλώ συμπληρώστε το ερωτηματολόγιο παρέχοντας πληροφορίες που αφορούν μόνο στο ένα σας παιδί. Αν θέλετε να συμπληρώσετε ξανά το ερωτηματολόγιο και για τα άλλα σας παιδιά, ακολουθείστε τις οδηγίες που θα εμφανιστούν στην οθόνη σας, μετά που θα έχετε πατήσει 'SUBMIT' στο τέλος του ερωτηματολογίου.

#### Κατάσταση υγείας

1. Έχει διαγνωστεί το παιδί σας από γιατρό με κάποια χρόνια ασθένεια (π.χ. Άσθμα, αλλεργίες, διαβήτη, χρόνια βρογχίτιδα);

◯ Ναι ◯ Όχι ◯ Δεν γνωρίζω ◯ Δεν απαντώ

Παρακαλώ σημειώστε με ποια χρόνια ασθένεια έχει διαγνωστεί το παιδί σας:

__________________________________

Για να απαντήσετε τις πιο κάτω ερωτήσεις σχετικά με τους εμβολιασμούς μπορείτε να λάβετε υπόψη ότι στην Κύπρο το πρόγραμμα εμβολιασμών περιλαμβάνει/συστήνει τα παρακάτω εμβόλια για όλους:

• Διφθερίτιδα, τέτανος, κοκκύτης (Diphtheria, Tetanus, Pertussis; DTaP) (>1 δόσεις)
• Πολιομυελίτιδα (Polio) (>1 δόσεις)
• Αιμόφιλο ινφλουέντζας τύπου b (Haemophilus influenza type b) (>1 δόσεις)
• Πνευμονιόκοκκο (Pneumococcal conjugated) (>1 δόσεις)
• Ηπατίτιδα Β (Hepatitis B) (>1 δόσεις)
• Μηνιγγιτιδόκοκκο C (Meningococcal C conjugated) (1 δόση)
• Ιλαρά, παρωτίτιδα, ερυθρά (Measles, Mumps, Rubella; MMR)
• Ανεμοβλογιά (Varicela)
• Ιό ανθρώπινων θηλωμάτων (Human Papilomavirus, HPV)

Και σε ειδικές περιπτώσεις τα παρακάτω εμβόλια για:
• Πνευμονιόκοκκο πολυσακχαριδικό (Pneumococcal polysaccharide, PPV23)
• Μηνιγγιτιδόκοκκο πολυσακαριδικό (Meningococcal polysaccharide)
• Ηπατίτιδα Α (Hepatitis A)
• Φυματίωση (tuberculosis, BCG)
• Αντιγριπικό (εποχιακή γρίπη, influenza)

Για περισσότερες πληροφορίες παρακάλω επισκεφτείτε τους πιο κάτω συνδέσμους:

Immunization Schedule 2012
Reccomended vaccinations for Cyprus

2. Το παιδί σας έχει κάνει τουλάχιστον ένα από τα εμβόλια της πιο πάνω λίστας;

◯ Ναι ◯ Όχι ◯ Δεν γνωρίζω/δεν είμαι σίγουρος/δεν θυμάμαι ◯ Δεν απαντώ

2α. Για τα εμβόλια που έχει κάνει το παιδί σας και απαιτούνται πάνω από μια δόσεις, έχει κάνει όλες τις δόσεις;

◯ Ναι
◯ Όχι γιατί το εμβόλιο δεν ήταν διαθέσιμο
◯ Όχι γιατί θεωρήσαμε ότι δεν χρειαζεται
◯ Όχι γιατί το αμελήσαμε αλλά θα τις κάνει
◯ Δεν χρειαζόταν πάνω από μία δόσεις στο/στα εμβόλιο/εμβόλια που έκανε
◯ Δεν γνωρίζω
◯ Δεν απαντώ

2β. Υπάρχει κάποιο εμβόλιο που επιλέξατε να μην κάνει το παιδί σας;

◯ Ναι
◯ Όχι (έχει κάνει όλα τα εμβόλια που συστήνονται για την ηλικία του)
◯ Δεν γνωρίζω
◯ Δεν απαντώ

Παρακαλώ συμπληρώστε ποιο/ποια εμβόλια επιλέξατε να μην κάνει το παιδί σας:

__________________________________

#### Σωματική άσκηση ΑΥΤΗ ΤΗΝ ΠΕΡΙΟΔΟ, με τη σταδιακή άρση των περιοριστικών μέτρων

3. Έχει κάνει το παιδί σας κάποια/κάποιες από τις πιο κάτω δραστηριότητες την προηγούμενη εβδομάδα (7 ημέρες); Εάν ναι, επιλέξτε πόσες φορές την εβδομάδα.

Έντονο περπάτημα

◯ Όχι
◯ 1-2 φορές την εβδομάδα
◯ 3-4 φορές την εβδομάδα
◯ 5-6 φορές την εβδομάδα
◯ 7 ή περισσότερες φορές την εβδομάδα
◯ Δεν γνωρίζω
◯ Δεν απαντώ

Ποδήλατο

◯ Όχι
◯ 1-2 φορές την εβδομάδα
◯ 3-4 φορές την εβδομάδα
◯ 5-6 φορές την εβδομάδα
◯ 7 ή περισσότερες φορές την εβδομάδα
◯ Δεν γνωρίζω
◯ Δεν απαντώ

Τρέξιμο ή τζόκινγκ

◯ Όχι
◯ 1-2 φορές την εβδομάδα
◯ 3-4 φορές την εβδομάδα
◯ 5-6 φορές την εβδομάδα
◯ 7 ή περισσότερες φορές την εβδομάδα
◯ Δεν γνωρίζω
◯ Δεν απαντώ

Κολύμβηση

◯ Όχι
◯ 1-2 φορές την εβδομάδα
◯ 3-4 φορές την εβδομάδα
◯ 5-6 φορές την εβδομάδα
◯ 7 ή περισσότερες φορές την εβδομάδα
◯ Δεν γνωρίζω
◯ Δεν απαντώ

Χορός ή μπαλέτο

◯ Όχι
◯ 1-2 φορές την εβδομάδα
◯ 3-4 φορές την εβδομάδα
◯ 5-6 φορές την εβδομάδα
◯ 7 ή περισσότερες φορές την εβδομάδα
◯ Δεν γνωρίζω
◯ Δεν απαντώ

Ποδόσφαιρο/Καλαθόσφαιρα/Πετόσφαιρα

◯ Όχι
◯ 1-2 φορές την εβδομάδα
◯ 3-4 φορές την εβδομάδα
◯ 5-6 φορές την εβδομάδα
◯ 7 ή περισσότερες φορές την εβδομάδα
◯ Δεν γνωρίζω
◯ Δεν απαντώ

Πολεμικές τέχνες

◯ Όχι
◯ 1-2 φορές την εβδομάδα
◯ 3-4 φορές την εβδομάδα
◯ 5-6 φορές την εβδομάδα
◯ 7 ή περισσότερες φορές την εβδομάδα
◯ Δεν γνωρίζω
◯ Δεν απαντώ

Άλλη δραστηριότητα

◯ Όχι
◯ 1-2 φορές την εβδομάδα
◯ 3-4 φορές την εβδομάδα
◯ 5-6 φορές την εβδομάδα
◯ 7 ή περισσότερες φορές την εβδομάδα
◯ Δεν γνωρίζω
◯ Δεν απαντώ

Παρακαλώ σημειώστε ποια άλλη δραστηριότητα κάνει το παιδί σας:

__________________________________

4. Ποια από τις πιο κάτω προτάσεις περιγράφει καλύτερα το παιδί σας για την περασμένη εβδομάδα, όσον αφορά τον ελεύθερο χρόνο του;

◯ Έκανε δραστηριότητες που χρειάζονται λίγη σωματική προσπάθεια
◯ Έκανε έντονες δραστηριότητες (π.χ. σπορ/άθλημα, τρέχει, κολυμπά) 1-2 φορές την εβδομάδα
◯ Έκανε έντονες δραστηριότητες (π.χ. σπορ/άθλημα, τρέχει, κολυμπά) 3-4 φορές την εβδομάδα
◯ Έκανει έντονες δραστηριότητες (π.χ. σπορ/άθλημα, τρέχει, κολυμπά) 5-6 φορές την εβδομάδα
◯ Έκανε έντονες δραστηριότητες (π.χ. σπορ/άθλημα, τρέχει, κολυμπά) 7 ή περισσότερες φορές την εβδομάδα
◯ Δεν γνωρίζω
◯ Δεν απαντώ

5. Τι έκανε το παιδί σας συνήθως κατά την διάρκεια του διαλείμματος την περασμένη εβδομάδα (7 ημέρες);

◯ Καθόταν (διάβαζε, μιλούσε, έτρωγε)
◯ Στεκόταν και τριγυρνούσε
◯ Έτρεχε και έπαιζε
◯ Έτρεχε και έπαιζε αρκετά
◯ Έτρεχε και έπαιζε έντονα την περισσότερη ώρα
◯ Δεν γνωρίζω
◯ Δεν απαντώ

6. Την περασμένη εβδομάδα (7 ημέρες) ήταν το παιδί σας άρρωστο ή το εμποδίζε κάτι άλλο από το να κάνει τις φυσικές δραστηριότητες που κάνει συνήθως;

◯ Ναι ◯ Όχι ◯ Δεν γνωρίζω ◯ Δεν απαντώ

Παρακαλώ σημειώστε τι το εμποδίζει:

__________________________________

#### Διατροφικές συνήθειες ΑΥΤΗ ΤΗΝ ΠΕΡΙΟΔΟ, με τη σταδιακή άρση των περιοριστικών μέτρων

7. Πόσο συχνά τρώει το παιδί σας πρωινό (π.χ. δημητριακά με γάλα, σάντουιτς κτλ), ΑΥΤΗ ΤΗΝ ΠΕΡΙΟΔΟ, με τη σταδιακή άρση των περιοριστικών μέτρων;

◯ Κάθε μέρα
◯ 4-6 φορές την εβδομάδα
◯ 2-3 φορές την εβδομάδα
◯ 1 φορά την εβδομάδα
◯ 2-3 φορές τον μήνα
◯ Ποτέ
◯ Δεν γνωρίζω
◯ Δεν απαντώ

---

8. Πόσο συχνά τρώει το παιδί σας έτοιμο φαγητό από έξω (π.χ. εστιατόριο, ψησταριά, fast-food κτλ.), ΑΥΤΗ ΤΗΝ ΠΕΡΙΟΔΟ, με τη σταδιακή άρση των περιοριστικών μέτρων;

◯ Κάθε μέρα
◯ 4-6 φορές την εβδομάδα
◯ 2-3 φορές την εβδομάδα
◯ 1 φορά την εβδομάδα
◯ 2-3 φορές τον μήνα
◯ Ποτέ
◯ Δεν γνωρίζω
◯ Δεν απαντώ

---

9. Λαμβάνει το παιδί σας συμπληρώματα διατροφής (βιταμίνες, ιχνοστοιχεία, ροφήματα) κλπ ΑΥΤΗ ΤΗΝ ΠΕΡΙΟΔΟ, με τη σταδιακή άρση των περιοριστικών μέτρων;

◯ Ναι ◯ Όχι ◯ Δεν γνωρίζω ◯ Δεν απαντώ

---

Παρακαλώ συμπληρώστε το είδος του συμπληρώματος διατροφής που λαμβάνει το παιδί σας ΑΥΤΗ ΤΗΝ ΠΕΡΙΟΔΟ, με τη σταδιακή άρση των περιοριστικών μέτρων:

__________________________________

---

10. Πόσο συχνά καταναλώνει φρούτα το παιδί σας ΑΥΤΗ ΤΗΝ ΠΕΡΙΟΔΟ, με τη σταδιακή άρση των περιοριστικών μέτρων;

◯ Κάθε μέρα
◯ 4-6 φορές την εβδομάδα
◯ 2-3 φορές την εβδομάδα
◯ 1 φορά την εβδομάδα
◯ 2-3 φορές τον μήνα
◯ 1 φορά τον μήνα
◯ Ποτέ
◯ Δεν γνωρίζω
◯ Δεν απαντώ

---

Κατά μέσο όρο, πόσες μερίδες φρούτων καταναλώνει τη μέρα το παιδί σας ΑΥΤΗ ΤΗΝ ΠΕΡΙΟΔΟ, με τη σταδιακή άρση των περιοριστικών μέτρων;

Σημείωση: Μια μερίδα ισοδυναμεί με: 1 μήλο, 1 αχλάδι, 1 ροδάκινο, 1 πορτοκάλι, 2 μανταρίνια, 1 φέτα καρπούζι/πεπόνι, 7 μικρές φράουλες, 14 κεράσια, 2 δαμάσκηνα, 2 φορμόζες, 2 ακτινίδια, 3 κουταλιές φρουτοσαλάτα (χωρίς κομπόστο ή ζάχαρη), 1 μικρό ποτήρι (150ml) φρέσκο χυμό φρούτων

__________________________________

11. Πόσο συχνά καταναλώνει λαχανικά το παιδί σας ΑΥΤΗ ΤΗΝ ΠΕΡΙΟΔΟ, με τη σταδιακή άρση των περιοριστικών μέτρων;

◯ Κάθε μέρα
◯ 4-6 φορές την εβδομάδα
◯ 2-3 φορές την εβδομάδα
◯ 1 φορά την εβδομάδα
◯ 2-3 φορές τον μήνα
◯ 1 φορά τον μήνα
◯ Ποτέ
◯ Δεν γνωρίζω
◯ Δεν απαντώ

Κατά μέσο όρο, πόσες μερίδες λαχανικών καταναλώνει τη μέρα το παιδί σας, ΑΥΤΗ ΤΗΝ ΠΕΡΙΟΔΟ, με τη σταδιακή άρση των περιοριστικών μέτρων;

Σημείωση: Μια μερίδα ισοδυναμεί με: 2 κομμάτια μπρόκολο, 2 μεγάλα κομμάτια κουνουπίδι, ½ φλιτζάνι μαγειρεμένα λαχανικά (π.χ. φασολάκι, κουνουπίδι), 1 μέτρια ντομάτα, 1 αγγουράκι, 1 καρότο, 1 φλιτζάνι σαλάτα από φρέσκα λαχανικά, 1 μικρό ποτήρι (150ml) φρέσκο χυμό λαχανικών

______________________________

12. Πόσο συχνά καταναλώνει κρέας το παιδί σας ΑΥΤΗ ΤΗΝ ΠΕΡΙΟΔΟ, με τη σταδιακή άρση των περιοριστικών μέτρων;

◯ Κάθε μέρα
◯ 4-6 φορές την εβδομάδα
◯ 2-3 φορές την εβδομάδα
◯ 1 φορά την εβδομάδα
◯ 2-3 φορές τον μήνα
◯ 1 φορά τον μήνα
◯ Ποτέ
◯ Δεν γνωρίζω
◯ Δεν απαντώ

Κατά μέσο όρο, πόσες μερίδες κρέατος καταναλώνει τη μέρα το παιδί σας, ΑΥΤΗ ΤΗΝ ΠΕΡΙΟΔΟ, με τη σταδιακή άρση των περιοριστικών μέτρων;

Σημείωση: Μια μερίδα ισοδυναμεί με: 90 γραμμάρια κρέας (χοιρινό, κοτόπουλο βοδινό, αρνί, κατσίκι, σούβλα, κιμάς, σουβλάκι, κλέφτικο, μπριζόλα, συκώτι), 90 γραμμάρια λούντζα/χαμ, 1 φέτα σαλάμι/μπέικον/μορταδέλα, 120 γραμμάρια κονσέρβα κρέατος (π.χ. ZWAN)

______________________________

13. Πόσο συχνά καταναλώνει όσπρια το παιδί σας ΑΥΤΗ ΤΗΝ ΠΕΡΙΟΔΟ, με τη σταδιακή άρση των περιοριστικών μέτρων;

◯ Κάθε μέρα
◯ 4-6 φορές την εβδομάδα
◯ 2-3 φορές την εβδομάδα
◯ 1 φορά την εβδομάδα
◯ 2-3 φορές τον μήνα
◯ 1 φορά τον μήνα
◯ Ποτέ
◯ Δεν γνωρίζω
◯ Δεν απαντώ

Κατά μέσο όρο, πόσες μερίδες όσπρια καταναλώνει τη μέρα το παιδί σας ΑΥΤΗ ΤΗΝ ΠΕΡΙΟΔΟ, με τη σταδιακή άρση των περιοριστικών μέτρων;

Σημείωση: Μια μερίδα ισοδυναμεί με: 90 γραμμάρια φασόλια, φακές, λουβιά, ρεβίθια, κουκιά, μπιζέλι, σόγια

__________________________________

14. Πόσο συχνά καταναλώνει ψάρι/θαλασσινά το παιδί σας ΑΥΤΗ ΤΗΝ ΠΕΡΙΟΔΟ, με τη σταδιακή άρση των περιοριστικών μέτρων;

◯ Κάθε μέρα
◯ 4-6 φορές την εβδομάδα
◯ 2-3 φορές την εβδομάδα
◯ 1 φορά την εβδομάδα
◯ 2-3 φορές τον μήνα
◯ 1 φορά τον μήνα
◯ Ποτέ
◯ Δεν γνωρίζω
◯ Δεν απαντώ

Κατά μέσο όρο, πόσες μερίδες ψάρι/θαλασσινά καταναλώνει τη μέρα το παιδί σας ΑΥΤΗ ΤΗΝ ΠΕΡΙΟΔΟ, με τη σταδιακή άρση των περιοριστικών μέτρων;

Σημείωση: Μια μερίδα ισοδυναμεί με: 90 γραμμάρια ψάρι ή θαλασσινά

__________________________________

15. Πόσο συχνά καταναλώνει το παιδί σας προϊόντα που περιέχουν ζάχαρη ΑΥΤΗ ΤΗΝ ΠΕΡΙΟΔΟ, με τη σταδιακή άρση των περιοριστικών μέτρων;

◯ Κάθε μέρα
◯ 4-6 φορές την εβδομάδα
◯ 2-3 φορές την εβδομάδα
◯ 1 φορά την εβδομάδα
◯ 2-3 φορές τον μήνα
◯ 1 φορά τον μήνα
◯ Ποτέ
◯ Δεν γνωρίζω
◯ Δεν απαντώ

Κατά μέσο όρο, πόσες μερίδες ζάχαρης καταναλώνει τη μέρα το παιδί σας ΑΥΤΗ ΤΗΝ ΠΕΡΙΟΔΟ, με τη σταδιακή άρση των περιοριστικών μέτρων;

Σημείωση: Μια μερίδα ισοδυναμεί με: 1 κουταλάκι γλυκού ζάχαρη/μέλι/μαρμελάδα/nutella, 45 γραμμάρια σοκολάτα, 1 μπισκότο, 1 μπάλα παγωτού, 1 κομμάτι κέικ/τάρτας, 1 μπολ κρέμας (π.χ. ρυζόγαλο)

__________________________________

16. Πόσο συχνά καταναλώνει το παιδί σας τσιπς, γαριδάκια, είδη αρτοποιείου και αλατισμένους ξηρούς καρπούς ΑΥΤΗ ΤΗΝ ΠΕΡΙΟΔΟ, με τη σταδιακή άρση των περιοριστικών μέτρων;

◯ Κάθε μέρα
◯ 4-6 φορές την εβδομάδα
◯ 2-3 φορές την εβδομάδα
◯ 1 φορά την εβδομάδα
◯ 2-3 φορές τον μήνα
◯ 1 φορά τον μήνα
◯ Ποτέ
◯ Δεν γνωρίζω
◯ Δεν απαντώ

Κατά μέσο όρο, πόσες μερίδες τσιπς, γαριδάκια, είδη αρτοποιείου και αλατισμένους ξηρούς καρπούς καταναλώνει τη μέρα το παιδί σας ΑΥΤΗ ΤΗΝ ΠΕΡΙΟΔΟ, με τη σταδιακή άρση των περιοριστικών μέτρων;

Σημείωση: Μια μερίδα ισοδυναμεί με: 4 αλμυρά (κοκτέιλ), 1 κουτί τσιπς/γαριδάκια, 15 ξηρούς καρπούς

__________________________________

#### Κάπνισμα ΑΥΤΗ ΤΗΝ ΠΕΡΙΟΔΟ, με τη σταδιακή άρση των περιοριστικών μέτρων

17. Καπνίζει κάποιο μέλος της οικογένειας μέσα στο σπίτι, ΑΥΤΗ ΤΗΝ ΠΕΡΙΟΔΟ, με τη σταδιακή άρση των περιοριστικών μέτρων;

◯ Ναι, καθημερινά
◯ Ναι, περιστασιακά
◯ Όχι
◯ Δεν γνωρίζω/δεν είμαι σίγουρος/δεν θυμάμαι
◯ Δεν απαντώ

---

Πόσα τσιγάρα καπνίζει κατά την διάρκεια της μέρας στο σπίτι, ΑΥΤΗ ΤΗΝ ΠΕΡΙΟΔΟ, με τη σταδιακή άρση των περιοριστικών μέτρων;

________________________________

---

Πόσα τσιγάρα καπνίζει κατά την διάρκεια της εβδομάδας στο σπίτι, ΑΥΤΗ ΤΗΝ ΠΕΡΙΟΔΟ, με τη σταδιακή άρση των περιοριστικών μέτρων;

________________________________

#### Κοινωνική συμπεριφορά ΑΥΤΗ ΤΗΝ ΠΕΡΙΟΔΟ, με τη σταδιακή άρση των περιοριστικών μέτρων

18. Πόσο συχνά επικοινωνεί το παιδί σας με φίλους/οικογένεια μέσω τηλεφώνου ή διαδικτύου (με email, viber, whatsapp, στα μέσα μαζικής δικτύωσης π.χ. Facebook, Instagram, κτλ) ΑΥΤΗ ΤΗΝ ΠΕΡΙΟΔΟ, με τη σταδιακή άρση των περιοριστικών μέτρων;

Φίλοι

◯ Κάθε μέρα
◯ Κάθε εβδομάδα αλλά όχι κάθε μέρα
◯ Κάποιες φορές το μήνα αλλά όχι κάθε εβδομάδα
◯ Ποτέ
◯ Δεν γνωρίζω
◯ Δεν απαντώ

Πόσες ώρες τη μέρα επικοινωνεί το παιδί σας με τους φίλους του/της μέσω τηλεφώνου ή διαδικτύου (με email, viber, whatsapp, στα μέσα μαζικής δικτύωσης π.χ. Facebook, Instagram, κτλ) ΑΥΤΗ ΤΗΝ ΠΕΡΙΟΔΟ, με τη σταδιακή άρση των περιοριστικών μέτρων;

______________________________
(Ώρες τη μέρα)

Οικογένεια/συγγενείς

◯ Κάθε μέρα
◯ Κάθε εβδομάδα αλλά όχι κάθε μέρα
◯ Κάποιες φορές το μήνα αλλά όχι κάθε εβδομάδα
◯ Ποτέ
◯ Δεν γνωρίζω
◯ Δεν απαντώ

Πόσες ώρες τη μέρα επικοινωνεί το παιδί σας με την οικογένεια/συγγενείς μέσω τηλεφώνου ή διαδικτύου (με email, viber, whatsapp, στα μέσα μαζικής δικτύωσης π.χ. Facebook, Instagram, κτλ) ΑΥΤΗ ΤΗΝ ΠΕΡΙΟΔΟ, με τη σταδιακή άρση των περιοριστικών μέτρων;

______________________________
(Ώρες τη μέρα)

19. Πόσες ώρες τη μέρα το παιδί σας παρακολουθεί τηλεόραση ή ασχολείται με τον υπολογιστή, το κινητό, το tablet ή με οποιαδήποτε ηλεκτρονική κονσόλα παιχνιδιών, ΑΥΤΗ ΤΗΝ ΠΕΡΙΟΔΟ, με τη σταδιακή άρση των περιοριστικών μέτρων;

◯ Λιγότερο από 1 ώρα τη μέρα
◯ 1-3 ώρες τη μέρα
◯ 4-7 ώρες τη μέρα
◯ 8-11 ώρες τη μέρα
◯ Περισσότερο από 11 ώρες τη μέρα
◯ Δεν γνωρίζω
◯ Δεν απαντώ

**Προσωπική υγιεινή και καθαριότητα σπιτιού ΑΥΤΗ ΤΗΝ ΠΕΡΙΟΔΟ, με τη σταδιακή άρση των περιοριστικών μέτρων**

20. Πόσες φορές την ημέρα, κατά μέσο όρο, το παιδί σας πλένει ή τρίβει τα χέρια του/της με σαπούνι/απολυμαντικά (αντισηπτικά) προϊόντα, ΑΥΤΗ ΤΗΝ ΠΕΡΙΟΔΟ, με τη σταδιακή άρση των περιοριστικών μέτρων;

Σαπούνι

◯ Ποτέ
◯ < 1 φορά τη μέρα
◯ 1-3 φορές τη μέρα
◯ 4-7 φορές τη μέρα
◯ >7 φορές τη μέρα
◯ Δεν γνωρίζω
◯ Δεν απαντώ

Απολυμαντικά (αντισηπτικά) προϊόντα

◯ Ποτέ
◯ < 1 φορά τη μέρα
◯ 1-3 φορές τη μέρα
◯ 4-7 φορές τη μέρα
◯ >7 φορές τη μέρα
◯ Δεν γνωρίζω
◯ Δεν απαντώ

21. Πόσες μέρες την εβδομάδα εκτελούνται, κατά μέσο όρο, οι ακόλουθες δραστηριότητες με τη χρήση απολυμαντικών/αντισηπτικών προϊόντων στο σπίτι, ΑΥΤΗ ΤΗΝ ΠΕΡΙΟΔΟ, με τη σταδιακή άρση των περιοριστικών μέτρων;

Καθαρισμός χώρων υγιεινής (π.χ. μπάνιο, τουαλέτα) με τη χρήση απολυμαντικών/αντισηπτικών προϊόντων:

________________________________
(Μέρες ανά εβδομάδα, ΑΥΤΗ ΤΗΝ ΠΕΡΙΟΔΟ, με τη σταδιακή άρση των περιοριστικών μέτρων)

Παρακαλώ σημειώστε το όνομα του προϊόντος που χρησιμοποιείτε για τον καθαρισμό χώρων υγιεινής:

________________________________

Καθαρισμός επιφανειών κουζίνας με τη χρήση απολυμαντικών/αντισηπτικών προϊόντων:

________________________________
(Μέρες ανά εβδομάδα, ΑΥΤΗ ΤΗΝ ΠΕΡΙΟΔΟ, με τη σταδιακή άρση των περιοριστικών μέτρων)

Παρακαλώ σημειώστε το όνομα του προϊόντος που χρησιμοποιείτε για τον καθαρισμό επιφανειών κουζίνας:

________________________________

Σφουγγάρισμα δαπέδων με τη χρήση απολυμαντικών/αντισηπτικών προϊόντων:

________________________________
(Μέρες ανά εβδομάδα, ΑΥΤΗ ΤΗΝ ΠΕΡΙΟΔΟ, με τη σταδιακή άρση των περιοριστικών μέτρων)

Παρακαλώ σημειώστε το όνομα του προϊόντος που χρησιμοποιείτε για το σφουγγάρισμα των δαπέδων:

________________________________

Καθαρισμός επιφανειών σε άλλους χώρους εκτός κουζίνας με τη χρήση απολυμαντικών/αντισηπτικών προϊόντων:

__________________________________

(Μέρες ανά εβδομάδα, ΑΥΤΗ  ΤΗΝ ΠΕΡΙΟΔΟ, με τη σταδιακή άρση των περιοριστικών μέτρων)

Παρακαλώ σημειώστε το όνομα του προϊόντος που χρησιμοποιείτε για τον καθαρισμό επιφανειών σε άλλους χώρους εκτός κουζίνας:

__________________________________

#### Χρόνος στο σπίτι, στο σχολείο και επαφή με άλλα άτομα ΑΥΤΗ ΤΗΝ ΠΕΡΙΟΔΟ, με τη σταδιακή άρση των περιοριστικών μέτρων

22α. Κατά μέσο όρο, πόσες ώρες βρίσκεται στο σπίτι το παιδί σας μια καθημερινή μέρα (Δευτέρα-Παρασκευή) ΑΥΤΗ ΤΗΝ ΠΕΡΙΟΔΟ, με τη σταδιακή άρση των περιοριστικών μέτρων;

__________________________________

22β. Με πόσα άτομα έρχεται σε επαφή το παιδί σας από Δευτέρα μέχρι Παρασκευή, κατά μέσο όρο, ΑΥΤΗ ΤΗΝ ΠΕΡΙΟΔΟ, με τη σταδιακή άρση των περιοριστικών μέτρων;

Σημείωση: επαφή ορίζεται ως είτε η αμφίδρομη συζήτηση με τρεις ή περισσότερες λέξεις μεταξύ της φυσικής παρουσίας δύο ατόμων είτε η φυσική επαφή (δέρμα με δέρμα) (π.χ. χειραψία, αγκαλιά, φιλί)

Αριθμός ατόμων στο σπίτι με τα οποία έρχεται σε επαφή το παιδί σας από Δευτέρα μέχρι Παρασκευή, ΑΥΤΗ ΤΗΝ ΠΕΡΙΟΔΟ, με τη σταδιακή άρση των περιοριστικών μέτρων:

__________________________________

Πόσα άτομα με τα οποία έρχεται σε επαφή το παιδί σας στο σπίτι από Δευτέρα μέχρι Παρασκευή ανήκουν σε ευπαθείς ομάδες, ΑΥΤΗ ΤΗΝ ΠΕΡΙΟΔΟ, με τη σταδιακή άρση των περιοριστικών μέτρων;
Στις ευπαθείς ομάδες ανήκουν τα άτομα άνω των 60 ετών, άτομα με χρόνιες παθήσεις και έγκυες γυναίκες.

__________________________________

Αριθμός ατόμων στο σχολείο με τα οποία έρχεται σε επαφή το παιδί σας από Δευτέρα μέχρι Παρασκευή, ΑΥΤΗ ΤΗΝ ΠΕΡΙΟΔΟ, με τη σταδιακή άρση των περιοριστικών μέτρων:

__________________________________

Αριθμός ατόμων εκτός σπιτιού και εκτός σχολείου με τα οποία έρχεται σε επαφή το παιδί σας από Δευτέρα μέχρι Παρασκευή, ΑΥΤΗ ΤΗΝ ΠΕΡΙΟΔΟ, με τη σταδιακή άρση των περιοριστικών μέτρων:

__________________________________

23α. Κατά μέσο όρο, πόσες ώρες βρίσκεται στο σπίτι το παιδί σας το σαββατοκύριακο ΑΥΤΗ ΤΗΝ ΠΕΡΙΟΔΟ, με τη σταδιακή άρση των περιοριστικών μέτρων;

__________________________________

23β. Με πόσα άτομα έρχεται σε επαφή το παιδί σας το σαββατοκύριακο, κατά μέσο όρο, ΑΥΤΗ ΤΗΝ ΠΕΡΙΟΔΟ, με τη σταδιακή άρση των περιοριστικών μέτρων;
Σημείωση: επαφή ορίζεται ως είτε η αμφίδρομη συζήτηση με τρεις ή περισσότερες λέξεις μεταξύ της φυσικής παρουσίας δύο ατόμων είτε η φυσική επαφή (δέρμα με δέρμα) (π.χ. χειραψία, αγκαλιά, φιλί)

Αριθμός ατόμων στο σπίτι με τα οποία έρχεται σε επαφή το παιδί σας το σαββατοκύριακο, ΑΥΤΗ ΤΗΝ ΠΕΡΙΟΔΟ, με τη σταδιακή άρση των περιοριστικών μέτρων:

__________________________________

Πόσα άτομα με τα οποία έρχεται σε επαφή το παιδί σας στο σπίτι το σαββατοκύριακο ανήκουν σε ευπαθείς ομάδες, ΑΥΤΗ ΤΗΝ ΠΕΡΙΟΔΟ, με τη σταδιακή άρση των περιοριστικών μέτρων;
Στις ευπαθείς ομάδες ανήκουν τα άτομα άνω των 60 ετών, άτομα με χρόνιες παθήσεις και έγκυες γυναίκες.

__________________________________

Αριθμός ατόμων εκτός σπιτιού με τα οποία έρχεται σε επαφή το παιδί σας το σαββατοκύριακο, ΑΥΤΗ ΤΗΝ ΠΕΡΙΟΔΟ, με τη σταδιακή άρση των περιοριστικών μέτρων:

__________________________________

#### Σωματική άσκηση ΠΡΙΝ ΤΗΝ ΠΑΝΔΗΜΙΑ, δηλαδή πριν την εφαρμογή των μέτρων περιορισμού

24. Έκανε το παιδί σας κάποια/κάποιες από τις πιο κάτω δραστηριότητες ΠΡΙΝ ΤΗΝ ΠΑΝΔΗΜΙΑ, δηλαδή πριν την εφαρμογή των μέτρων περιορισμού; Εάν ναι, επιλέξτε πόσες φορές την εβδομάδα.

Έντονο περπάτημα

- ◯ Όχι
- ◯ 1-2 φορές την εβδομάδα
- ◯ 3-4 φορές την εβδομάδα
- ◯ 5-6 φορές την εβδομάδα
- ◯ 7 ή περισσότερες φορές την εβδομάδα
- ◯ Δεν γνωρίζω
- ◯ Δεν απαντώ

Ποδήλατο

- ◯ Όχι
- ◯ 1-2 φορές την εβδομάδα
- ◯ 3-4 φορές την εβδομάδα
- ◯ 5-6 φορές την εβδομάδα
- ◯ 7 ή περισσότερες φορές την εβδομάδα
- ◯ Δεν γνωρίζω
- ◯ Δεν απαντώ

Τρέξιμο ή τζόκινγκ

- ◯ Όχι
- ◯ 1-2 φορές την εβδομάδα
- ◯ 3-4 φορές την εβδομάδα
- ◯ 5-6 φορές την εβδομάδα
- ◯ 7 ή περισσότερες φορές την εβδομάδα
- ◯ Δεν γνωρίζω
- ◯ Δεν απαντώ

Κολύμβηση

- ◯ Όχι
- ◯ 1-2 φορές την εβδομάδα
- ◯ 3-4 φορές την εβδομάδα
- ◯ 5-6 φορές την εβδομάδα
- ◯ 7 ή περισσότερες φορές την εβδομάδα
- ◯ Δεν γνωρίζω
- ◯ Δεν απαντώ

Χορός ή μπαλέτο

- ◯ Όχι
- ◯ 1-2 φορές την εβδομάδα
- ◯ 3-4 φορές την εβδομάδα
- ◯ 5-6 φορές την εβδομάδα
- ◯ 7 ή περισσότερες φορές την εβδομάδα
- ◯ Δεν γνωρίζω
- ◯ Δεν απαντώ

Ποδόσφαιρο/Καλαθόσφαιρα/Πετόσφαιρα

◯ Όχι
◯ 1-2 φορές την εβδομάδα
◯ 3-4 φορές την εβδομάδα
◯ 5-6 φορές την εβδομάδα
◯ 7 ή περισσότερες φορές την εβδομάδα
◯ Δεν γνωρίζω
◯ Δεν απαντώ

Πολεμικές τέχνες

◯ Όχι
◯ 1-2 φορές την εβδομάδα
◯ 3-4 φορές την εβδομάδα
◯ 5-6 φορές την εβδομάδα
◯ 7 ή περισσότερες φορές την εβδομάδα
◯ Δεν γνωρίζω
◯ Δεν απαντώ

Άλλη δραστηριότητα

◯ Όχι
◯ 1-2 φορές την εβδομάδα
◯ 3-4 φορές την εβδομάδα
◯ 5-6 φορές την εβδομάδα
◯ 7 ή περισσότερες φορές την εβδομάδα
◯ Δεν γνωρίζω
◯ Δεν απαντώ

Παρακαλώ σημειώστε ποια άλλη δραστηριότητα έκανε το παιδί σας:

__________________________________

25. Ποια από τις πιο κάτω προτάσεις περιγράφει καλύτερα το παιδί σας όσον αφορά τον ελεύθερο χρόνο του ΠΡΙΝ ΤΗΝ ΠΑΝΔΗΜΙΑ, δηλαδή πριν την εφαρμογή των μέτρων περιορισμού;

◯ Έκανε δραστηριότητες που χρειάζονται λίγη σωματική προσπάθεια
◯ Έκανε έντονες δραστηριότητες (π.χ. σπορ/άθλημα, τρέχει, κολυμπά) 1-2 φορές την εβδομάδα
◯ Έκανε έντονες δραστηριότητες (π.χ. σπορ/άθλημα, τρέχει, κολυμπά) 3-4 φορές την εβδομάδα
◯ Έκανε έντονες δραστηριότητες (π.χ. σπορ/άθλημα, τρέχει, κολυμπά) 5-6 φορές την εβδομάδα
◯ Έκανε έντονες δραστηριότητες (π.χ. σπορ/άθλημα, τρέχει, κολυμπά) 7 ή περισσότερες φορές την εβδομάδα
◯ Δεν γνωρίζω
◯ Δεν απαντώ

26. Τι έκανε το παιδί σας συνήθως κατά την διάρκεια του διαλείμματος ΠΡΙΝ ΤΗΝ ΠΑΝΔΗΜΙΑ, δηλαδή πριν την εφαρμογή των μέτρων περιορισμού;

◯ Καθόταν (διάβαζε, μιλούσε)
◯ Στεκόταν και τριγυρνούσε
◯ Έτρεχε και έπαιζε
◯ Έτρεχε και έπαιζε αρκετά
◯ Έτρεχε και έπαιζε έντονα την περισσότερη ώρα
◯ Έτρωγε
◯ Δεν γνωρίζω
◯ Δεν απαντώ

27. Ήταν το παιδί σας άρρωστο ή το εμπόδισε κάτι άλλο από το να κάνει τις φυσικές δραστηριότητες που κάνει συνήθως ΠΡΙΝ ΤΗΝ ΠΑΝΔΗΜΙΑ, δηλαδή πριν την εφαρμογή των μέτρων περιορισμού;

◯ Ναι ◯ Όχι ◯ Δεν γνωρίζω ◯ Δεν απαντώ

Παρακαλώ σημειώστε τι το εμπόδιζε:

__________________________________

**Διατροφικές συνήθειες ΠΡΙΝ ΤΗΝ ΠΑΝΔΗΜΙΑ, δηλαδή πριν την εφαρμογή των μέτρων περιορισμού**

28. Πόσο συχνά έτρωγε το παιδί σας πρωινό (π.χ. δημητριακά με γάλα, σάντουιτς κτλ) ΠΡΙΝ ΤΗΝ ΠΑΝΔΗΜΙΑ, δηλαδή πριν την εφαρμογή των μέτρων περιορισμού;

◯ Κάθε μέρα
◯ 4-6 φορές την εβδομάδα
◯ 2-3 φορές την εβδομάδα
◯ 1 φορά την εβδομάδα
◯ 2-3 φορές τον μήνα
◯ Ποτέ
◯ Δεν γνωρίζω
◯ Δεν απαντώ

29. Πόσο συχνά έτρωγε το παιδί σας έτοιμο φαγητό από έξω (π.χ. εστιατόριο, ψησταριά, fast-food κτλ.) ΠΡΙΝ ΤΗΝ ΠΑΝΔΗΜΙΑ, δηλαδή πριν την εφαρμογή των μέτρων περιορισμού;

◯ Κάθε μέρα
◯ 4-6 φορές την εβδομάδα
◯ 2-3 φορές την εβδομάδα
◯ 1 φορά την εβδομάδα
◯ 2-3 φορές τον μήνα
◯ Ποτέ
◯ Δεν γνωρίζω
◯ Δεν απαντώ

30.  Λάμβανε το παιδί σας συμπληρώματα διατροφής (βιταμίνες, ιχνοστοιχεία, ροφήματα) κλπ ΠΡΙΝ ΤΗΝ ΠΑΝΔΗΜΙΑ, δηλαδή πριν την εφαρμογή των μέτρων περιορισμού;

◯ Ναι ◯ Όχι ◯ Δεν γνωρίζω ◯ Δεν απαντώ

Παρακαλώ συμπληρώστε το είδος του συμπληρώματος διατροφής που λαμβάνει το παιδί σας ΠΡΙΝ ΤΗΝ ΠΑΝΔΗΜΙΑ, δηλαδή πριν την εφαρμογή των μέτρων περιορισμού:

__________________________________

31. Πόσο συχνά κατανάλωνε φρούτα το παιδί σας, ΠΡΙΝ ΤΗΝ ΠΑΝΔΗΜΙΑ, δηλαδή πριν την εφαρμογή των μέτρων περιορισμού;

◯ Κάθε μέρα
◯ 4-6 φορές την εβδομάδα
◯ 2-3 φορές την εβδομάδα
◯ 1 φορά την εβδομάδα
◯ 2-3 φορές τον μήνα
◯ 1 φορά τον μήνα
◯ Ποτέ
◯ Δεν γνωρίζω
◯ Δεν απαντώ

Κατά μέσο όρο, πόσες μερίδες φρούτων κατανάλωνε τη μέρα το παιδί σας ΠΡΙΝ ΤΗΝ ΠΑΝΔΗΜΙΑ, δηλαδή πριν την εφαρμογή των μέτρων περιορισμού;

Σημείωση: Μια μερίδα ισοδυναμεί με: 1 μήλο, 1 αχλάδι, 1 ροδάκινο, 1 πορτοκάλι, 2 μανταρίνια, 1 φέτα καρπούζι/πεπόνι, 7 μικρές φράουλες, 14 κεράσια, 2 δαμάσκηνα, 2 φορμόζες, 2 ακτινίδια, 3 κουταλιές φρουτοσαλάτα (χωρίς κομπόστο ή ζάχαρη), 1 μικρό ποτήρι (150ml) φρέσκο χυμό φρούτων

__________________________________

32. Πόσο συχνά κατανάλωνε λαχανικά το παιδί σας, ΠΡΙΝ ΤΗΝ ΠΑΝΔΗΜΙΑ, δηλαδή πριν την εφαρμογή των μέτρων περιορισμού;

◯ Κάθε μέρα
◯ 4-6 φορές την εβδομάδα
◯ 2-3 φορές την εβδομάδα
◯ 1 φορά την εβδομάδα
◯ 2-3 φορές τον μήνα
◯ 1 φορά τον μήνα
◯ Ποτέ
◯ Δεν γνωρίζω
◯ Δεν απαντώ

Κατά μέσο όρο, πόσες μερίδες λαχανικών κατανάλωνε τη μέρα το παιδί σας, ΠΡΙΝ ΤΗΝ ΠΑΝΔΗΜΙΑ, δηλαδή πριν την εφαρμογή των μέτρων περιορισμού;

Σημείωση: Μια μερίδα ισοδυναμεί με: 2 κομμάτια μπρόκολο, 2 μεγάλα κομμάτια κουνουπίδι, ½ φλιτζάνι μαγειρεμένα λαχανικά (π.χ. φασολάκι, κουνουπίδι), 1 μέτρια ντομάτα, 1 αγγουράκι, 1 καρότο, 1 φλιτζάνι σαλάτα από φρέσκα λαχανικά, 1 μικρό ποτήρι (150ml) φρέσκο χυμό λαχανικών

__________________________________

33. Πόσο συχνά κατανάλωνε κρέας το παιδί σας, ΠΡΙΝ ΤΗΝ ΠΑΝΔΗΜΙΑ, δηλαδή πριν την εφαρμογή των μέτρων περιορισμού;

◯ Κάθε μέρα
◯ 4-6 φορές την εβδομάδα
◯ 2-3 φορές την εβδομάδα
◯ 1 φορά την εβδομάδα
◯ 2-3 φορές τον μήνα
◯ 1 φορά τον μήνα
◯ Ποτέ
◯ Δεν γνωρίζω
◯ Δεν απαντώ

Κατά μέσο όρο, πόσες μερίδες κρέατος κατανάλωνε τη μέρα το παιδί σας, ΠΡΙΝ ΤΗΝ ΠΑΝΔΗΜΙΑ, δηλαδή πριν την εφαρμογή των μέτρων περιορισμού;

Σημείωση: Μια μερίδα ισοδυναμεί με: 90 γραμμάρια κρέας (χοιρινό, κοτόπουλο βοδινό, αρνί, κατσίκι, σούβλα, κιμάς, σουβλάκι, κλέφτικο, μπριζόλα, συκώτι), 90 γραμμάρια λούντζα/χαμ, 1 φέτα σαλάμι/μπέικον/μορταδέλα, 120 γραμμάρια κονσέρβα κρέατος (π.χ. ZWAN)

__________________________________

34. Πόσο συχνά κατανάλωνε όσπρια το παιδί σας, ΠΡΙΝ ΤΗΝ ΠΑΝΔΗΜΙΑ, δηλαδή πριν την εφαρμογή των μέτρων περιορισμού;

◯ Κάθε μέρα
◯ 4-6 φορές την εβδομάδα
◯ 2-3 φορές την εβδομάδα
◯ 1 φορά την εβδομάδα
◯ 2-3 φορές τον μήνα
◯ 1 φορά τον μήνα
◯ Ποτέ
◯ Δεν γνωρίζω
◯ Δεν απαντώ

Κατά μέσο όρο, πόσες μερίδες όσπρια κατανάλωνε τη μέρα το παιδί σας, ΠΡΙΝ ΤΗΝ ΠΑΝΔΗΜΙΑ, δηλαδή πριν την εφαρμογή των μέτρων περιορισμού;

Σημείωση: Μια μερίδα ισοδυναμεί με: 90 γραμμάρια φασόλια, φακές, λουβιά, ρεβίθια, κουκιά, μπιζέλι, σόγια

__________________________________

35. Πόσο συχνά κατανάλωνε ψάρι/θαλασσινά το παιδί σας, ΠΡΙΝ ΤΗΝ ΠΑΝΔΗΜΙΑ, δηλαδή πριν την εφαρμογή των μέτρων περιορισμού;

◯ Κάθε μέρα
◯ 4-6 φορές την εβδομάδα
◯ 2-3 φορές την εβδομάδα
◯ 1 φορά την εβδομάδα
◯ 2-3 φορές τον μήνα
◯ 1 φορά τον μήνα
◯ Ποτέ
◯ Δεν γνωρίζω
◯ Δεν απαντώ

Κατά μέσο όρο, πόσες μερίδες ψάρι/θαλασσινά κατανάλωνε τη μέρα το παιδί σας, ΠΡΙΝ ΤΗΝ ΠΑΝΔΗΜΙΑ, δηλαδή πριν την εφαρμογή των μέτρων περιορισμού;

Σημείωση: Μια μερίδα ισοδυναμεί με: 90 γραμμάρια ψάρι ή θαλασσινά

__________________________________

36. Πόσο συχνά κατανάλωνε το παιδί σας προϊόντα που περιείχαν ζάχαρη, ΠΡΙΝ ΤΗΝ ΠΑΝΔΗΜΙΑ, δηλαδή πριν την εφαρμογή των μέτρων περιορισμού;

◯ Κάθε μέρα
◯ 4-6 φορές την εβδομάδα
◯ 2-3 φορές την εβδομάδα
◯ 1 φορά την εβδομάδα
◯ 2-3 φορές τον μήνα
◯ 1 φορά τον μήνα
◯ Ποτέ
◯ Δεν γνωρίζω
◯ Δεν απαντώ

Κατά μέσο όρο, πόσες μερίδες ζάχαρης κατανάλωνε τη μέρα το παιδί σας ΠΡΙΝ ΤΗΝ ΠΑΝΔΗΜΙΑ, δηλαδή πριν την εφαρμογή των μέτρων περιορισμού;

Σημείωση: Μια μερίδα ισοδυναμεί με: 1 κουταλάκι γλυκού ζάχαρη/μέλι/μαρμελάδα/nutella, 45 γραμμάρια σοκολάτα, 1 μπισκότο, 1 μπάλα παγωτού, 1 κομμάτι κέικ/τάρτας, 1 μπολ κρέμας (π.χ. ρυζόγαλο)

__________________________________

37. Πόσο συχνά κατανάλωνε το παιδί σας τσιπς, γαριδάκια, είδη αρτοποιείου και αλατισμένους ξηρούς καρπούς, ΠΡΙΝ ΤΗΝ ΠΑΝΔΗΜΙΑ, δηλαδή πριν την εφαρμογή των μέτρων περιορισμού;

◯ Κάθε μέρα
◯ 4-6 φορές την εβδομάδα
◯ 2-3 φορές την εβδομάδα
◯ 1 φορά την εβδομάδα
◯ 2-3 φορές τον μήνα
◯ 1 φορά τον μήνα
◯ Ποτέ
◯ Δεν γνωρίζω
◯ Δεν απαντώ

Κατά μέσο όρο, πόσες μερίδες τσιπς, γαριδάκια, είδη αρτοποιείου και αλατισμένους ξηρούς καρπούς κατανάλωνε τη μέρα το παιδί σας ΠΡΙΝ ΤΗΝ ΠΑΝΔΗΜΙΑ, δηλαδή πριν την εφαρμογή των μέτρων περιορισμού;

Σημείωση: Μια μερίδα ισοδυναμεί με: 4 αλμυρά (κοκτέιλ), 1 κουτί τσιπς/γαριδάκια, 15 ξηρούς καρπούς

__________________________________

#### Κάπνισμα ΠΡΙΝ ΤΗΝ ΠΑΝΔΗΜΙΑ, δηλαδή πριν την εφαρμογή των μέτρων περιορισμού

38. Κάπνιζε κάποιο μέλος της οικογένειας μέσα στο σπίτι, ΠΡΙΝ ΤΗΝ ΠΑΝΔΗΜΙΑ, δηλαδή πριν την εφαρμογή των μέτρων περιορισμού;

◯ Ναι, καθημερινά
◯ Ναι, περιστασιακά
◯ Όχι
◯ Δεν γνωρίζω/δεν είμαι σίγουρος/δεν θυμάμαι
◯ Δεν απαντώ

Πόσα τσιγάρα κάπνιζε κατά την διάρκεια της μέρας στο σπίτι, ΠΡΙΝ ΤΗΝ ΠΑΝΔΗΜΙΑ, δηλαδή πριν την εφαρμογή των μέτρων περιορισμού;

__________________________________

Πόσα τσιγάρα κάπνιζε κατά την διάρκεια της εβδομάδας στο σπίτι, ΠΡΙΝ ΤΗΝ ΠΑΝΔΗΜΙΑ, δηλαδή πριν την εφαρμογή των μέτρων περιορισμού;

__________________________________

#### Κοινωνική συμπεριφορά ΠΡΙΝ ΤΗΝ ΠΑΝΔΗΜΙΑ, δηλαδή πριν την εφαρμογή των μέτρων περιορισμού

39. Πόσο συχνά επικοινωνούσε το παιδί σας με φίλους/οικογένεια μέσω τηλεφώνου ή διαδικτύου (με email, viber, whatsapp, στα μέσα μαζικής δικτύωσης π.χ. Facebook, Instagram, κτλ) ΠΡΙΝ ΤΗΝ ΠΑΝΔΗΜΙΑ, δηλαδή πριν την εφαρμογή των μέτρων περιορισμού;

Φίλοι

◯ Κάθε μέρα
◯ Κάθε εβδομάδα αλλά όχι κάθε μέρα
◯ Κάποιες φορές το μήνα αλλά όχι κάθε εβδομάδα
◯ Ποτέ
◯ Δεν γνωρίζω
◯ Δεν απαντώ

Πόσες ώρες τη μέρα επικοινωνούσε το παιδί σας με τους φίλους του/της μέσω τηλεφώνου ή διαδικτύου (με email, viber, whatsapp, στα μέσα μαζικής δικτύωσης π.χ. Facebook, Instagram, κτλ) ΠΡΙΝ ΤΗΝ ΠΑΝΔΗΜΙΑ, δηλαδή πριν την εφαρμογή των μέτρων περιορισμού;

__________________________________
(Ώρες τη μέρα)

Οικογένεια/συγγενείς

◯ Κάθε μέρα
◯ Κάθε εβδομάδα αλλά όχι κάθε μέρα
◯ Κάποιες φορές το μήνα αλλά όχι κάθε εβδομάδα
◯ Ποτέ
◯ Δεν γνωρίζω
◯ Δεν απαντώ

Πόσες ώρες τη μέρα επικοινωνούσε το παιδί σας με την οικογένεια/συγγενείς μέσω τηλεφώνου ή διαδικτύου (με email, viber, whatsapp, στα μέσα μαζικής δικτύωσης π.χ. Facebook, Instagram, κτλ) ΠΡΙΝ ΤΗΝ ΠΑΝΔΗΜΙΑ, δηλαδή πριν την εφαρμογή των μέτρων περιορισμού;

__________________________________
(Ώρες τη μέρα)

40. Πόσες ώρες τη μέρα το παιδί σας παρακολουθούσε τηλεόραση ή ασχολιόταν με τον υπολογιστή, το κινητό, το tablet ή με οποιαδήποτε ηλεκτρονική κονσόλα παιχνιδιών, ΠΡΙΝ ΤΗΝ ΠΑΝΔΗΜΙΑ, δηλαδή πριν την εφαρμογή των μέτρων περιορισμού;

◯ Λιγότερο από 1 ώρα τη μέρα
◯ 1-3 ώρες τη μέρα
◯ 4-7 ώρες τη μέρα
◯ 8-11 ώρες τη μέρα
◯ Περισσότερο από 11 ώρες τη μέρα
◯ Δεν γνωρίζω
◯ Δεν απαντώ

**Προσωπική υγιεινή και καθαριότητα σπιτιού ΠΡΙΝ ΤΗΝ ΠΑΝΔΗΜΙΑ, δηλαδή πριν την εφαρμογή των περιοριστικών μέτρων**

41. Πόσες φορές την ημέρα, κατά μέσο όρο, το παιδί σας έπλενε ή έτριβε τα χέρια του/της με σαπούνι/απολυμαντικά (αντισηπτικά) προϊόντα, ΠΡΙΝ ΤΗΝ ΠΑΝΔΗΜΙΑ, δηλαδή πριν την εφαρμογή των μέτρων περιορισμού;

Σαπούνι

◯ Ποτέ
◯ < 1 φορά τη μέρα
◯ 1-3 φορές τη μέρα
◯ 4-7 φορές τη μέρα
◯ >7 φορές τη μέρα
◯ Δεν γνωρίζω
◯ Δεν απαντώ

Απολυμαντικά (αντισηπτικά) προϊόντα

◯ Ποτέ
◯ < 1 φορά τη μέρα
◯ 1-3 φορές τη μέρα
◯ 4-7 φορές τη μέρα
◯ >7 φορές τη μέρα
◯ Δεν γνωρίζω
◯ Δεν απαντώ

42. Πόσες μέρες την εβδομάδα εκτελούνταν, κατά μέσο όρο, οι ακόλουθες δραστηριότητες με τη χρήση απολυμαντικών/αντισηπτικών προϊόντων στο σπίτι, ΠΡΙΝ ΤΗΝ ΠΑΝΔΗΜΙΑ, δηλαδή πριν την εφαρμογή των μέτρων περιορισμού;

Καθαρισμός χώρων υγιεινής (π.χ. μπάνιο, τουαλέτα) με τη χρήση απολυμαντικών/αντισηπτικών προϊόντων:

__________________________________
(Μέρες ανά εβδομάδα ΠΡΙΝ ΤΗΝ ΠΑΝΔΗΜΙΑ, δηλαδή πριν την εφαρμογή των μέτρων περιορισμού)

Παρακαλώ σημειώστε το όνομα του προϊόντος που χρησιμοποιούσατε για τον καθαρισμό χώρων υγιεινής:

__________________________________

Καθαρισμός επιφανειών κουζίνας με τη χρήση απολυμαντικών/αντισηπτικών προϊόντων:

__________________________________
(Μέρες ανά εβδομάδα ΠΡΙΝ ΤΗΝ ΠΑΝΔΗΜΙΑ, δηλαδή πριν την εφαρμογή των μέτρων περιορισμού)

Παρακαλώ σημειώστε το όνομα του προϊόντος που χρησιμοποιούσατε για τον καθαρισμό επιφανειών κουζίνας:

__________________________________

Σφουγγάρισμα δαπέδων με τη χρήση απολυμαντικών/αντισηπτικών προϊόντων:

__________________________________
(Μέρες ανά εβδομάδα ΠΡΙΝ ΤΗΝ ΠΑΝΔΗΜΙΑ, δηλαδή πριν την εφαρμογή των μέτρων περιορισμού)

Παρακαλώ σημειώστε το όνομα του προϊόντος που χρησιμοποιούσατε για το σφουγγάρισμα των δαπέδων:

__________________________________

Καθαρισμός επιφανειών σε άλλους χώρους εκτός κουζίνας με τη χρήση απολυμαντικών/αντισηπτικών προϊόντων:

__________________________________
(Μέρες ανά εβδομάδα ΠΡΙΝ ΤΗΝ ΠΑΝΔΗΜΙΑ, δηλαδή πριν την εφαρμογή των μέτρων περιορισμού)

Παρακαλώ σημειώστε το όνομα του προϊόντος που χρησιμοποιούσατε για τον καθαρισμό επιφανειών σε άλλους χώρους εκτός κουζίνας:

__________________________________

**Χρόνος στο σπίτι, στο σχολείο και επαφή με άλλα άτομα ΠΡΙΝ ΤΗΝ ΠΑΝΔΗΜΙΑ, δηλαδή πριν την εφαρμογή των μέτρων περιορισμού;**

43α. Κατά μέσο όρο, πόσες ώρες βρισκόταν στο σπίτι το παιδί σας μια καθημερινή μέρα (Δευτέρα-Παρασκευή) ΠΡΙΝ ΤΗΝ ΠΑΝΔΗΜΙΑ, δηλαδή πριν την εφαρμογή των μέτρων περιορισμού;

__________________________________

43β. Με πόσα άτομα ερχόταν σε επαφή το παιδί σας από Δευτέρα μέχρι Παρασκευή, κατά μέσο όρο, ΠΡΙΝ ΤΗΝ ΠΑΝΔΗΜΙΑ, δηλαδή πριν την εφαρμογή των μέτρων περιορισμού;
Σημείωση: επαφή ορίζεται ως είτε η αμφίδρομη συζήτηση με τρεις ή περισσότερες λέξεις μεταξύ της φυσικής παρουσίας δύο ατόμων είτε η φυσική επαφή (δέρμα με δέρμα) (π.χ. χειραψία, αγκαλιά, φιλί)

Αριθμός ατόμων με τα οποία ερχόταν σε επαφή το παιδί σας στο σπίτι από Δευτέρα μέχρι Παρασκευή, ΠΡΙΝ ΤΗΝ ΠΑΝΔΗΜΙΑ, δηλαδή πριν την εφαρμογή των μέτρων περιορισμού:

__________________________________

Πόσα άτομα με τα οποία ερχόταν σε επαφή το παιδί σας στο σπίτι από Δευτέρα μέχρι Παρασκευή ανήκουν σε ευπαθείς ομάδες ΠΡΙΝ ΤΗΝ ΠΑΝΔΗΜΙΑ, δηλαδή πριν την εφαρμογή των μέτρων περιορισμού;
Στις ευπαθείς ομάδες ανήκουν τα άτομα άνω των 60 ετών, άτομα με χρόνιες παθήσεις και έγκυες γυναίκες.

__________________________________

Αριθμός ατόμων με τα οποία ερχόταν σε επαφή το παιδί σας στο σχολείο από Δευτέρα μέχρι Παρασκευή, ΠΡΙΝ ΤΗΝ ΠΑΝΔΗΜΙΑ, δηλαδή πριν την εφαρμογή των μέτρων περιορισμού:

__________________________________

Αριθμός ατόμων με τα οποία ερχόταν σε επαφή το παιδί σας εκτός σπιτιού και εκτός σχολείου από Δευτέρα μέχρι Παρασκευή, ΠΡΙΝ ΤΗΝ ΠΑΝΔΗΜΙΑ, δηλαδή πριν την εφαρμογή των μέτρων περιορισμού:

__________________________________

44α. Κατά μέσο όρο, πόσες ώρες βρισκόταν στο σπίτι το παιδί σας το σαββατοκύριακο ΠΡΙΝ ΤΗΝ ΠΑΝΔΗΜΙΑ, δηλαδή πριν την εφαρμογή των μέτρων περιορισμού;

__________________________________

44β. Με πόσα άτομα ερχόταν σε επαφή το παιδί σας το σαββατοκύριακο, κατά μέσο όρο, ΠΡΙΝ ΤΗΝ ΠΑΝΔΗΜΙΑ, δηλαδή πριν την εφαρμογή των μέτρων περιορισμού:
Σημείωση: επαφή ορίζεται ως είτε η αμφίδρομη συζήτηση με τρεις ή περισσότερες λέξεις μεταξύ της φυσικής παρουσίας δύο ατόμων είτε η φυσική επαφή (δέρμα με δέρμα) (π.χ. χειραψία, αγκαλιά, φιλί)

Αριθμός ατόμων με τα οποία ερχόταν σε επαφή το παιδί σας στο σπίτι το σαββατοκύριακο, ΠΡΙΝ ΤΗΝ ΠΑΝΔΗΜΙΑ, δηλαδή πριν την εφαρμογή των μέτρων περιορισμού:

__________________________________

Πόσα άτομα με τα οποία ερχόταν σε επαφή το παιδί σας στο σπίτι το σαββατοκύριακο ανήκουν σε ευπαθείς ομάδες, ΠΡΙΝ ΤΗΝ ΠΑΝΔΗΜΙΑ, δηλαδή πριν την εφαρμογή των μέτρων περιορισμού;
Στις ευπαθείς ομάδες ανήκουν τα άτομα άνω των 60 ετών, άτομα με χρόνιες παθήσεις και έγκυες γυναίκες.

__________________________________

Αριθμός ατόμων με τα οποία ερχόταν σε επαφή το παιδί σας εκτός σπιτιού το σαββατοκύριακο, ΠΡΙΝ ΤΗΝ ΠΑΝΔΗΜΙΑ, δηλαδή πριν την εφαρμογή των μέτρων περιορισμού:

__________________________________

#### Γενικά και Δημογραφικά Στοιχεία

45. Φύλο παιδιού:

◯ Αγόρι
◯ Κορίτσι
◯ Δεν θα ήθελα να απαντήσω

---

46. Τόπος γέννησης παιδιού:

◯ Κύπρος
◯ Σε άλλη χώρα της Ευρωπαϊκής Ένωσης
◯ Σε άλλη χώρα εκτός Ευρωπαϊκής Ένωσης

---

Παρακαλώ σημειώστε σε ποια:

__________________________________

---

47. Έτος γέννησης παιδιού:

__________________________________

---

48.Ταχυδρομικός κώδικας κατοικίας:

__________________________________

---

49. Πόλη διαμονής:

◯ Λεμεσός
◯ Πάφος
◯ Λάρνακα
◯ Λευκωσία
◯ Αμμόχωστος

---

50. Δήμος / Κοινότητα:

__________________________________

---

51. Σχολείο:

__________________________________

---

52. Πόσα χρόνια μένει στην Κύπρο το παιδί σας;

__________________________________

53α. Ποιο είναι το υψηλότερο επίπεδο εκπαίδευσης που έχει ολοκληρώσει με επιτυχία η μητέρα του παιδιού;

◯ Δεν έχει φοιτήσει ποτέ σε σχολείο
◯ Δεν έχει τελειώσει το δημοτικό
◯ Δημοτικό σχολείο
◯ Γυμνάσιο (3 χρόνια)
◯ Λύκειο/Τεχνική σχολή (απολυτήριο)
◯ Μεταλυκειακή εκπαίδευση μη τριτοβάθμια
◯ Τριτοβάθμια μη Πανεπιστημιακή
◯ Πανεπιστήμιο (πρώτο πτυχίο)
◯ Πανεπιστήμιο-Μεταπτυχιακό (μόνο Master's degree)
◯ Διδακτορικό
◯ Δεν γνωρίζω/Δεν απαντώ

53β. Ποιο είναι το υψηλότερο επίπεδο εκπαίδευσης που έχει ολοκληρώσει με επιτυχία ο πατέρας του παιδιού;

◯ Δεν έχει φοιτήσει ποτέ σε σχολείο
◯ Δεν έχει τελειώσει το δημοτικό
◯ Δημοτικό σχολείο
◯ Γυμνάσιο (3 χρόνια)
◯ Λύκειο/Τεχνική σχολή (απολυτήριο)
◯ Μεταλυκειακή εκπαίδευση μη τριτοβάθμια
◯ Τριτοβάθμια μη Πανεπιστημιακή
◯ Πανεπιστήμιο (πρώτο πτυχίο)
◯ Πανεπιστήμιο-Μεταπτυχιακό (μόνο Master's degree)
◯ Διδακτορικό
◯ Δεν γνωρίζω/Δεν απαντώ

54. Πόσο είναι το ύψος του παιδιού σας χωρίς παπούτσια;

______________________________
(Σε εκατοστά)

55. Πόσο ζυγίζει το παιδί σας χωρίς ρούχα και παπούτσια;

______________________________
(Σε κιλά)

56. Το παιδί σας παρακολουθεί τα μαθήματα του κανονικά στο σχολείο, μετά την επαναλειτουργία τους, ή επιλέξατε να παραμείνει στο σπίτι;

◯ Παρακολουθεί τα μαθήματα του κανονικά στο σχολείο
◯ Παραμένει στο σπίτι

Συμμετέχει το παιδί σας στο πρόγραμμα διαγνωστικών εξετάσεων για τη νόσο COVID-19;

◯ Ναι
◯ Όχι

Παρακαλώ επιλέξτε τι από τα παρακάτω ισχύει για το αποτέλεσμα της εξέτασης για τη νόσο COVID-19 για το παιδί σας:

◯ Το αποτέλεσμα ήταν αρνητικό
◯ Το αποτέλεσμα ήταν θετικό
◯ Δεν το γνωρίζω
◯ Δεν απαντώ
